# Characterising complex health needs and the use of preventative therapies in the older population: a population-based cohort analysis of UK primary care and hospital linked data

**DOI:** 10.1101/2022.09.30.22280548

**Authors:** Leena Elhussein, Annika M. Jödicke, Ying He, Antonella Delmestri, Danielle E. Robinson, Victoria Y. Strauss, Daniel Prieto-Alhambra

**Author notes:** **Corresponding Author** Annika M. Jödicke, Pharmaco- and Device Epidemiology, Centre for Statistics in Medicine, Nuffield Department of Orthopaedics, Rheumatology and Musculoskeletal Sciences (NDORMS), University of Oxford, Oxford, UK.

## Abstract

**Background:** Several definitions exist for multimorbidity, frailty or polypharmacy, but no formal definition exists for “complex health needs”. We aimed to identify and characterise older people with complex health needs based on healthcare resource use (unplanned hospitalisations or polypharmacy) or frailty.

**Methods:** In this cohort study, data was extracted from UK primary care records (CPRD GOLD), with linked Hospital Episode Statistics inpatient data. People aged >65 on 1st January 2010, registered in CPRD for ≥1 year were included. We identified complex health needs as the top quintile of unplanned hospitalisations, number of prescribed medicines, and electronic frailty index. We characterised all three cohorts, and quantified point-prevalence and incidence rates of preventative medicines use.

**Results:** Overall, 90597, 110225 and 116076 individuals were included in the hospitalisation, frailty, and polypharmacy cohorts respectively. Frailty and polypharmacy cohorts had the highest bi- directional overlap. Most comorbidities such as diabetes and chronic kidney disease were more common in the frailty and polypharmacy cohorts compared to the hospitalisation cohort. Generally, prevalence of preventative medicines use was highest in the polypharmacy cohort compared to the other two cohorts: For instance, one-year point-prevalence of statins was 64.2% in the polypharmacy cohort vs. 60.5% in the frailty cohort.

**Conclusions:** Three distinct groups of older people with complex health needs were identified. Compared to the hospitalisation cohort, frailty and polypharmacy cohorts had more comorbidities and higher preventative therapies use. Research is needed into the benefit-risk of different definitions of complex health needs and use of preventative therapies in the older population.

## INTRODUCTION

With society growing older, an increasing number of people in the UK is suffering from multimorbidity and receive poly-pharmaceutical treatment: Studies suggest that up to 55% of the population aged ≥65 and 80% of people aged ≥80 were diagnosed with multiple chronic conditions in 2007(1). At the same time, prevalence of polypharmacy rose, with the number of older patients receiving ≥10 drugs tripled between 1995 and 2010.(2) Drug treatments targeting multiple single conditions often cumulate, increasing the risk for drug-drug interactions (2) and adverse events. While clinical guidelines provide recommendations for treatment of individual diseases, guidance on the simultaneous treatment of multiple conditions considering the cumulative impact of treatment recommendations is often scarce(3). Aside from comorbidity, a patient’s treatment requirement is influenced by functional status, preferences, and life expectancy. With a lack of clear evidence of treatment risks and benefits, both over- and under-treatment occur in frail patients. For instance, controversial recommendations exist for the use of common preventative treatments such as statins, anti-hypertensives and oral bisphosphonates in multimorbid patients with limited life expectancy and frailty.

As part of its strategy to optimise care for multimorbid patients, the UK’s National Institute for Health and Care Excellence (NICE) published a multimorbidity guideline(4), suggesting the use of primary care electronic health records to identify patients with complex health needs. In particular, the guideline suggested to consider validated tools such as the electronic frailty index (eFI) (5) to identify multimorbid patients at risk for adverse events and unplanned hospital admissions, as well as markers of polypharmacy to identify patients with high treatment burden. Whilst several definitions exist for polypharmacy and frailty, a formal definition to identify patients with “complex health needs” is lacking, precluding good quality research into this important patient group.

The objective of this study was to identify and characterise older patients with complex health needs, using three distinct definitions: 1) unplanned hospitalisation, 2) frailty and 3) polypharmacy. We described patient characteristics for the three individual cohorts, compared them to the background population and explored the overlap between the respective cohorts. In addition, we investigated the population-level use of key preventative therapies in the individual cohorts.

## MATERIALS AND METHODS

### Data source

We extracted data from CPRD GOLD, a UK primary care database containing >2.8million anonymised records of active patients (September 2019)(6). CPRD was representative of the UK population in terms of age, sex and ethnicity (7). We linked CPRD to Hospital Episodes Statistics inpatient and emergency admission data (HES APC and A&E) and Office for National statistics (ONS) mortality data.

### Population

All subjects eligible for CPRD-HES at study start (01/01/2010), aged >65 years, acceptable for clinical research, who were registered with an ‘up-to-standard’ practice(7) for at least 1 year comprised the source population.

### Cohort definitions

Among the source population, we identified three cohorts of older patients with ‘complex health needs’ based on the following health-care markers:

a) Hospitalisation cohort: defined by the number of unplanned hospital admissions in 2009.

b) Frailty cohort: defined using the validated eFI score developed by Clegg et al (5), and based on the count of frailty markers/deficits as recorded during 2009.

c) Polypharmacy cohort: defined by the number of different drug substances prescribed in 2009.

After examining the data distribution of these markers, patients in the top quintile (20%) constituted the respective cohorts. If there was no clear cut-off to determine the top quintile due to count data, the nearest division based on the data distribution was chosen. Patients could be included to multiple cohorts at the same time. Patients who belonged to all three cohorts constituted the overlap group, whereas those who did not belong to any of the three cohorts comprised the background population.

### Utilisation of preventive therapies

The NICE multimorbidity guideline recommended research into continuing and stopping of three preventive therapies, namely bisphosphonates, statins, and anti-hypertensives, as those may potentially be avoidable in patients with limited life expectancy and frailty (4). We evaluated the use of oral bisphosphonates, statins, and anti-hypertensives, separately in each cohort. Prescriptions of preventative treatments were identified based on product codes in CPRD (Code lists available in the Supplements). Anti-hypertensives were analysed as a group, and separately by drug class according to the WHO-ATC classification. We calculated treatment episodes by combining individual prescriptions, allowing for a maximum 90-day gap between prescriptions and with adding a 90-day washout period at the end of each episode.

### Statistical analyses

We calculated one month, three month and one year point prevalence (PP) prior to start date to identify prevalent users of oral bisphosphonates, statins, and anti-hypertensives in each cohort, the overlap group and background population, separately. Furthermore, we calculated one-year treatment incidence rates (IR) in the first year after start date after excluding one-year prevalent patients of each drug class for the same groups. We assumed a Poisson distribution for both analyses. Incidences were counted as the first prescription after start date and IRs were calculated per 1000 person years. Patients exited the study at the earliest of practice last collection date, patient transfer-out of practice date or death date(8). For IR analysis, a patient would contribute to the sum of person-years until a prescription of the preventative therapy is reported. We used STATA (Version 15.1) for cohort identification, and R (Version 3.6.0) for cohort characterisation and calculation of PPs and IRs.

## RESULTS

### Cohort identification

Cut-offs for inclusion to each of the three cohorts were: ≥1 unplanned hospital admission for the hospitalisation cohort, an eFI score ≥3 for the frailty cohort, and ≥10 medicines for the polypharmacy cohort. Figure S1 and Table S1 in the Supplements describe the distribution of these markers.

Figure 1 shows the distribution of the 475 371 eligible people belonging to the three cohorts. Frailty and polypharmacy had the highest bi-directional overlap (>50%).

**Figure 1:**
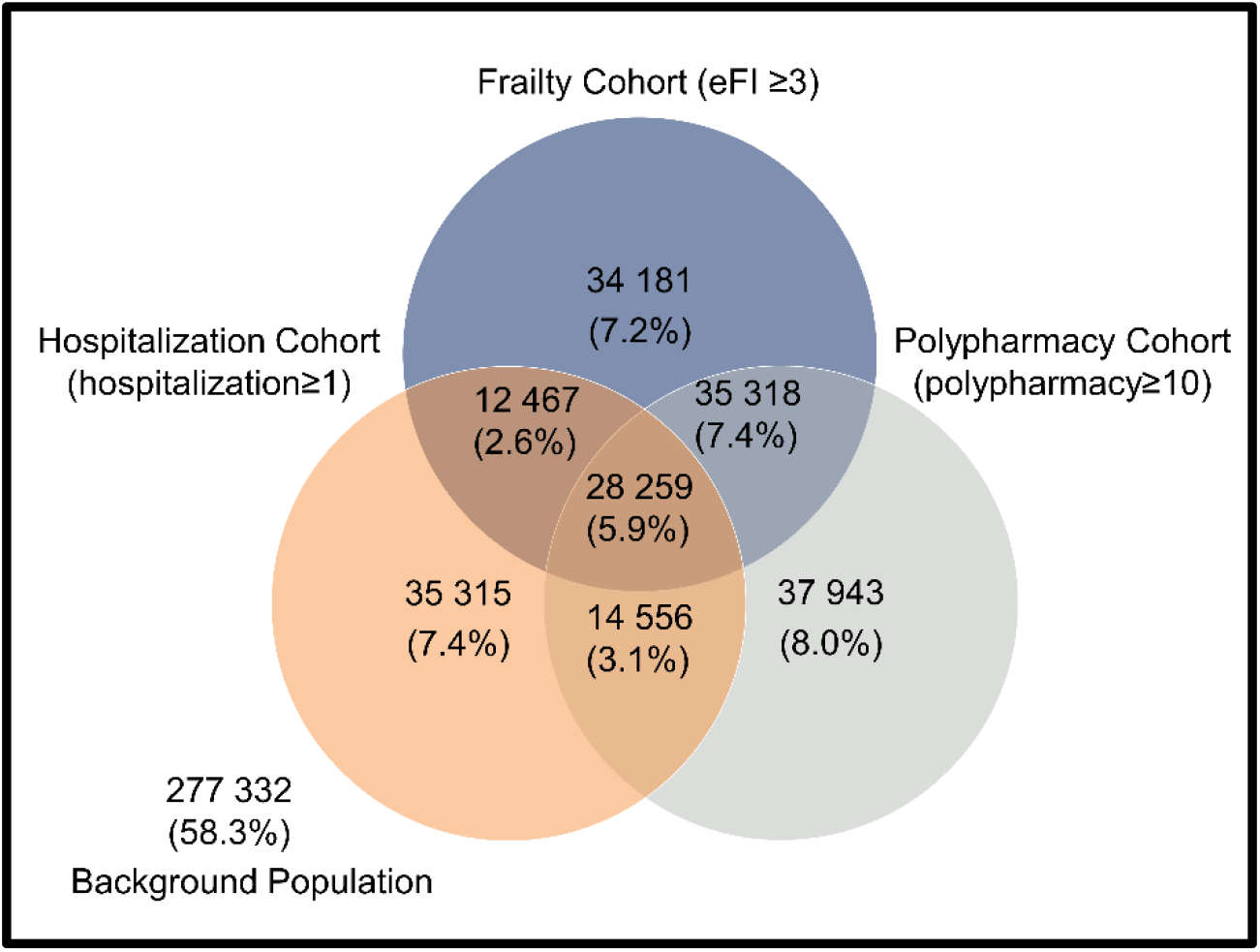
Venn diagram: Number of patients in each cohort of complex health needs, and cohort overlap

### Cohort characteristics

Mean age was similar in the three cohorts and the overlap group, (between 78.2 years and 79.7 years), and slightly lower in the background population (74.7 years). The proportion of males was lower in the three cohorts and the overlap group (between 39.3% and 43.3%) compared to the background population, (46.7%). Table 1 describes the baseline characteristics of the three complex health needs cohorts, the overlap group and the background population.

**Table 1:** Baseline characteristics

|  | **Hospitalisation Cohort** | **Frailty Cohort** | **Polypharmacy Cohort** | **Overlap group** | **Background population** |
| --- | --- | --- | --- | --- | --- |
| n | 90597 | 110225 | 116076 | 28259 | 277332 |
| Hospitalisation Cohort | 90597 (100.0%) | 40726 (36.9%) | 42815 (36.9%) | 28259 (100.0%) | 0 (0) |
| Frailty Cohort | 40726 (45.0%) | 110225 (100.0%) | 63577 (54.8%) | 28259 (100.0%) | 0 (0) |
| Polypharmacy Cohort | 42815 (47.3%) | 63577 (57.7%) | 116076 (100.0%) | 28259 (100.0%) | 0 (0) |
| Gender = Male | 39201 (43.3%) | 45272 (41.1%) | 45578 (39.3%) | 11470 (40.6%) | 129406 (46.7%) |
| Age (mean (SD)) | 78.16 (7.76) | 78.74 (7.41) | 78.19 (7.38) | 79.66 (7.41) | 74.73 (6.97) |
| Socio-economic status |  |  |  |  |  |
| 1 (least deprived) | 20559 (22.7%) | 24226 (22.0%) | 25520 (22.0%) | 5632 (19.9%) | 78067 (28.1%) |
| 2 | 21641 (23.9%) | 25823 (23.4%) | 26496 (22.8%) | 6340 (22.4%) | 72208 (26.0%) |
| 3 | 19291 (21.3%) | 23633 (21.4%) | 24482 (21.1%) | 6003 (21.2%) | 57973 (20.9%) |
| 4 | 16921 (18.7%) | 21331 (19.4%) | 22730 (19.6%) | 5760 (20.4%) | 44379 (16.0%) |
| 5 (most deprived) | 12112 (13.4%) | 15134 (13.7%) | 16744 (14.4%) | 4501 (15.9%) | 24525 (8.8%) |
| Smoking status |  |  |  |  |  |
| Ex-smoker | 34409 (38.0%) | 46401 (42.1%) | 48782 (42.0%) | 12628 (44.7%) | 85474 (30.8%) |
| Non-smoker | 45804 (50.6%) | 53183 (48.2%) | 55991 (48.2%) | 12973 (45.9%) | 147173 (53.1%) |
| Current smoker | 8368 (9.2%) | 9323 (8.5%) | 9853 (8.5%) | 2320 (8.2%) | 27988 (10.1%) |
| Drinking status |  |  |  |  |  |
| Ex-drinker | 3369 (3.7%) | 4866 (4.4%) | 5007 (4.3%) | 1503 (5.3%) | 5228 (1.9%) |
| Non-drinker | 17779 (19.6%) | 24343 (22.1%) | 25945 (22.4%) | 6832 (24.2%) | 36386 (13.1%) |
| Current drinker | 50181 (55.4%) | 62745 (56.9%) | 64124 (55.2%) | 15267 (54.0%) | 159807 (57.6%) |
| BMI* (mean (SD)) | 27.10 (5.55) | 27.95 (5.78) | 28.31 (5.86) | 27.59 (5.97) | 26.73 (4.66) |
| Charlson Comorbidity Index<sup>a</sup> (mean (SD)) | 0.37 (0.92) | 0.51 (1.02) | 0.38 (0.91) | 0.64 (1.15) | 0.08 (0.42) |
| eFI<sup>a</sup> (median [IQR]) | 2 [1, 3] | 3 [3, 4] | 3 [2, 4] | 4 [3, 5] | 1 [0, 2] |
| Unplanned hospital admissions<sup>a</sup> (median [IQR]) | 1 [1, 2] | 0 [0, 1] | 0 [0, 1] | 1 [1, 2] | 0 [0, 0] |
| Number of different drugs<sup>a</sup> (median [IQR]) | 9 [6, 13] | 10 [8, 14] | 13 [11, 15] | 14 [12, 18] | 5 [3, 9] |

BMI = Body mass index; eFI = electronic Frailty Index
<sup>a</sup> Calculated based on the year prior to start
Missingness was around 0.1% for socio-economic status, ranged between 1.2% to 6.0% for smoking, 16.5% to 27.4% for drinking, and 19.8% to 64.1% for BMI

### Comorbidities

Comorbidity burden was higher in the three complex health needs cohorts and the overlap group compared to the background population, especially for diabetes, ischemic heart disease, respiratory disease and diseases of the urinary system. For most conditions, frailty cohort had the highest prevalence, followed by the polypharmacy cohort. The hospitalisation cohort had the highest prevalence of myocardial infarction, cancer and fragility fractures. Figure 2 shows the prevalence of each individual condition included in the Charlson Comorbidity Index and eFI score in the year prior to start date.

**Figure 2:**
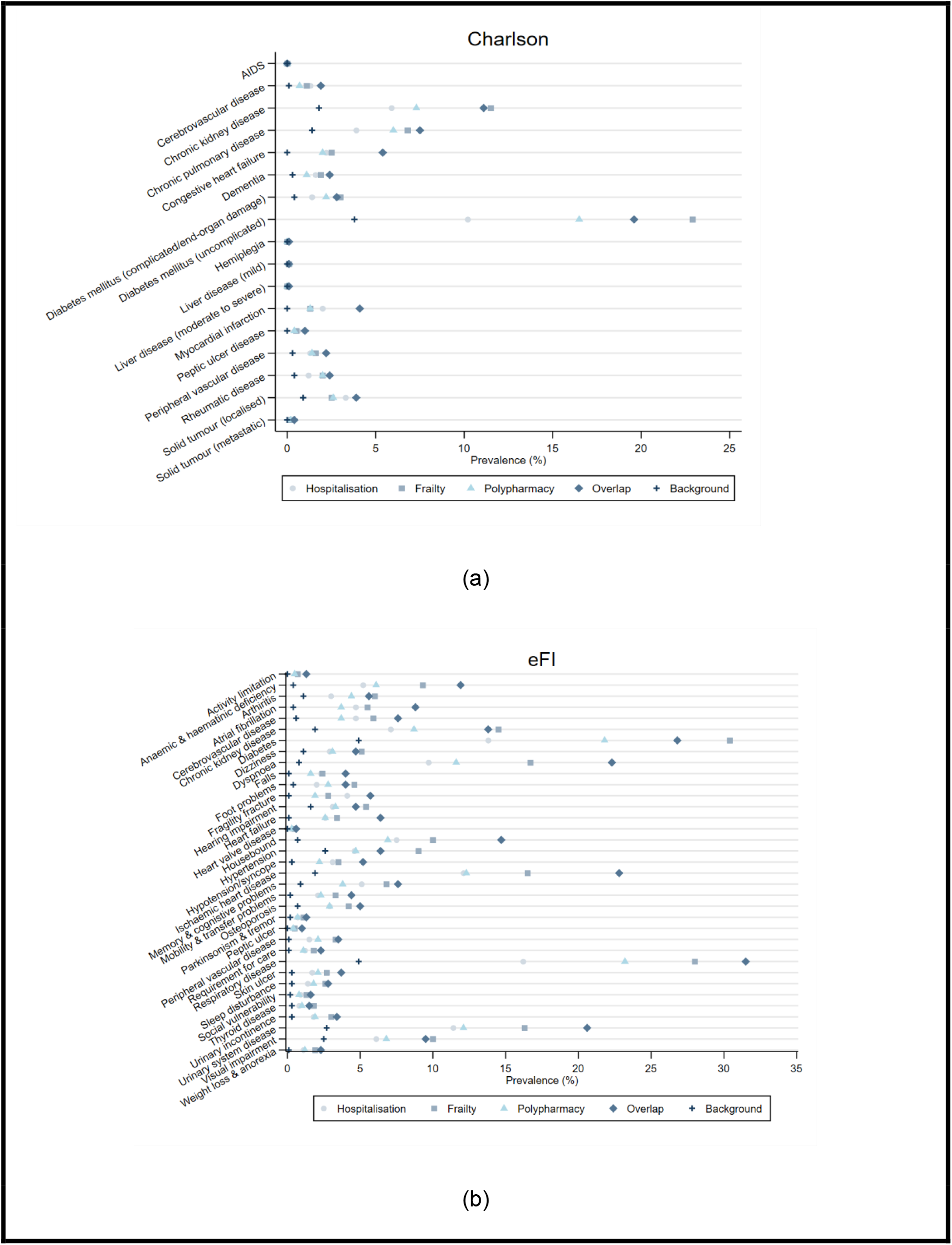
Prevalence of deficits/conditions - in the year prior to start - for all cohorts, overlap and background population for (a) Charlson morbidity index, and (b) eFI

### Preventative therapy use

#### Point prevalence

Figure 3 describes one-year PP of oral bisphosphonates, statins and anti-hypertensives for all cohorts. Compared to the hospitalisation and frailty cohort, the polypharmacy cohort had a higher prevalence of preventative treatment use. Prevalence was highest in the overlap group and lowest in the background population. Among the three preventative therapies, the use of anti-hypertensives was substantially higher than for oral bisphosphonates and statins for all cohorts (≥70.0%). Figure S2 in the Supplements describes one and three-month PP, and Tables S2a-e report PP of the different anti-hypertensive classes.

**Figure 3:**
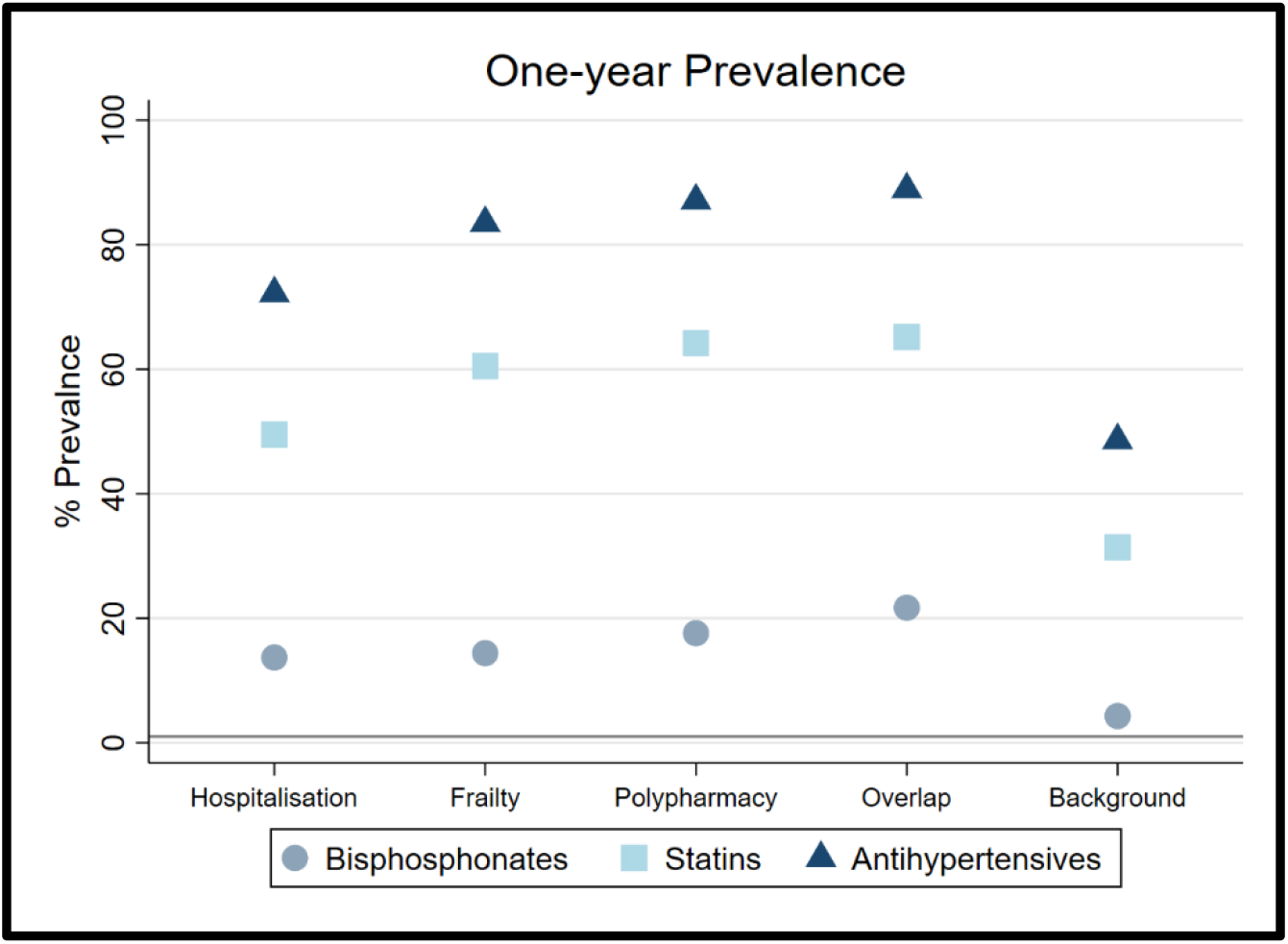
One-year Point Prevalence for oral bisphosphonates, statins and anti-hypertensives, for all cohorts

#### Incidence

Table 2 shows one-year IRs of oral bisphosphonates, statins and anti-hypertensives for all cohorts. Both the frailty and polypharmacy cohort had similar IRs for the preventative therapies, with hospitalisation cohort having the lowest IRs among the three cohorts in most cases. As with point prevalence, the overlap group had generally higher IRs than the three identified cohorts. Tables S3a-e in the Supplements describe the one-year IRs for the different classes of anti-hypertensives.

**Table 2:** One-year IRs – excluding patients with one-year prevalence

|  | **Bisphosphonates** |  |  |
| --- | --- | --- | --- |
|  | **N** | **PY** | **IR (95% CI)** |
| **Hospitalisation Cohort** | 2233 | 69984.6 | 31.91 (30.6, 33.2) |
| **Frailty Cohort** | 2441 | 85996.6 | 28.39 (27.3, 29.5) |
| **Polypharmacy Cohort** | 2477 | 87269.8 | 28.38 (27.3, 29.5) |
| **Overlap group** | 780 | 19081.4 | 40.88 (38.0, 43.8) |
| **Background population** | 2604 | 253071.6 | 10.29 (9.9, 10.7) |
|  | **Statins** |  |  |
|  | **N** | **PY** | **IR (95% CI)** |
| **Hospitalisation Cohort** | 2472 | 39902.3 | 61.95 (59.5, 64.4) |
| **Frailty Cohort** | 2520 | 38216.8 | 65.94 (63.4, 68.5) |
| **Polypharmacy Cohort** | 2327 | 36515.6 | 63.73 (61.1, 66.3) |
| **Overlap group** | 551 | 8115.8 | 67.89 (62.2, 73.6) |
| **Background population** | 8425 | 177430.1 | 47.48 (46.5, 48.5) |
|  | **Anti-hypertensives** |  |  |
|  | **N** | **PY** | **IR (95% CI)** |
| **Hospitalisation Cohort** | 2793 | 21610.1 | 129.25 (124.5, 134.0) |
| **Frailty Cohort** | 2400 | 15449.0 | 155.35 (149.1, 161.6) |
| **Polypharmacy Cohort** | 1972 | 12759.6 | 154.55 (147.7, 161.4) |
| **Overlap group** | 491 | 2481.901 | 197.83 (180.3, 215.3) |
| **Background population** | 9622 | 131709.3 | 73.06 (71.6, 74.5) |

N= Number of incident users, PY = person years, IR = incidence rate

## DISCUSSION

### Principal findings

We identified three cohorts of older patients with complex healthcare needs, based on healthcare resource use (unplanned hospitalisations, polypharmacy) or frailty. Compared to the background population, patients included in either of the three cohorts were older, had a higher comorbidity burden, higher number of prescribed drugs, and a substantially higher prevalence and incidence of preventative drug use.

### Assessment of complex healthcare needs in the older population

#### Identification of patients with complex health needs

Following the recommendation of the NICE multimorbidity guideline(4), we identified patients with complex health needs in UK primary care data and inpatient records. The utilisation of primary care data allows for a systematic and comprehensive approach to identify potential target groups for preventative measures, who might benefit most from care optimisation. We defined patients with “complex health needs” as those individuals belonging to the top quintile of the distribution of the respective heath markers. While multiple definitions for polypharmacy and frailty are used side-by-side, we opted for a data-driven approach to identify the most vulnerable among the older population, allowing for a uniform definition applicable in different healthcare settings, data types and patient collectives. Access to large and representative population-based data allowed us to anchor our definitions based on their public health relevance.

Although the association between frailty, polypharmacy and healthcare utilisation has been described previously (5, 9-16), no direct comparison of patient groups identified by these distinct markers has been done.

##### Polypharmacy

While various definitions of polypharmacy are applied in the literature (17), no standard is currently agreed upon (18, 19). Frequently, the concomitant prescription of ≥5 medicines is considered polypharmacy, with ≥10 prescriptions referred to as excessive, severe or hyper-polypharmacy (2, 19). Using our data-driven approach, we identified a cut-off of ≥10 medicines per year for the polypharmacy cohort. While our approach to cumulatively assess drug use might overestimate the number of drugs actually taken concomitantly, it reflects the patients’ treatment burden with respect to both acute and chronic indications over a long time period, providing a pragmatic screening tool.

##### Unplanned hospitalisations

Prevention of unplanned and avoidable hospitalisations are a public health priority, as they present a major burden for the affected patients and substantial costs for the health care system. Our study found that 19.1% of the study population was admitted to hospital for any unplanned reason in 2009, leading to the inclusion of all these patients to the hospitalisation cohort.

##### Frailty

While drug prescriptions and unplanned hospitalisations are well recorded in real world data, the assessment of a patients’ frailty status is challenging. Physical and mental fitness vary greatly among patients of the same age and while a patient’s frailty will greatly influence a general practitioners treatment decision, such characteristics are typically not recorded in primary care databases. Therefore, frailty is one of the most important confounders in pharmaco-epidemiological studies among the older population.

The concept of frailty rapidly evolved over the last two decades, with the two major concepts of “frailty phenotype” and “frailty indexes” being established (9). In recent years, different approaches have been developed to identify frailty in routinely collected data (5, 20- 23).

#### Association between polypharmacy, frailty and unplanned hospitalisation

To our knowledge, this is the first study directly comparing three different markers of complex heath needs to characterise patient groups. The overlap of the three individual cohorts identified in our study illustrates the association between polypharmacy, frailty and hospitalisation. A recent systematic review summarised the bi-directional association between frailty and polypharmacy(9): mean drug consumption was reported to be higher among frail older patients compared to robust individuals, and increased likelihood for frailty was described among patients with polypharmacy (OR 1.77–2.55)(12-14) and hyper-polypharmacy (OR 4.47–5.8) (12-14), respectively. In addition, frailty among the UKs community-dwelling older was associated with reduced quality of life, (15) increased annual GP consultations, hospital admissions and elevated healthcare costs (5, 24).

Our results are in line with the associations described in the literature, with a large pairwise overlap between hospitalisation, frailty and polypharmacy cohort. However, we also found a substantial number of patients only included in *one* of the respective cohorts. This highlights that, while there is a strong association, the markers of complex heath needs are distinct and should not be used interchangeably but rather complement each other. Future pharmaco- epidemiological studies should consider multiple patient characteristics when designing studies among geriatric patients.

### Cohort characteristics and use of preventative treatments in the older people

#### Comorbidity burden

Multimorbidity is often poorly reported and definitions vary greatly between individual studies(25). Consistent across different measures, comorbidity burden in our complex health needs cohorts was substantially higher than in the background population. As counts of deficits were used to define the frailty cohort, it was to be expected that the highest prevalence of comorbidities would be found in the frailty cohort and overlap group.

#### Preventative treatments

Patients with complex healthcare needs were commonly prescribed preventative treatments, with both incidence and prevalence being significantly higher than for the background population. Direct comparison of use of preventative therapies to previous observational studies is difficult considering differences in patient collectives, age-group definitions, and treatment indication (26, 27) between individual studies. However, in the UK, annual initiation rates of statins previously reported for people aged 60-84 years were between 30-50/1000 person-years(28), which is largely comparable to our results. Likewise, incidence rates for anti-osteoporosis treatment (12.5 to 26.0/1000 person-years) were previously reported for women aged 65-84 years (29), with considerably lower rates reported in males. UK based studies reporting incidence rates of antihypertensive treatment among the older population are scarce. A CPRD based study reports annual IR of approximately 15% in 2001 assessing hypertensive patients aged ≥40 years with ≥3 cardiovascular risk factors (30). In our study, incidence rates varied across the different cohorts, with IR 73.1/1000 and 197.8/1000 patient years for background population and overlap group.

### Strengths and limitations

Our study comes with both strengths and limitations. We consider the use of CPRD, which is frequently used for pharmaco-epidemiological studies(31) and provides good quality records for research, a particular strength. In addition, CPRD is representative for the UK population, which enables the use of results from our study to inform local public health strategies for older residents. Another strength of this study is the direct comparison of the different definitions of complex healthcare needs and their impact on which patients are included into the cohort.

We defined our frailty cohort based on the eFI score. When describing the association between frailty and polypharmacy, it is important to consider that polypharmacy (defined as ≥5 medicines per year) is included as a deficit in the eFI score. Thus, there is an operational association, which may overestimate the true overlap between cohorts. While primary care records include all prescriptions issued by GPs, drugs purchased over-the-counter and prescribed in hospital are not included. Therefore, the true burden of polypharmacy may be underestimated. In addition, while we identified patients with multiple prescribed drugs, no information on the adequateness of polypharmacy in the individual patients is available. Lastly, for practical reasons, we assessed all patient information using a baseline period of one year. Therefore, underreporting of chronic deficits cannot be ruled out.

## Conclusion

Our study shows that depending on the definition used, cohorts of patients with complex healthcare needs slightly differ with respect to their demographic characteristics, comorbidity pattern and drug utilisation. Our results highlight that several indicators of complex healthcare needs should be used in any future research among geriatric populations, as they may complement each other. We found the use of preventative therapies to be common among older patients with complex healthcare needs. Future studies should therefore evaluate the risk-benefit of these medications in older patients with reduced lifespan and high potential for adverse drug reactions (32).

## NOTES

## Data Availability

The data used in this article were obtained from the UK Clinical Practice Research Datalink (CPRD) under the Oxford University CPRD multi-study license. This data cannot be shared publically for ethical reasons.

## Author Contributions

DPA, VS, DR and AD planned the study. AD extracted and curated the data. LE and YH performed the statistical analysis. LE, DPA and AJ interpreted the results. LE and AJ drafted the manuscript. All authors reviewed all versions of the manuscript, agreed to the final version, agreed on the journal the manuscript was submitted to. All authors agree to take responsibility and be accountable for the contents of the manuscript.

### Funding

The research was supported by the National Institute for Health and Care Research (NIHR) Oxford Biomedical Research Centre (BRC). DPA is funded through a NIHR Senior Research Fellowship (Grant number SRF-2018-11-ST2-004). The views expressed in this publication are those of the author(s) and not necessarily those of the NHS, the National Institute for Health and Care Research or the Department of Health.

### Data Availability

The data used in this article were obtained from CPRD under the Oxford University CPRD multi-study license. This data cannot be shared publically for ethical reasons.

### Ethics Approval

The study protocol was approved by the Independent Scientific Advisory Committee (ISAC) for UK Medicines and Healthcare products Regulatory Agency database research (Protocol No 19_132A2), and can be made available to the journal reviewers upon request. The data accessed met the relevant data privacy and protection regulations.

### Patient and Public Involvement

A frail patient on anti-osteoporosis therapy and two charity representatives have reviewed the protocol and provided helpful feedback.

## Acknowledgement

This study is based in part on data from the Clinical Practice Research Datalink obtained under the University of Oxford multi-study license from the UK Medicines and Healthcare products Regulatory Agency. The data is provided by patients and collected by the NHS as part of their care and support. The interpretation and conclusions contained in this study are those of the authors alone.

