## Supplements for "Characterising complex health needs and the use of preventative therapies in the older population: a population-based cohort analysis of UK primary care and hospital linked data"

### Tables and figures

Table 1.1 and Figure 1.1 describe the distribution of the healthcare markers used to identify the cohorts to create the cohorts.

Table S1 descriptive statistics of variables used to identify cohorts

| Overall population (n=475371) | | | | | |
|---|---|---|---|---|---|
| **Variable** | **Min** | **P25** | **P50 (median)** | **P75** | **Max** |
| **Unplanned hospital admission[1]** | 0 | 0 | 0 | 0 | 76 |
| **eFI[2]** | 0 | 0 | 1 | 2 | 13 |
| **Polypharmacy[3]** | 0 | 3 | 6 | 9 | 52 |

1 Defined by the number of unplanned hospital admissions (through accidents and emergency rooms or admitted patient care), identified in the linked HES data in 2009
2 Defined using the validated eFI score developed by Clegg et al(5). and currently used by the NHS to support routine frailty identification(8). The eFI was calculated based on a count of frailty markers/deficits as recorded during 2009 in CPRD using a pre-specified list of Read codes(5).
3 Defined by the number of different drug substances prescribed in 2009. For each patient, all prescriptions issued by GPs in 2009 were identified and for these prescriptions, the drug substance was retrieved using the “PRODUCT” dictionary and “THERAPY” table in CPRD. Fixed combinations of multiple substances in products were counted as one substance.
P25 and P75: 25th and 75th percentile. P50: median, eFI = electronic Frailty Index

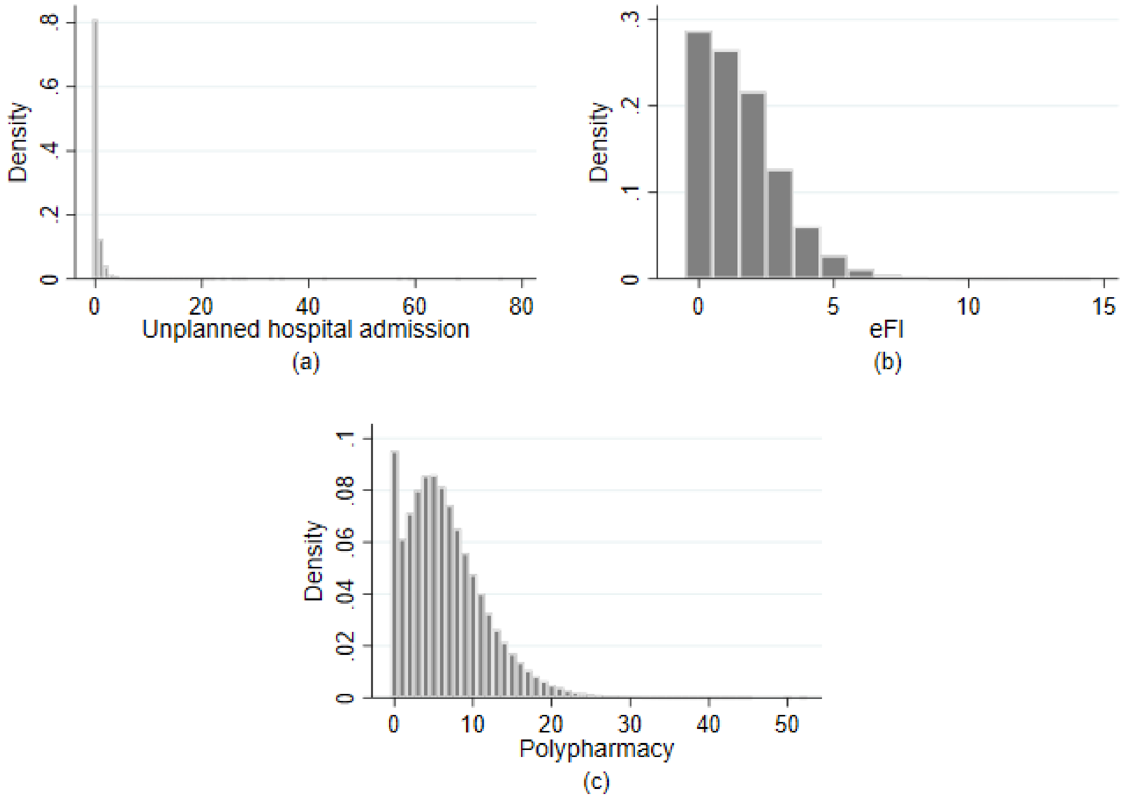

Figure S1 Histograms of variables used to identify cohorts of complex health needs: (a) unplanned hospital admission, (b) eFI and (c) polypharmacy

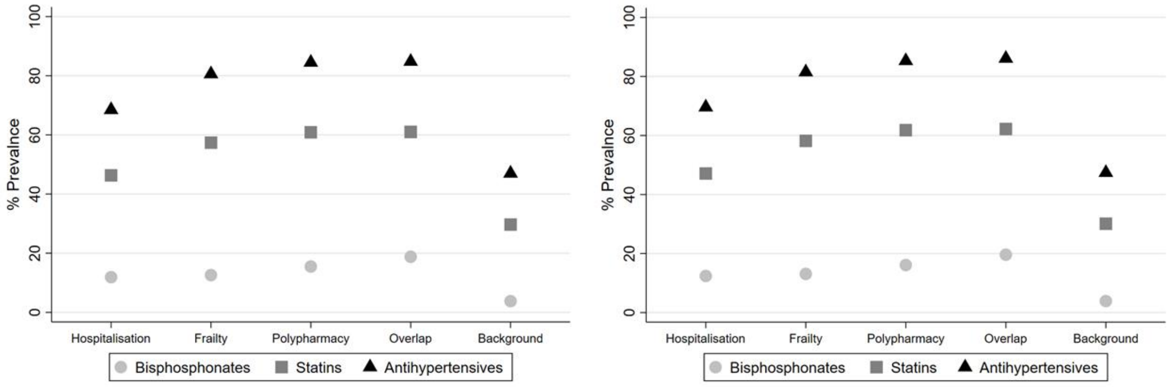

One-month Point Prevalence
(a)

Three-month Point Prevalence
(b)

Figure S2 Point Prevalence for oral bisphosphonates, statins and anti-hypertensives, for all cohorts at (a) 1 month and (b) 3 months

Table S2a: Point prevalence of the different antihypertensives classes for the Hospitalisation cohort

| Hospitalisation cohort N=90597 | | | | | | |
|---|---|---|---|---|---|---|
| **Drug name** | **1 month PP** | | **3 months PP** | | **1 year PP** | |
| | **N prevalent** | **PP (95% CI)** | **N prevalent** | **PP (95% CI)** | **N prevalent** | **PP (95% CI)** |
| ACE-I + CCB | 11 | 0 (0, 0) | 11 | 0 (0, 0) | 15 | 0 (0, 0) |
| ACE-I + Diuretics | 281 | 0.3 (0.3, 0.3) | 300 | 0.3 (0.3, 0.4) | 388 | 0.4 (0.4, 0.5) |
| ARB + CCB | 24 | 0 (0, 0) | 25 | 0 (0, 0) | 26 | 0 (0, 0) |
| ARB + Diuretics | 141 | 0.2 (0.1, 0.2) | 149 | 0.2 (0.1, 0.2) | 180 | 0.2 (0.2, 0.2) |
| Betablockers + CCB | 26 | 0 (0, 0) | 27 | 0 (0, 0) | 32 | 0 (0, 0) |
| Betablockers + Diuretics | 303 | 0.3 (0.3, 0.4) | 323 | 0.4 (0.3, 0.4) | 422 | 0.5 (0.4, 0.5) |
| Renin-Inhibitors | 60 | 0.1 (0.1, 0.1) | 63 | 0.1 (0.1, 0.1) | 87 | 0.1 (0.1, 0.1) |
| ACE-I | 27588 | 30.5 (30.1, 30.8) | 28486 | 31.4 (31.1, 31.8) | 31419 | 34.7 (34.3, 35.1) |
| ARB | 11724 | 12.9 (12.7, 13.2) | 12045 | 13.3 (13.1, 13.5) | 13076 | 14.4 (14.2, 14.7) |
| All antihypertensives | 62056 | 68.5 (68.0, 69.0) | 63017 | 69.6 (69.0, 70.1) | 65387 | 72.2 (71.6, 72.7) |
| Other antihypertensives* | 5427 | 6.0 (5.8, 6.2) | 5749 | 6.3 (6.2, 6.5) | 6760 | 7.5 (7.3, 7.6) |
| Betablockers | 22774 | 25.1 (24.8, 25.5) | 23406 | 25.8 (25.5, 26.2) | 25467 | 28.1 (27.8, 28.5) |
| CCB | 23376 | 25.8 (25.5, 26.1) | 24378 | 26.9 (26.6, 27.2) | 27540 | 30.4 (30.0, 30.8) |
| Diuretics | 32672 | 36.1 (35.7, 36.5) | 34173 | 37.7 (37.3, 38.1) | 38126 | 42.1 (41.7, 42.5) |

Note: ACE-I = Angiotensin-converting-enzyme inhibitor, CCB = calcium channel blockers, ARB = Angiotensin-II receptor blockers. *other antihypertensives comprise: Ambrisentan, Betanidine, Bosentan, Clonidine,Debrisoquine, Doxazosin,Guanethidine, Hydralazine, Indoramin, Ketanserin, Macitentan, Methoserpidine ,Methyldopa, Methyldopate, Metirosine, Minoxidil, Moxonidine, Prazosin, Reserpine, Riociguat, Sitaxentan, Sodium nitroprusside and Trimetaphan camsilate

Table S2b: Point prevalence of the different antihypertensives classes for the frailty cohort

| Frailty cohort N=110225 | | | | | | |
|---|---|---|---|---|---|---|
| **Drug name** | **1 month PP** | | **3 months PP** | | **1 year PP** | |
| | **N prevalent** | **PP (95% CI)** | **N prevalent** | **PP (95% CI)** | **N prevalent** | **PP (95% CI)** |
| ACE-I + CCB | 29 | 0 (0, 0) | 30 | 0 (0, 0) | 38 | 0 (0, 0) |
| ACE-I + Diuretics | 473 | 0.4 (0.4, 0.5) | 501 | 0.5 (0.4, 0.5) | 617 | 0.6 (0.5, 0.6) |
| ARB + CCB | 22 | 0 (0, 0) | 23 | 0 (0, 0) | 23 | 0 (0, 0) |
| ARB + Diuretics | 228 | 0.2 (0.2, 0.2) | 238 | 0.2 (0.2, 0.2) | 301 | 0.3 (0.2, 0.3) |
| Betablockers + CCB | 30 | 0 (0, 0) | 31 | 0 (0, 0) | 40 | 0 (0, 0) |
| Betablockers + Diuretics | 484 | 0.4 (0.4, 0.5) | 501 | 0.5 (0.4, 0.5) | 623 | 0.6 (0.5, 0.6) |
| Renin-Inhibitors | 123 | 0.1 (0.1, 0.1) | 131 | 0.1 (0.1, 0.1) | 154 | 0.1 (0.1, 0.2) |
| ACE-I | 42538 | 38.6 (38.2, 39.0) | 43651 | 39.6 (39.2, 40.0) | 47382 | 43.0 (42.6, 43.4) |
| ARB | 19638 | 17.8 (17.6, 18.1) | 20027 | 18.2 (17.9, 18.4) | 21325 | 19.3 (19.1, 19.6) |
| All antihypertensives | 88863 | 80.6 (80.1, 81.2) | 89797 | 81.5 (80.9, 82.0) | 92014 | 83.5 (82.9, 84.0) |
| Other antihypertensives* | 9739 | 8.8 (8.7, 9.0) | 10137 | 9.2 (9.0, 9.4) | 11480 | 10.4 (10.2, 10.6) |
| Betablockers | 31101 | 28.2 (27.9, 28.5) | 31767 | 28.8 (28.5, 29.1) | 34106 | 30.9 (30.6, 31.3) |
| CCB | 35338 | 32.1 (31.7, 32.4) | 36522 | 33.1 (32.8, 33.5) | 40453 | 36.7 (36.3, 37.1) |
| Diuretics | 48364 | 43.9 (43.5, 44.3) | 50160 | 45.5 (45.1, 45.9) | 55022 | 49.9 (49.5, 50.3) |

Note: ACE-I = Angiotensin-converting-enzyme inhibitor, CCB = calcium channel blockers, ARB = Angiotensin-II receptor blockers. *other antihypertensives comprise: Ambrisentan, Betanidine, Bosentan, Clonidine,Debrisoquine, Doxazosin,Guanethidine, Hydralazine, Indoramin, Ketanserin, Macitentan, Methoserpidine ,Methyldopa, Methyldopate, Metirosine, Minoxidil, Moxonidine, Prazosin, Reserpine, Riociguat, Sitaxentan, Sodium nitroprusside and Trimetaphan camsilate

Table S2c: Point prevalence of the different antihypertensives classes for the polypharmacy cohort

| Polypharmacy cohort N=116076 | | | | | | |
|---|---|---|---|---|---|---|
| **Drug name** | **1 month PP** | | **3 months PP** | | **1 year PP** | |
| | **N prevalent** | **PP (95% CI)** | **N prevalent** | **PP (95% CI)** | **N prevalent** | **PP (95% CI)** |
| ACE-I + CCB | 18 | 0 (0, 0) | 19 | 0 (0, 0) | 33 | 0 (0, 0) |
| ACE-I + Diuretics | 503 | 0.4 (0.4, 0.5) | 536 | 0.5 (0.4, 0.5) | 660 | 0.6 (0.5, 0.6) |
| ARB + CCB | 33 | 0 (0, 0) | 34 | 0 (0, 0) | 36 | 0 (0, 0) |
| ARB + Diuretics | 308 | 0.3 (0.2, 0.3) | 326 | 0.3 (0.3, 0.3) | 396 | 0.3 (0.3, 0.4) |
| Betablockers + CCB | 29 | 0 (0, 0) | 32 | 0 (0, 0) | 39 | 0 (0, 0) |
| Betablockers + Diuretics | 461 | 0.4 (0.4, 0.4) | 490 | 0.4 (0.4, 0.5) | 620 | 0.5 (0.5, 0.6) |
| Renin-Inhibitors | 173 | 0.1 (0.1, 0.2) | 183 | 0.2 (0.1, 0.2) | 215 | 0.2 (0.2, 0.2) |
| ACE-I | 45732 | 39.4 (39.0, 39.8) | 46947 | 40.4 (40.1, 40.8) | 50964 | 43.9 (43.5, 44.3) |
| ARB | 23327 | 20.1 (19.8, 20.4) | 23785 | 20.5 (20.2, 20.8) | 25216 | 21.7 (21.5, 22.0) |
| All antihypertensives | 98069 | 84.5 (84.0, 85.0) | 98995 | 85.3 (84.8, 85.8) | 101121 | 87.1 (86.6, 87.7) |
| Other antihypertensives* | 11867 | 10.2 (10.0, 10.4) | 12373 | 10.7 (10.5, 10.8) | 13995 | 12.1 (11.9, 12.3) |
| Betablockers | 37388 | 32.2 (31.9, 32.5) | 38131 | 32.9 (32.5, 33.2) | 40731 | 35.1 (34.7, 35.4) |
| CCB | 41720 | 35.9 (35.6, 36.3) | 43092 | 37.1 (36.8, 37.5) | 47568 | 41.0 (40.6, 41.3) |
| Diuretics | 57259 | 49.3 (48.9, 49.7) | 59339 | 51.1 (50.7, 51.5) | 64681 | 55.7 (55.3, 56.2) |

Note: ACE-I = Angiotensin-converting-enzyme inhibitor, CCB = calcium channel blockers, ARB = Angiotensin-II receptor blockers. *other antihypertensives comprise: Ambrisentan, Betanidine, Bosentan, Clonidine,Debrisoquine, Doxazosin,Guanethidine, Hydralazine, Indoramin, Ketanserin, Macitentan, Methoserpidine ,Methyldopa, Methyldopate, Metirosine, Minoxidil, Moxonidine, Prazosin, Reserpine, Riociguat, Sitaxentan, Sodium nitroprusside and Trimetaphan camsilate

Table S2d: Point prevalence of the different antihypertensives classes for the overlap group

| Overlap group N=28259 | | | | | | |
|---|---|---|---|---|---|---|
| Drug name | 1 month PP | | 3 months PP | | 1 year PP | |
| | N prevalent | PP (95% CI) | N prevalent | PP (95% CI) | N prevalent | PP (95% CI) |
| ACE-I + CCB | 6 | 0 (0, 0) | 6 | 0 (0, 0) | 8 | 0 (0, 0.001) |
| ACE-I + Diuretics | 94 | 0.3 (0.3, 0.4) | 106 | 0.4 (0.3, 0.5) | 145 | 0.5 (0.4, 0.6) |
| ARB + CCB | 6 | 0 (0, 0) | 7 | 0 (0, 0.1) | 7 | 0 (0, 0.1) |
| ARB + Diuretics | 41 | 0.1 (0.1, 0.2) | 45 | 0.2 (0.1, 0.2) | 61 | 0.2 (0.2, 0.3) |
| Betablockers + CCB | 0 | NA | <5 | NA | <5 | NA |
| Betablockers + Diuretics | 51 | 0.2 (0.1, 0.2) | 57 | 0.2 (0.2, 0.3) | 103 | 0.4 (0.3, 0.4) |
| Renin-Inhibitors | 31 | 0.1 (0.1, 0.2) | 31 | 0.1 (0.1, 0.2) | 43 | 0.2 (0.1, 0.2) |
| ACE-I | 11314 | 40.0 (39.3, 40.8) | 11818 | 41.8 (41.1, 42.6) | 13253 | 46.9 (46.1, 47.7) |
| ARB | 5254 | 18.6 (18.1, 19.1) | 5435 | 19.2 (18.7, 19.8) | 6016 | 21.3 (20.8, 21.8) |
| All antihypertensives | 23957 | 84.8 (83.7, 85.9) | 24323 | 86.1 (85.0, 87.2) | 25112 | 88.9 (87.8, 90.0) |
| Other antihypertensives* | 2559 | 9.1 (8.7, 9.4) | 2741 | 97. (9.3, 10.1) | 3300 | 11.7 (11.3, 12.1) |
| Betablockers | 9314 | 33.0 (32.3, 33.6) | 9626 | 34.1 (33.4, 34.8) | 10562 | 37.4 (36.7, 38.1) |
| CCB | 9208 | 32.6 (31.9, 33.3) | 9737 | 34.5 (33.8, 35.1) | 11318 | 40.1 (39.3, 40.8) |
| Diuretics | 14835 | 52.5 (51.7, 53.3) | 15547 | 55.0 (54.2, 55.9) | 17300 | 61.2 (60.3, 62.1) |

Note: ACE-I = Angiotensin-converting-enzyme inhibitor, CCB = calcium channel blockers, ARB = Angiotensin-II receptor blockers. *other antihypertensives comprise: Ambrisentan, Betanidine, Bosentan, Clonidine,Debrisoquine, Doxazosin,Guanethidine, Hydralazine, Indoramin, Ketanserin, Macitentan, Methoserpidine ,Methyldopa, Methyldopate, Metirosine, Minoxidil, Moxonidine, Prazosin, Reserpine, Riociguat, Sitaxentan, Sodium nitroprusside and Trimetaphan camsilate

Table S2e: Point prevalence of the different antihypertensives classes for the background population

| Background population N=277332 | | | | | | |
|---|---|---|---|---|---|---|
| **Drug name** | **1 month PP** | | **3 months PP** | | **1 year PP** | |
| | **N prevalent** | **PP (95% CI)** | **N prevalent** | **PP (95% CI)** | **N prevalent** | **PP (95% CI)** |
| ACE-I + CCB | 50 | 0 (0, 0) | 52 | 0 (0, 0) | 57 | 0 (0, 0) |
| ACE-I + Diuretics | 1052 | 0.4 (0.4, 0.4) | 1066 | 0.4 (0.4, 0.4) | 1155 | 0.4 (0.4, 0.4) |
| ARB + CCB | 25 | 0 (0, 0) | 27 | 0 (0, 0) | 30 | 0 (0, 0) |
| ARB + Diuretics | 426 | 0.2 (0.1, 0.2) | 436 | 0.2 (0.1, 0.2) | 459 | 0.2 (0.2, 0.2) |
| Betablockers + CCB | 120 | 0 (0, 0.1) | 121 | 0 (0, 0.1) | 124 | 0 (0, 0.1) |
| Betablockers + Diuretics | 1634 | 0.6 (0.6, 0.6) | 1663 | 0.6 (0.6, 0.6) | 1798 | 0.6 (0.6, 0.7) |
| Renin-Inhibitors | 83 | 0 (0, 0) | 89 | 0 (0, 0) | 99 | 0 (0, 0) |
| ACE-I | 54058 | 19.5 (19.3, 19.7) | 54653 | 19.7 (19.5, 19.9) | 57730 | 20.8 (20.6, 21.0) |
| ARB | 21582 | 7.8 (7.7, 7.9) | 21762 | 7.8 (7.7, 8.0) | 22541 | 8.1 (8.0, 8.2) |
| All antihypertensives | 130411 | 47.0 (46.8, 47.3) | 131354 | 47.4 (47.1, 47.6) | 134675 | 48.6 (48.3, 48.8) |
| Other antihypertensives* | 9762 | 3.5 (3.5, 3.6) | 9959 | 3.6 (3.5, 3.7) | 10781 | 3.9 (3.8, 4.0) |
| Betablockers | 41161 | 14.8 (14.7, 15.0) | 41629 | 15.0 (14.9, 15.2) | 43793 | 15.8 (15.6, 15.9) |
| CCB | 51332 | 18.5 (18.3, 18.7) | 52055 | 18.8 (18.6, 18.9) | 55234 | 19.9 (19.8, 20.1) |
| Diuretics | 56577 | 20.4 (20.2, 20.6) | 57695 | 20.8 (20.6, 21.0) | 61844 | 22.3 (22.1, 22.5) |

Note: ACE-I = Angiotensin-converting-enzyme inhibitor, CCB = calcium channel blockers, ARB = Angiotensin-II receptor blockers. *other antihypertensives comprise: Ambrisentan, Betanidine, Bosentan, Clonidine,Debrisoquine, Doxazosin,Guanethidine, Hydralazine, Indoramin, Ketanserin, Macitentan, Methoserpidine ,Methyldopa, Methyldopate, Metirosine, Minoxidil, Moxonidine, Prazosin, Reserpine, Riociguat, Sitaxentan, Sodium nitroprusside and Trimetaphan camsilate

Table S3a: One-year IRs – excluding one-year prevalence of the different antihypertensives classes for the Hospitalisation cohort

| Drug name | Hospitalisation cohort N=90597 | | |
|---|---|---|---|
| | N incidence | PY | IR (95% CI) |
| **ACE-I + CCB** | <5 | NA | NA |
| **ACE-I + Diuretics** | 18 | 81946.7 | 0.2 (0.1, 0.3) |
| **ARB + CCB** | 9 | 82284.4 | 0.1 (0.0, 0.2) |
| **ARB + Diuretics** | 13 | 82145.1 | 0.2 (0.1, 0.2) |
| **Betablockers + CCB** | 0 | NA | NA |
| **Betablockers + Diuretics** | <5 | NA | NA |
| **Renin-Inhibitors** | 25 | 82217.3 | 0.3 (0.2, 0.4) |
| **ACE-I** | 2905 | 52211.7 | 55.6 (53.6, 57.7) |
| **ARB** | 1230 | 69601.9 | 17.7 (16.7, 18.7) |
| **All antihypertensives** | 2793 | 21610.1 | 129.3 (124.5, 134.0) |
| **Other antihypertensives*** | 848 | 75686.8 | 11.2 (10.5, 12.0) |
| **Betablockers** | 2445 | 57871.6 | 42.3 (40.6, 43.9) |
| **CCB** | 2783 | 55607.2 | 50.1 (48.2, 51.9) |
| **Diuretics** | 4141 | 46198.4 | 89.6 (86.9, 92.4) |

PY=person years

Note: ACE-I = Angiotensin-converting-enzyme inhibitor, CCB = calcium channel blockers, ARB = Angiotensin-II receptor blockers.

*other antihypertensives comprise: Ambrisentan, Betanidine, Bosentan, Clonidine,Debrisoquine, Doxazosin,Guanethidine, Hydralazine, Indoramin, Ketanserin, Macitentan, Methoserpidine ,Methyldopa, Methyldopate, Metirosine, Minoxidil, Moxonidine, Prazosin, Reserpine, Riociguat, Sitaxentan, Sodium nitroprusside and Trimetaphan camsilate

Table S3b: One-year IRs – excluding one-year prevalence of the different antihypertensives classes for the frailty cohort

| Drug name | Frailty cohort N=110225 | | |
|---|---|---|---|
| | N incidence | PY | IR (95% CI) |
| **ACE-I + CCB** | 5 | 101713.0 | 0.1 (0.0, 0.1) |
| **ACE-I + Diuretics** | 29 | 101161.0 | 0.3 (0.2, 0.4) |
| **ARB + CCB** | 19 | 101717.8 | 0.2 (0.1, 0.3) |
| **ARB + Diuretics** | 32 | 101448.5 | 0.3 (0.2, 0.3) |
| **Betablockers + CCB** | <5 | NA | NA |
| **Betablockers + Diuretics** | 8 | 101159.6 | 0.1 (0.0, 0.1) |
| **Renin-Inhibitors** | 43 | 101577.2 | 0.4 (0.3, 0.6) |
| **ACE-I** | 3400 | 55998.0 | 60.7 (58.7, 62.8) |
| **ARB** | 1716 | 80840.6 | 21.2 (20.2, 22.2) |
| **All antihypertensives** | 2400 | 15449.0 | 155.4 (149.1, 161.6) |
| **Other antihypertensives*** | 1291 | 90359.1 | 14.3 (13.5, 15.1) |
| **Betablockers** | 2933 | 68671.5 | 42.7 (41.2, 44.3) |
| **CCB** | 3334 | 62256.8 | 53.6 (51.7, 55.4) |
| **Diuretics** | 5102 | 48913.8 | 104.3 (101.4, 107.2) |

PY=person years
Note: ACE-I = Angiotensin-converting-enzyme inhibitor, CCB = calcium channel blockers, ARB = Angiotensin-II receptor blockers.
*other antihypertensives comprise: Ambrisentan, Betanidine, Bosentan, Clonidine,Debrisoquine, Doxazosin,Guanethidine, Hydralazine, Indoramin, Ketanserin, Macitentan, Methoserpidine ,Methyldopa, Methyldopate, Metirosine, Minoxidil, Moxonidine, Prazosin, Reserpine, Riociguat, Sitaxentan, Sodium nitroprusside and Trimetaphan camsilate

Table S3c: One-year IRs – excluding one-year prevalence of the different antihypertensives classes for the polypharmacy cohort

| Drug name | Polypharmacy cohort N=116076 | | |
|---|---|---|---|
| | N incidence | PY | IR (95% CI) |
| ACE-I + CCB | 5 | 107295.7 | 0.05 (0.01, 0.09) |
| ACE-I + Diuretics | 40 | 106684.8 | 0.38 (0.26, 0.49) |
| ARB + CCB | 16 | 107286.8 | 0.15 (0.08, 0.22) |
| ARB + Diuretics | 39 | 106932.9 | 0.37 (0.25, 0.48) |
| Betablockers + CCB | <5 | NA | NA |
| Betablockers + Diuretics | 12 | 106732.5 | 0.11 (0.05, 0.18) |
| Renin-Inhibitors | 53 | 107091.2 | 0.50 (0.36, 0.63) |
| ACE-I | 3339 | 58366.1 | 57.21 (55.27, 59.15) |
| ARB | 1936 | 82641.2 | 23.43 (22.38, 24.47) |
| All antihypertensives | 1972 | 12759.6 | 154.55 (147.73, 161.37) |
| Other antihypertensives* | 1454 | 93430.1 | 15.56 (14.76, 16.36) |
| Betablockers | 2894 | 68095.8 | 42.50 (40.95, 44.05) |
| CCB | 3368 | 61092.4 | 55.13 (53.27, 56.99) |
| Diuretics | 4983 | 45487.4 | 109.55 (106.51, 112.59) |

PY=person years
Note: ACE-I = Angiotensin-converting-enzyme inhibitor, CCB = calcium channel blockers, ARB = Angiotensin-II receptor blockers.
*other antihypertensives comprise: Ambrisentan, Betanidine, Bosentan, Clonidine,Debrisoquine, Doxazosin,Guanethidine, Hydralazine, Indoramin, Ketanserin, Macitentan, Methoserpidine ,Methyldopa, Methyldopate, Metirosine, Minoxidil, Moxonidine, Prazosin, Reserpine, Riociguat, Sitaxentan, Sodium nitroprusside and Trimetaphan camsilate

Table S3d: One-year IRs – excluding one-year prevalence of the different antihypertensives classes for the overlap group

| **Drug name** | **Overlap group N=28259** | | |
|---|---|---|---|
| | **N incidence** | **PY** | **IR (95% CI)** |
| **ACE-I + CCB** | 0 | NA | NA |
| **ACE-I + Diuretics** | 5 | 24729.1 | 0.20 (0.03, 0.38) |
| **ARB + CCB** | <5 | NA | NA |
| **ARB + Diuretics** | 7 | 24802.3 | 0.28 (0.07, 0.49) |
| **Betablockers + CCB** | 0 | NA | NA |
| **Betablockers + Diuretics** | <5 | NA | NA |
| **Renin-Inhibitors** | 9 | 24814.1 | 0.36 (0.13, 0.6) |
| **ACE-I** | 866 | 12665.3 | 68.38 (63.82, 72.9) |
| **ARB** | 472 | 19174.3 | 24.62 (22.40, 26.84) |
| **All antihypertensives** | 491 | 2481.9 | 197.83 (180.33, 215.33) |
| **Other antihypertensives*** | 315 | 21729.2 | 14.50 (12.90, 16.10) |
| **Betablockers** | 782 | 15105.2 | 51.77 (48.14, 55.40) |
| **CCB** | 828 | 14269.9 | 58.03 (54.08, 61.98) |
| **Diuretics** | 1381 | 9117.8 | 151.46 (143.47, 159.45) |

PY=person years
Note: ACE-I = Angiotensin-converting-enzyme inhibitor, CCB = calcium channel blockers, ARB = Angiotensin-II receptor blockers.
*other antihypertensives comprise: Ambrisentan, Betanidine, Bosentan, Clonidine,Debrisoquine, Doxazosin,Guanethidine, Hydralazine, Indoramin, Ketanserin, Macitentan, Methoserpidine ,Methyldopa, Methyldopate, Metirosine, Minoxidil, Moxonidine, Prazosin, Reserpine, Riociguat, Sitaxentan, Sodium nitroprusside and Trimetaphan camsilate

Table S3e: One-year IRs – excluding one-year prevalence of the different antihypertensives classes for the background population

| Drug name | Background population N=277332 | | |
|---|---|---|---|
| | N incidence | PY | IR (95% CI) |
| **ACE-I + CCB** | 5 | 265656.0 | 0.02 (0, 0.04) |
| **ACE-I + Diuretics** | 79 | 264560.6 | 0.30 (0.23, 0.36) |
| **ARB + CCB** | 25 | 265671.2 | 0.09 (0.06, 0.13) |
| **ARB + Diuretics** | 33 | 265244.8 | 0.12 (0.08, 0.17) |
| **Betablockers + CCB** | <5 | NA | NA |
| **Betablockers + Diuretics** | 24 | 263954.9 | 0.09 (0.06, 0.13) |
| **Renin-Inhibitors** | 45 | 265589.5 | 0.17 (0.12, 0.22) |
| **ACE-I** | 7348 | 206580.2 | 35.57 (34.76, 36.38) |
| **ARB** | 2400 | 242782.7 | 9.89 (9.49, 10.28) |
| **All antihypertensives** | 9622 | 131709.3 | 73.06 (71.60, 74.52) |
| **Other antihypertensives*** | 1553 | 254571.4 | 6.10 (5.80, 6.40) |
| **Betablockers** | 5137 | 221275.9 | 23.22 (22.58, 23.85) |
| **CCB** | 7555 | 208727.9 | 36.20 (35.38, 37.01) |
| **Diuretics** | 8059 | 202584.7 | 39.78 (38.91, 40.65) |

PY=person years
Note: ACE-I = Angiotensin-converting-enzyme inhibitor, CCB = calcium channel blockers, ARB = Angiotensin-II receptor blockers.
*other antihypertensives comprise: Ambrisentan, Betanidine, Bosentan, Clonidine,Debrisoquine, Doxazosin,Guanethidine, Hydralazine, Indoramin, Ketanserin, Macitentan, Methoserpidine ,Methyldopa, Methyldopate, Metirosine, Minoxidil, Moxonidine, Prazosin, Reserpine, Riociguat, Sitaxentan, Sodium nitroprusside and Trimetaphan camsilate

#### Code lists

| Product code | dmd code | Drug substance name | Product name |
|---|---|---|---|
| **ANTIHYPERTENSIVES** | | | |
| **ACE-inhibitors** | | | |
| 4103 | 318925003 | Trandolapril | Trandolapril 1mg capsules |
| 5047 | 318926002 | Trandolapril | Trandolapril 2mg capsules |
| 7419 | 318924004 | Trandolapril | Trandolapril 500microgram capsules |
| 8025 | 346811000001101 | Trandolapril | Gopten 1mg capsules (Abbott Laboratories Ltd) |
| 8026 | 273111000001109 | Trandolapril | Gopten 2mg capsules (Abbott Laboratories Ltd) |
| 9948 | 410958005 | Trandolapril | Trandolapril 4mg capsules |
| 16710 | 253511000001102 | Trandolapril | Gopten 500microgram capsules (Abbott Laboratories Ltd) |
| 28902 | 140511000001108 | Trandolapril | Odrik 2mg capsules (Aventis Pharma) |
| 29130 | 5651611000001103 | Trandolapril | Gopten 4mg capsules (Abbott Laboratories Ltd) |
| 31307 | 227511000001106 | Trandolapril | Odrik 500microgram capsules (Aventis Pharma) |
| 31810 | 432911000001102 | Trandolapril | Odrik 1mg capsules (Aventis Pharma) |
| 54345 | 15162111000001105 | Trandolapril | Trandolapril 4mg capsules (Arrow Generics Ltd) |
| 60757 | 13433411000001109 | Trandolapril | Trandolapril 500microgram capsules (Teva UK Ltd) |
| 65389 | 5372611000001104 | Trandolapril | Gopten 500microgram capsules (Waymade Healthcare Plc) |
| 65570 | 13494811000001100 | Trandolapril | Trandolapril 4mg capsules (Teva UK Ltd) |
| 66623 | 13470311000001106 | Trandolapril | Trandolapril 2mg capsules (A A H Pharmaceuticals Ltd) |
| 74209 | 14407611000001105 | Trandolapril | Trandolapril 1mg capsules (Actavis UK Ltd) |
| 80 | 318902004 | Ramipril | Ramipril 5mg capsules |
| 82 | 318906001 | Ramipril | Ramipril 10mg capsules |
| 147 | 318900007 | Ramipril | Ramipril 1.25mg capsules |
| 654 | 212915001000027107 | Ramipril | Ramipril 2.5/ 5mg/ 10mg capsule |
| 709 | 318901006 | Ramipril | Ramipril 2.5mg capsules |
| 756 | 408052000 | Ramipril | Ramipril 10mg tablets |
| 761 | 408040007 | Ramipril | Ramipril 1.25mg tablets |
| 5275 | 835411000001105 | Ramipril | Tritace 2.5mg capsules (Sanofi) |
| 5735 | 802311000001101 | Ramipril | Tritace 5mg capsules (Sanofi) |
| 6261 | 5010511000001106 | Ramipril | Tritace 1.25mg tablets (Sanofi) |
| 6288 | 408051007 | Ramipril | Ramipril 5mg tablets |
| 6314 | 408050008 | Ramipril | Ramipril 2.5mg tablets |
| 6362 | 5011111000001108 | Ramipril | Tritace 5mg tablets (Sanofi) |
| 6364 | 5010811000001109 | Ramipril | Tritace 2.5mg tablets (Sanofi) |
| 9646 | 111611000001109 | Ramipril | Tritace 1.25mg capsules (Aventis Pharma) |
| 9693 | 43711000001100 | Ramipril | Tritace 10mg capsules (Sanofi) |

| | | | |
|---|---|---|---|
| 9915 | 5011411000001103 | Ramipril | Tritace 10mg tablets (Sanofi) |
| 11937 | 8720711000001102 | Ramipril | Ramipril 2.5mg/ 5ml oral suspension |
| 28586 | 7948911000001100 | Ramipril | Lopace 5mg capsules (Discovery Pharmaceuticals) |
| 29627 | 7948711000001102 | Ramipril | Lopace 2.5mg capsules (Discovery Pharmaceuticals) |
| 32857 | 5587511000001109 | Ramipril | Ramipril 1.25mg capsules (Teva UK Ltd) |
| 32934 | 7949111000001105 | Ramipril | Lopace 10mg capsules (Discovery Pharmaceuticals) |
| 33811 | 7817811000001108 | Ramipril | Ramipril 2.5mg capsules (Ranbaxy (UK) Ltd) |
| 33894 | 5589711000001107 | Ramipril | Ramipril 10mg capsules (Teva UK Ltd) |
| 34357 | 5629611000001101 | Ramipril | Ramipril 10mg capsules (Genus Pharmaceuticals Ltd) |
| 34382 | 5631411000001102 | Ramipril | Ramipril 5mg capsules (Zentiva) |
| 34390 | 5629311000001106 | Ramipril | Ramipril 5mg capsules (Genus Pharmaceuticals Ltd) |
| 34412 | 5588511000001108 | Ramipril | Ramipril 5mg capsules (Teva UK Ltd) |
| 34429 | 7433211000001103 | Ramipril | Ramipril 5mg capsules (Mylan) |
| 34431 | 5631211000001101 | Ramipril | Ramipril 2.5mg capsules (Zentiva) |
| 34432 | 5628711000001101 | Ramipril | Ramipril 2.5mg capsules (Genus Pharmaceuticals Ltd) |
| 34490 | 5588011000001100 | Ramipril | Ramipril 2.5mg capsules (Teva UK Ltd) |
| 34505 | 5623511000001102 | Ramipril | Ramipril 2.5mg capsules (Sandoz Ltd) |
| 34528 | 5875911000001104 | Ramipril | Ramipril 2.5mg capsules (A A H Pharmaceuticals Ltd) |
| 34539 | 5624411000001103 | Ramipril | Ramipril 5mg capsules (Sandoz Ltd) |
| 34540 | 5878511000001103 | Ramipril | Ramipril 5mg capsules (A A H Pharmaceuticals Ltd) |
| 34567 | 7433011000001108 | Ramipril | Ramipril 2.5mg capsules (Mylan) |
| 34583 | 147765001000027101 | Ramipril | Ramipril 10mg Capsule (Dexcel-Pharma Ltd) |
| 34589 | 147725001000027109 | Ramipril | Ramipril 5mg Capsule (Dexcel-Pharma Ltd) |
| 34651 | 7433511000001100 | Ramipril | Ramipril 10mg capsules (Mylan) |
| 34652 | 147155001000027104 | Ramipril | Ramipril 5mg Capsule (Sovereign Medical Ltd) |
| 34657 | 5631611000001104 | Ramipril | Ramipril 10mg capsules (Zentiva) |
| 34698 | 7338611000001103 | Ramipril | Ramipril 1.25mg capsules (Zentiva) |
| 34710 | 5624611000001100 | Ramipril | Ramipril 10mg capsules (Sandoz Ltd) |
| 34732 | 147685001000027107 | Ramipril | Ramipril 2.5mg Capsule (Dexcel-Pharma Ltd) |
| 34877 | 147215001000027107 | Ramipril | Ramipril 10mg Capsule (Sovereign Medical Ltd) |
| 34893 | 153475001000027100 | Ramipril | Ramipril 10mg Capsule (IVAX Pharmaceuticals UK Ltd) |
| 34943 | 5880511000001109 | Ramipril | Ramipril 10mg capsules (A A H Pharmaceuticals Ltd) |
| 35007 | 8720511000001107 | Ramipril | Ramipril 10mg/ 5ml oral suspension |
| 38308 | 251535001000027101 | Ramipril | Ramipril 2.5/ 5mg/ 10mg tablet |
| 39355 | 237055001000027109 | Ramipril | Tritace 10mg Tablet (Sterwin Medicines) |
| 40384 | 5885911000001106 | Ramipril | Ramipril 10mg tablets (A A H Pharmaceuticals Ltd) |
| 42081 | 237025001000027108 | Ramipril | Tritace 1.25mg Tablet (Sterwin Medicines) |
| 45264 | 8266111000001102 | Ramipril | Ramipril 1.25mg capsules (Actavis UK Ltd) |
| 45340 | 157665001000027100 | Ramipril | Ramipril 10mg Capsule (Actavis UK Ltd) |
| 45554 | 8720811000001105 | Ramipril | Ramipril 5mg/ 5ml oral solution |
| 46890 | 8720911000001100 | Ramipril | Ramipril 5mg/ 5ml oral suspension |
| 47021 | 19877111000001100 | Ramipril | Ramipril 2.5mg/ 5ml oral solution sugar free |
| 47998 | 8266411000001107 | Ramipril | Ramipril 2.5mg capsules (Actavis UK Ltd) |

| | | | |
|---|---|---|---|
| 48008 | 8267111000001104 | Ramipril | Ramipril 5mg capsules (Actavis UK Ltd) |
| 48053 | 9805611000001106 | Ramipril | Ramipril 2.5mg capsules (Almus Pharmaceuticals Ltd) |
| 49164 | 8267511000001108 | Ramipril | Ramipril 10mg capsules (Actavis UK Ltd) |
| 50509 | 8720411000001108 | Ramipril | Ramipril 10mg/ 5ml oral solution |
| 51701 | 16065011000001105 | Ramipril | Ramipril 5mg capsules (Bristol Laboratories Ltd) |
| 51714 | 5921211000001103 | Ramipril | Ramipril 2.5mg capsules (Alliance Healthcare (Distribution) Ltd) |
| 52197 | 15183911000001103 | Ramipril | Ramipril 5mg capsules (Sigma Pharmaceuticals Plc) |
| 52399 | 5632211000001108 | Ramipril | Ramipril 1.25mg capsules (Kent Pharmaceuticals Ltd) |
| 52407 | 5632811000001109 | Ramipril | Ramipril 10mg capsules (Kent Pharmaceuticals Ltd) |
| 53612 | 5937511000001108 | Ramipril | Ramipril 10mg tablets (Alliance Healthcare (Distribution) Ltd) |
| 53621 | 16064811000001100 | Ramipril | Ramipril 2.5mg capsules (Bristol Laboratories Ltd) |
| 54298 | 10412211000001105 | Ramipril | Ramipril 2.5mg capsules (Arrow Generics Ltd) |
| 54620 | 15183111000001101 | Ramipril | Ramipril 2.5mg capsules (Sigma Pharmaceuticals Plc) |
| 54941 | 5923011000001109 | Ramipril | Ramipril 5mg capsules (Alliance Healthcare (Distribution) Ltd) |
| 55299 | 5873711000001102 | Ramipril | Ramipril 1.25mg capsules (A A H Pharmaceuticals Ltd) |
| 55798 | 21878111000001106 | Ramipril | Ramipril 5mg capsules (Waymade Healthcare Plc) |
| 56013 | 21877911000001108 | Ramipril | Ramipril 2.5mg capsules (Waymade Healthcare Plc) |
| 56038 | 20005711000001100 | Ramipril | Ramipril 10mg tablets (Pfizer Ltd) |
| 56129 | 5632611000001105 | Ramipril | Ramipril 5mg capsules (Kent Pharmaceuticals Ltd) |
| 56148 | 7562211000001109 | Ramipril | Ramipril 1.25mg tablets (Kent Pharmaceuticals Ltd) |
| 56169 | 10412711000001103 | Ramipril | Ramipril 10mg capsules (Arrow Generics Ltd) |
| 56356 | 5925111000001106 | Ramipril | Ramipril 10mg capsules (Alliance Healthcare (Distribution) Ltd) |
| 56704 | 5919811000001106 | Ramipril | Ramipril 1.25mg capsules (Alliance Healthcare (Distribution) Ltd) |
| 56763 | 17921311000001109 | Ramipril | Ramipril 10mg capsules (Phoenix Healthcare Distribution Ltd) |
| 56855 | 15182211000001101 | Ramipril | Ramipril 10mg capsules (Sigma Pharmaceuticals Plc) |
| 57073 | 21877711000001106 | Ramipril | Ramipril 1.25mg capsules (Waymade Healthcare Plc) |
| 57235 | 11533411000001106 | Ramipril | Ramipril 1.25mg tablets (Sandoz Ltd) |
| 57346 | 21878311000001108 | Ramipril | Ramipril 10mg capsules (Waymade Healthcare Plc) |
| 57658 | 5881111000001106 | Ramipril | Ramipril 1.25mg tablets (A A H Pharmaceuticals Ltd) |
| 57864 | 15184311000001102 | Ramipril | Ramipril 5mg tablets (Sigma Pharmaceuticals Plc) |
| 59557 | 5632411000001107 | Ramipril | Ramipril 2.5mg capsules (Kent Pharmaceuticals Ltd) |
| 59603 | 17920911000001103 | Ramipril | Ramipril 2.5mg capsules (Phoenix Healthcare Distribution Ltd) |
| 59788 | 16065411000001101 | Ramipril | Ramipril 10mg capsules (Bristol Laboratories Ltd) |
| 60730 | 17921111000001107 | Ramipril | Ramipril 5mg capsules (Phoenix Healthcare Distribution Ltd) |
| 61067 | 11407911000001108 | Ramipril | Ramipril 5mg capsules (Almus Pharmaceuticals Ltd) |
| 61339 | 11408211000001100 | Ramipril | Ramipril 10mg capsules (Almus Pharmaceuticals Ltd) |
| 61499 | 10720911000001101 | Ramipril | Ramipril 2.5mg tablets (Actavis UK Ltd) |
| 61694 | 7339211000001105 | Ramipril | Ramipril 5mg tablets (Zentiva) |
| 61985 | 5590011000001101 | Ramipril | Ramipril 1.25mg tablets (Teva UK Ltd) |
| 62036 | 21879211000001105 | Ramipril | Ramipril 5mg tablets (Waymade Healthcare Plc) |
| 62039 | 7338811000001104 | Ramipril | Ramipril 1.25mg tablets (Zentiva) |
| 62918 | 8720611000001106 | Ramipril | Ramipril 2.5mg/ 5ml oral solution |
| 62958 | 5591311000001108 | Ramipril | Ramipril 5mg tablets (Teva UK Ltd) |

| | | | |
|---|---|---|---|
| 63010 | 17922811000001109 | Ramipril | Ramipril 10mg tablets (Phoenix Healthcare Distribution Ltd) |
| 63442 | 5590811000001107 | Ramipril | Ramipril 2.5mg tablets (Teva UK Ltd) |
| 64055 | 21878511000001102 | Ramipril | Ramipril 2.5mg/ 5ml oral solution sugar free (Waymade Healthcare Plc) |
| 65443 | 147275001000027104 | Ramipril | Ramipril 1.25mg Tablet (Sovereign Medical Ltd) |
| 65599 | 588521000001102 | Ramipril | Ramipril 5mg tablets (A A H Pharmaceuticals Ltd) |
| 65749 | 28780011000001109 | Ramipril | Ramipril 5mg capsules (Ennogen Pharma Ltd) |
| 65936 | 30087011000001109 | Ramipril | Ramipril 5mg capsules (DE Pharmaceuticals) |
| 66162 | 559191000001109 | Ramipril | Ramipril 10mg tablets (Teva UK Ltd) |
| 66329 | 244065001000027109 | Ramipril | Ramipril oral solution |
| 66669 | 30087411000001100 | Ramipril | Ramipril 10mg capsules (DE Pharmaceuticals) |
| 67719 | 30866711000001105 | Ramipril | Ramipril 5mg capsules (Mawdsley-Brooks & Company Ltd) |
| 67741 | 9806011000001108 | Ramipril | Ramipril 1.25mg capsules (Almus Pharmaceuticals Ltd) |
| 68192 | 32497211000001100 | Ramipril | Ramipril 5mg capsules (Brown & Burk UK Ltd) |
| 68372 | 10721111000001105 | Ramipril | Ramipril 5mg tablets (Actavis UK Ltd) |
| 68480 | 30867211000001101 | Ramipril | Ramipril 10mg capsules (Mawdsley-Brooks & Company Ltd) |
| 69288 | 32497411000001101 | Ramipril | Ramipril 10mg capsules (Brown & Burk UK Ltd) |
| 70072 | 30865311000001101 | Ramipril | Ramipril 1.25mg capsules (Mawdsley-Brooks & Company Ltd) |
| 70709 | 10721411000001100 | Ramipril | Ramipril 10mg tablets (Actavis UK Ltd) |
| 71025 | 19834511000001104 | Ramipril | Ramipril 1.25mg tablets (APC Pharmaceuticals & Chemicals (Europe) Ltd) |
| 71040 | 20005511000001105 | Ramipril | Ramipril 5mg tablets (Pfizer Ltd) |
| 71068 | 19834711000001109 | Ramipril | Ramipril 2.5mg tablets (APC Pharmaceuticals & Chemicals (Europe) Ltd) |
| 71491 | 28779811000001103 | Ramipril | Ramipril 2.5mg capsules (Ennogen Pharma Ltd) |
| 72341 | 34747411000001106 | Ramipril | Ramipril 2.5mg capsules (Wockhardt UK Ltd) |
| 72842 | 34747811000001108 | Ramipril | Ramipril 10mg capsules (Wockhardt UK Ltd) |
| 73459 | 20007111000001105 | Ramipril | Ramipril 2.5mg/ 5ml oral solution sugar free (A A H Pharmaceuticals Ltd) |
| 73484 | 237045001000027106 | Ramipril | Tritace 5mg Tablet (Sterwin Medicines) |
| 74066 | 13762311000001106 | Ramipril | Ramipril 10mg tablets (Tillomed Laboratories Ltd) |
| 74618 | 30866311000001106 | Ramipril | Ramipril 2.5mg capsules (Mawdsley-Brooks & Company Ltd) |
| 74632 | 147375001000027107 | Ramipril | Ramipril 5mg Tablet (Sovereign Medical Ltd) |
| 75410 | | Ramipril | Ramipril 2.5mg capsules (Brown & Burk UK Ltd) |
| 76548 | | Ramipril | Ramipril 1.25mg capsules (Phoenix Healthcare Distribution Ltd) |
| 76567 | | Ramipril | Ramipril 1.25mg tablets (Sigma Pharmaceuticals Plc) |
| 76784 | | Ramipril | Ramipril 5mg capsules (Wockhardt UK Ltd) |
| 77129 | | Ramipril | Ramipril 10mg capsules (Ennogen Pharma Ltd) |
| 77486 | | Ramipril | Ramipril 2.5mg tablets (Alliance Healthcare (Distribution) Ltd) |
| 77615 | | Ramipril | Ramipril 1.25mg/ 5ml oral solution |
| 3929 | 318886000 | Quinapril | Quinapril 10mg tablets |
| 5159 | 318887009 | Quinapril | Quinapril 20mg tablets |
| 6765 | 318885001 | Quinapril | Quinapril 5mg tablets |
| 7314 | 582611000001106 | Quinapril | Accupro 5mg tablets (Pfizer Ltd) |
| 9731 | 318894007 | Quinapril | Quinapril 40mg tablets |
| 14477 | 231111000001106 | Quinapril | Accupro 10mg tablets (Pfizer Ltd) |
| 14478 | 829111000001100 | Quinapril | Accupro 20mg tablets (Pfizer Ltd) |

| | | | |
|---|---|---|---|
| 15096 | 86411000001109 | Quinapril | Accupro 40mg tablets (Pfizer Ltd) |
| 38854 | 252775001000027101 | Quinapril | Quinapril 20mg/ 5ml oral solution |
| 38899 | 9208111000001100 | Quinapril | Quinil 10mg tablets (Tillomed Laboratories Ltd) |
| 40355 | 9207711000001100 | Quinapril | Quinil 5mg tablets (Tillomed Laboratories Ltd) |
| 42285 | 9208711000001104 | Quinapril | Quinil 40mg tablets (Tillomed Laboratories Ltd) |
| 46365 | 9208411000001105 | Quinapril | Quinil 20mg tablets (Tillomed Laboratories Ltd) |
| 61292 | 7392611000001105 | Quinapril | Quinapril 40mg tablets (Mylan) |
| 77116 | | Quinapril | Quinapril 5mg tablets (Alliance Healthcare (Distribution) Ltd) |
| 56079 | 21939811000001102 | Perindopril tosilate | Perindopril tosilate 10mg tablets |
| 57333 | 21940111000001100 | Perindopril tosilate | Perindopril tosilate 5mg tablets |
| 57944 | 21939911000001107 | Perindopril tosilate | Perindopril tosilate 2.5mg tablets |
| 97 | 318897000 | Perindopril erbumine | Perindopril erbumine 4mg tablets |
| 593 | 318896009 | Perindopril erbumine | Perindopril erbumine 2mg tablets |
| 5612 | 48211000001104 | Perindopril erbumine | Coversyl 2mg tablets (Servier Laboratories Ltd) |
| 5800 | 902211000001100 | Perindopril erbumine | Coversyl 4mg tablets (Servier Laboratories Ltd) |
| 6078 | 374667004 | Perindopril erbumine | Perindopril erbumine 8mg tablets |
| 11983 | 8671311000001103 | Perindopril erbumine | Perindopril erbumine 4mg/ 5ml oral suspension |
| 14960 | 3803711000001101 | Perindopril erbumine | Coversyl 8mg tablets (Servier Laboratories Ltd) |
| 33095 | 10828711000001103 | Perindopril erbumine | Perindopril erbumine 4mg tablets (A A H Pharmaceuticals Ltd) |
| 35731 | 10829111000001106 | Perindopril erbumine | Perindopril erbumine 8mg tablets (A A H Pharmaceuticals Ltd) |
| 38285 | 11879911000001102 | Perindopril erbumine | Perindopril erbumine 4mg tablets (Teva UK Ltd) |
| 38510 | 13767811000001104 | Perindopril erbumine | Perindopril erbumine 4mg tablets (Apotex UK Ltd) |
| 43012 | 244345001000027102 | Perindopril Erbumine | Perindopril erbumine oral solution |
| 43813 | 12498511000001100 | Perindopril erbumine | Perindopril erbumine 2mg tablets (Actavis UK Ltd) |
| 45319 | 10828511000001108 | Perindopril erbumine | Perindopril erbumine 2mg tablets (A A H Pharmaceuticals Ltd) |
| 45938 | 11880111000001100 | Perindopril erbumine | Perindopril erbumine 8mg tablets (Teva UK Ltd) |
| 48049 | 16631411000001104 | Perindopril erbumine | Perindopril erbumine 2mg tablets (Mylan) |
| 48180 | 13652011000001107 | Perindopril erbumine | Perindopril erbumine 4mg tablets (Sandoz Ltd) |
| 48214 | 12498711000001105 | Perindopril erbumine | Perindopril erbumine 4mg tablets (Actavis UK Ltd) |
| 49491 | 14126011000001109 | Perindopril erbumine | Perindopril erbumine 2mg tablets (Consilient Health Ltd) |
| 50402 | 193395001000027104 | Perindopril erbumine | Perindopril 2mg Tablet (Servier Laboratories Ltd) |
| 53058 | 20319211000001109 | Perindopril erbumine | Perindopril erbumine 8mg tablets (Sandoz Ltd) |
| 54733 | 17218111000001108 | Perindopril erbumine | Perindopril erbumine 8mg tablets (Consilient Health Ltd) |
| 54899 | 11879711000001104 | Perindopril erbumine | Perindopril erbumine 2mg tablets (Teva UK Ltd) |
| 54942 | 16631011000001108 | Perindopril erbumine | Perindopril erbumine 8mg tablets (Mylan) |
| 54986 | 14057411000001109 | Perindopril erbumine | Perindopril erbumine 8mg/ 5ml oral suspension |
| 56162 | 14126311000001107 | Perindopril erbumine | Perindopril erbumine 4mg tablets (Consilient Health Ltd) |
| 56472 | 10743211000001103 | Perindopril erbumine | Perindopril erbumine 4mg tablets (Kent Pharmaceuticals Ltd) |
| 56473 | 15156311000001100 | Perindopril erbumine | Perindopril erbumine 2mg tablets (Sigma Pharmaceuticals Plc) |
| 56506 | 5535211000001106 | Perindopril erbumine | Coversyl 2mg tablets (Dowelhurst Ltd) |
| 56508 | 5538211000001104 | Perindopril erbumine | Coversyl 4mg tablets (Dowelhurst Ltd) |
| 56516 | 13651711000001102 | Perindopril erbumine | Perindopril erbumine 2mg tablets (Sandoz Ltd) |
| 57701 | 12498911000001107 | Perindopril erbumine | Perindopril erbumine 8mg tablets (Actavis UK Ltd) |

| | | | |
|---|---|---|---|
| 57801 | 15639211000001104 | Perindopril erbumine | Perindopril erbumine 4mg tablets (Glenmark Pharmaceuticals Europe Ltd) |
| 58843 | 10742711000001102 | Perindopril erbumine | Perindopril erbumine 2mg tablets (Kent Pharmaceuticals Ltd) |
| 58874 | 18426311000001107 | Perindopril erbumine | Perindopril erbumine 2mg tablets (Somex Pharma) |
| 59770 | 22378411000001100 | Perindopril erbumine | Perindopril erbumine 4mg tablets (Aurobindo Pharma Ltd) |
| 59790 | 20168911000001102 | Perindopril erbumine | Perindopril erbumine 8mg tablets (Accord Healthcare Ltd) |
| 59972 | 12060711000001101 | Perindopril erbumine | Perindopril erbumine 2mg tablets (Alliance Healthcare (Distribution) Ltd) |
| 60065 | 15156711000001101 | Perindopril erbumine | Perindopril erbumine 4mg tablets (Sigma Pharmaceuticals Plc) |
| 61117 | 23471511000001106 | Perindopril erbumine | Perindopril erbumine 4mg/ 5ml oral solution |
| 61270 | 21296211000001103 | Perindopril erbumine | Perindopril erbumine 4mg tablets (Accord Healthcare Ltd) |
| 61693 | 22378211000001104 | Perindopril erbumine | Perindopril erbumine 8mg tablets (Aurobindo Pharma Ltd) |
| 64602 | 21845111000001102 | Perindopril erbumine | Perindopril erbumine 2mg tablets (Waymade Healthcare Plc) |
| 65273 | 16631211000001103 | Perindopril erbumine | Perindopril erbumine 4mg tablets (Mylan) |
| 66060 | 30075311000001109 | Perindopril erbumine | Perindopril erbumine 2mg tablets (DE Pharmaceuticals) |
| 67269 | 5352011000001109 | Perindopril erbumine | Coversyl 2mg tablets (Waymade Healthcare Plc) |
| 67789 | 21295411000001108 | Perindopril erbumine | Perindopril erbumine 2mg tablets (Accord Healthcare Ltd) |
| 68021 | 12061211000001102 | Perindopril erbumine | Perindopril erbumine 4mg tablets (Alliance Healthcare (Distribution) Ltd) |
| 68381 | 30856611000001105 | Perindopril erbumine | Perindopril erbumine 4mg tablets (Mawdsley-Brooks & Company Ltd) |
| 68759 | 22378611000001102 | Perindopril erbumine | Perindopril erbumine 2mg tablets (Aurobindo Pharma Ltd) |
| 69016 | 30075711000001108 | Perindopril erbumine | Perindopril erbumine 8mg tablets (DE Pharmaceuticals) |
| 70916 | 15639611000001102 | Perindopril erbumine | Perindopril erbumine 8mg tablets (Glenmark Pharmaceuticals Europe Ltd) |
| 70917 | 15639011000001109 | Perindopril erbumine | Perindopril erbumine 2mg tablets (Glenmark Pharmaceuticals Europe Ltd) |
| 71004 | 21296511000001100 | Perindopril erbumine | Perindopril erbumine 8mg tablets (Accord Healthcare Ltd) |
| 72295 | 196145001000027107 | Perindopril erbumine | Perindopril 2mg Tablet (Neo Laboratories Ltd) |
| 72941 | 13011311000001104 | Perindopril erbumine | Perindopril erbumine 1mg/ 5ml oral suspension |
| 75021 | 16213911000001100 | Perindopril erbumine | Perindopril erbumine 4mg tablets (Ranbaxy (UK) Ltd) |
| 75024 | 17955711000001106 | Perindopril erbumine | Perindopril erbumine 4mg tablets (Phoenix Healthcare Distribution Ltd) |
| 75847 | | Perindopril erbumine | Perindopril erbumine 8mg/ 5ml oral solution |
| 77665 | | Perindopril erbumine | Perindopril erbumine 8mg tablets (Somex Pharma) |
| 37930 | 13454411000001108 | Perindopril arginine | Perindopril arginine 5mg tablets |
| 37964 | 13454211000001109 | Perindopril arginine | Perindopril arginine 2.5mg tablets |
| 37965 | 13444911000001103 | Perindopril arginine | Coversyl Arginine 5mg tablets (Servier Laboratories Ltd) |
| 37971 | 13454111000001103 | Perindopril arginine | Perindopril arginine 10mg tablets |
| 38026 | 13444611000001109 | Perindopril arginine | Coversyl Arginine 10mg tablets (Servier Laboratories Ltd) |
| 38034 | 13445211000001108 | Perindopril arginine | Coversyl Arginine 2.5mg tablets (Servier Laboratories Ltd) |
| 50347 | 15416811000001102 | Perindopril arginine | Coversyl Arginine 5mg tablets (Waymade Healthcare Plc) |
| 51807 | 19720911000001102 | Perindopril arginine | Coversyl Arginine 5mg tablets (DE Pharmaceuticals) |
| 15121 | 318934008 | Moexipril | Moexipril 7.5mg tablets |
| 17120 | 318935009 | Moexipril | Moexipril 15mg tablets |
| 28724 | 4041111000001109 | Moexipril | Perdix 7.5mg tablets (UCB Pharma Ltd) |
| 28725 | 4040311000001101 | Moexipril | Perdix 15mg tablets (UCB Pharma Ltd) |
| 65 | 318859000 | Lisinopril | Lisinopril 10mg tablets |
| 69 | 318860005 | Lisinopril | Lisinopril 20mg tablets |
| 78 | 318858008 | Lisinopril | Lisinopril 5mg tablets |

| | | | |
|---|---|---|---|
| 277 | 318857003 | Lisinopril | Lisinopril 2.5mg tablets |
| 3720 | 593111000001108 | Lisinopril | Zestril 2.5mg tablets (AstraZeneca UK Ltd) |
| 6806 | 825311000001100 | Lisinopril | Zestril 10mg tablets (AstraZeneca UK Ltd) |
| 6807 | 823211000001109 | Lisinopril | Zestril 5mg tablets (AstraZeneca UK Ltd) |
| 8268 | 891711000001107 | Lisinopril | Zestril 20mg tablets (AstraZeneca UK Ltd) |
| 10882 | 321011000001103 | Lisinopril | Carace 2.5mg tablets (Bristol-Myers Squibb Pharmaceuticals Ltd) |
| 11987 | 8622511000001100 | Lisinopril | Lisinopril 5mg/ 5ml oral solution |
| 12313 | 56711000001109 | Lisinopril | Carace 20mg tablets (Bristol-Myers Squibb Pharmaceuticals Ltd) |
| 14387 | 778011000001109 | Lisinopril | Carace 5mg tablets (Bristol-Myers Squibb Pharmaceuticals Ltd) |
| 16701 | 315211000001104 | Lisinopril | Carace 10mg tablets (Bristol-Myers Squibb Pharmaceuticals Ltd) |
| 19198 | 487411000001102 | Lisinopril | Lisinopril 20mg tablets (Teva UK Ltd) |
| 19204 | 528611000001100 | Lisinopril | Lisinopril 5mg tablets (Teva UK Ltd) |
| 19223 | 640511000001107 | Lisinopril | Lisinopril 10mg tablets (Teva UK Ltd) |
| 20975 | 8622911000001107 | Lisinopril | Lisinopril 7.5mg/ 5ml oral suspension |
| 30921 | 60211000001105 | Lisinopril | Lisinopril 2.5mg tablets (Teva UK Ltd) |
| 32597 | 145811000001100 | Lisinopril | Lisinopril 10mg tablets (Sandoz Ltd) |
| 33977 | 656411000001108 | Lisinopril | Lisinopril 10mg tablets (Mylan) |
| 34471 | 35611000001109 | Lisinopril | Lisinopril 5mg tablets (Mylan) |
| 34696 | 576511000001101 | Lisinopril | Lisinopril 20mg tablets (Mylan) |
| 34799 | 360711000001100 | Lisinopril | Lisinopril 20mg tablets (Zentiva) |
| 37778 | 8622611000001101 | Lisinopril | Lisinopril 5mg/ 5ml oral suspension |
| 41522 | 7315011000001107 | Lisinopril | Lisopress 20mg tablets (Teva UK Ltd) |
| 41532 | 7314511000001100 | Lisinopril | Lisopress 5mg tablets (Teva UK Ltd) |
| 41538 | 7314011000001108 | Lisinopril | Lisopress 2.5mg tablets (Teva UK Ltd) |
| 41573 | 7314811000001102 | Lisinopril | Lisopress 10mg tablets (Teva UK Ltd) |
| 43412 | 874311000001100 | Lisinopril | Lisinopril 2.5mg tablets (A A H Pharmaceuticals Ltd) |
| 43413 | 668811000001102 | Lisinopril | Lisinopril 20mg tablets (A A H Pharmaceuticals Ltd) |
| 43416 | 532711000001109 | Lisinopril | Lisinopril 10mg tablets (A A H Pharmaceuticals Ltd) |
| 43418 | 18111000001103 | Lisinopril | Lisinopril 5mg tablets (A A H Pharmaceuticals Ltd) |
| 43566 | 877611000001104 | Lisinopril | Lisinopril 2.5mg tablets (Sandoz Ltd) |
| 45300 | 247511000001109 | Lisinopril | Lisinopril 10mg tablets (Actavis UK Ltd) |
| 45324 | 460111000001109 | Lisinopril | Lisinopril 20mg tablets (Actavis UK Ltd) |
| 45337 | 34511000001109 | Lisinopril | Lisinopril 5mg tablets (Actavis UK Ltd) |
| 45816 | 9810211000001103 | Lisinopril | Lisinopril 5mg tablets (Almus Pharmaceuticals Ltd) |
| 46975 | 683211000001108 | Lisinopril | Lisinopril 5mg tablets (Sandoz Ltd) |
| 46979 | 637111000001101 | Lisinopril | Lisinopril 20mg tablets (Sandoz Ltd) |
| 47159 | 9810411000001104 | Lisinopril | Lisinopril 10mg tablets (Almus Pharmaceuticals Ltd) |
| 51433 | 13754411000001103 | Lisinopril | Lisinopril 20mg tablets (Tillomed Laboratories Ltd) |
| 52088 | 17937211000001108 | Lisinopril | Lisinopril 5mg tablets (Phoenix Healthcare Distribution Ltd) |
| 53271 | 912911000001107 | Lisinopril | Lisinopril 10mg tablets (Alliance Healthcare (Distribution) Ltd) |
| 53551 | 17937611000001105 | Lisinopril | Lisinopril 20mg tablets (Phoenix Healthcare Distribution Ltd) |
| 53820 | 10386011000001109 | Lisinopril | Lisinopril 5mg tablets (Arrow Generics Ltd) |
| 54037 | 10290011000001104 | Lisinopril | Lisinopril 10mg tablets (Relonchem Ltd) |

| | | | |
|---|---|---|---|
| 54283 | 8594211000001106 | Lisinopril | Lisinopril 5mg/ 5ml oral suspension (Special Order) |
| 54288 | 10386411000001100 | Lisinopril | Lisinopril 10mg tablets (Arrow Generics Ltd) |
| 54512 | 243735001000027100 | Lisinopril | Lisinopril Oral solution |
| 54928 | 16060611000001106 | Lisinopril | Lisinopril 10mg tablets (Bristol Laboratories Ltd) |
| 55002 | 18463211000001101 | Lisinopril | Lisinopril 20mg tablets (Accord Healthcare Ltd) |
| 55456 | 72111000001106 | Lisinopril | Lisinopril 5mg tablets (Alliance Healthcare (Distribution) Ltd) |
| 55588 | 15107811000001101 | Lisinopril | Lisinopril 20mg tablets (Sigma Pharmaceuticals Plc) |
| 55639 | 18463011000001106 | Lisinopril | Lisinopril 10mg tablets (Accord Healthcare Ltd) |
| 55896 | 460011000001108 | Lisinopril | Lisinopril 2.5mg tablets (Actavis UK Ltd) |
| 56279 | 8622111000001109 | Lisinopril | Lisinopril 2.5mg/ 5ml oral solution |
| 56505 | 16450011000001100 | Lisinopril | Zestril 5mg tablets (Lexon (UK) Ltd) |
| 56510 | 14767811000001102 | Lisinopril | Zestril 20mg tablets (Sigma Pharmaceuticals Plc) |
| 57048 | 499711000001101 | Lisinopril | Lisinopril 10mg tablets (Zentiva) |
| 57588 | 17480011000001107 | Lisinopril | Zestril 2.5mg tablets (Mawdsley-Brooks & Company Ltd) |
| 58258 | 8622211000001103 | Lisinopril | Lisinopril 2.5mg/ 5ml oral suspension |
| 58294 | 18463411000001102 | Lisinopril | Lisinopril 5mg tablets (Accord Healthcare Ltd) |
| 58451 | 9809811000001108 | Lisinopril | Lisinopril 2.5mg tablets (Almus Pharmaceuticals Ltd) |
| 58461 | 709811000001102 | Lisinopril | Lisinopril 2.5mg tablets (Kent Pharmaceuticals Ltd) |
| 58682 | 5253711000001106 | Lisinopril | Lisinopril 2.5mg tablets (Mylan) |
| 58863 | 17937411000001107 | Lisinopril | Lisinopril 10mg tablets (Phoenix Healthcare Distribution Ltd) |
| 58871 | 22072611000001100 | Lisinopril | Lisinopril 10mg tablets (Waymade Healthcare Plc) |
| 59109 | 13754011000001107 | Lisinopril | Lisinopril 5mg tablets (Tillomed Laboratories Ltd) |
| 59111 | 580411000001107 | Lisinopril | Lisinopril 20mg tablets (Alliance Healthcare (Distribution) Ltd) |
| 60010 | 832711000001108 | Lisinopril | Lisinopril 10mg tablets (Kent Pharmaceuticals Ltd) |
| 60097 | 461611000001106 | Lisinopril | Lisinopril 2.5mg tablets (Zentiva) |
| 60232 | 574111000001103 | Lisinopril | Lisinopril 5mg tablets (Zentiva) |
| 60309 | 10291711000001105 | Lisinopril | Lisinopril 5mg tablets (Relonchem Ltd) |
| 61262 | 16060811000001105 | Lisinopril | Lisinopril 20mg tablets (Bristol Laboratories Ltd) |
| 62564 | 8621911000001101 | Lisinopril | Lisinopril 10mg/ 5ml oral solution |
| 63030 | 24367411000001102 | Lisinopril | Lisinopril 10mg tablets (DE Pharmaceuticals) |
| 63559 | 607011000001108 | Lisinopril | Lisinopril 20mg tablets (Kent Pharmaceuticals Ltd) |
| 63824 | 8622011000001108 | Lisinopril | Lisinopril 10mg/ 5ml oral suspension |
| 64902 | 30251811000001106 | Lisinopril | Lisinopril 5mg/ 5ml oral solution sugar free |
| 65102 | 15107411000001103 | Lisinopril | Lisinopril 10mg tablets (Sigma Pharmaceuticals Plc) |
| 65416 | 27377411000001101 | Lisinopril | Lisinopril 5mg tablets (Lupin Healthcare (UK) Ltd) |
| 65536 | 337211000001101 | Lisinopril | Lisinopril 2.5mg tablets (Alliance Healthcare (Distribution) Ltd) |
| 65983 | 27377211000001100 | Lisinopril | Lisinopril 2.5mg tablets (Lupin Healthcare (UK) Ltd) |
| 65985 | 24368011000001107 | Lisinopril | Lisinopril 2.5mg tablets (DE Pharmaceuticals) |
| 66558 | 24368811000001101 | Lisinopril | Lisinopril 5mg tablets (DE Pharmaceuticals) |
| 66622 | 24368611000001100 | Lisinopril | Lisinopril 20mg tablets (DE Pharmaceuticals) |
| 66772 | 22072211000001102 | Lisinopril | Lisinopril 2.5mg tablets (Waymade Healthcare Plc) |
| 67075 | 30221411000001105 | Lisinopril | Lisinopril 2.5mg tablets (Mawdsley-Brooks & Company Ltd) |
| 67194 | 16060211000001109 | Lisinopril | Lisinopril 2.5mg tablets (Bristol Laboratories Ltd) |

| | | | |
|---|---|---|---|
| 67795 | 9810811000001102 | Lisinopril | Lisinopril 20mg tablets (Almus Pharmaceuticals Ltd) |
| 68094 | 15108011000001108 | Lisinopril | Lisinopril 5mg tablets (Sigma Pharmaceuticals Plc) |
| 68247 | 16060411000001108 | Lisinopril | Lisinopril 5mg tablets (Bristol Laboratories Ltd) |
| 69074 | 10291011000001108 | Lisinopril | Lisinopril 20mg tablets (Relonchem Ltd) |
| 69269 | 22072811000001101 | Lisinopril | Lisinopril 20mg tablets (Waymade Healthcare Plc) |
| 70667 | 30221611000001108 | Lisinopril | Lisinopril 5mg tablets (Mawdsley-Brooks & Company Ltd) |
| 71562 | 129245001000027105 | Lisinopril | Lisinopril 10mg Tablet (Niche Generics Ltd) |
| 72038 | 10290411000001108 | Lisinopril | Lisinopril 2.5mg tablets (Relonchem Ltd) |
| 72336 | 22072411000001103 | Lisinopril | Lisinopril 5mg tablets (Waymade Healthcare Plc) |
| 73672 | 35165411000001106 | Lisinopril | Lisinopril 10mg tablets (Aurobindo Pharma Ltd) |
| 73716 | 35391311000001107 | Lisinopril | Lisinopril 2.5mg tablets (Crescent Pharma Ltd) |
| 74155 | 35165211000001107 | Lisinopril | Lisinopril 5mg tablets (Aurobindo Pharma Ltd) |
| 76100 | | Lisinopril | Lisinopril 5mg tablets (Kent Pharmaceuticals Ltd) |
| 76128 | | Lisinopril | Lisinopril 20mg tablets (Lupin Healthcare (UK) Ltd) |
| 77378 | | Lisinopril | Zestril 10mg tablets (Dowelhurst Ltd) |
| 77401 | | Lisinopril | Zestril 2.5mg tablets (Waymade Healthcare Plc) |
| 77402 | | Lisinopril | Zestril 5mg tablets (Dowelhurst Ltd) |
| 77407 | | Lisinopril | Zestril 5mg tablets (Waymade Healthcare Plc) |
| 6408 | 797911000001103 | Imidapril | Tanatril 5mg tablets (Mitsubishi Tanabe Pharma Europe Ltd) |
| 12815 | 680811000001102 | Imidapril | Tanatril 10mg tablets (Mitsubishi Tanabe Pharma Europe Ltd) |
| 12858 | 318944005 | Imidapril | Imidapril 10mg tablets |
| 16924 | 318943004 | Imidapril | Imidapril 5mg tablets |
| 18219 | 318942009 | Imidapril | Imidapril 20mg tablets |
| 32560 | 533511000001106 | Imidapril | Tanatril 20mg tablets (Mitsubishi Tanabe Pharma Europe Ltd) |
| 633 | 318909008 | Fosinopril sodium | Fosinopril 10mg tablets |
| 4571 | 462511000001104 | Fosinopril sodium | Staril 10mg tablets (Bristol-Myers Squibb Pharmaceuticals Ltd) |
| 5861 | 318910003 | Fosinopril sodium | Fosinopril 20mg tablets |
| 13589 | 348111000001107 | Fosinopril sodium | Staril 20mg tablets (Bristol-Myers Squibb Pharmaceuticals Ltd) |
| 67307 | 5543711000001104 | Fosinopril sodium | Staril 20mg tablets (Dowelhurst Ltd) |
| 196 | 318851002 | Enalapril maleate | Enalapril 5mg tablets |
| 448 | 318850001 | Enalapril maleate | Enalapril 2.5mg tablets |
| 1299 | 318853004 | Enalapril maleate | Enalapril 10mg tablets |
| 1904 | 318855006 | Enalapril maleate | Enalapril 20mg tablets |
| 8105 | 302311000001104 | Enalapril maleate | Innovace 20mg tablets (Merck Sharp & Dohme Ltd) |
| 8106 | 749611000001107 | Enalapril maleate | Innovace 2.5mg tablets (Merck Sharp & Dohme Ltd) |
| 8800 | 730211000001104 | Enalapril maleate | Innovace 5mg tablets (Merck Sharp & Dohme Ltd) |
| 8830 | 316111000001104 | Enalapril maleate | Innovace 10mg tablets (Merck Sharp & Dohme Ltd) |
| 11197 | 233035001000027101 | Enalapril Maleate | Innovace melt 5mg Wafer (Merck Sharp & Dohme Ltd) |
| 13755 | 206205001000027109 | Enalapril Maleate | Enalapril 10mg wafer |
| 15085 | 82345001000027107 | Enalapril Maleate | Innovace Titration pack (Merck Sharp & Dohme Ltd) |
| 16708 | 98645001000027107 | Enalapril Maleate | Enalapril titration pack |
| 19208 | 676011000001102 | Enalapril maleate | Enalapril 10mg tablets (Actavis UK Ltd) |
| 20188 | 98655001000027105 | Enalapril Maleate | Enalapril 2.5mg wafer |

| | | | |
|---|---|---|---|
| 22439 | 222975001000027100 | Enalapril maleate | Ednyt 20mg Tablet (Dominion Pharma) |
| 22708 | 206195001000027109 | Enalapril Maleate | Enalapril 5mg wafer |
| 23252 | 761411000001101 | Enalapril maleate | Pralenal 10 tablets (Opus Pharmaceuticals Ltd) |
| 24041 | 206215001000027106 | Enalapril Maleate | Enalapril 20mg wafer |
| 27871 | 233045001000027100 | Enalapril Maleate | Innovace melt 10mg Wafer (Merck Sharp & Dohme Ltd) |
| 28127 | 795111000001105 | Enalapril maleate | Enalapril 2.5mg tablets (Teva UK Ltd) |
| 29530 | 233025001000027103 | Enalapril Maleate | Innovace melt 2.5mg Wafer (Merck Sharp & Dohme Ltd) |
| 31587 | 233005001000027105 | Enalapril Maleate | Innovace melt 20mg Wafer (Merck Sharp & Dohme Ltd) |
| 31716 | 665211000001105 | Enalapril maleate | Enalapril 20mg tablets (Actavis UK Ltd) |
| 32241 | 225811000001104 | Enalapril maleate | Enalapril 10mg tablets (A A H Pharmaceuticals Ltd) |
| 33057 | 195715001000027106 | Enalapril maleate | Ednyt 5mg Tablet (Dominion Pharma) |
| 33078 | 526511000001100 | Enalapril maleate | Enalapril 20mg tablets (A A H Pharmaceuticals Ltd) |
| 34400 | 61665001000027109 | Enalapril maleate | Enalapril 5mg Tablet (Dowelhurst Ltd) |
| 34453 | 923711000001107 | Enalapril maleate | Enalapril 20mg tablets (Mylan) |
| 34712 | 233511000001103 | Enalapril maleate | Enalapril 20mg tablets (Kent Pharmaceuticals Ltd) |
| 34768 | 381411000001100 | Enalapril maleate | Enalapril 20mg tablets (IVAX Pharmaceuticals UK Ltd) |
| 34798 | 728411000001104 | Enalapril maleate | Enalapril 20mg tablets (Sandoz Ltd) |
| 34952 | 401711000001103 | Enalapril maleate | Enalapril 10mg tablets (Mylan) |
| 34953 | 346311000001105 | Enalapril maleate | Enalapril 20mg tablets (Zentiva) |
| 35794 | 26111000001109 | Enalapril maleate | Enalapril 5mg tablets (A A H Pharmaceuticals Ltd) |
| 36753 | 195725001000027102 | Enalapril maleate | Ednyt 10mg Tablet (Dominion Pharma) |
| 37080 | 8486911000001103 | Enalapril maleate | Enalapril 5mg/ 5ml oral solution |
| 37087 | 8487011000001104 | Enalapril maleate | Enalapril 5mg/ 5ml oral suspension |
| 41417 | 4811000001104 | Enalapril maleate | Enalapril 2.5mg tablets (A A H Pharmaceuticals Ltd) |
| 41694 | 456011000001101 | Enalapril maleate | Enalapril 2.5mg tablets (IVAX Pharmaceuticals UK Ltd) |
| 41746 | 450811000001105 | Enalapril maleate | Enalapril 10mg tablets (Sandoz Ltd) |
| 42723 | 578511000001100 | Enalapril maleate | Pralenal 5 tablets (Opus Pharmaceuticals Ltd) |
| 42894 | 115211000001105 | Enalapril maleate | Enalapril 10mg tablets (Teva UK Ltd) |
| 42901 | 319411000001102 | Enalapril maleate | Enalapril 5mg tablets (Teva UK Ltd) |
| 42902 | 695511000001106 | Enalapril maleate | Enalapril 20mg tablets (Teva UK Ltd) |
| 42908 | 856011000001106 | Enalapril maleate | Enalapril 5mg tablets (IVAX Pharmaceuticals UK Ltd) |
| 43411 | 51511000001108 | Enalapril maleate | Enalapril 5mg tablets (Sandoz Ltd) |
| 43563 | 102611000001100 | Enalapril maleate | Enalapril 2.5mg tablets (Zentiva) |
| 44657 | 195705001000027109 | Enalapril maleate | Ednyt 2.5mg Tablet (Dominion Pharma) |
| 45217 | 50211000001104 | Enalapril maleate | Enalapril 5mg tablets (Kent Pharmaceuticals Ltd) |
| 46974 | 766411000001107 | Enalapril maleate | Enalapril 5mg tablets (Mylan) |
| 50334 | 8486811000001108 | Enalapril maleate | Enalapril 4mg/ 5ml oral suspension |
| 50780 | 12086111000001102 | Enalapril maleate | Enalapril 2mg/ 5ml oral solution |
| 50863 | 8463911000001106 | Enalapril maleate | Enalapril 5mg/ 5ml oral solution (Drug Tariff Special Order) |
| 52010 | 688811000001104 | Enalapril maleate | Enalapril 10mg tablets (Alliance Healthcare (Distribution) Ltd) |
| 52882 | 20092911000001108 | Enalapril maleate | Enalapril 5mg/ 5ml oral suspension sugar free |
| 53719 | 367611000001108 | Enalapril maleate | Enalapril 20mg tablets (Alliance Healthcare (Distribution) Ltd) |
| 53915 | 11553911000001104 | Enalapril maleate | Enalapril 5mg tablets (Dexcel-Pharma Ltd) |

| | | | |
|---|---|---|---|
| 55903 | 11554111000001100 | Enalapril maleate | Enalapril 10mg tablets (Dexcel-Pharma Ltd) |
| 57378 | 12086211000001108 | Enalapril maleate | Enalapril 2mg/ 5ml oral suspension |
| 57882 | 8489611000001101 | Enalapril maleate | Enalapril 2.5mg/ 5ml oral suspension |
| 58751 | 8485611000001104 | Enalapril maleate | Enalapril 1.25mg/ 5ml oral suspension |
| 59996 | 19191211000001101 | Enalapril maleate | Enalapril 20mg tablets (Milpharm Ltd) |
| 60143 | 19727011000001100 | Enalapril maleate | Enalapril 5mg tablets (Medreich Plc) |
| 61133 | 17901611000001106 | Enalapril maleate | Enalapril 10mg tablets (Phoenix Healthcare Distribution Ltd) |
| 62860 | 23881911000001104 | Enalapril maleate | Enalapril 5mg tablets (DE Pharmaceuticals) |
| 63322 | 9794411000001107 | Enalapril maleate | Enalapril 10mg tablets (Almus Pharmaceuticals Ltd) |
| 64062 | 12085811000001101 | Enalapril maleate | Enalapril 1mg/ 5ml oral suspension |
| 64877 | 11553711000001101 | Enalapril maleate | Enalapril 2.5mg tablets (Dexcel-Pharma Ltd) |
| 66895 | 8485811000001100 | Enalapril maleate | Enalapril 10mg/ 5ml oral suspension |
| 68496 | 127311000001103 | Enalapril maleate | Enalapril 10mg tablets (Kent Pharmaceuticals Ltd) |
| 71668 | 9794611000001105 | Enalapril maleate | Enalapril 20mg tablets (Almus Pharmaceuticals Ltd) |
| 71737 | 9793911000001101 | Enalapril maleate | Enalapril 2.5mg tablets (Almus Pharmaceuticals Ltd) |
| 72017 | 8485511000001103 | Enalapril maleate | Enalapril 1.25mg/ 5ml oral solution |
| 73389 | 23881711000001101 | Enalapril maleate | Enalapril 2.5mg tablets (DE Pharmaceuticals) |
| 73617 | 12085411000001103 | Enalapril maleate | Enalapril 1.5mg/ 5ml oral suspension |
| 74237 | 12085911000001106 | Enalapril maleate | Enalapril 25mg/ 5ml oral solution |
| 76486 | | Enalapril maleate | Enalapril 2.5mg tablets (Mylan) |
| 76619 | | Enalapril maleate | Enalapril 20mg tablets (Dexcel-Pharma Ltd) |
| 77271 | | Enalapril maleate | Enalapril 20mg Tablet (Neo Laboratories Ltd) |
| 77315 | | Enalapril maleate | Enalapril 2.5mg/ 5ml oral solution |
| 77316 | | Enalapril maleate | Enalapril 4mg/ 5ml oral solution |
| 77345 | | Enalapril maleate | Innovace 10mg tablets (Dowelhurst Ltd) |
| 77690 | | Enalapril maleate | Enalapril 10mg tablets (IVAX Pharmaceuticals UK Ltd) |
| 77831 | | Enalapril maleate | Enalapril 500micrograms/ 5ml oral solution |
| 12411 | 318915008 | Cilazapril monohydrate | Cilazapril 500microgram tablets |
| 12412 | 318917000 | Cilazapril monohydrate | Cilazapril 2.5mg tablets |
| 12574 | 318916009 | Cilazapril monohydrate | Cilazapril 1mg tablets |
| 13026 | 318923005 | Cilazapril monohydrate | Cilazapril 5mg tablets |
| 16196 | 3671311000001104 | Cilazapril monohydrate | Vascace 5mg tablets (Roche Products Ltd) |
| 16197 | 3672411000001101 | Cilazapril monohydrate | Vascace 2.5mg tablets (Roche Products Ltd) |
| 16212 | 3669711000001105 | Cilazapril monohydrate | Vascace 1mg tablets (Roche Products Ltd) |
| 21053 | 3740211000001108 | Cilazapril monohydrate | Vascace 500microgram tablets (Roche Products Ltd) |
| 15605 | 11055001000027102 | Cilazapril | Cilazapril 250micrograms tablets |
| 23642 | 20325001000027107 | Cilazapril | Vascace 0.25mg Tablet (Roche Products Ltd) |
| 1121 | 318820009 | Captopril | Captopril 12.5mg tablets |
| 1143 | 318821008 | Captopril | Captopril 25mg tablets |
| 1144 | 455611000001103 | Captopril | Capoten 25mg tablets (Bristol-Myers Squibb Pharmaceuticals Ltd) |
| 1807 | 318824000 | Captopril | Captopril 50mg tablets |
| 3069 | 517711000001103 | Captopril | Acepril 25mg tablets (Bristol-Myers Squibb Pharmaceuticals Ltd) |
| 3310 | 134511000001103 | Captopril | Capoten 12.5mg tablets (Bristol-Myers Squibb Pharmaceuticals Ltd) |

| | | | |
|---|---|---|---|
| 3839 | 386611000001109 | Captopril | Capoten 50mg tablets (Bristol-Myers Squibb Pharmaceuticals Ltd) |
| 15958 | 219545001000027103 | Captopril | Captopril 2mg tablets |
| 17624 | 8351811000001102 | Captopril | Captopril 5mg/ 5ml oral suspension |
| 17633 | 236435001000027104 | Captopril | Captopril 3mg/ 5ml oral solution |
| 18269 | 660111000001107 | Captopril | Acepril 12.5mg tablets (Bristol-Myers Squibb Pharmaceuticals Ltd) |
| 18325 | 814111000001108 | Captopril | Acepril 50mg tablets (Bristol-Myers Squibb Pharmaceuticals Ltd) |
| 20849 | 673711000001107 | Captopril | Tensopril 12.5mg tablets (Teva UK Ltd) |
| 21943 | 777711000001105 | Captopril | Kaplon 12.5mg tablets (Teva UK Ltd) |
| 23478 | 653111000001108 | Captopril | Tensopril 50mg tablets (Teva UK Ltd) |
| 24482 | 760211000001106 | Captopril | Captomex 50mg tablets (Actavis UK Ltd) |
| 25998 | 372511000001104 | Captopril | Captomex 12.5mg tablets (Actavis UK Ltd) |
| 26995 | 477111000001100 | Captopril | Kaplon 25mg tablets (Teva UK Ltd) |
| 28486 | 8352011000001100 | Captopril | Captopril 6.25mg/ 5ml oral suspension |
| 28820 | 562011000001101 | Captopril | Captomex 25mg tablets (Actavis UK Ltd) |
| 30039 | 597511000001109 | Captopril | Tensopril 25mg tablets (Teva UK Ltd) |
| 32048 | 572511000001102 | Captopril | Kaplon 50mg tablets (Teva UK Ltd) |
| 32514 | 817411000001108 | Captopril | Ecopace 25mg tablets (AMCo) |
| 33336 | 160135001000027103 | Captopril | Captopril 5mg/ 5ml Oral suspension (Eldon Laboratories) |
| 33646 | 68395001000027109 | Captopril | Captopril 12.5mg Tablet (Generics (UK) Ltd) |
| 34544 | 68435001000027105 | Captopril | Captopril 12.5mg Tablet (IVAX Pharmaceuticals UK Ltd) |
| 34562 | 68445001000027106 | Captopril | Captopril 25mg Tablet (IVAX Pharmaceuticals UK Ltd) |
| 34719 | 68415001000027104 | Captopril | Captopril 50mg Tablet (Generics (UK) Ltd) |
| 34936 | 68705001000027105 | Captopril | Captopril 25mg Tablet (Lagap) |
| 34937 | 68455001000027109 | Captopril | Captopril 50mg Tablet (IVAX Pharmaceuticals UK Ltd) |
| 35302 | 8347211000001105 | Captopril | Captopril 12.5mg/ 5ml oral suspension |
| 36742 | 248405001000027101 | Captopril | Captopril 2mg/ 5ml oral suspension |
| 37655 | 424411000001101 | Captopril | Captopril 25mg tablets (Teva UK Ltd) |
| 39512 | 256275001000027106 | Captopril | Captopril 25mg/ 5ml oral suspension |
| 41617 | 501411000001104 | Captopril | Captopril 25mg tablets (Actavis UK Ltd) |
| 41633 | 605411000001105 | Captopril | Captopril 12.5mg tablets (Actavis UK Ltd) |
| 41743 | 3911000001107 | Captopril | Captopril 50mg tablets (Teva UK Ltd) |
| 43432 | 245485001000027106 | Captopril | Captopril 6.25mg tablets |
| 43507 | 68405001000027101 | Captopril | Captopril 25mg Tablet (Generics (UK) Ltd) |
| 43649 | 303411000001102 | Captopril | Captopril 25mg tablets (A A H Pharmaceuticals Ltd) |
| 44527 | 7659911000001107 | Captopril | Captopril 5mg/ ml oral solution sugar free |
| 45228 | 243265001000027100 | Captopril | Captopril capsules |
| 46851 | 8351611000001101 | Captopril | Captopril 5mg/ 5ml oral solution |
| 46951 | 226411000001105 | Captopril | Captopril 12.5mg tablets (A A H Pharmaceuticals Ltd) |
| 46957 | 13741311000001109 | Captopril | Captopril 12.5mg tablets (Tillomed Laboratories Ltd) |
| 52293 | 8791811000001109 | Captopril | Captopril 2mg capsules |
| 52499 | 8348511000001107 | Captopril | Captopril 25mg/ 5ml oral solution |
| 54544 | 8348611000001106 | Captopril | Captopril 25mg/ 5ml oral suspension |
| 56509 | 5528711000001108 | Captopril | Capoten 12.5mg tablets (Dowelhurst Ltd) |

| | | | |
|---|---|---|---|
| 56850 | 221411000001104 | Captopril | Ecopace 12.5mg tablets (AMCo) |
| 58195 | 8347111000001104 | Captopril | Captopril 12.5mg/ 5ml oral solution |
| 59699 | 23707511000001102 | Captopril | Captopril 5mg/ 5ml oral solution sugar free |
| 59915 | 23707311000001108 | Captopril | Captopril 25mg/ 5ml oral solution sugar free |
| 60349 | 23681711000001107 | Captopril | Noyada 25mg/ 5ml oral solution (Martindale Pharmaceuticals Ltd) |
| 60823 | 23682011000001102 | Captopril | Noyada 5mg/ 5ml oral solution (Martindale Pharmaceuticals Ltd) |
| 64739 | 8317011000001108 | Captopril | Captopril 25mg/ 5ml oral solution (Special Order) |
| 66597 | 8346811000001109 | Captopril | Captopril 10mg/ 5ml oral suspension |
| 69192 | 243255001000027109 | Captopril | Captopril oral solution |
| 69599 | 8351111000001109 | Captopril | Captopril 500micrograms/ 5ml oral suspension |
| 69600 | 8347911000001101 | Captopril | Captopril 1mg/ 5ml oral suspension |
| 70994 | 654711000001106 | Captopril | Captopril 12.5mg tablets (Sandoz Ltd) |
| 71277 | 8353911000001108 | Captopril | Captopril 7.5mg/ 5ml oral suspension |
| 73659 | 291911000001104 | Captopril | Captopril 50mg tablets (Kent Pharmaceuticals Ltd) |
| 74417 | 8348311000001101 | Captopril | Captopril 20mg/ 5ml oral suspension |
| 74627 | 68765001000027106 | Captopril | Captopril 25mg Tablet (C P Pharmaceuticals Ltd) |
| 76433 | | Captopril | Captopril 1.5mg capsules |
| 77046 | | Captopril | Captopril 3mg oral powder sachets |
| 77361 | | Captopril | Capoten 25mg tablets (Waymade Healthcare Plc) |
| 77364 | | Captopril | Capoten 25mg tablets (Stephar (U.K.) Ltd) |
| 77400 | | Captopril | Capoten 25mg tablets (Mawdsley-Brooks & Company Ltd) |
| 77410 | | Captopril | Capoten 50mg tablets (Dowelhurst Ltd) |
| 77415 | | Captopril | Capoten 50mg tablets (Waymade Healthcare Plc) |
| 77597 | | Captopril | Captopril 50mg/ 5ml oral suspension |
| 217 | 118915001000027101 | | CAPTOPRIL 4 MG/ ML LIQ |
| 2927 | 98615001000027100 | | PERINDOPRIL/ TERT-BUTYLAMINE 2 MG TAB |
| 3509 | 82805001000027100 | | ENALAPRIL MALEATE 40 MG TAB |
| 6200 | 3042211000001107 | | Tritace titration pack capsules (Sanofi) |
| 8923 | 91075001000027109 | | CAPTOPRIL 100 MG TAB |
| 22004 | | | CARACE (SPECIAL COMPLIANCE PACK) |
| 22882 | | | RAMIPRIL |
| 23382 | | | CARACE (SPECIAL COMPLIANCE PACK) |
| 24214 | 178845001000027106 | | TRITACE 1.25 MG TAB |
| 24693 | | | CARACE (SPECIAL COMPLIANCE PACK) |
| 27890 | | | ENALAPRIL MALEATE |
| 29964 | 172005001000027105 | | TRITACE 2.5 MG TAB |
| 31288 | | | TRITACE |
| 39421 | 13600911000001104 | | Tritace titration pack tablets (Sanofi) |
| 63594 | 13610111000001104 | | Generic Tritace titration pack tablets |
| **ACE-inhibitors in combination with calcium channel blockers** | | | |
| 18223 | 232125001000027103 | Verapamil / Trandolapril | Trandolapril with verapamil 2mg + 180mg Modified-release capsule |

| | | | |
|---|---|---|---|
| 19690 | 36149211000001102 | Verapamil / Trandolapril | Verapamil 180mg modified-release / Trandolapril 2mg capsules |
| 20579 | 3691211000001101 | Verapamil / Trandolapril | Tarka modified-release capsules (Abbott Laboratories Ltd) |
| 60684 | 23984911000001108 | Perindopril erbumine/ Amlodipine besilate | Perindopril erbumine 4mg / Amlodipine 10mg tablets |
| 11567 | 21665500100002710 0 | Felodipine/ Ramipril | Ramipril 5mg with felodipine 5mg modified-release tablet |
| 11965 | 216645001000027103 | Felodipine/ Ramipril | Ramipril 2.5mg with felodipine 2.5mg modified-release tablet |
| 17006 | 3887911000001109 | Felodipine/ Ramipril | Triapin 5mg/ 5mg modified-release tablets (Sanofi) |
| 17474 | 318177008 | Felodipine/ Ramipril | Felodipine 5mg modified-release / Ramipril 5mg tablets |
| 21162 | 318176004 | Felodipine/ Ramipril | Felodipine 2.5mg modified-release / Ramipril 2.5mg tablets |
| 28438 | 4093211000001109 | Felodipine/ Ramipril | Triapin 2.5mg/ 2.5mg modified-release tablets (Sanofi) |
| 60067 | 23985011000001108 | Amlodipine besilate/ Perindopril erbumine | Perindopril erbumine 4mg / Amlodipine 5mg tablets |
| 60744 | 23985211000001103 | Amlodipine besilate/ Perindopril erbumine | Perindopril erbumine 8mg / Amlodipine 5mg tablets |
| 63149 | 23985111000001109 | Amlodipine besilate/ Perindopril erbumine | Perindopril erbumine 8mg / Amlodipine 10mg tablets |
| **ACE- inhibitors in combination with diuretics** | | | |
| 6794 | 3437611000001100 | Perindopril erbumine/ Indapamide | Perindopril erbumine 4mg / Indapamide 1.25mg tablets |
| 14228 | 562511000001109 | Perindopril erbumine/ Indapamide | Coversyl Plus tablets (Servier Laboratories Ltd) |
| 48098 | 263725001000027105 | Perindopril Erbumine/ Indapamide | Perindopril arginine 4mg with Indapamide 1.25mg tablet |
| 50607 | 246725001000027107 | Perindopril Erbumine/ Indapamide | Perindopril arginine 2mg with Indapamide 625 micrograms tablet |
| 37908 | 13444311000001104 | Perindopril arginine/ Indapamide | Coversyl Arginine Plus 5mg/ 1.25mg tablets (Servier Laboratories Ltd) |
| 37978 | 13454311000001101 | Perindopril arginine/ Indapamide | Perindopril arginine 5mg / Indapamide 1.25mg tablets |
| 51258 | 18571111000001108 | Perindopril arginine/ Indapamide | Coversyl Arginine Plus 5mg/ 1.25mg tablets (DE Pharmaceuticals) |
| 2982 | 27665001000027103 | Lisinopril/ Hydrochlorothiazide | Zestoretic 20- 20mg+12.5mg Tablet (AstraZeneca UK Ltd) |
| 6359 | 27675001000027105 | Lisinopril/ Hydrochlorothiazide | Zestoretic 10- 10mg+12.5mg Tablet (AstraZeneca UK Ltd) |
| 6468 | 318880006 | Lisinopril/ Hydrochlorothiazide | Lisinopril 20mg / Hydrochlorothiazide 12.5mg tablets |
| 6786 | 318884002 | Lisinopril/ Hydrochlorothiazide | Lisinopril 10mg / Hydrochlorothiazide 12.5mg tablets |
| 9764 | 178295001000027105 | Lisinopril/ Hydrochlorothiazide | Carace 20 Tablet (Bristol-Myers Squibb Pharmaceuticals Ltd) |
| 17655 | 178305001000027103 | Lisinopril/ Hydrochlorothiazide | Carace 10 Tablet (Bristol-Myers Squibb Pharmaceuticals Ltd) |
| 21231 | 7385711000001106 | Lisinopril/ Hydrochlorothiazide | Caralpha 20mg/ 12.5mg tablets (Actavis UK Ltd) |
| 33353 | 7334911000001102 | Lisinopril/ Hydrochlorothiazide | Lisinopril 20mg / Hydrochlorothiazide 12.5mg tablets (Teva UK Ltd) |
| 37710 | 7334711000001104 | Lisinopril/ Hydrochlorothiazide | Lisinopril 10mg / Hydrochlorothiazide 12.5mg tablets (Teva UK Ltd) |
| 38995 | 3143111000001100 | Lisinopril/ Hydrochlorothiazide | Zestoretic 20 tablets (AstraZeneca UK Ltd) |
| 39137 | 3144311000001101 | Lisinopril/ Hydrochlorothiazide | Zestoretic 10 tablets (AstraZeneca UK Ltd) |
| 39147 | 3143511000001109 | Lisinopril/ Hydrochlorothiazide | Carace 20 Plus tablets (Merck Sharp & Dohme Ltd) |
| 39242 | 3144511000001107 | Lisinopril/ Hydrochlorothiazide | Carace 10 Plus tablets (Merck Sharp & Dohme Ltd) |
| 54201 | 13845711000001107 | Lisinopril/ Hydrochlorothiazide | Lisinopril 20mg / Hydrochlorothiazide 12.5mg tablets (Almus Pharmaceuticals Ltd) |
| 55399 | 7495911000001104 | Lisinopril/ Hydrochlorothiazide | Lisinopril 20mg / Hydrochlorothiazide 12.5mg tablets (A A H Pharmaceuticals Ltd) |
| 56244 | 13754811000001101 | Lisinopril/ Hydrochlorothiazide | Lisinopril 20mg / Hydrochlorothiazide 12.5mg tablets (Tillomed Laboratories Ltd) |
| 57539 | 14767411000001104 | Lisinopril/ Hydrochlorothiazide | Zestoretic 10 tablets (Sigma Pharmaceuticals Plc) |
| 67767 | 13845511000001102 | Lisinopril/ Hydrochlorothiazide | Lisinopril 10mg / Hydrochlorothiazide 12.5mg tablets (Almus Pharmaceuticals Ltd) |
| 71115 | 10483611000001104 | Lisinopril/ Hydrochlorothiazide | Zestoretic 10 tablets (Waymade Healthcare Plc) |

| | | | |
|---|---|---|---|
| 74008 | 5426911000001100 | Lisinopril/ Hydrochlorothiazide | Zestoretic 20 tablets (Waymade Healthcare Plc) |
| 74040 | 16449811000001104 | Lisinopril/ Hydrochlorothiazide | Zestoretic 20 tablets (Lexon (UK) Ltd) |
| 74874 | 13983811000001102 | Lisinopril/ Hydrochlorothiazide | Zestoretic 20 tablets (DE Pharmaceuticals) |
| 56157 | 21940011000001101 | Indapamide/ Perindopril tosilate | Perindopril tosilate 5mg / Indapamide 1.25mg tablets |
| 15031 | 260211000001104 | Hydrochlorothiazide/ Quinapril | Accuretic 12.5mg/ 10mg tablets (Pfizer Ltd) |
| 15108 | 318892006 | Hydrochlorothiazide/ Quinapril | Quinapril 10mg / Hydrochlorothiazide 12.5mg tablets |
| 3203 | 152555001000027101 | Hydrochlorothiazide/ Captopril | Capozide LS Tablet (E R Squibb and Sons Ltd) |
| 11561 | 318806002 | Hydrochlorothiazide/ Captopril | Co-zidocapt 12.5mg/ 25mg tablets |
| 39227 | 546711000001107 | Hydrochlorothiazide/ Captopril | Capozide LS 12.5mg/ 25mg tablets (Bristol-Myers Squibb Pharmaceuticals Ltd) |
| 1021 | 146811000001108 | Enalapril maleate/ Hydrochlorothiazide | Innozide 20mg/ 12.5mg tablets (Merck Sharp & Dohme Ltd) |
| 5189 | 318849001 | Enalapril maleate/ Hydrochlorothiazide | Enalapril 20mg / Hydrochlorothiazide 12.5mg tablets |
| 76920 | | Enalapril maleate/ Hydrochlorothiazide | Enalapril 20mg / Hydrochlorothiazide 12.5mg tablets (Tillomed Laboratories Ltd) |
| 1520 | 17311000001102 | Captopril/ Hydrochlorothiazide | Capozide 25mg/ 50mg tablets (Bristol-Myers Squibb Pharmaceuticals Ltd) |
| 10902 | 152515001000027106 | Captopril/ Hydrochlorothiazide | Captopril 50mg with Hydrochlorothiazide 25mg tablets |
| 11133 | 159305001000027103 | Captopril/ Hydrochlorothiazide | Hydrochlorothiazide with captopril 25mg with 50mg Tablet |
| 11351 | 318807006 | Captopril/ Hydrochlorothiazide | Co-zidocapt 25mg/ 50mg tablets |
| 11641 | 152525001000027102 | Captopril/ Hydrochlorothiazide | Captopril 25mg with Hydrochlorothiazide 12.5mg tablets |
| 15135 | 159315001000027101 | Captopril/ Hydrochlorothiazide | Hydrochlorothiazide with captopril 12.5mg with 25mg Tablet |
| 18263 | 263311000001101 | Captopril/ Hydrochlorothiazide | Acezide 25mg/ 50mg tablets (Bristol-Myers Squibb Pharmaceuticals Ltd) |
| 32166 | 214775001000027102 | Captopril/ Hydrochlorothiazide | Capto-co 25mg+50mg Tablet (IVAX Pharmaceuticals UK Ltd) |
| 77457 | | Captopril/ Hydrochlorothiazide | Capozide 25mg/ 50mg tablets (Dowelhurst Ltd) |
| **Angiotensin II receptor blocker** | | | |
| 575 | 318961008 | Valsartan | Valsartan 40mg capsules |
| 3222 | 318962001 | Valsartan | Valsartan 80mg capsules |
| 4645 | 318963006 | Valsartan | Valsartan 160mg capsules |
| 6518 | 117011000001107 | Valsartan | Diovan 160mg capsules (Novartis Pharmaceuticals UK Ltd) |
| 11251 | 777611000001101 | Valsartan | Diovan 40mg capsules (Novartis Pharmaceuticals UK Ltd) |
| 11252 | 554511000001105 | Valsartan | Diovan 80mg capsules (Novartis Pharmaceuticals UK Ltd) |
| 14943 | 416515008 | Valsartan | Valsartan 40mg tablets |
| 24359 | 8263211000001101 | Valsartan | Diovan 40mg tablets (Novartis Pharmaceuticals UK Ltd) |
| 37573 | 376487009 | Valsartan | Valsartan 320mg tablets |
| 38395 | 375034009 | Valsartan | Valsartan 80mg tablets |
| 39199 | 13143311000001102 | Valsartan | Diovan 320mg tablets (Novartis Pharmaceuticals UK Ltd) |
| 44778 | 375035005 | Valsartan | Valsartan 160mg tablets |
| 45600 | 261655001000027107 | Valsartan | Diovan 160mg Tablet (Novartis Pharmaceuticals UK Ltd) |
| 53833 | 19827711000001100 | Valsartan | Valsartan 160mg capsules (Mylan) |
| 54726 | 19631111000001104 | Valsartan | Valsartan 40mg capsules (Teva UK Ltd) |
| 55187 | 20021711000001108 | Valsartan | Valsartan 160mg capsules (Arrow Generics Ltd) |
| 55821 | 19630911000001108 | Valsartan | Valsartan 160mg capsules (Teva UK Ltd) |
| 58669 | 19631111000001104 | Valsartan | Valsartan 40mg capsules (Teva UK Ltd) |

| | | | |
|---|---|---|---|
| 58910 | 14690811000001105 | Valsartan | Valsartan 80mg capsules (Sigma Pharmaceuticals Plc) |
| 59029 | 20007411000001100 | Valsartan | Valsartan 3mg/ ml oral solution |
| 59448 | 19803311000001101 | Valsartan | Valsartan 80mg capsules (A A H Pharmaceuticals Ltd) |
| 60076 | 21923911000001107 | Valsartan | Valsartan 160mg capsules (Waymade Healthcare Plc) |
| 61442 | 19630911000001108 | Valsartan | Valsartan 160mg capsules (Teva UK Ltd) |
| 67663 | 19683111000001105 | Valsartan | Valsartan 160mg capsules (Dexcel-Pharma Ltd) |
| 68948 | 19688111000001101 | Valsartan | Valsartan 160mg capsules (Actavis UK Ltd) |
| 70628 | 20001711000001102 | Valsartan | Diovan 3mg/ 1ml oral solution (Novartis Pharmaceuticals UK Ltd) |
| 71028 | 20021411000001102 | Valsartan | Valsartan 80mg capsules (Arrow Generics Ltd) |
| 74013 | 13866611000001102 | Valsartan | Diovan 160mg capsules (DE Pharmaceuticals) |
| 74055 | 15775511000001107 | Valsartan | Valsartan 40mg/ 5ml oral solution |
| 74057 | 15775611000001106 | Valsartan | Valsartan 40mg/ 5ml oral suspension |
| 75409 | | Valsartan | Valsartan 80mg capsules (Teva UK Ltd) |
| 5988 | 318986004 | Telmisartan | Telmisartan 40mg tablets |
| 6243 | 134463001 | Telmisartan | Telmisartan 20mg tablets |
| 12874 | 318987008 | Telmisartan | Telmisartan 80mg tablets |
| 13821 | 924911000001106 | Telmisartan | Micardis 40mg tablets (Boehringer Ingelheim Ltd) |
| 17545 | 527411000001102 | Telmisartan | Micardis 80mg tablets (Boehringer Ingelheim Ltd) |
| 17686 | 648711000001100 | Telmisartan | Micardis 20mg tablets (Boehringer Ingelheim Ltd) |
| 61177 | 14625011000001101 | Telmisartan | Telmisartan 20mg tablets (Sigma Pharmaceuticals Plc) |
| 65274 | 23669311000001109 | Telmisartan | Telmisartan 40mg tablets (Actavis UK Ltd) |
| 70251 | 23613311000001107 | Telmisartan | Telmisartan 20mg tablets (Teva UK Ltd) |
| 6217 | 408055003 | Olmesartan medoxomil | Olmesartan medoxomil 10mg tablets |
| 6285 | 385542009 | Olmesartan medoxomil | Olmesartan medoxomil 20mg tablets |
| 6351 | 385543004 | Olmesartan medoxomil | Olmesartan medoxomil 40mg tablets |
| 14983 | 4624011000001101 | Olmesartan medoxomil | Olmetec 10mg tablets (Daiichi Sankyo UK Ltd) |
| 18910 | 4624311000001103 | Olmesartan medoxomil | Olmetec 20mg tablets (Daiichi Sankyo UK Ltd) |
| 20117 | 4624611000001108 | Olmesartan medoxomil | Olmetec 40mg tablets (Daiichi Sankyo UK Ltd) |
| 39786 | 14680711000001103 | Olmesartan medoxomil | Olmesartan medoxomil 10mg/ 5ml oral suspension |
| 520 | 318955005 | Losartan potassium | Losartan 25mg tablets |
| 624 | 407784004 | Losartan potassium | Losartan 100mg tablets |
| 1780 | 318956006 | Losartan potassium | Losartan 50mg tablets |
| 4226 | 266511000001104 | Losartan potassium | Cozaar 25mg tablets (Merck Sharp & Dohme Ltd) |
| 5723 | 53611000001106 | Losartan potassium | Cozaar 50mg tablets (Merck Sharp & Dohme Ltd) |
| 14965 | 245811000001102 | Losartan potassium | Cozaar 100mg tablets (Merck Sharp & Dohme Ltd) |
| 39944 | 15148111000001100 | Losartan potassium | Losartan 12.5mg tablets |
| 40571 | 15138911000001101 | Losartan potassium | Cozaar 12.5mg tablets (Merck Sharp & Dohme Ltd) |
| 40711 | 15507411000001105 | Losartan potassium | Losartan 2.5mg/ ml oral suspension sugar free |
| 41232 | 15506811000001105 | Losartan potassium | Cozaar 2.5mg/ ml oral suspension (Merck Sharp & Dohme Ltd) |
| 47006 | 17024511000001106 | Losartan potassium | Losartan 100mg tablets (Teva UK Ltd) |
| 48398 | 16998411000001102 | Losartan potassium | Losartan 25mg tablets (Dexcel-Pharma Ltd) |
| 49492 | 16971611000001102 | Losartan potassium | Losartan 25mg tablets (Mylan) |
| 49588 | 16732611000001102 | Losartan potassium | Losartan 100mg tablets (A A H Pharmaceuticals Ltd) |

| | | | |
|---|---|---|---|
| 50971 | 17015511000001104 | Losartan potassium | Losartan 25mg tablets (A A H Pharmaceuticals Ltd) |
| 51186 | 17660711000001105 | Losartan potassium | Losartan 25mg tablets (Arrow Generics Ltd) |
| 51601 | 16749111000001100 | Losartan potassium | Losartan 50mg tablets (Actavis UK Ltd) |
| 52427 | 18226311000001106 | Losartan potassium | Cozaar 100mg tablets (Necessity Supplies Ltd) |
| 52658 | 15451211000001101 | Losartan potassium | Losartan 100mg/ 5ml oral suspension |
| 52659 | 14159411000001106 | Losartan potassium | Losartan 50mg/ 5ml oral solution |
| 52886 | 17015311000001105 | Losartan potassium | Losartan 12.5mg tablets (A A H Pharmaceuticals Ltd) |
| 54049 | 18464011000001108 | Losartan potassium | Losartan 50mg tablets (Accord Healthcare Ltd) |
| 54057 | 17024311000001100 | Losartan potassium | Losartan 50mg tablets (Teva UK Ltd) |
| 54404 | 16749311000001103 | Losartan potassium | Losartan 100mg tablets (Actavis UK Ltd) |
| 54735 | 17026111000001109 | Losartan potassium | Losartan 50mg tablets (Alliance Healthcare (Distribution) Ltd) |
| 54740 | 16748711000001108 | Losartan potassium | Losartan 25mg tablets (Actavis UK Ltd) |
| 54843 | 16998911000001105 | Losartan potassium | Losartan 50mg tablets (Dexcel-Pharma Ltd) |
| 55296 | 16971811000001103 | Losartan potassium | Losartan 50mg tablets (Mylan) |
| 55446 | 21037511000001102 | Losartan potassium | Losartan 100mg tablets (Bristol Laboratories Ltd) |
| 55718 | 18140111000001105 | Losartan potassium | Losartan 25mg tablets (Phoenix Healthcare Distribution Ltd) |
| 56104 | 16732411000001100 | Losartan potassium | Losartan 50mg tablets (A A H Pharmaceuticals Ltd) |
| 56970 | 18169911000001109 | Losartan potassium | Losartan 100mg tablets (Pfizer Ltd) |
| 57028 | 16972011000001101 | Losartan potassium | Losartan 100mg tablets (Mylan) |
| 58274 | 18463811000001100 | Losartan potassium | Losartan 25mg tablets (Accord Healthcare Ltd) |
| 58649 | 21033711000001102 | Losartan potassium | Losartan 25mg tablets (Bristol Laboratories Ltd) |
| 58967 | 17025711000001102 | Losartan potassium | Losartan 12.5mg tablets (Alliance Healthcare (Distribution) Ltd) |
| 59086 | 20571411000001104 | Losartan potassium | Losartan 25mg tablets (Wockhardt UK Ltd) |
| 59271 | 20308011000001106 | Losartan potassium | Losartan 25mg tablets (Sandoz Ltd) |
| 59340 | 16998111000001107 | Losartan potassium | Losartan 12.5mg tablets (Dexcel-Pharma Ltd) |
| 59351 | 18170211000001103 | Losartan potassium | Losartan 50mg tablets (Pfizer Ltd) |
| 59750 | 22338211000001101 | Losartan potassium | Losartan 50mg tablets (Aptil Pharma Ltd) |
| 59903 | 14159511000001105 | Losartan potassium | Losartan 50mg/ 5ml oral suspension |
| 60506 | 16999411000001105 | Losartan potassium | Losartan 100mg tablets (Dexcel-Pharma Ltd) |
| 61053 | 17026311000001106 | Losartan potassium | Losartan 100mg tablets (Alliance Healthcare (Distribution) Ltd) |
| 61288 | 18463611000001104 | Losartan potassium | Losartan 100mg tablets (Accord Healthcare Ltd) |
| 61495 | 22338011000001106 | Losartan potassium | Losartan 25mg tablets (Aptil Pharma Ltd) |
| 61754 | 14204111000001106 | Losartan potassium | Losartan 25mg/ 5ml oral suspension |
| 62388 | 24372011000001100 | Losartan potassium | Losartan 12.5mg tablets (DE Pharmaceuticals) |
| 63222 | 18170411000001104 | Losartan potassium | Losartan 25mg tablets (Pfizer Ltd) |
| 63918 | 17024111000001102 | Losartan potassium | Losartan 25mg tablets (Teva UK Ltd) |
| 64888 | 29898911000001106 | Losartan potassium | Losartan 12.5mg tablets (Sigma Pharmaceuticals Plc) |
| 65094 | 20308411000001102 | Losartan potassium | Losartan 50mg tablets (Sandoz Ltd) |
| 66114 | 30811311000001102 | Losartan potassium | Losartan 12.5mg tablets (Mawdsley-Brooks & Company Ltd) |
| 66551 | 21036211000001102 | Losartan potassium | Losartan 50mg tablets (Bristol Laboratories Ltd) |
| 67902 | 20308611000001104 | Losartan potassium | Losartan 100mg tablets (Sandoz Ltd) |
| 68340 | 32661211000001105 | Losartan potassium | Losartan 12.5mg tablets (Consilient Health Ltd) |
| 68603 | 20571811000001102 | Losartan potassium | Losartan 50mg tablets (Wockhardt UK Ltd) |

| | | | |
|---|---|---|---|
| 69667 | 15451111000001107 | Losartan potassium | Losartan 100mg/ 5ml oral solution |
| 69858 | 17025911000001100 | Losartan potassium | Losartan 25mg tablets (Alliance Healthcare (Distribution) Ltd) |
| 70325 | 32617311000001107 | Losartan potassium | Losartan 50mg tablets (Almus Pharmaceuticals Ltd) |
| 70765 | 32616911000001105 | Losartan potassium | Losartan 100mg tablets (Almus Pharmaceuticals Ltd) |
| 71910 | 18494111000001106 | Losartan potassium | Losartan 50mg tablets (Necessity Supplies Ltd) |
| 74243 | 35657011000001102 | Losartan potassium | Losartan 50mg tablets (Consilient Health Ltd) |
| 74589 | 32753911000001104 | Losartan potassium | Losartan 25mg tablets (Genesis Pharmaceuticals Ltd) |
| 74904 | 24372211000001105 | Losartan potassium | Losartan 25mg tablets (DE Pharmaceuticals) |
| 77196 | | Losartan potassium | Losartan 25mg tablets (Kent Pharmaceuticals Ltd) |
| 77408 | | Losartan potassium | Cozaar 50mg tablets (Waymade Healthcare Plc) |
| 828 | 318968002 | Irbesartan | Irbesartan 75mg tablets |
| 1293 | 318969005 | Irbesartan | Irbesartan 150mg tablets |
| 2971 | 318970006 | Irbesartan | Irbesartan 300mg tablets |
| 7338 | 434511000001104 | Irbesartan | Aprovel 75mg tablets (Sanofi) |
| 9196 | 859711000001103 | Irbesartan | Aprovel 150mg tablets (Sanofi) |
| 11348 | 323211000001107 | Irbesartan | Aprovel 300mg tablets (Sanofi) |
| 36939 | 12639511000001103 | Irbesartan | Irbesartan 300mg/ 5ml oral suspension |
| 52972 | 14261011000001108 | Irbesartan | Irbesartan 300mg tablets (Sigma Pharmaceuticals Plc) |
| 55017 | 21285511000001106 | Irbesartan | Irbesartan 300mg tablets (Accord Healthcare Ltd) |
| 58108 | 21230711000001102 | Irbesartan | Irbesartan 150mg tablets (A A H Pharmaceuticals Ltd) |
| 58201 | 21219211000001108 | Irbesartan | Irbesartan 150mg tablets (Actavis UK Ltd) |
| 59393 | 21993211000001106 | Irbesartan | Irbesartan 300mg tablets (Sandoz Ltd) |
| 60597 | 21112911000001107 | Irbesartan | Irbesartan 150mg tablets (Teva UK Ltd) |
| 61781 | 21113111000001103 | Irbesartan | Irbesartan 300mg tablets (Teva UK Ltd) |
| 62415 | 21100511000001103 | Irbesartan | Irbesartan 300mg tablets (A A H Pharmaceuticals Ltd) |
| 63385 | 21522211000001102 | Irbesartan | Sabervel 75mg tablets (Aspire Pharma Ltd) |
| 63411 | 21123811000001104 | Irbesartan | Irbesartan 300mg tablets (Alliance Healthcare (Distribution) Ltd) |
| 63717 | 24177711000001109 | Irbesartan | Irbesartan 300mg tablets (DE Pharmaceuticals) |
| 65065 | 21230511000001107 | Irbesartan | Irbesartan 75mg tablets (A A H Pharmaceuticals Ltd) |
| 70431 | 27588011000001102 | Irbesartan | Irbesartan 150mg tablets (Lupin Healthcare (UK) Ltd) |
| 70955 | 194235001000027100 | Irbesartan | Irbesartan 300mg/ 5ml Oral suspension (Martindale Pharmaceuticals Ltd) |
| 71019 | 21623611000001102 | Irbesartan | Irbesartan 75mg tablets (Dr Reddy's Laboratories (UK) Ltd) |
| 71096 | 21624111000001107 | Irbesartan | Irbesartan 150mg tablets (Dr Reddy's Laboratories (UK) Ltd) |
| 71215 | 21219011000001103 | Irbesartan | Irbesartan 75mg tablets (Actavis UK Ltd) |
| 72000 | 21123311000001108 | Irbesartan | Irbesartan 150mg tablets (Alliance Healthcare (Distribution) Ltd) |
| 76202 | | Irbesartan | Irbesartan 150mg/ 5ml oral suspension |
| 76797 | | Irbesartan | Ifirmasta 75mg tablets (Consilient Health Ltd) |
| 77350 | | Irbesartan | Aprovel 150mg tablets (Dowelhurst Ltd) |
| 77443 | | Irbesartan | Aprovel 300mg tablets (Mawdsley-Brooks & Company Ltd) |
| 78012 | | Irbesartan | Ifirmasta 150mg tablets (Consilient Health Ltd) |
| 6939 | 318994006 | Eprosartan mesilate | Eprosartan 300mg tablets |
| 9745 | 401211000001105 | Eprosartan mesilate | Teveten 300mg tablets (Mylan) |
| 12836 | 318996008 | Eprosartan mesilate | Eprosartan 600mg tablets |

| | | | |
|---|---|---|---|
| 13123 | 318995007 | Eprosartan mesilate | Eprosartan 400mg tablets |
| 16285 | 151411000001103 | Eprosartan mesilate | Teveten 400mg tablets (Abbott Healthcare Products Ltd) |
| 16371 | 872011000001109 | Eprosartan mesilate | Teveten 600mg tablets (Mylan) |
| 63337 | 21229711000001107 | Eprosartan mesilate | Eprosartan 600mg tablets (A A H Pharmaceuticals Ltd) |
| 529 | 318977009 | Candesartan cilexetil | Candesartan 2mg tablets |
| 531 | 318978004 | Candesartan cilexetil | Candesartan 4mg tablets |
| 4155 | 97311000001103 | Candesartan cilexetil | Amias 2mg tablets (Takeda UK Ltd) |
| 4685 | 857411000001100 | Candesartan cilexetil | Amias 4mg tablets (Takeda UK Ltd) |
| 4741 | 318980005 | Candesartan cilexetil | Candesartan 16mg tablets |
| 4818 | 318979007 | Candesartan cilexetil | Candesartan 8mg tablets |
| 5013 | 36011000001106 | Candesartan cilexetil | Amias 8mg tablets (Takeda UK Ltd) |
| 5117 | 908511000001100 | Candesartan cilexetil | Amias 16mg tablets (Takeda UK Ltd) |
| 7043 | 376998003 | Candesartan cilexetil | Candesartan 32mg tablets |
| 31072 | 8983911000001107 | Candesartan cilexetil | Amias 32mg tablets (Takeda UK Ltd) |
| 50185 | 20476311000001109 | Candesartan cilexetil | Candesartan 8mg tablets (Teva UK Ltd) |
| 51117 | 13840411000001108 | Candesartan cilexetil | Candesartan 8mg tablets (DE Pharmaceuticals) |
| 51519 | 20495911000001109 | Candesartan cilexetil | Candesartan 8mg tablets (A A H Pharmaceuticals Ltd) |
| 51647 | 16353011000001102 | Candesartan cilexetil | Candesartan 4mg tablets (Mawdsley-Brooks & Company Ltd) |
| 52208 | 20496211000001106 | Candesartan cilexetil | Candesartan 16mg tablets (A A H Pharmaceuticals Ltd) |
| 52559 | 20913011000001106 | Candesartan cilexetil | Candesartan 8mg tablets (Zentiva) |
| 53680 | 20476511000001103 | Candesartan cilexetil | Candesartan 16mg tablets (Teva UK Ltd) |
| 53755 | 20476011000001106 | Candesartan cilexetil | Candesartan 4mg tablets (Teva UK Ltd) |
| 54326 | 20476711000001108 | Candesartan cilexetil | Candesartan 32mg tablets (Teva UK Ltd) |
| 54414 | 20530511000001105 | Candesartan cilexetil | Candesartan 16mg tablets (Consilient Health Ltd) |
| 57026 | 21813511000001100 | Candesartan cilexetil | Candesartan 8mg tablets (Waymade Healthcare Plc) |
| 57266 | 21666411000001103 | Candesartan cilexetil | Candesartan 2mg tablets (Actavis UK Ltd) |
| 57273 | 20483811000001104 | Candesartan cilexetil | Candesartan 8mg tablets (Actavis UK Ltd) |
| 57977 | 20509411000001107 | Candesartan cilexetil | Candesartan 16mg tablets (Alliance Healthcare (Distribution) Ltd) |
| 58646 | 20483511000001102 | Candesartan cilexetil | Candesartan 4mg tablets (Actavis UK Ltd) |
| 59690 | 20530311000001104 | Candesartan cilexetil | Candesartan 8mg tablets (Consilient Health Ltd) |
| 59802 | 20595811000001108 | Candesartan cilexetil | Candesartan 2mg tablets (Teva UK Ltd) |
| 62035 | 21813311000001106 | Candesartan cilexetil | Candesartan 16mg tablets (Waymade Healthcare Plc) |
| 62140 | 24505711000001100 | Candesartan cilexetil | Candesartan 4mg tablets (Sandoz Ltd) |
| 64359 | 22626711000001100 | Candesartan cilexetil | Candesartan 4mg tablets (DE Pharmaceuticals) |
| 65228 | 16353811000001108 | Candesartan cilexetil | Candesartan 16mg tablets (Mawdsley-Brooks & Company Ltd) |
| 65479 | 20493511000001101 | Candesartan cilexetil | Candesartan 2mg tablets (A A H Pharmaceuticals Ltd) |
| 66624 | 24506211000001101 | Candesartan cilexetil | Candesartan 16mg tablets (Sandoz Ltd) |
| 66958 | 24506011000001106 | Candesartan cilexetil | Candesartan 8mg tablets (Sandoz Ltd) |
| 67929 | 32554811000001106 | Candesartan cilexetil | Candesartan 16mg tablets (Tillomed Laboratories Ltd) |
| 68647 | 32746311000001107 | Candesartan cilexetil | Candesartan 8mg tablets (Genesis Pharmaceuticals Ltd) |
| 68718 | 20495511000001102 | Candesartan cilexetil | Candesartan 4mg tablets (A A H Pharmaceuticals Ltd) |
| 68751 | 32746511000001101 | Candesartan cilexetil | Candesartan 16mg tablets (Genesis Pharmaceuticals Ltd) |
| 69802 | 33582711000001106 | Candesartan cilexetil | Candesartan 4mg tablets (Mylan) |

| | | | |
|---|---|---|---|
| 70455 | 15825311000001103 | Candesartan cilexetil | Candesartan 4mg/ 5ml oral suspension |
| 70805 | 34007811000001104 | Candesartan cilexetil | Candesartan 8mg tablets (Crescent Pharma Ltd) |
| 71080 | 20912811000001108 | Candesartan cilexetil | Candesartan 4mg tablets (Zentiva) |
| 72215 | 20530011000001102 | Candesartan cilexetil | Candesartan 4mg tablets (Consilient Health Ltd) |
| 72924 | 23646611000001101 | Candesartan cilexetil | Candesartan 4mg tablets (Phoenix Healthcare Distribution Ltd) |
| 73528 | 35139711000001105 | Candesartan cilexetil | Candesartan 4mg tablets (Milpharm Ltd) |
| 73811 | 22947511000001109 | Candesartan cilexetil | Candesartan 2mg tablets (Ranbaxy (UK) Ltd) |
| 73813 | 21812911000001100 | Candesartan cilexetil | Candesartan 4mg tablets (Waymade Healthcare Plc) |
| 75407 | | Candesartan cilexetil | Candesartan 32mg tablets (Sandoz Ltd) |
| 75418 | | Candesartan cilexetil | Candesartan 16mg tablets (Mylan) |
| 76070 | | Candesartan cilexetil | Candesartan 8mg tablets (Almus Pharmaceuticals Ltd) |
| 76209 | | Candesartan cilexetil | Candesartan 8mg tablets (Mylan) |
| 77128 | | Candesartan cilexetil | Candesartan 16mg tablets (Actavis UK Ltd) |
| 51368 | 449333009 | Azilsartan medoxomil | Azilsartan medoxomil 80mg tablets |
| 51897 | 20350911000001100 | Azilsartan medoxomil | Edarbi 20mg tablets (Takeda UK Ltd) |
| 56606 | 449109006 | Azilsartan medoxomil | Azilsartan medoxomil 40mg tablets |
| 76119 | | Azilsartan medoxomil | Azilsartan medoxomil 20mg tablets |
| 76121 | | Azilsartan medoxomil | Edarbi 40mg tablets (Takeda UK Ltd) |
| 31160 | | | IRBESARTAN |
| **Angiotensin II receptor blockers in combination with calcium channel blockers** | | | |
| 35096 | 11160711000001108 | Valsartan/ Amlodipine besilate | Exforge 10mg/ 160mg tablets (Novartis Pharmaceuticals UK Ltd) |
| 35189 | 11160111000001107 | Valsartan/ Amlodipine besilate | Amlodipine 10mg / Valsartan 160mg tablets |
| 35317 | 11161811000001108 | Valsartan/ Amlodipine besilate | Exforge 5mg/ 80mg tablets (Novartis Pharmaceuticals UK Ltd) |
| 35329 | 11160311000001109 | Valsartan/ Amlodipine besilate | Amlodipine 5mg / Valsartan 80mg tablets |
| 35343 | 11160211000001101 | Valsartan/ Amlodipine besilate | Amlodipine 5mg / Valsartan 160mg tablets |
| 35697 | 11161511000001105 | Valsartan/ Amlodipine besilate | Exforge 5mg/ 160mg tablets (Novartis Pharmaceuticals UK Ltd) |
| 47727 | 18987311000001103 | Olmesartan medoxomil/ Hydrochlorothiazide/ Amlodipine besilate | Sevikar HCT 40mg/ 5mg/ 25mg tablets (Daiichi Sankyo UK Ltd) |
| 53220 | 18987611000001108 | Olmesartan medoxomil/ Hydrochlorothiazide/ Amlodipine besilate | Sevikar HCT 40mg/ 10mg/ 25mg tablets (Daiichi Sankyo UK Ltd) |
| 39984 | 15773211000001105 | Olmesartan medoxomil/ Amlodipine besilate | Sevikar 20mg/ 5mg tablets (Daiichi Sankyo UK Ltd) |
| 40316 | 429502004 | Olmesartan medoxomil/ Amlodipine besilate | Olmesartan medoxomil 20mg / Amlodipine 5mg tablets |
| 40639 | 429503009 | Olmesartan medoxomil/ Amlodipine besilate | Olmesartan medoxomil 40mg / Amlodipine 5mg tablets |
| 40668 | 429678006 | Olmesartan medoxomil/ Amlodipine besilate | Olmesartan medoxomil 40mg / Amlodipine 10mg tablets |
| 41203 | 15772611000001102 | Olmesartan medoxomil/ Amlodipine besilate | Sevikar 40mg/ 10mg tablets (Daiichi Sankyo UK Ltd) |

| | | | |
|---|---|---|---|
| 41205 | 15772911000001108 | Olmesartan medoxomil/ Amlodipine besilate | Sevikar 40mg/ 5mg tablets (Daiichi Sankyo UK Ltd) |
| 46355 | 18986411000001108 | Hydrochlorothiazide/ Olmesartan medoxomil/ Amlodipine besilate | Sevikar HCT 20mg/ 5mg/ 12.5mg tablets (Daiichi Sankyo UK Ltd) |
| 60780 | 18987811000001107 | Hydrochlorothiazide/ Olmesartan medoxomil/ Amlodipine besilate | Generic Sevikar HCT 20mg/ 5mg/ 12.5mg tablets |
| 46687 | 26494500100002710 3 | Amlodipine/ Hydrochlorothiazide/ Olmesartan Medoxomil | Olmesartan medoxomil with amlodipine and hydrochlorothiazide 20mg + 5mg + 12.5mg Tablet |
| 46715 | 264965001000027109 | Amlodipine/ Hydrochlorothiazide/ Olmesartan Medoxomil | Olmesartan medoxomil with amlodipine and hydrochlorothiazide 40mg + 10mg + 12.5mg Tablet |
| 46792 | 264955001000027100 | Amlodipine/ Hydrochlorothiazide/ Olmesartan Medoxomil | Olmesartan medoxomil with amlodipine and hydrochlorothiazide 40mg + 5mg + 12.5mg Tablet |
| 47467 | 264975001000027102 | Amlodipine/ Hydrochlorothiazide/ Olmesartan Medoxomil | Olmesartan medoxomil with amlodipine and hydrochlorothiazide 40mg + 5mg + 25mg Tablet |
| 55358 | 264985001000027107 | Amlodipine/ Hydrochlorothiazide/ Olmesartan Medoxomil | Olmesartan medoxomil with amlodipine and hydrochlorothiazide 40mg + 10mg + 25mg Tablet |
| 35173 | 246845001000027107 | Amlodipine Besilate/ Valsartan | Valsartan 160mg with amlodipine 5mg tablets |
| 35174 | 246835001000027106 | Amlodipine Besilate/ Valsartan | Valsartan 80mg with amlodipine 5mg tablets |
| 35304 | 246855001000027105 | Amlodipine Besilate/ Valsartan | Valsartan 160mg with amlodipine 10mg tablets |
| 47573 | 18986711000001102 | Amlodipine besilate/ Hydrochlorothiazide/ Olmesartan medoxomil | Sevikar HCT 40mg/ 5mg/ 12.5mg tablets (Daiichi Sankyo UK Ltd) |
| 47616 | 18987011000001101 | Amlodipine besilate/ Hydrochlorothiazide/ Olmesartan medoxomil | Sevikar HCT 40mg/ 10mg/ 12.5mg tablets (Daiichi Sankyo UK Ltd) |
| 60007 | 18987911000001102 | Amlodipine besilate/ Hydrochlorothiazide/ Olmesartan medoxomil | Generic Sevikar HCT 40mg/ 10mg/ 12.5mg tablets |
| **Angiotensin II receptor blockers and diuretics** | | | |
| 764 | 8150111000001108 | Valsartan/ Hydrochlorothiazide | Co-Diovan 80mg/ 12.5mg tablets (Novartis Pharmaceuticals UK Ltd) |
| 6877 | 7668611000001104 | Valsartan/ Hydrochlorothiazide | Co-Diovan 160mg/ 12.5mg tablets (Novartis Pharmaceuticals UK Ltd) |
| 11864 | 395521005 | Valsartan/ Hydrochlorothiazide | Valsartan 160mg / Hydrochlorothiazide 12.5mg tablets |
| 14283 | 409298002 | Valsartan/ Hydrochlorothiazide | Valsartan 160mg / Hydrochlorothiazide 25mg tablets |
| 16060 | 377488008 | Valsartan/ Hydrochlorothiazide | Valsartan 80mg / Hydrochlorothiazide 12.5mg tablets |
| 23456 | 238185001000027100 | Valsartan/ Hydrochlorothiazide | Hydrochlorothiazide with valsartan 25mg with 160mg Tablet |
| 24268 | 239025001000027101 | Valsartan/ Hydrochlorothiazide | Hydrochlorothiazide with valsartan 12.5mg with 80mg Tablet |
| 24484 | 238175001000027105 | Valsartan/ Hydrochlorothiazide | Hydrochlorothiazide with valsartan 12.5mg with 160mg Tablet |
| 25382 | 7668911000001105 | Valsartan/ Hydrochlorothiazide | Co-Diovan 160mg/ 25mg tablets (Novartis Pharmaceuticals UK Ltd) |
| 52858 | 14206911000001101 | Valsartan/ Hydrochlorothiazide | Co-Diovan 80mg/ 12.5mg tablets (Sigma Pharmaceuticals Plc) |
| 67664 | 19631911000001101 | Valsartan/ Hydrochlorothiazide | Valsartan 160mg / Hydrochlorothiazide 12.5mg tablets (Teva UK Ltd) |
| 72086 | 19688511000001105 | Valsartan/ Hydrochlorothiazide | Valsartan 160mg / Hydrochlorothiazide 12.5mg tablets (Actavis UK Ltd) |
| 14870 | 407855002 | Telmisartan/ Hydrochlorothiazide | Telmisartan 40mg / Hydrochlorothiazide 12.5mg tablets |
| 16161 | 407856001 | Telmisartan/ Hydrochlorothiazide | Telmisartan 80mg / Hydrochlorothiazide 12.5mg tablets |

| | | | |
|---|---|---|---|
| 17689 | 3806911000001105 | Telmisartan/ Hydrochlorothiazide | MicardisPlus 80mg/ 12.5mg tablets (Boehringer Ingelheim Ltd) |
| 18202 | 3806311000001109 | Telmisartan/ Hydrochlorothiazide | MicardisPlus 40mg/ 12.5mg tablets (Boehringer Ingelheim Ltd) |
| 62376 | 24412511000001109 | Telmisartan/ Hydrochlorothiazide | Actelsar HCT 80mg/ 12.5mg tablets (Actavis UK Ltd) |
| 63890 | 10540911000001100 | Telmisartan/ Hydrochlorothiazide | MicardisPlus 80mg/ 12.5mg tablets (Waymade Healthcare Plc) |
| 66997 | 29749811000001100 | Telmisartan/ Hydrochlorothiazide | MicardisPlus 40mg/ 12.5mg tablets (Waymade Healthcare Plc) |
| 76840 | | Telmisartan/ Hydrochlorothiazide | Actelsar HCT 40mg/ 12.5mg tablets (Actavis UK Ltd) |
| 43322 | 409185001 | Olmesartan medoxomil/ Hydrochlorothiazide | Olmesartan medoxomil 40mg / Hydrochlorothiazide 12.5mg tablets |
| 43915 | 17220911000001102 | Olmesartan medoxomil/ Hydrochlorothiazide | Olmetec Plus 40mg/ 12.5mg tablets (Daiichi Sankyo UK Ltd) |
| 4540 | 255911000001105 | Losartan potassium/ Hydrochlorothiazide | Cozaar-Comp 50mg/ 12.5mg tablets (Merck Sharp & Dohme Ltd) |
| 6437 | 318959004 | Losartan potassium/ Hydrochlorothiazide | Losartan 50mg / Hydrochlorothiazide 12.5mg tablets |
| 10323 | 395497004 | Losartan potassium/ Hydrochlorothiazide | Losartan 100mg / Hydrochlorothiazide 25mg tablets |
| 14738 | 209115001000027101 | Losartan Potassium/ Hydrochlorothiazide | Hydrochlorothiazide with losartan 12.5mg with 50mg Tablet |
| 21423 | 9566911000001105 | Losartan potassium/ Hydrochlorothiazide | Cozaar-Comp 100mg/ 25mg tablets (Merck Sharp & Dohme Ltd) |
| 24632 | 241785001000027103 | Losartan Potassium/ Hydrochlorothiazide | Hydrochlorothiazide with losartan 25mg with 100mg Tablet |
| 38367 | 250095001000027100 | Losartan Potassium/ Hydrochlorothiazide | Hydrochlorothiazide with losartan 12.5mg with 100mg Tablet |
| 52189 | 17015111000001108 | Losartan potassium/ Hydrochlorothiazide | Losartan 100mg / Hydrochlorothiazide 25mg tablets (A A H Pharmaceuticals Ltd) |
| 55160 | 14210911000001101 | Losartan potassium/ Hydrochlorothiazide | Cozaar-Comp 50mg/ 12.5mg tablets (Sigma Pharmaceuticals Plc) |
| 56204 | 18165111000001109 | Losartan potassium/ Hydrochlorothiazide | Losartan 50mg / Hydrochlorothiazide 12.5mg tablets (Actavis UK Ltd) |
| 56975 | 17014911000001107 | Losartan potassium/ Hydrochlorothiazide | Losartan 50mg / Hydrochlorothiazide 12.5mg tablets (A A H Pharmaceuticals Ltd) |
| 57796 | 19541911000001108 | Losartan potassium/ Hydrochlorothiazide | Cozaar-Comp 50mg/ 12.5mg tablets (DE Pharmaceuticals) |
| 62911 | 17024711000001101 | Losartan potassium/ Hydrochlorothiazide | Losartan 50mg / Hydrochlorothiazide 12.5mg tablets (Teva UK Ltd) |
| 66598 | 27475411000001100 | Losartan potassium/ Hydrochlorothiazide | Losartan 50mg / Hydrochlorothiazide 12.5mg tablets (Lupin Healthcare (UK) Ltd) |
| 70754 | 18552511000001109 | Losartan potassium/ Hydrochlorothiazide | Losartan 50mg / Hydrochlorothiazide 12.5mg tablets (Ranbaxy (UK) Ltd) |
| 71682 | 30826311000001104 | Losartan potassium/ Hydrochlorothiazide | Losartan 100mg / Hydrochlorothiazide 25mg tablets (Mawdsley-Brooks & Company Ltd) |
| 74215 | 24371611000001101 | Losartan potassium/ Hydrochlorothiazide | Losartan 50mg / Hydrochlorothiazide 12.5mg tablets (DE Pharmaceuticals) |

| | | | |
|---|---|---|---|
| 11469 | 134460003 | Irbesartan/ Hydrochlorothiazide | Irbesartan 300mg / Hydrochlorothiazide 12.5mg tablets |
| 11526 | 682711000001109 | Irbesartan/ Hydrochlorothiazide | CoAprovel 300mg/ 12.5mg tablets (Sanofi) |
| 35196 | 10968611000001106 | Irbesartan/ Hydrochlorothiazide | CoAprovel 300mg/ 25mg tablets (Sanofi) |
| 35481 | 10970311000001105 | Irbesartan/ Hydrochlorothiazide | Irbesartan 300mg / Hydrochlorothiazide 25mg tablets |
| 62337 | 23472611000001104 | Irbesartan/ Hydrochlorothiazide | Irbesartan 300mg / Hydrochlorothiazide 12.5mg tablets (Actavis UK Ltd) |
| 77641 | | Irbesartan/ Hydrochlorothiazide | CoAprovel 300mg/ 12.5mg tablets (Mawdsley-Brooks & Company Ltd) |
| 38459 | 13731911000001109 | Hydrochlorothiazide/ Telmisartan | Telmisartan 80mg / Hydrochlorothiazide 25mg tablets |
| 38889 | 13719711000001103 | Hydrochlorothiazide/ Telmisartan | MicardisPlus 80mg/ 25mg tablets (Boehringer Ingelheim Ltd) |
| 71748 | 24413611000001103 | Hydrochlorothiazide/ Telmisartan | Actelsar HCT 80mg/ 25mg tablets (Actavis UK Ltd) |
| 18200 | 409184002 | Hydrochlorothiazide/ Olmesartan medoxomil | Olmesartan medoxomil 20mg / Hydrochlorothiazide 12.5mg tablets |
| 18903 | 10270711000001105 | Hydrochlorothiazide/ Olmesartan medoxomil | Olmesartan medoxomil 20mg / Hydrochlorothiazide 25mg tablets |
| 27520 | 10261811000001100 | Hydrochlorothiazide/ Olmesartan medoxomil | Olmetec Plus 20mg/ 25mg tablets (Daiichi Sankyo UK Ltd) |
| 29634 | 10261511000001103 | Hydrochlorothiazide/ Olmesartan medoxomil | Olmetec Plus 20mg/ 12.5mg tablets (Daiichi Sankyo UK Ltd) |
| 35380 | 242645001000027102 | Hydrochlorothiazide/ Olmesartan Medoxomil | Hydrochlorothiazide with olmesartan medoxomil 12.5mg with 20mg tablet |
| 39021 | 242635001000027103 | Hydrochlorothiazide/ Olmesartan Medoxomil | Hydrochlorothiazide with olmesartan medoxomil 25mg with 20mg tablet |
| 37650 | 13112711000001103 | Hydrochlorothiazide/ Losartan potassium | Losartan 100mg / Hydrochlorothiazide 12.5mg tablets |
| 37747 | 13094111000001102 | Hydrochlorothiazide/ Losartan potassium | Cozaar-Comp 100mg/ 12.5mg tablets (Merck Sharp & Dohme Ltd) |
| 48039 | 19485511000001100 | Hydrochlorothiazide/ Losartan potassium | Losartan 100mg / Hydrochlorothiazide 12.5mg tablets (Teva UK Ltd) |
| 70922 | 18618811000001108 | Hydrochlorothiazide/ Losartan potassium | Losartan 100mg / Hydrochlorothiazide 12.5mg tablets (Phoenix Healthcare Distribution Ltd) |
| 71618 | 18276311000001107 | Hydrochlorothiazide/ Losartan potassium | Losartan 100mg / Hydrochlorothiazide 12.5mg tablets (A A H Pharmaceuticals Ltd) |
| 10316 | 792411000001108 | Hydrochlorothiazide/ Irbesartan | CoAprovel 150mg/ 12.5mg tablets (Sanofi) |
| 11448 | 134461004 | Hydrochlorothiazide/ Irbesartan | Irbesartan 150mg / Hydrochlorothiazide 12.5mg tablets |
| **Other fixed combinations of antihypertensives** | | | |
| 47727 | 18987311000001103 | Olmesartan medoxomil/ Hydrochlorothiazide/ Amlodipine besilate | Sevikar HCT 40mg/ 5mg/ 25mg tablets (Daiichi Sankyo UK Ltd) |
| 53220 | 18987611000001108 | Olmesartan medoxomil/ Hydrochlorothiazide/ Amlodipine besilate | Sevikar HCT 40mg/ 10mg/ 25mg tablets (Daiichi Sankyo UK Ltd) |
| 46355 | 18986411000001108 | Hydrochlorothiazide/ Olmesartan medoxomil/ Amlodipine besilate | Sevikar HCT 20mg/ 5mg/ 12.5mg tablets (Daiichi Sankyo UK Ltd) |

| | | | |
|---|---|---|---|
| 60780 | 18987811000001107 | Hydrochlorothiazide/ Olmesartan medoxomil/ Amlodipine besilate | Generic Sevikar HCT 20mg/ 5mg/ 12.5mg tablets |
| 18606 | 216735001000027106 | Diltiazem / Hydrochlorothiazide | Diltiazem and hydrochlorothiazide 150mg+12.5mg modified-release capsules |
| 23505 | 216725001000027109 | Diltiazem / Hydrochlorothiazide | Adizem xl plus 150mg+12.5mg Modified-release capsule (Napp Pharmaceuticals Ltd) |
| 4406 | 48465001000027107 | Benzthiazide/ methoserpidine | Decaserpyl plus Tablet (Roussel Laboratories Ltd) |
| 29696 | 124515001000027108 | Benzthiazide/ methoserpidine | Methoserpidine with benzthiazide Tablet |
| 46687 | 264945001000027103 | Amlodipine/ Hydrochlorothiazide/ Olmesartan Medoxomil | Olmesartan medoxomil with amlodipine and hydrochlorothiazide 20mg + 5mg + 12.5mg Tablet |
| 46715 | 264965001000027109 | Amlodipine/ Hydrochlorothiazide/ Olmesartan Medoxomil | Olmesartan medoxomil with amlodipine and hydrochlorothiazide 40mg + 10mg + 12.5mg Tablet |
| 46792 | 264955001000027100 | Amlodipine/ Hydrochlorothiazide/ Olmesartan Medoxomil | Olmesartan medoxomil with amlodipine and hydrochlorothiazide 40mg + 5mg + 12.5mg Tablet |
| 47467 | 264975001000027102 | Amlodipine/ Hydrochlorothiazide/ Olmesartan Medoxomil | Olmesartan medoxomil with amlodipine and hydrochlorothiazide 40mg + 5mg + 25mg Tablet |
| 55358 | 264985001000027107 | Amlodipine/ Hydrochlorothiazide/ Olmesartan Medoxomil | Olmesartan medoxomil with amlodipine and hydrochlorothiazide 40mg + 10mg + 25mg Tablet |
| 47573 | 18986711000001102 | Amlodipine besilate/ Hydrochlorothiazide/ Olmesartan medoxomil | Sevikar HCT 40mg/ 5mg/ 12.5mg tablets (Daiichi Sankyo UK Ltd) |
| 47616 | 18987011000001101 | Amlodipine besilate/ Hydrochlorothiazide/ Olmesartan medoxomil | Sevikar HCT 40mg/ 10mg/ 12.5mg tablets (Daiichi Sankyo UK Ltd) |
| 60007 | 18987911000001102 | Amlodipine besilate/ Hydrochlorothiazide/ Olmesartan medoxomil | Generic Sevikar HCT 40mg/ 10mg/ 12.5mg tablets |
| **Other antihypertensives** | | | |
| 29757 | 36143911000001108 | Trimetaphan camsilate | Trimetaphan camsilate 250mg/ 5ml solution for injection ampoules |
| 36612 | 375899001 | Sodium nitroprusside dihydrate | Sodium nitroprusside 50mg powder for solution for infusion vials |
| 37085 | 11394311000001108 | Sitaxentan sodium | Sitaxentan 100mg tablets |
| 40899 | 11392011000001104 | Sitaxentan sodium | Thelin 100mg tablets (Pfizer Ltd) |
| 64930 | 24408611000001108 | Riociguat | Riociguat 2mg tablets |
| 74720 | 24399711000001103 | Riociguat | Adempas 2mg tablets (Merck Sharp & Dohme Ltd) |
| 75040 | 24408411000001105 | Riociguat | Riociguat 1mg tablets |
| 75041 | 24408311000001103 | Riociguat | Riociguat 1.5mg tablets |
| 76902 | | Riociguat | Riociguat 500microgram tablets |
| 20656 | 76555001000027105 | Reserpine | Serpasil 250microgram Tablet (Novartis Pharmaceuticals UK Ltd) |
| 20690 | 139815001000027109 | Reserpine | Reserpine 250micrograms tablet |
| 21502 | 189705001000027101 | Reserpine | Serpasil 100microgram Tablet (Novartis Pharmaceuticals UK Ltd) |
| 22853 | 139805001000027107 | Reserpine | Reserpine 100micrograms tablet |
| 591 | 318768008 | Prazosin | Prazosin 1mg tablets |
| 726 | 318769000 | Prazosin | Prazosin 2mg tablets |
| 1292 | 347411000001101 | Prazosin | Hypovase 1mg tablets (Pfizer Ltd) |
| 1455 | 318767003 | Prazosin | Prazosin 500microgram tablets |

| | | | |
|---|---|---|---|
| 3715 | 318770004 | Prazosin | Prazosin 5mg tablets |
| 4111 | 321311000001100 | Prazosin | Hypovase 500microgram tablets (Pfizer Ltd) |
| 5183 | 150911000001104 | Prazosin | Hypovase 2mg tablets (Pfizer Ltd) |
| 8198 | 14555001000027108 | Prazosin | Hypovase 5mg Tablet (Pfizer Ltd) |
| 8863 | 15362501000027101 | Prazosin | Hypovase benign prostatic hyperplasia 1mg Tablet (Pfizer Ltd) |
| 13610 | 570011000001106 | Prazosin | Alphavase 2 tablets (Ashbourne Pharmaceuticals Ltd) |
| 19823 | 74711000001102 | Prazosin | Alphavase 5 tablets (Ashbourne Pharmaceuticals Ltd) |
| 23459 | 15365501000027100 | Prazosin | Hypovase benign prostatic hyperplasia 2mg Tablet (Pfizer Ltd) |
| 25047 | 15361501000027105 | Prazosin | Hypovase benign prostatic hyperplasia 500microgram Tablet (Pfizer Ltd) |
| 26237 | 30515001000027104 | Prazosin | Alphavase 500microgram Tablet (Ashbourne Pharmaceuticals Ltd) |
| 26238 | 934411000001100 | Prazosin | Alphavase 1 tablets (Ashbourne Pharmaceuticals Ltd) |
| 26693 | 15360501000027108 | Prazosin | Hypovase benign prostatic hyperplasia bd BD Starter pack (Pfizer Ltd) |
| 41651 | 52515001000027104 | Prazosin | Prazosin 500microgram Tablet (Approved Prescription Services Ltd) |
| 41652 | 89711000001108 | Prazosin | Prazosin 500microgram tablets (A A H Pharmaceuticals Ltd) |
| 41721 | 778311000001107 | Prazosin | Prazosin 1mg tablets (A A H Pharmaceuticals Ltd) |
| 43547 | 898711000001102 | Prazosin | Prazosin 500microgram tablets (IVAX Pharmaceuticals UK Ltd) |
| 46922 | 559411000001105 | Prazosin | Prazosin 1mg tablets (IVAX Pharmaceuticals UK Ltd) |
| 55826 | 739211000001106 | Prazosin | Prazosin 5mg tablets (A A H Pharmaceuticals Ltd) |
| 60316 | 52525001000027108 | Prazosin | Prazosin 1mg Tablet (Approved Prescription Services Ltd) |
| 4993 | 318707000 | Moxonidine | Moxonidine 200microgram tablets |
| 7174 | 318708005 | Moxonidine | Moxonidine 400microgram tablets |
| 9749 | 142811000001107 | Moxonidine | Physiotens 400microgram tablets (Mylan) |
| 9876 | 522011000001109 | Moxonidine | Physiotens 300microgram tablets (Mylan) |
| 10253 | 408604009 | Moxonidine | Moxonidine 300microgram tablets |
| 11177 | 41111000001102 | Moxonidine | Physiotens 200microgram tablets (Mylan) |
| 33322 | 8098911000001106 | Moxonidine | Moxonidine 200microgram tablets (Sandoz Ltd) |
| 40310 | 8390411000001102 | Moxonidine | Moxonidine 200microgram tablets (Teva UK Ltd) |
| 43531 | 8099411000001106 | Moxonidine | Moxonidine 400microgram tablets (Sandoz Ltd) |
| 60898 | 8171411000001109 | Moxonidine | Moxonidine 200microgram tablets (Mylan) |
| 61036 | 21407711000001105 | Moxonidine | Physiotens 300microgram tablets (Actavis UK Ltd) |
| 62853 | 8936111000001107 | Moxonidine | Moxonidine 200microgram tablets (A A H Pharmaceuticals Ltd) |
| 63938 | 8099111000001101 | Moxonidine | Moxonidine 300microgram tablets (Sandoz Ltd) |
| 67665 | 8265311000001105 | Moxonidine | Moxonidine 400microgram tablets (Actavis UK Ltd) |
| 67808 | 21406211000001100 | Moxonidine | Physiotens 200microgram tablets (Actavis UK Ltd) |
| 76479 | | Moxonidine | Moxonidine 400microgram tablets (A A H Pharmaceuticals Ltd) |
| 2967 | 318656009 | Minoxidil | Minoxidil 5mg tablets |
| 2968 | 318657000 | Minoxidil | Minoxidil 10mg tablets |
| 2970 | 318655008 | Minoxidil | Minoxidil 2.5mg tablets |
| 9463 | 3666411000001106 | Minoxidil | Loniten 5mg tablets (Pfizer Ltd) |
| 9697 | 3667011000001104 | Minoxidil | Loniten 2.5mg tablets (Pfizer Ltd) |
| 14495 | 3666711000001100 | Minoxidil | Loniten 10mg tablets (Pfizer Ltd) |
| 22454 | 16780501000027109 | Metirosine | Demser 250mg Capsule (Merck Sharp & Dohme Ltd) |
| 33788 | 16777501000027100 | Metirosine | Metirosine 250mg Capsule |

| | | | |
|---|---|---|---|
| 7416 | 51165001000027107 | Methyldopate | Aldomet 50mg/ ml Injection (Merck Sharp & Dohme Ltd) |
| 26919 | 125025001000027109 | Methyldopate | Methyldopa 50mg/ ml Injection |
| 21346 | 125115001000027106 | Methyldopa / Hydrochlorothiazide | Hydromet Tablet (MSD Thomas Morson Pharmaceuticals) |
| 28738 | 125055001000027105 | Methyldopa / Hydrochlorothiazide | Methyldopa with hydrochlorothiazide Tablet |
| 1707 | 318672001 | Methyldopa | Methyldopa 250mg tablets |
| 3049 | 318671008 | Methyldopa | Methyldopa 125mg tablets |
| 3070 | 318673006 | Methyldopa | Methyldopa 500mg tablets |
| 7626 | 655001000027106 | Methyldopa | Aldomet 250mg Tablet (Merck Sharp & Dohme Ltd) |
| 7642 | 665001000027102 | Methyldopa | Aldomet 500mg Tablet (Merck Sharp & Dohme Ltd) |
| 8033 | 645001000027108 | Methyldopa | Aldomet 125mg Tablet (Merck Sharp & Dohme Ltd) |
| 9225 | 204605001000027101 | Methyldopa | Methyldopa 250mg Capsule |
| 14390 | 51145001000027101 | Methyldopa | Aldomet 250mg/ 5ml Liquid (Merck Sharp & Dohme Ltd) |
| 18252 | 29695001000027106 | Methyldopa | Metalpha 250mg Tablet (Ashbourne Pharmaceuticals Ltd) |
| 23761 | 8667311000001100 | Methyldopa | Methyldopa 250mg/ 5ml oral suspension |
| 24196 | 54625001000027106 | Methyldopa | Dopamet 250mg Tablet (Berk Pharmaceuticals Ltd) |
| 25275 | 29705001000027102 | Methyldopa | Metalpha 500mg Tablet (Ashbourne Pharmaceuticals Ltd) |
| 25289 | 54635001000027108 | Methyldopa | Dopamet 500mg Tablet (Berk Pharmaceuticals Ltd) |
| 29570 | 54615001000027102 | Methyldopa | Dopamet 125mg Tablet (Berk Pharmaceuticals Ltd) |
| 32913 | 874511000001106 | Methyldopa | Methyldopa 250mg tablets (Actavis UK Ltd) |
| 41661 | 15965001000027104 | Methyldopa | Methyldopa 250mg Tablet (C P Pharmaceuticals Ltd) |
| 43988 | 253711000001107 | Methyldopa | Aldomet 250mg tablets (Aspen Pharma Trading Ltd) |
| 43989 | 73611000001108 | Methyldopa | Aldomet 500mg tablets (Aspen Pharma Trading Ltd) |
| 62513 | 17204211000001105 | Methyldopa | Methyldopa 250mg tablets (Sovereign Medical Ltd) |
| 71110 | 17895811000001103 | Methyldopa | Aldomet 500mg tablets (Sigma Pharmaceuticals Plc) |
| 71385 | 17895611000001102 | Methyldopa | Aldomet 250mg tablets (Sigma Pharmaceuticals Plc) |
| 72819 | 17940911000001101 | Methyldopa | Methyldopa 250mg tablets (Phoenix Healthcare Distribution Ltd) |
| 73640 | 87911000001103 | Methyldopa | Methyldopa 250mg tablets (Sandoz Ltd) |
| 77825 | | Methyldopa | Methyldopa 250mg tablets (A A H Pharmaceuticals Ltd) |
| 10713 | 6855001000027101 | Methoserpidine | Decaserpyl 5mg Tablet (Roussel Laboratories Ltd) |
| 10714 | 124465001000027102 | Methoserpidine | Methoserpidine 5mg Tablet |
| 25393 | 6865001000027105 | Methoserpidine | Decaserpyl 10mg Tablet (Roussel Laboratories Ltd) |
| 29187 | 124475001000027109 | Methoserpidine | Methoserpidine 10mg Tablet |
| 63780 | 23707811000001104 | Macitentan | Macitentan 10mg tablets |
| 54940 | 14696611000001108 | Ketanserin | Ketanserin 20mg tablets |
| 2117 | 3354611000001100 | Indoramin | Doralese Tiltab 20mg tablets (Chemidex Pharma Ltd) |
| 2816 | 318739007 | Indoramin | Indoramin 20mg tablets |
| 5815 | 318740009 | Indoramin | Indoramin 25mg tablets |
| 9019 | 118045001000027102 | Indoramin | Indoramin 50mg Tablet |
| 11394 | 2415001000027104 | Indoramin | Baratol 25mg Tablet (Shire Pharmaceuticals Ltd) |
| 16198 | 2425001000027108 | Indoramin | Baratol 50mg Tablet (Shire Pharmaceuticals Ltd) |
| 40256 | 3689111000001107 | Indoramin | Baratol 25mg tablets (Amdipharm Plc) |
| 504 | 34193811000001100 | Hydralazine | Hydralazine 20mg powder for solution for injection ampoules |
| 573 | 318649003 | Hydralazine | Hydralazine 25mg tablets |

| | | | |
|---|---|---|---|
| 1296 | 318650003 | Hydralazine | Hydralazine 50mg tablets |
| 2362 | 657011000001101 | Hydralazine | Apresoline 25mg tablets (AMCo) |
| 2680 | 1655001000027106 | Hydralazine | Apresoline 50mg Tablet (Sovereign Medical Ltd) |
| 13317 | 3925011000001100 | Hydralazine | Apresoline 20mg powder for solution for injection ampoules (AMCo) |
| 18861 | 8528611000001105 | Hydralazine | Hydralazine 10mg/ 5ml oral suspension |
| 31220 | 193711000001103 | Hydralazine | Hydralazine 25mg tablets (A A H Pharmaceuticals Ltd) |
| 41639 | 147111000001103 | Hydralazine | Hydralazine 50mg tablets (Actavis UK Ltd) |
| 43500 | 364711000001106 | Hydralazine | Hydralazine 25mg tablets (Actavis UK Ltd) |
| 59512 | 8528711000001101 | Hydralazine | Hydralazine 50mg/ 5ml oral solution |
| 61116 | 8528811000001109 | Hydralazine | Hydralazine 50mg/ 5ml oral suspension |
| 63652 | 245915001000027107 | Hydralazine | Hydralazine Tablet |
| 64253 | 12537811000001103 | Hydralazine | Hydralazine 5mg/ 5ml oral suspension |
| 70655 | 8581011000001103 | Hydralazine | Hydralazine 25mg/ 5ml oral suspension |
| 71097 | 19819011000001104 | Hydralazine | Hydralazine 50mg tablets (Almus Pharmaceuticals Ltd) |
| 71256 | 30172911000001106 | Hydralazine | Hydralazine 20mg powder for concentrate for solution for injection ampoules (AMCo) |
| 74749 | 414426001 | Hydralazine | Hydralazine 10mg tablets |
| 75259 | 12536611000001104 | Hydralazine | Hydralazine 1mg/ 5ml oral suspension |
| 7922 | 1530501000027108 | Guanethidine Monosulphate | Ismelin 10mg Tablet (Sovereign Medical Ltd) |
| 7923 | 189785001000027102 | Guanethidine Monosulphate | Guanethidine 10mg Tablet |
| 13379 | 1531501000027105 | Guanethidine Monosulphate | Ismelin 25mg Tablet (Sovereign Medical Ltd) |
| 17291 | 189795001000027103 | Guanethidine Monosulphate | Guanethidine 25mg Tablet |
| 30127 | 1532501000027101 | Guanethidine monosulfate | Ismelin 10mg/ ml Injection (Sovereign Medical Ltd) |
| 31080 | 36053311000001103 | Guanethidine monosulfate | Guanethidine 10mg/ 1ml solution for injection ampoules |
| 48189 | 4369611000001106 | Guanethidine monosulfate | Ismelin 10mg/ 1ml solution for injection ampoules (Amdipharm Plc) |
| 119 | 318781001 | Doxazosin mesilate | Doxazosin 1mg tablets |
| 493 | 318782008 | Doxazosin mesilate | Doxazosin 2mg tablets |
| 582 | 134456001 | Doxazosin mesilate | Doxazosin 4mg modified-release tablets |
| 755 | 123911000001106 | Doxazosin mesilate | Cardura XL 4mg tablets (Pfizer Ltd) |
| 1294 | 318783003 | Doxazosin mesilate | Doxazosin 4mg tablets |
| 4449 | 907711000001109 | Doxazosin mesilate | Cardura 1mg tablets (Pfizer Ltd) |
| 4802 | 41811000001109 | Doxazosin mesilate | Cardura 2mg tablets (Pfizer Ltd) |
| 5496 | 135921004 | Doxazosin mesilate | Doxazosin 8mg modified-release tablets |
| 5618 | 873411000001109 | Doxazosin mesilate | Cardura XL 8mg tablets (Pfizer Ltd) |
| 7547 | 4857711000001103 | Doxazosin mesilate | Doxadura 2mg tablets (Discovery Pharmaceuticals) |
| 7549 | 4857511000001108 | Doxazosin mesilate | Doxadura 1mg tablets (Discovery Pharmaceuticals) |
| 8086 | 159565001000027104 | Doxazosin mesilate | Cardura 4mg Tablet (Pfizer Ltd) |
| 10088 | 4858111000001103 | Doxazosin mesilate | Doxadura 4mg tablets (Discovery Pharmaceuticals) |
| 19193 | 481211000001106 | Doxazosin mesilate | Doxazosin 2mg tablets (Teva UK Ltd) |
| 19216 | 525011000001104 | Doxazosin mesilate | Doxazosin 4mg tablets (IVAX Pharmaceuticals UK Ltd) |
| 20369 | 8483411000001101 | Doxazosin mesilate | Doxazosin 1mg/ 5ml oral suspension |
| 25487 | 904511000001107 | Doxazosin mesilate | Cascor 2mg tablets (Ranbaxy (UK) Ltd) |
| 25551 | 179311000001107 | Doxazosin mesilate | Cascor 4mg tablets (Ranbaxy (UK) Ltd) |
| 33094 | 674011000001107 | Doxazosin mesilate | Doxazosin 2mg tablets (Mylan) |

| | | | |
|---|---|---|---|
| 34342 | 647711000001101 | Doxazosin mesilate | Doxazosin 1mg tablets (Teva UK Ltd) |
| 34553 | 221311000001106 | Doxazosin mesilate | Doxazosin 4mg tablets (Mylan) |
| 34601 | 565111000001107 | Doxazosin mesilate | Doxazosin 1mg tablets (Mylan) |
| 34625 | 554711000001100 | Doxazosin mesilate | Doxazosin 2mg tablets (A A H Pharmaceuticals Ltd) |
| 34715 | 228711000001100 | Doxazosin mesilate | Doxazosin 1mg tablets (A A H Pharmaceuticals Ltd) |
| 35272 | 11269911000001101 | Doxazosin mesilate | Doxadura XL 4mg tablets (Discovery Pharmaceuticals) |
| 35603 | 8483511000001102 | Doxazosin mesilate | Doxazosin 4mg/ 5ml oral suspension |
| 36023 | 244615001000027103 | Doxazosin mesilate | Cardozin xl 4mg Tablet (Hillcross Pharmaceuticals Ltd) |
| 36740 | 11098311000001104 | Doxazosin mesilate | Slocinx XL 4mg tablets (Zentiva) |
| 37243 | 248505001000027109 | Doxazosin mesilate | Cardozin xl 4mg Tablet (Teva UK Ltd) |
| 38461 | 11752411000001104 | Doxazosin mesilate | Cardozin XL 4mg tablets (Arrow Generics Ltd) |
| 40678 | 653011000001107 | Doxazosin mesilate | Doxazosin 4mg tablets (Teva UK Ltd) |
| 40891 | 840911000001105 | Doxazosin mesilate | Doxazosin 2mg tablets (IVAX Pharmaceuticals UK Ltd) |
| 41543 | 854611000001108 | Doxazosin mesilate | Doxazosin 1mg tablets (IVAX Pharmaceuticals UK Ltd) |
| 43695 | 11757711000001107 | Doxazosin mesilate | Colixil XL 4mg tablets (Sandoz Ltd) |
| 45040 | 17338211000001104 | Doxazosin mesilate | Larbex XL 4mg tablets (Teva UK Ltd) |
| 45265 | 198435001000027102 | Doxazosin mesilate | Doxazosin sr 4mg Tablet (Generics (UK) Ltd) |
| 45328 | 462611000001100 | Doxazosin mesilate | Doxazosin 1mg tablets (Sandoz Ltd) |
| 45342 | 761311000001108 | Doxazosin mesilate | Doxazosin 4mg tablets (Sandoz Ltd) |
| 45583 | 11554711000001104 | Doxazosin mesilate | Doxazosin 2mg tablets (Dexcel-Pharma Ltd) |
| 46066 | 18197411000001107 | Doxazosin mesilate | Cardozin XL 4mg tablets (Almus Pharmaceuticals Ltd) |
| 46526 | 18164911000001108 | Doxazosin mesilate | Raporsin XL 4mg tablets (Actavis UK Ltd) |
| 47807 | 203155001000027108 | Doxazosin mesilate | Doxazosin xl 4mg Tablet (Hillcross Pharmaceuticals Ltd) |
| 48150 | 4250111000001105 | Doxazosin mesilate | Doxazosin 1mg tablets (Actavis UK Ltd) |
| 50467 | 430311000001102 | Doxazosin mesilate | Doxazosin 2mg tablets (Alliance Healthcare (Distribution) Ltd) |
| 51685 | 4250511000001101 | Doxazosin mesilate | Doxazosin 4mg tablets (Actavis UK Ltd) |
| 53033 | 13811611000001108 | Doxazosin mesilate | Doxzogen XL 4mg tablets (Mylan) |
| 53322 | 16050211000001103 | Doxazosin mesilate | Doxazosin 4mg tablets (Bristol Laboratories Ltd) |
| 54785 | 19730511000001106 | Doxazosin mesilate | Doxazosin 4mg tablets (Medreich Plc) |
| 55906 | 11554511000001109 | Doxazosin mesilate | Doxazosin 1mg tablets (Dexcel-Pharma Ltd) |
| 55916 | 870611000001103 | Doxazosin mesilate | Doxazosin 1mg tablets (Alliance Healthcare (Distribution) Ltd) |
| 56145 | 4250311000001107 | Doxazosin mesilate | Doxazosin 2mg tablets (Actavis UK Ltd) |
| 57074 | 15083011000001109 | Doxazosin mesilate | Doxazosin 2mg tablets (Sigma Pharmaceuticals Plc) |
| 57448 | 588211000001106 | Doxazosin mesilate | Doxazosin 4mg tablets (A A H Pharmaceuticals Ltd) |
| 57784 | 12083611000001104 | Doxazosin mesilate | Doxazosin 2mg/ 5ml oral suspension |
| 58276 | 19730311000001100 | Doxazosin mesilate | Doxazosin 2mg tablets (Medreich Plc) |
| 58325 | 17894411000001101 | Doxazosin mesilate | Doxazosin 4mg tablets (Phoenix Healthcare Distribution Ltd) |
| 59209 | 252111000001100 | Doxazosin mesilate | Doxazosin 1mg tablets (Kent Pharmaceuticals Ltd) |
| 59862 | 11554911000001102 | Doxazosin mesilate | Doxazosin 4mg tablets (Dexcel-Pharma Ltd) |
| 60200 | 23970811000001104 | Doxazosin mesilate | Doxazosin 4mg tablets (DE Pharmaceuticals) |
| 60319 | 16049611000001105 | Doxazosin mesilate | Doxazosin 1mg tablets (Bristol Laboratories Ltd) |
| 61066 | 16049911000001104 | Doxazosin mesilate | Doxazosin 2mg tablets (Bristol Laboratories Ltd) |
| 61123 | 23466611000001105 | Doxazosin mesilate | Doxazosin 4mg/ 5ml oral solution |

| | | | |
|---|---|---|---|
| 61283 | 578111000001109 | Doxazosin mesilate | Doxazosin 4mg tablets (Alliance Healthcare (Distribution) Ltd) |
| 62019 | 9792611000001102 | Doxazosin mesilate | Doxazosin 1mg tablets (Almus Pharmaceuticals Ltd) |
| 62158 | 9793211000001105 | Doxazosin mesilate | Doxazosin 4mg tablets (Almus Pharmaceuticals Ltd) |
| 62351 | 17894211000001100 | Doxazosin mesilate | Doxazosin 2mg tablets (Phoenix Healthcare Distribution Ltd) |
| 63158 | 9792911000001108 | Doxazosin mesilate | Doxazosin 2mg tablets (Almus Pharmaceuticals Ltd) |
| 63314 | 17198011000001109 | Doxazosin mesilate | Doxazosin 1mg tablets (Sovereign Medical Ltd) |
| 64233 | 23591811000001109 | Doxazosin mesilate | Doxazosin 1mg tablets (Waymade Healthcare Plc) |
| 65159 | 23592211000001101 | Doxazosin mesilate | Doxazosin 4mg tablets (Waymade Healthcare Plc) |
| 65853 | 30122411000001102 | Doxazosin mesilate | Doxazosin 1mg tablets (Mawdsley-Brooks & Company Ltd) |
| 66065 | 225411000001101 | Doxazosin mesilate | Doxazosin 2mg tablets (Kent Pharmaceuticals Ltd) |
| 68022 | 17198511000001101 | Doxazosin mesilate | Doxazosin 4mg tablets (Sovereign Medical Ltd) |
| 68161 | 30123811000001108 | Doxazosin mesilate | Doxazosin 4mg tablets (Mawdsley-Brooks & Company Ltd) |
| 69319 | 23970611000001103 | Doxazosin mesilate | Doxazosin 2mg tablets (DE Pharmaceuticals) |
| 69757 | 12084811000001102 | Doxazosin mesilate | Doxazosin 8mg/ 5ml oral suspension |
| 71405 | 13846611000001108 | Doxazosin mesilate | Cardura XL 4mg tablets (DE Pharmaceuticals) |
| 72346 | 12083511000001103 | Doxazosin mesilate | Doxazosin 2mg/ 5ml oral solution |
| 72348 | 24509711000001103 | Doxazosin mesilate | Doxazosin 1mg/ 5ml oral solution |
| 73116 | 17198311000001107 | Doxazosin mesilate | Doxazosin 2mg tablets (Sovereign Medical Ltd) |
| 73637 | 4469911000001108 | Doxazosin mesilate | Doxazosin 1mg tablets (Sterwin Medicines) |
| 74000 | 5346711000001108 | Doxazosin mesilate | Cardura XL 4mg tablets (Waymade Healthcare Plc) |
| 74567 | 421069003 | Doxazosin mesilate | Doxazosin 8mg tablets |
| 74831 | 15083211000001104 | Doxazosin mesilate | Doxazosin 4mg tablets (Sigma Pharmaceuticals Plc) |
| 76149 | | Doxazosin mesilate | Doxazosin 5mg/ 5ml oral suspension |
| 77362 | | Doxazosin mesilate | Cardura 2mg tablets (Sigma Pharmaceuticals Plc) |
| 77436 | | Doxazosin mesilate | Cardura XL 4mg tablets (Mawdsley-Brooks & Company Ltd) |
| 4374 | 108525001000027101 | Debrisoquine Sulphate | Debrisoquine 20mg tablets |
| 8342 | 6915001000027102 | Debrisoquine Sulphate | Declinax 20mg Tablet (Roche Products Ltd) |
| 4375 | 318724006 | Debrisoquine sulfate | Debrisoquine 10mg tablets |
| 7911 | 6905001000027104 | Debrisoquine sulfate | Declinax 10mg Tablet (Roche Products Ltd) |
| 338 | 322840006 | Clonidine | Clonidine 25microgram tablets |
| 2630 | 344511000001105 | Clonidine | Dixarit 25microgram tablets (Boehringer Ingelheim Ltd) |
| 2878 | 318667005 | Clonidine | Clonidine 100microgram tablets |
| 4215 | 215111000001101 | Clonidine | Catapres 100microgram tablets (Boehringer Ingelheim Ltd) |
| 5289 | 36089511000001100 | Clonidine | Clonidine 250microgram modified-release capsules |
| 6694 | 318668000 | Clonidine | Clonidine 300microgram tablets |
| 8296 | 4536811000001109 | Clonidine | Catapres PL Perlongets 250microgram capsules (Boehringer Ingelheim Ltd) |
| 16248 | 36089211000001103 | Clonidine | Clonidine 150micrograms/ 1ml solution for injection ampoules |
| 23380 | 368711000001103 | Clonidine | Catapres 300microgram tablets (Boehringer Ingelheim Ltd) |
| 30293 | 364911000001108 | Clonidine | Catapres 150micrograms/ 1ml solution for injection ampoules (Boehringer Ingelheim Ltd) |
| 33093 | 3963611000001104 | Clonidine | Clonidine 25microgram tablets (Sandoz Ltd) |
| 45578 | 4920211000001104 | Clonidine | Clonidine 25microgram tablets (A A H Pharmaceuticals Ltd) |
| 49684 | 10448511000001104 | Clonidine | Clonidine 100micrograms/ 24hours transdermal patches |
| 52555 | 8398511000001100 | Clonidine | Clonidine 50micrograms/ 5ml oral solution |

| | | | |
|---|---|---|---|
| 53142 | 8398611000001101 | Clonidine | Clonidine 50micrograms/ 5ml oral suspension |
| 54467 | 10449111000001101 | Clonidine | Clonidine 300micrograms/ 24hours transdermal patches |
| 55797 | 8398311000001106 | Clonidine | Clonidine 25micrograms/ 5ml oral solution |
| 58090 | 11814111000001107 | Clonidine | Clonidine 75micrograms/ 5ml oral solution |
| 58529 | 7660211000001104 | Clonidine | Clonidine 200micrograms/ 24hours transdermal patches |
| 60089 | 21922811000001105 | Clonidine | Clonidine 25microgram tablets (Waymade Healthcare Plc) |
| 60136 | 11812911000001101 | Clonidine | Clonidine 100micrograms/ 5ml oral solution |
| 61256 | 8426311000001108 | Clonidine | Clonidine 5micrograms/ 5ml oral suspension |
| 61710 | 18402811000001107 | Clonidine | Clonidine 25microgram tablets (Teva UK Ltd) |
| 63971 | 15077611000001101 | Clonidine | Clonidine 25microgram tablets (Sigma Pharmaceuticals Plc) |
| 64284 | 16159511000001104 | Clonidine | Dixarit 25microgram tablets (Lexon (UK) Ltd) |
| 67200 | 8397911000001106 | Clonidine | Clonidine 10micrograms/ 5ml oral solution |
| 72130 | 11813011000001109 | Clonidine | Clonidine 100micrograms/ 5ml oral suspension |
| 72227 | 13867511000001104 | Clonidine | Dixarit 25microgram tablets (DE Pharmaceuticals) |
| 72807 | 11813311000001107 | Clonidine | Clonidine 15micrograms/ 5ml oral solution |
| 74534 | 32392011000001103 | Clonidine | Clonidine 250micrograms/ 5ml oral suspension |
| 75207 | 36392711000001102 | Clonidine | Clonidine 50micrograms/ 5ml oral solution sugar free |
| 76766 | | Clonidine | Catapres TTS 3 patches (Imported (Germany)) |
| 77591 | | Clonidine | Clonidine 12.5micrograms/ 5ml oral suspension |
| 29560 | 407815004 | Bosentan monohydrate | Bosentan 62.5mg tablets |
| 29561 | 407816003 | Bosentan monohydrate | Bosentan 125mg tablets |
| 47654 | 4765211000001107 | Bosentan monohydrate | Tracleer 62.5mg tablets (Actelion Pharmaceuticals UK Ltd) |
| 58632 | 4765711000001100 | Bosentan monohydrate | Tracleer 125mg tablets (Actelion Pharmaceuticals UK Ltd) |
| 10879 | 122435001000027102 | Betanidine Sulphate | Bethanidine sulphate 10mg tablets |
| 14442 | 10505001000027103 | Betanidine Sulphate | Esbatal 50mg Tablet (Wellcome Medical Division) |
| 18247 | 10495001000027109 | Betanidine Sulphate | Esbatal 10mg Tablet (Wellcome Medical Division) |
| 28676 | 122445001000027101 | Betanidine Sulphate | Bethanidine sulphate 50mg tablets |
| 29443 | 122475001000027100 | Betanidine Sulphate | Bendogen 10mg Tablet (Lagap) |
| 40421 | 122485001000027105 | Betanidine Sulphate | Bendogen 50mg Tablet (Lagap) |
| 40527 | 428480001 | Ambrisentan | Ambrisentan 10mg tablets |
| 40528 | 429662009 | Ambrisentan | Ambrisentan 5mg tablets |
| 214 | 168745001000027105 | | HYDRALAZINE 1 MG SYR |
| 445 | 4078311000001100 | | Prazosin 1mg tablets and Prazosin 500microgram tablets |
| 2649 | 70895001000027105 | | METHYLDOPA 250 MG CAP |
| 4507 | 145525001000027109 | | HYDRALAZINE 12.5 MG TAB |
| 19635 | | | DIXARIT |
| 20808 | | | CATAPRES PERLONGETS |
| 21749 | 168685001000027105 | | HYDRALAZINE 100 MG TAB |
| 23746 | 179545001000027108 | | HYDRALAZINE 10 MG TAB |
| 25088 | | | CATAPRES |
| 25836 | 141085001000027104 | | METHYLDOPA 200 MG TAB |
| 27894 | | | CLONIDINE |
| 28790 | | | CATAPRES |

| | | | |
|---|---|---|---|
| 31971 | 145575001000027108 | | HYDRALAZINE 6.25 MG SYR |
| **Beta-blockers** | | | |
| 7852 | 51195001000027109 | Timolol maleate | Blocadren 10mg Tablet (Merck Sharp & Dohme Ltd) |
| 7853 | 318534000 | Timolol maleate | Timolol 10mg tablets |
| 12037 | 3315001000027104 | Timolol maleate | Betim 10mg Tablet (ICN Pharmaceuticals France S.A.) |
| 29610 | 72811000001104 | Timolol maleate | Betim 10mg tablets (Meda Pharmaceuticals Ltd) |
| 786 | 318525005 | Sotalol | Sotalol 40mg tablets |
| 1572 | 318526006 | Sotalol | Sotalol 80mg tablets |
| 4004 | 463811000001109 | Sotalol | Sotacor 80mg tablets (Bristol-Myers Squibb Pharmaceuticals Ltd) |
| 5858 | 900511000001103 | Sotalol | Beta-Cardone 40mg tablets (Focus Pharmaceuticals Ltd) |
| 6751 | 104011000001101 | Sotalol | Beta-Cardone 80mg tablets (Focus Pharmaceuticals Ltd) |
| 9292 | 318528007 | Sotalol | Sotalol 160mg tablets |
| 11380 | 446211000001108 | Sotalol | Sotacor 160mg tablets (Bristol-Myers Squibb Pharmaceuticals Ltd) |
| 13051 | 318527002 | Sotalol | Sotalol 200mg tablets |
| 13487 | 447111000001104 | Sotalol | Beta-Cardone 200mg tablets (Focus Pharmaceuticals Ltd) |
| 17679 | 141245001000027103 | Sotalol | Sotalol 10mg/ ml injection |
| 24635 | 141365001000027103 | Sotalol | Sotacor 10mg/ ml Injection (Bristol-Myers Squibb Pharmaceuticals Ltd) |
| 27727 | 141255001000027100 | Sotalol | Sotalol 2mg/ ml injection |
| 33578 | 694711000001100 | Sotalol | Sotacor 40mg/ 4ml solution for injection ampoules (Bristol-Myers Squibb Pharmaceuticals Ltd) |
| 34371 | 936211000001109 | Sotalol | Sotalol 40mg tablets (A A H Pharmaceuticals Ltd) |
| 34520 | 680311000001106 | Sotalol | Sotalol 80mg tablets (Mylan) |
| 34600 | 3659211000001106 | Sotalol | Sotalol 40mg tablets (Teva UK Ltd) |
| 34640 | 89165001000027103 | Sotalol | Sotalol 40mg Tablet (Tillomed Laboratories Ltd) |
| 34690 | 170011000001101 | Sotalol | Sotalol 80mg tablets (Sandoz Ltd) |
| 35710 | 8726911000001102 | Sotalol | Sotalol 25mg/ 5ml oral suspension |
| 38498 | 35930811000001109 | Sotalol | Sotalol 40mg/ 4ml solution for injection ampoules |
| 39423 | 909811000001109 | Sotalol | Sotalol 80mg tablets (A A H Pharmaceuticals Ltd) |
| 43549 | 4230411000001104 | Sotalol | Sotalol 40mg tablets (IVAX Pharmaceuticals UK Ltd) |
| 51492 | 8726811000001107 | Sotalol | Sotalol 25mg/ 5ml oral solution |
| 70161 | 8727111000001102 | Sotalol | Sotalol 40mg/ 5ml oral solution |
| 70162 | 8727211000001108 | Sotalol | Sotalol 40mg/ 5ml oral suspension |
| 70734 | 11427411000001105 | Sotalol | Sotalol 40mg tablets (Almus Pharmaceuticals Ltd) |
| 71479 | 16067611000001105 | Sotalol | Sotalol 40mg tablets (Bristol Laboratories Ltd) |
| 72347 | 8727511000001106 | Sotalol | Sotalol 80mg/ 5ml oral suspension |
| 73481 | 3658411000001102 | Sotalol | Sotalol 160mg tablets (Teva UK Ltd) |
| 74628 | 3659411000001105 | Sotalol | Sotalol 80mg tablets (Teva UK Ltd) |
| 74829 | 17799811000001102 | Sotalol | Sotalol 40mg tablets (Phoenix Healthcare Distribution Ltd) |
| 75060 | 243155001000027107 | Sotalol | Sotalol oral solution |
| 75821 | | Sotalol | Sotalol 40mg tablets (Alliance Healthcare (Distribution) Ltd) |
| 220 | 89655001000027106 | Propranolol | Propranolol 5mg/ 5ml oral solution |
| 297 | 318352004 | Propranolol | Propranolol 10mg tablets |
| 707 | 318353009 | Propranolol | Propranolol 40mg tablets |
| 769 | 318407001 | Propranolol | Propranolol 80mg modified-release capsules |

| | | | |
|---|---|---|---|
| 940 | 318354003 | Propranolol | Propranolol 80mg tablets |
| 1006 | 226211000001106 | Propranolol | Half Inderal LA 80mg capsules (AstraZeneca UK Ltd) |
| 1048 | 821011000001100 | Propranolol | Inderal 80mg tablets (AstraZeneca UK Ltd) |
| 1050 | 930211000001107 | Propranolol | Inderal 40mg tablets (AstraZeneca UK Ltd) |
| 1448 | 318406005 | Propranolol | Propranolol 160mg modified-release capsules |
| 2414 | 567911000001100 | Propranolol | Inderal 10mg tablets (AstraZeneca UK Ltd) |
| 3005 | 154311000001108 | Propranolol | Inderal LA 160mg capsules (AstraZeneca UK Ltd) |
| 3087 | 15356211000001102 | Propranolol | Propranolol 40mg/ 5ml oral solution sugar free |
| 3167 | 318355002 | Propranolol | Propranolol 160mg tablets |
| 3827 | 121565001000027103 | Propranolol | Propanix 40mg Tablet (Ashbourne Pharmaceuticals Ltd) |
| 5478 | 35932311000001101 | Propranolol | Propranolol 10mg/ 5ml oral solution sugar free |
| 8331 | 51875001000027109 | Propranolol | Inderal 160mg Tablet (AstraZeneca UK Ltd) |
| 8978 | 120695001000027100 | Propranolol | Propanix 160mg Modified-release capsule (Ashbourne Pharmaceuticals Ltd) |
| 9185 | 196405001000027103 | Propranolol | Propranolol 80mg/ 5ml oral solution |
| 10294 | 4371111000001106 | Propranolol | Inderal 1mg/ 1ml solution for injection ampoules (AstraZeneca UK Ltd) |
| 11711 | 196395001000027106 | Propranolol | Propranolol 50mg/ 5ml oral solution |
| 12495 | 54385001000027105 | Propranolol | Berkolol 10mg Tablet (Berk Pharmaceuticals Ltd) |
| 14552 | 121555001000027107 | Propranolol | Propanix 10mg Tablet (Ashbourne Pharmaceuticals Ltd) |
| 14808 | 573711000001104 | Propranolol | Bedranol SR 80mg capsules (Sandoz Ltd) |
| 15619 | 72255001000027102 | Propranolol | Half-betadur cr 80mg Capsule (Monmouth Pharmaceuticals Ltd) |
| 17082 | 295911000001102 | Propranolol | Syprol 5mg/ 5ml oral solution (Rosemont Pharmaceuticals Ltd) |
| 20468 | 724111000001103 | Propranolol | Half Beta-Prograne 80mg modified-release capsules (Tillomed Laboratories Ltd) |
| 21838 | 121575001000027105 | Propranolol | Propanix 80mg Tablet (Ashbourne Pharmaceuticals Ltd) |
| 21839 | 54405001000027101 | Propranolol | Berkolol 80mg Tablet (Berk Pharmaceuticals Ltd) |
| 21866 | 54395001000027109 | Propranolol | Berkolol 40mg Tablet (Berk Pharmaceuticals Ltd) |
| 22208 | 227945001000027104 | Propranolol | Half propanix la 80mg Modified-release capsule (Ashbourne Pharmaceuticals Ltd) |
| 23326 | 175605001000027100 | Propranolol | Betadur cr 160mg Modified-release capsule (Monmouth Pharmaceuticals Ltd) |
| 23587 | 138495001000027100 | Propranolol | Sloprolol 160mg Capsule (C P Pharmaceuticals Ltd) |
| 24218 | 54445001000027106 | Propranolol | Berkolol 160mg Tablet (Berk Pharmaceuticals Ltd) |
| 25359 | 2899211000001100 | Propranolol | Rapranol SR 160mg capsules (Ranbaxy (UK) Ltd) |
| 25367 | 2899011000001105 | Propranolol | Rapranol SR 80mg capsules (Ranbaxy (UK) Ltd) |
| 26228 | 231365001000027104 | Propranolol | Propanix LA 160mg Modified-release capsule (Ashbourne Pharmaceuticals Ltd) |
| 26229 | 510611000001108 | Propranolol | Beta-Prograne 160mg modified-release capsules (Tillomed Laboratories Ltd) |
| 26255 | 207165001000027106 | Propranolol | Lopranol la 160mg Capsule (Opus Pharmaceuticals Ltd) |
| 26895 | 28711000001109 | Propranolol | Syprol 10mg/ 5ml oral solution (Rosemont Pharmaceuticals Ltd) |
| 27486 | 35932611000001106 | Propranolol | Propranolol 1mg/ 1ml solution for injection ampoules |
| 27700 | 92011000001103 | Propranolol | Propranolol 40mg tablets (Actavis UK Ltd) |
| 27964 | 75515001000027103 | Propranolol | Apsolol 40mg Tablet (Approved Prescription Services Ltd) |
| 28048 | 90305001000027107 | Propranolol | Angilol 10mg Tablet (DDSA Pharmaceuticals Ltd) |
| 28128 | 49655001000027103 | Propranolol | Propranolol 80mg Modified-release capsule (Actavis UK Ltd) |
| 28788 | 232405001000027109 | Propranolol | Half propatard la 80mg Modified-release capsule (Galen Ltd) |
| 28996 | 549611000001106 | Propranolol | Bedranol SR 160mg capsules (Sandoz Ltd) |
| 29763 | 120685001000027104 | Propranolol | Propanix 160mg Tablet (Ashbourne Pharmaceuticals Ltd) |

| | | | |
|---|---|---|---|
| 31214 | 694511000001105 | Propranolol | Propranolol 80mg tablets (Mylan) |
| 31776 | 23611000001107 | Propranolol | Propranolol 40mg tablets (Mylan) |
| 31833 | 90325001000027100 | Propranolol | Angilol 80mg Tablet (DDSA Pharmaceuticals Ltd) |
| 32162 | 90295001000027100 | Propranolol | Propranolol 80mg Modified-release capsule (Lagap) |
| 33376 | 208225001000027105 | Propranolol | Probeta LA 160mg Capsule (Trinity Pharmaceuticals Ltd) |
| 33602 | 240711000001100 | Propranolol | Slo-Pro 160mg capsules (Mylan) |
| 33644 | 763811000001100 | Propranolol | Propranolol 80mg tablets (A A H Pharmaceuticals Ltd) |
| 33836 | 75555001000027108 | Propranolol | Apsolol 160mg Tablet (Approved Prescription Services Ltd) |
| 34185 | 65085001000027102 | Propranolol | Propranolol LA 80mg Modified-release capsule (Approved Prescription Services Ltd) |
| 34208 | 30275001000027102 | Propranolol | Propranolol SR 160mg Modified-release capsule (C P Pharmaceuticals Ltd) |
| 34214 | 696011000001107 | Propranolol | Propranolol 160mg tablets (Actavis UK Ltd) |
| 34378 | 695911000001104 | Propranolol | Propranolol 10mg tablets (A A H Pharmaceuticals Ltd) |
| 34783 | 98911000001101 | Propranolol | Propranolol 10mg tablets (Actavis UK Ltd) |
| 34804 | 212711000001109 | Propranolol | Propranolol 10mg tablets (Teva UK Ltd) |
| 34867 | 63965001000027101 | Propranolol | Propranolol 80mg Capsule (IVAX Pharmaceuticals UK Ltd) |
| 34868 | 859211000001105 | Propranolol | Propranolol 40mg tablets (Teva UK Ltd) |
| 34884 | 141395001000027101 | Propranolol | Propranolol 160mg Modified-release capsule (Sandoz Ltd) |
| 34945 | 90285001000027104 | Propranolol | Propranolol 160mg Modified-release capsule (Lagap) |
| 34949 | 49665001000027107 | Propranolol | Propranolol 160mg Modified-release capsule (Actavis UK Ltd) |
| 35938 | 168311000001107 | Propranolol | Propranolol 80mg modified-release capsules (A A H Pharmaceuticals Ltd) |
| 36576 | 397911000001104 | Propranolol | Propranolol 10mg tablets (Mylan) |
| 36603 | 22555001000027104 | Propranolol | Propranolol SR 160mg Modified-release capsule (Hillcross Pharmaceuticals Ltd) |
| 38433 | 60395001000027107 | Propranolol | Propranolol 50mg/ 5ml Oral solution (Rosemont Pharmaceuticals Ltd) |
| 39233 | 8393911000001102 | Propranolol | Propranolol 80mg modified-release capsules (Teva UK Ltd) |
| 40241 | 65075001000027107 | Propranolol | Propranolol LA 160mg Capsule (Approved Prescription Services Ltd) |
| 41555 | 188311000001109 | Propranolol | Propranolol 40mg tablets (A A H Pharmaceuticals Ltd) |
| 42152 | 823311000001101 | Propranolol | Syprol 50mg/ 5ml oral solution (Rosemont Pharmaceuticals Ltd) |
| 43525 | 395011000001105 | Propranolol | Propranolol 10mg tablets (IVAX Pharmaceuticals UK Ltd) |
| 45297 | 502911000001101 | Propranolol | Propranolol 40mg tablets (IVAX Pharmaceuticals UK Ltd) |
| 45343 | 30285001000027107 | Propranolol | Propranolol SR 80mg Modified-release capsule (C P Pharmaceuticals Ltd) |
| 45494 | 9808311000001105 | Propranolol | Propranolol 10mg tablets (Almus Pharmaceuticals Ltd) |
| 45765 | 15307311000001100 | Propranolol | Syprol 40mg/ 5ml oral solution (Rosemont Pharmaceuticals Ltd) |
| 45877 | 15610411000001104 | Propranolol | Beta-Prograne 160mg modified-release capsules (Teva UK Ltd) |
| 46363 | 15610611000001101 | Propranolol | Half Beta-Prograne 80mg modified-release capsules (Teva UK Ltd) |
| 47543 | 18069311000001100 | Propranolol | Half Beta-Prograne 80mg modified-release capsules (Actavis UK Ltd) |
| 47833 | 18694811000001101 | Propranolol | Bedranol SR 80mg capsules (Almus Pharmaceuticals Ltd) |
| 47907 | 18695311000001109 | Propranolol | Bedranol SR 160mg capsules (Almus Pharmaceuticals Ltd) |
| 48682 | 35932711000001102 | Propranolol | Propranolol 50mg/ 5ml oral solution sugar free |
| 49863 | 35932811000001105 | Propranolol | Propranolol 5mg/ 5ml oral solution sugar free |
| 52136 | 160775001000027108 | Propranolol | Bedranol sr 160mg Capsule (Lagap) |
| 52609 | 14252111000001108 | Propranolol | Inderal LA 160mg capsules (Sigma Pharmaceuticals Plc) |
| 52777 | 587411000001106 | Propranolol | Propranolol 40mg tablets (Kent Pharmaceuticals Ltd) |
| 53177 | 244465001000027101 | Propranolol | Propranolol oral solution |

| | | | |
|---|---|---|---|
| 54297 | 13199511000001100 | Propranolol | Propranolol 50mg/ 5ml oral solution |
| 54623 | 18069011000001103 | Propranolol | Beta-Prograne 160mg modified-release capsules (Actavis UK Ltd) |
| 55228 | 14782911000001107 | Propranolol | Propranolol 40mg tablets (Boston Healthcare Ltd) |
| 55416 | 9808611000001100 | Propranolol | Propranolol 40mg tablets (Almus Pharmaceuticals Ltd) |
| 55849 | 582811000001105 | Propranolol | Propranolol 160mg tablets (Mylan) |
| 55949 | 8672711000001106 | Propranolol | Propranolol 40mg/ 5ml oral solution |
| 56173 | 18069311000001100 | Propranolol | Half Beta-Prograne 80mg modified-release capsules (Actavis UK Ltd) |
| 56764 | 21870611000001101 | Propranolol | Propranolol 40mg tablets (Waymade Healthcare Plc) |
| 57063 | 18694811000001101 | Propranolol | Bedranol SR 80mg capsules (Almus Pharmaceuticals Ltd) |
| 57342 | 17919711000001101 | Propranolol | Propranolol 40mg tablets (Phoenix Healthcare Distribution Ltd) |
| 57567 | 13160911000001104 | Propranolol | Propranolol 10mg/ 5ml oral suspension |
| 58297 | 163111000001101 | Propranolol | Propranolol 10mg tablets (Kent Pharmaceuticals Ltd) |
| 58407 | 59811000001107 | Propranolol | Propranolol 80mg tablets (Teva UK Ltd) |
| 58491 | 316911000001101 | Propranolol | Propranolol 40mg tablets (Alliance Healthcare (Distribution) Ltd) |
| 59415 | 23489711000001109 | Propranolol | Propranolol 40mg tablets (Accord Healthcare Ltd) |
| 59597 | 4577511000001105 | Propranolol | Propranolol 160mg modified-release capsules (A A H Pharmaceuticals Ltd) |
| 60565 | 9153911000001101 | Propranolol | Propranolol 40mg tablets (Ranbaxy (UK) Ltd) |
| 60934 | 388811000001109 | Propranolol | Propranolol 80mg modified-release capsules (Kent Pharmaceuticals Ltd) |
| 61727 | 23489111000001108 | Propranolol | Propranolol 10mg tablets (Accord Healthcare Ltd) |
| 62711 | 23601111000001107 | Propranolol | Propranolol 80mg modified-release capsules (Waymade Healthcare Plc) |
| 64160 | 22352811000001103 | Propranolol | Propranolol 5mg/ 5ml oral solution sugar free (AM Distributions (Yorkshire) Ltd) |
| 65435 | 21870211000001103 | Propranolol | Propranolol 10mg tablets (Waymade Healthcare Plc) |
| 65986 | 795211000001104 | Propranolol | Propranolol 10mg tablets (Alliance Healthcare (Distribution) Ltd) |
| 66555 | 14782711000001105 | Propranolol | Propranolol 10mg tablets (Boston Healthcare Ltd) |
| 68400 | 30081311000001101 | Propranolol | Propranolol 160mg tablets (DE Pharmaceuticals) |
| 69661 | 13172311000001106 | Propranolol | Propranolol 3mg/ 5ml oral solution |
| 70680 | 13200011000001100 | Propranolol | Propranolol 6mg/ 5ml oral suspension |
| 70681 | 13171211000001105 | Propranolol | Propranolol 3mg/ 5ml oral suspension |
| 71150 | 13199711000001105 | Propranolol | Propranolol 5mg/ 5ml oral solution |
| 71173 | 30860011000001105 | Propranolol | Propranolol 80mg modified-release capsules (Mawdsley-Brooks & Company Ltd) |
| 72220 | 24412311000001103 | Propranolol | Propranolol 10mg/ 5ml oral solution sugar free (CST Pharma Ltd) |
| 72222 | 30080311000001106 | Propranolol | Propranolol 80mg modified-release capsules (DE Pharmaceuticals) |
| 73653 | 9154111000001102 | Propranolol | Propranolol 80mg tablets (Ranbaxy (UK) Ltd) |
| 73765 | 9153711000001103 | Propranolol | Propranolol 10mg tablets (Ranbaxy (UK) Ltd) |
| 75278 | 13199811000001102 | Propranolol | Propranolol 5mg/ 5ml oral suspension |
| 75289 | 32759511000001108 | Propranolol | Propranolol 5mg/ 5ml oral solution sugar free (Rosemont Pharmaceuticals Ltd) |
| 75312 | 36106711000001107 | Propranolol | Bedranol 80mg tablets (Ennogen Pharma Ltd) |
| 75507 | | Propranolol | Propranolol 160mg Capsule (IVAX Pharmaceuticals UK Ltd) |
| 76252 | | Propranolol | Propranolol 160mg modified-release capsules (Teva UK Ltd) |
| 76431 | | Propranolol | Cardinol 10mg Tablet (C P Pharmaceuticals Ltd) |
| 76633 | | Propranolol | Propranolol 10mg tablets (DE Pharmaceuticals) |
| 77413 | | Propranolol | Inderal LA 160mg capsules (Waymade Healthcare Plc) |
| 77649 | | Propranolol | Half Inderal LA 80mg capsules (DE Pharmaceuticals) |

| | | | |
|---|---|---|---|
| 20169 | 137695001000027105 | Practolol | Practolol 2mg/ ml injection |
| 4588 | 34545001000027108 | Pindolol | Visken 5mg Tablet (Sovereign Medical Ltd) |
| 5284 | 318512002 | Pindolol | Pindolol 5mg tablets |
| 14673 | 318513007 | Pindolol | Pindolol 15mg tablets |
| 20012 | 34555001000027106 | Pindolol | Visken 15mg Tablet (Sovereign Medical Ltd) |
| 32787 | 3887411000001101 | Pindolol | Visken 15mg tablets (AMCo) |
| 35695 | 3706111000001105 | Pindolol | Visken 5mg tablets (AMCo) |
| 55853 | 96905001000027105 | Pindolol | Pindolol 15mg Tablet (Hillcross Pharmaceuticals Ltd) |
| 73413 | 3707011000001107 | Pindolol | Pindolol 5mg tablets (A A H Pharmaceuticals Ltd) |
| 1333 | 318484005 | Oxprenolol | Oxprenolol 40mg tablets |
| 1334 | 3602301100000 1102 | Oxprenolol | Oxprenolol 160mg modified-release tablets |
| 2361 | 54835001000027104 | Oxprenolol | Trasicor 80mg Tablet (Novartis Pharmaceuticals UK Ltd) |
| 2780 | 318485006 | Oxprenolol | Oxprenolol 80mg tablets |
| 3516 | 318483004 | Oxprenolol | Oxprenolol 20mg tablets |
| 3748 | 89605001000027103 | Oxprenolol | Oxprenolol 160mg Tablet |
| 4025 | 473011000001107 | Oxprenolol | Slow-Trasicor 160mg tablets (AMCo) |
| 7474 | 54815001000027105 | Oxprenolol | Trasicor 20mg Tablet (Novartis Pharmaceuticals UK Ltd) |
| 8290 | 54825001000027101 | Oxprenolol | Trasicor 40mg Tablet (Novartis Pharmaceuticals UK Ltd) |
| 10777 | 54865001000027109 | Oxprenolol | Trasicor 160mg Tablet (Novartis Pharmaceuticals UK Ltd) |
| 21885 | 195255001000027109 | Oxprenolol | Oxyprenix SR 160mg tablets |
| 24094 | 592911000001104 | Oxprenolol | Trasicor 40mg tablets (Amdipharm Plc) |
| 25644 | 75595001000027102 | Oxprenolol | Apsolox 80mg Tablet (Approved Prescription Services Ltd) |
| 27357 | 9205001000027107 | Oxprenolol | Oxprenolol 40mg Tablet (Actavis UK Ltd) |
| 29180 | 568511000001106 | Oxprenolol | Trasicor 80mg tablets (Amdipharm Plc) |
| 29230 | 90575001000027104 | Oxprenolol | Slow-pren 160mg Tablet (IVAX Pharmaceuticals UK Ltd) |
| 33569 | 9405001000027108 | Oxprenolol | Oxprenolol sr 160mg Modified-release tablet (Hillcross Pharmaceuticals Ltd) |
| 35062 | 3379011000001106 | Oxprenolol | Trasicor 20mg tablets (Amdipharm Plc) |
| 74015 | 17595711000001107 | Oxprenolol | Slow-Trasicor 160mg tablets (Sigma Pharmaceuticals Plc) |
| 77445 | | Oxprenolol | Slow-Trasicor 160mg tablets (DE Pharmaceuticals) |
| 77461 | | Oxprenolol | Slow-Trasicor 160mg tablets (Waymade Healthcare Plc) |
| 751 | 318640004 | Nebivolol | Nebivolol 5mg tablets |
| 7528 | 726011000001109 | Nebivolol | Nebilet 5mg tablets (A. Menarini Farmaceutica Internazionale SRL) |
| 40761 | 432118006 | Nebivolol | Nebivolol 2.5mg tablets |
| 44808 | 16551311000001103 | Nebivolol | Nebivolol 2.5mg tablets (A A H Pharmaceuticals Ltd) |
| 47300 | 15638611000001106 | Nebivolol | Nebivolol 2.5mg tablets (Glenmark Pharmaceuticals Europe Ltd) |
| 54487 | 21266011000001105 | Nebivolol | Nebivolol 2.5mg tablets (Sigma Pharmaceuticals Plc) |
| 59961 | 432159000 | Nebivolol | Nebivolol 10mg tablets |
| 64703 | 15638211000001109 | Nebivolol | Nebivolol 5mg tablets (Glenmark Pharmaceuticals Europe Ltd) |
| 66559 | 5393311000001106 | Nebivolol | Nebilet 5mg tablets (Waymade Healthcare Plc) |
| 67595 | 24483711000001109 | Nebivolol | Nebivolol 5mg tablets (Almus Pharmaceuticals Ltd) |
| 68677 | 14404711000001108 | Nebivolol | Nebivolol 5mg tablets (Accord Healthcare Ltd) |
| 69115 | 14033711000001109 | Nebivolol | Nebivolol 5mg tablets (A A H Pharmaceuticals Ltd) |
| 71032 | 20312411000001102 | Nebivolol | Nebivolol 5mg tablets (Sandoz Ltd) |

| | | | |
|---|---|---|---|
| 74076 | 13801211000001108 | Nebivolol | Nebivolol 5mg tablets (PLIVA Pharma Ltd) |
| 76484 | | Nebivolol | Nebivolol 5mg tablets (Sigma Pharmaceuticals Plc) |
| 2499 | 318481002 | Nadolol | Nadolol 80mg tablets |
| 8935 | 318480001 | Nadolol | Nadolol 40mg tablets |
| 10716 | 3687411000001106 | Nadolol | Corgard 80mg tablets (Sanofi) |
| 13415 | 3686511000001108 | Nadolol | Corgard 40mg tablets (Sanofi-Synthelabo Ltd) |
| 66464 | 12300711000001104 | Nadolol | Nadolol 40mg/ 5ml oral solution |
| 66779 | 16110011000001107 | Nadolol | Nadolol 30mg/ 5ml oral suspension |
| 67424 | 15227211000001106 | Nadolol | Nadolol 80mg/ 5ml oral suspension |
| 72159 | 16131011000001109 | Nadolol | Nadolol 20mg/ 5ml oral suspension |
| 73219 | 12300811000001107 | Nadolol | Nadolol 40mg/ 5ml oral suspension |
| 74929 | 250335001000027108 | Nadolol | Nadolol Oral solution |
| 74930 | 16104311000001104 | Nadolol | Nadolol 30mg/ 5ml oral suspension (Drug Tariff Special Order) |
| 77002 | | Nadolol | Corgard 80mg tablets (Lexon (UK) Ltd) |
| 77945 | | Nadolol | Nadolol 20mg/ 5ml oral solution |
| 739 | 318475005 | Metoprolol tartrate | Metoprolol 50mg tablets |
| 753 | 318474009 | Metoprolol tartrate | Metoprolol 100mg tablets |
| 3344 | 193911000001101 | Metoprolol tartrate | Betaloc 100mg tablets (AstraZeneca UK Ltd) |
| 3474 | 185111000001102 | Metoprolol tartrate | Betaloc-SA 200mg tablets (AstraZeneca UK Ltd) |
| 8068 | 36035411000001108 | Metoprolol tartrate | Metoprolol 200mg modified-release tablets |
| 8071 | 31811000001105 | Metoprolol tartrate | Betaloc 50mg tablets (AstraZeneca UK Ltd) |
| 10429 | 17735001000027102 | Metoprolol tartrate | Lopresor 50mg Tablet (Novartis Pharmaceuticals UK Ltd) |
| 11793 | 8669111000001108 | Metoprolol tartrate | Metoprolol 50mg/ 5ml oral suspension |
| 13499 | 17745001000027101 | Metoprolol tartrate | Lopresor 100mg Tablet (Novartis Pharmaceuticals UK Ltd) |
| 14502 | 3631211000001109 | Metoprolol tartrate | Metoprolol 5mg/ 5ml solution for injection ampoules |
| 20082 | 916111000001108 | Metoprolol tartrate | Lopresor SR 200mg tablets (Recordati Pharmaceuticals Ltd) |
| 24461 | 3615711000001109 | Metoprolol tartrate | Betaloc I.V. 5mg/ 5ml solution for injection ampoules (AstraZeneca UK Ltd) |
| 29762 | 120275001000027107 | Metoprolol tartrate | Mepranix 50mg Tablet (Ashbourne Pharmaceuticals Ltd) |
| 30400 | 120285001000027102 | Metoprolol tartrate | Mepranix 100mg Tablet (Ashbourne Pharmaceuticals Ltd) |
| 32836 | 683311000001100 | Metoprolol tartrate | Metoprolol 50mg tablets (Mylan) |
| 34092 | 89911000001105 | Metoprolol tartrate | Metoprolol 100mg tablets (Teva UK Ltd) |
| 34094 | 393511000001101 | Metoprolol tartrate | Metoprolol 50mg tablets (A A H Pharmaceuticals Ltd) |
| 34125 | 121711000001108 | Metoprolol tartrate | Metoprolol 100mg tablets (A A H Pharmaceuticals Ltd) |
| 34407 | 126811000001105 | Metoprolol tartrate | Metoprolol 50mg tablets (Teva UK Ltd) |
| 34430 | 499011000001103 | Metoprolol tartrate | Metoprolol 50mg tablets (Actavis UK Ltd) |
| 34509 | 497611000001100 | Metoprolol tartrate | Metoprolol 100mg tablets (Mylan) |
| 34584 | 407511000001102 | Metoprolol tartrate | Metoprolol 50mg tablets (IVAX Pharmaceuticals UK Ltd) |
| 34854 | 834111000001104 | Metoprolol tartrate | Metoprolol 100mg tablets (Actavis UK Ltd) |
| 34890 | 43455001000027104 | Metoprolol tartrate | Metoprolol 50mg Tablet (Berk Pharmaceuticals Ltd) |
| 34925 | 519211000001104 | Metoprolol tartrate | Metoprolol 50mg tablets (Sandoz Ltd) |
| 40167 | 203711000001103 | Metoprolol tartrate | Metoprolol 100mg tablets (IVAX Pharmaceuticals UK Ltd) |
| 45289 | 243965001000027101 | Metoprolol Tartrate | Metoprolol tartrate Oral solution |
| 46614 | 113511000001109 | Metoprolol tartrate | Lopresor 50mg tablets (Recordati Pharmaceuticals Ltd) |

| | | | |
|---|---|---|---|
| 46740 | 756911000001107 | Metoprolol tartrate | Lopresor 100mg tablets (Recordati Pharmaceuticals Ltd) |
| 47536 | 304045001000027103 | Metoprolol Tartrate | Metoprolol tartrate 12.5mg/ 5ml Oral suspension |
| 51447 | 8668411000001106 | Metoprolol tartrate | Metoprolol 12.5mg/ 5ml oral suspension |
| 55979 | 8668811000001108 | Metoprolol tartrate | Metoprolol 25mg/ 5ml oral suspension |
| 57240 | 8635811000001107 | Metoprolol tartrate | Metoprolol 50mg/ 5ml oral suspension (Special Order) |
| 63724 | 21799011000001109 | Metoprolol tartrate | Metoprolol 100mg tablets (Waymade Healthcare Plc) |
| 65227 | 8668211000001107 | Metoprolol tartrate | Metoprolol 12.5mg/ 5ml oral solution |
| 66670 | 610811000001104 | Metoprolol tartrate | Metoprolol 100mg tablets (Alliance Healthcare (Distribution) Ltd) |
| 68881 | 16632411000001109 | Metoprolol tartrate | Metoprolol 12.5mg capsules |
| 70116 | 8668911000001103 | Metoprolol tartrate | Metoprolol 50mg/ 5ml oral solution |
| 71098 | 21286311000001105 | Metoprolol tartrate | Metoprolol 50mg tablets (Accord Healthcare Ltd) |
| 74854 | 8668711000001100 | Metoprolol tartrate | Metoprolol 25mg/ 5ml oral solution |
| 75010 | 16235001000027106 | Metoprolol tartrate | Metoprolol tartrate 50mg Tablet (C P Pharmaceuticals Ltd) |
| 76180 | | Metoprolol tartrate | Metoprolol 5mg/ 5ml oral solution |
| 76531 | | Metoprolol tartrate | Metoprolol 10mg/ 5ml oral suspension |
| 27719 | 160665001000027109 | Metoprolol | Metoros ls 95mg Tablet (Geigy Pharmaceuticals) |
| 29998 | 160625001000027101 | Metoprolol | Metoros 190mg Tablet (Novartis Pharmaceuticals UK Ltd) |
| 77282 | | Metoprolol | Metoprolol 190mg Modified-release tablet |
| 1295 | 318447008 | Labetalol | Labetalol 400mg tablets |
| 1597 | 318445000 | Labetalol | Labetalol 100mg tablets |
| 2775 | 318446004 | Labetalol | Labetalol 200mg tablets |
| 4725 | 318458004 | Labetalol | Labetalol 50mg tablets |
| 8707 | 77311000001105 | Labetalol | Trandate 200mg tablets (RPH Pharmaceuticals AB) |
| 8807 | 674711000001109 | Labetalol | Trandate 400mg tablets (RPH Pharmaceuticals AB) |
| 9016 | 639011000001106 | Labetalol | Trandate 100mg tablets (RPH Pharmaceuticals AB) |
| 9273 | 42611000001104 | Labetalol | Trandate 50mg tablets (RPH Pharmaceuticals AB) |
| 16645 | 82695001000027107 | Labetalol | Labrocol 400mg Tablet (Lagap) |
| 19068 | 7978711000001103 | Labetalol | Labetalol 50mg/ 10ml solution for injection pre-filled syringes |
| 19998 | 4413911000001106 | Labetalol | Trandate 100mg/ 20ml solution for injection ampoules (Focus Pharmaceuticals Ltd) |
| 22793 | 82685001000027108 | Labetalol | Labrocol 200mg Tablet (Lagap) |
| 30770 | 22311000001107 | Labetalol | Labetalol 200mg tablets (A A H Pharmaceuticals Ltd) |
| 34171 | 14865001000027100 | Labetalol | Labetalol 100mg Tablet (C P Pharmaceuticals Ltd) |
| 34177 | 50911000001108 | Labetalol | Labetalol 100mg tablets (A A H Pharmaceuticals Ltd) |
| 34188 | 27395001000027109 | Labetalol | Labetalol 200mg Tablet (Celltech Pharma Europe Ltd) |
| 35778 | 82675001000027103 | Labetalol | Labrocol 100mg Tablet (Lagap) |
| 38370 | 36037611000001104 | Labetalol | Labetalol 100mg/ 20ml solution for injection ampoules |
| 40240 | 246511000001107 | Labetalol | Labetalol 400mg tablets (A A H Pharmaceuticals Ltd) |
| 41827 | 2211000001109 | Labetalol | Labetalol 100mg tablets (Mylan) |
| 44083 | 265911000001104 | Labetalol | Labetalol 200mg tablets (Actavis UK Ltd) |
| 45250 | 929111000001109 | Labetalol | Labetalol 400mg tablets (Sandoz Ltd) |
| 47673 | 52485001000027108 | Labetalol | Labetalol 400mg Tablet (Approved Prescription Services Ltd) |
| 47674 | 14875001000027107 | Labetalol | Labetalol 200mg Tablet (C P Pharmaceuticals Ltd) |
| 59222 | 21024711000001102 | Labetalol | Labetalol 100mg/ 20ml solution for injection ampoules (RPH Pharmaceuticals AB) |

| | | | |
|---|---|---|---|
| 62638 | 281111000001101 | Labetalol | Labetalol 100mg tablets (Actavis UK Ltd) |
| 63736 | 22055311000001102 | Labetalol | Labetalol 100mg tablets (Waymade Healthcare Plc) |
| 72514 | 12643411000001100 | Labetalol | Labetalol 1.5mg/ 5ml oral solution |
| 75179 | 99411000001101 | Labetalol | Labetalol 200mg tablets (Mylan) |
| 77958 | | Labetalol | Labetalol 50mg/ 5ml oral suspension |
| 26922 | 10070311000001104 | Esmolol | Brevibloc Premixed 100mg/ 10ml solution for injection vials (Baxter Healthcare Ltd) |
| 30541 | 71225001000027104 | Esmolol | Esmolol 250mg/ ml concentrate solution for infusion |
| 32135 | 10070411000001106 | Esmolol | Brevibloc Concentrate 2.5g/ 10ml solution for infusion ampoules (Baxter Healthcare Ltd) |
| 39819 | 17033711000001101 | Esmolol | Esmolol 2.5g/ 250ml infusion bags |
| 4265 | 157565001000027107 | Celiprolol | Celectol 200mg Tablet (Pantheon Healthcare Ltd) |
| 7974 | 318622003 | Celiprolol | Celiprolol 400mg tablets |
| 8262 | 318619000 | Celiprolol | Celiprolol 200mg tablets |
| 16776 | 157575001000027100 | Celiprolol | Celectol 400mg Tablet (Pantheon Healthcare Ltd) |
| 35054 | 864811000001100 | Celiprolol | Celectol 200mg tablets (Zentiva) |
| 35940 | 479211000001100 | Celiprolol | Celectol 400mg tablets (Zentiva) |
| 41740 | 749111000001104 | Celiprolol | Celiprolol 200mg tablets (Teva UK Ltd) |
| 42795 | 354811000001105 | Celiprolol | Celiprolol 200mg tablets (Mylan) |
| 56485 | 11028811000001103 | Celiprolol | Celectol 200mg tablets (Waymade Healthcare Plc) |
| 57573 | 5530311000001105 | Celiprolol | Celectol 200mg tablets (Dowelhurst Ltd) |
| 67292 | 14493011000001107 | Celiprolol | Celectol 200mg tablets (Sigma Pharmaceuticals Plc) |
| 74062 | 14493611000001100 | Celiprolol | Celectol 400mg tablets (Sigma Pharmaceuticals Plc) |
| 74623 | 684111000001100 | Celiprolol | Celiprolol 400mg tablets (Mylan) |
| 817 | 318633000 | Carvedilol | Carvedilol 3.125mg tablets |
| 2629 | 318631003 | Carvedilol | Carvedilol 12.5mg tablets |
| 4410 | 318635007 | Carvedilol | Carvedilol 6.25mg tablets |
| 7049 | 318632005 | Carvedilol | Carvedilol 25mg tablets |
| 14117 | 873111000001104 | Carvedilol | Eucardic 3.125mg tablets (Roche Products Ltd) |
| 14146 | 690511000001101 | Carvedilol | Eucardic 6.25mg tablets (Roche Products Ltd) |
| 18414 | 334711000001101 | Carvedilol | Eucardic 12.5mg tablets (Roche Products Ltd) |
| 19202 | 7333911000001104 | Carvedilol | Carvedilol 6.25mg tablets (Teva UK Ltd) |
| 19437 | 709911000001107 | Carvedilol | Eucardic 25mg tablets (Roche Products Ltd) |
| 33374 | 9034211000001104 | Carvedilol | Carvedilol 12.5mg tablets (Genus Pharmaceuticals Ltd) |
| 34501 | 7332611000001100 | Carvedilol | Carvedilol 12.5mg tablets (Actavis UK Ltd) |
| 34740 | 7332411000001103 | Carvedilol | Carvedilol 6.25mg tablets (Actavis UK Ltd) |
| 34741 | 7402011000001106 | Carvedilol | Carvedilol 3.125mg tablets (IVAX Pharmaceuticals UK Ltd) |
| 46935 | 7398911000001101 | Carvedilol | Carvedilol 3.125mg tablets (Actavis UK Ltd) |
| 46936 | 7493711000001102 | Carvedilol | Carvedilol 3.125mg tablets (A A H Pharmaceuticals Ltd) |
| 47107 | 8356811000001107 | Carvedilol | Carvedilol 5mg/ 5ml oral suspension |
| 49142 | 12422611000001106 | Carvedilol | Carvedilol 3.125mg/ 5ml oral suspension |
| 54106 | 14012311000001106 | Carvedilol | Carvedilol 1.5mg/ 5ml oral suspension |
| 59549 | 8356811000001107 | Carvedilol | Carvedilol 5mg/ 5ml oral suspension |
| 61663 | 7333711000001101 | Carvedilol | Carvedilol 3.125mg tablets (Teva UK Ltd) |
| 63422 | 21820811000001101 | Carvedilol | Carvedilol 12.5mg tablets (Waymade Healthcare Plc) |

| | | | |
|---|---|---|---|
| 67661 | 15072711000001108 | Carvedilol | Carvedilol 6.25mg tablets (Sigma Pharmaceuticals Plc) |
| 72507 | 7334311000001103 | Carvedilol | Carvedilol 25mg tablets (Teva UK Ltd) |
| 73451 | 11401811000001105 | Carvedilol | Carvedilol 6.25mg tablets (Almus Pharmaceuticals Ltd) |
| 74619 | 7334111000001100 | Carvedilol | Carvedilol 12.5mg tablets (Teva UK Ltd) |
| 77648 | | Carvedilol | Carvedilol 12.5mg tablets (Almus Pharmaceuticals Ltd) |
| 472 | 318590006 | Bisoprolol fumarate | Bisoprolol 5mg tablets |
| 594 | 318605000 | Bisoprolol fumarate | Bisoprolol 2.5mg tablets |
| 599 | 318604001 | Bisoprolol fumarate | Bisoprolol 1.25mg tablets |
| 822 | 243085001000027105 | Bisoprolol Fumarate | Bisoprolol 1.5mg/ 5ml oral suspension |
| 1290 | 318591005 | Bisoprolol fumarate | Bisoprolol 10mg tablets |
| 3588 | 349611000001108 | Bisoprolol fumarate | Monocor 5mg tablets (Wyeth Pharmaceuticals) |
| 4771 | 483911000001101 | Bisoprolol fumarate | Emcor LS 5mg tablets (Merck Serono Ltd) |
| 5713 | 318607008 | Bisoprolol fumarate | Bisoprolol 7.5mg tablets |
| 5968 | 329611000001106 | Bisoprolol fumarate | Monocor 10mg tablets (Wyeth Pharmaceuticals) |
| 7091 | 318606004 | Bisoprolol fumarate | Bisoprolol 3.75mg tablets |
| 7553 | 8304811000001108 | Bisoprolol fumarate | Bisoprolol 5mg/ 5ml oral suspension |
| 10892 | 106611000001104 | Bisoprolol fumarate | Emcor 10mg tablets (Merck Serono Ltd) |
| 14030 | 628711000001102 | Bisoprolol fumarate | Cardicor 2.5mg tablets (Merck Serono Ltd) |
| 14058 | 712511000001106 | Bisoprolol fumarate | Cardicor 1.25mg tablets (Merck Serono Ltd) |
| 17615 | 19711000001106 | Bisoprolol fumarate | Cardicor 5mg tablets (Merck Serono Ltd) |
| 18185 | 404511000001105 | Bisoprolol fumarate | Cardicor 7.5mg tablets (Merck Serono Ltd) |
| 19178 | 78411000001104 | Bisoprolol fumarate | Bisoprolol 10mg tablets (Ranbaxy (UK) Ltd) |
| 19200 | 433811000001104 | Bisoprolol fumarate | Bisoprolol 5mg tablets (IVAX Pharmaceuticals UK Ltd) |
| 19853 | 823111000001103 | Bisoprolol fumarate | Cardicor 3.75mg tablets (Merck Serono Ltd) |
| 19858 | 890111000001100 | Bisoprolol fumarate | Cardicor 10mg tablets (Merck Serono Ltd) |
| 21905 | 405411000001107 | Bisoprolol fumarate | Bipranix 10mg tablets (Ashbourne Pharmaceuticals Ltd) |
| 21966 | 501211000001103 | Bisoprolol fumarate | Bipranix 5mg tablets (Ashbourne Pharmaceuticals Ltd) |
| 24083 | 853911000001104 | Bisoprolol fumarate | Bisoprolol 5mg tablets (Teva UK Ltd) |
| 32114 | 573811000001107 | Bisoprolol fumarate | Bisoprolol 5mg tablets (Mylan) |
| 32552 | 10241511000001108 | Bisoprolol fumarate | Congescor 2.5mg tablets (Tillomed Laboratories Ltd) |
| 32630 | 9739311000001105 | Bisoprolol fumarate | Vivacor 10mg tablets (Lexon (UK) Ltd) |
| 33839 | 13211000001101 | Bisoprolol fumarate | Bisoprolol 10mg tablets (Actavis UK Ltd) |
| 33909 | 10241311000001102 | Bisoprolol fumarate | Congescor 1.25mg tablets (Tillomed Laboratories Ltd) |
| 34821 | 502311000001102 | Bisoprolol fumarate | Bisoprolol 10mg tablets (Mylan) |
| 34963 | 706111000001101 | Bisoprolol fumarate | Bisoprolol 5mg tablets (Actavis UK Ltd) |
| 37118 | 10743711000001105 | Bisoprolol fumarate | Bisoprolol 2.5mg tablets (A A H Pharmaceuticals Ltd) |
| 37837 | 187815001000027109 | Bisoprolol fumarate | Bisoprolol 2.5mg Tablet (Teva UK Ltd) |
| 38991 | 13919811000001103 | Bisoprolol fumarate | Bisoprolol 7.5mg tablets (A A H Pharmaceuticals Ltd) |
| 39646 | 256485001000027100 | Bisoprolol Fumarate | Bisoprolol 0.625mg/ 5ml oral solution |
| 39846 | 9105811000001100 | Bisoprolol fumarate | Vivacor 5mg tablets (Lexon (UK) Ltd) |
| 41591 | 888911000001101 | Bisoprolol fumarate | Bisoprolol 10mg tablets (Teva UK Ltd) |
| 43251 | 13436011000001108 | Bisoprolol fumarate | Bisoprolol 1.25mg tablets (Mylan) |
| 43564 | 112235001000027100 | Bisoprolol fumarate | Bisoprolol 5mg Tablet (PLIVA Pharma Ltd) |

| | | | |
|---|---|---|---|
| 44000 | 12287611000001100 | Bisoprolol fumarate | Bisoprolol 2.5mg/ 5ml oral suspension |
| 47041 | 13436211000001103 | Bisoprolol fumarate | Bisoprolol 2.5mg tablets (Mylan) |
| 50224 | 20475811000001108 | Bisoprolol fumarate | Congescor 2.5mg tablets (Teva UK Ltd) |
| 50300 | 20475611000001109 | Bisoprolol fumarate | Congescor 1.25mg tablets (Teva UK Ltd) |
| 50403 | 187775001000027103 | Bisoprolol fumarate | Bisoprolol 1.25mg Tablet (Teva UK Ltd) |
| 50514 | 17012411000001106 | Bisoprolol fumarate | Bisoprolol 2.5mg tablets (Chanelle Medical UK Ltd) |
| 51528 | 20358011000001102 | Bisoprolol fumarate | Bisoprolol 1.25mg tablets (Actavis UK Ltd) |
| 52548 | 18506211000001103 | Bisoprolol fumarate | Bisoprolol 1.25mg tablets (Almus Pharmaceuticals Ltd) |
| 52611 | 12287311000001105 | Bisoprolol fumarate | Bisoprolol 10mg/ 5ml oral solution |
| 52635 | 557311000001108 | Bisoprolol fumarate | Bisoprolol 5mg tablets (Alliance Healthcare (Distribution) Ltd) |
| 52686 | 12287511000001104 | Bisoprolol fumarate | Bisoprolol 2.5mg/ 5ml oral solution |
| 53334 | 632011000001104 | Bisoprolol fumarate | Bisoprolol 10mg tablets (A A H Pharmaceuticals Ltd) |
| 53664 | 20283611000001102 | Bisoprolol fumarate | Bisoprolol 2.5mg tablets (Sandoz Ltd) |
| 53885 | 11252511000001108 | Bisoprolol fumarate | Bisoprolol 1.25mg tablets (A A H Pharmaceuticals Ltd) |
| 53916 | 18506411000001104 | Bisoprolol fumarate | Bisoprolol 2.5mg tablets (Almus Pharmaceuticals Ltd) |
| 54479 | 10740611000001105 | Bisoprolol fumarate | Bisoprolol 1.25mg tablets (Alliance Healthcare (Distribution) Ltd) |
| 55298 | 15058011000001107 | Bisoprolol fumarate | Bisoprolol 10mg tablets (Sigma Pharmaceuticals Plc) |
| 55791 | 20357211000001108 | Bisoprolol fumarate | Bisoprolol 3.75mg tablets (Actavis UK Ltd) |
| 55929 | 21281511000001108 | Bisoprolol fumarate | Bisoprolol 5mg tablets (Accord Healthcare Ltd) |
| 56240 | 20283811000001103 | Bisoprolol fumarate | Bisoprolol 3.75mg tablets (Sandoz Ltd) |
| 56459 | 21281311000001102 | Bisoprolol fumarate | Bisoprolol 2.5mg tablets (Accord Healthcare Ltd) |
| 56486 | 5582411000001107 | Bisoprolol fumarate | Monocor 10mg tablets (Dowelhurst Ltd) |
| 56768 | 18553111000001106 | Bisoprolol fumarate | Bisoprolol 2.5mg tablets (Niche Generics Ltd) |
| 57023 | 18506411000001104 | Bisoprolol fumarate | Bisoprolol 2.5mg tablets (Almus Pharmaceuticals Ltd) |
| 57176 | 21281711000001103 | Bisoprolol fumarate | Bisoprolol 10mg tablets (Accord Healthcare Ltd) |
| 57578 | 17619411000001108 | Bisoprolol fumarate | Cardicor 2.5mg tablets (Necessity Supplies Ltd) |
| 57626 | 8305311000001100 | Bisoprolol fumarate | Bisoprolol 1.25mg/ 5ml oral solution |
| 57934 | 7376711000001106 | Bisoprolol fumarate | Bisoprolol 5mg tablets (Sandoz Ltd) |
| 58109 | 8305211000001108 | Bisoprolol fumarate | Bisoprolol 1.25mg/ 5ml oral suspension |
| 58455 | 20284011000001106 | Bisoprolol fumarate | Bisoprolol 7.5mg tablets (Sandoz Ltd) |
| 58498 | 22496711000001105 | Bisoprolol fumarate | Bisoprolol 2.5mg tablets (Medreich Plc) |
| 58511 | 20283411000001100 | Bisoprolol fumarate | Bisoprolol 1.25mg tablets (Sandoz Ltd) |
| 58763 | 21802011000001102 | Bisoprolol fumarate | Bisoprolol 2.5mg tablets (Waymade Healthcare Plc) |
| 58973 | 10437911000001106 | Bisoprolol fumarate | Bisoprolol 10mg tablets (Niche Generics Ltd) |
| 58974 | 10740811000001109 | Bisoprolol fumarate | Bisoprolol 2.5mg tablets (Alliance Healthcare (Distribution) Ltd) |
| 58982 | 19734511000001102 | Bisoprolol fumarate | Bisoprolol 10mg tablets (Medreich Plc) |
| 59037 | 249811000001104 | Bisoprolol fumarate | Bisoprolol 5mg tablets (A A H Pharmaceuticals Ltd) |
| 59148 | 20577811000001109 | Bisoprolol fumarate | Bisoprolol 2.5mg tablets (Zentiva) |
| 59495 | 21105111000001104 | Bisoprolol fumarate | Bisoprolol 1.25mg tablets (Teva UK Ltd) |
| 59969 | 18497711000001100 | Bisoprolol fumarate | Bisoprolol 5mg tablets (Almus Pharmaceuticals Ltd) |
| 60502 | 19708211000001104 | Bisoprolol fumarate | Bisoprolol 3.75mg tablets (DE Pharmaceuticals) |
| 60761 | 22495911000001100 | Bisoprolol fumarate | Bisoprolol 1.25mg tablets (Medreich Plc) |
| 60896 | 19734211000001100 | Bisoprolol fumarate | Bisoprolol 5mg tablets (Medreich Plc) |

| | | | |
|---|---|---|---|
| 61115 | 8304911000001103 | Bisoprolol fumarate | Bisoprolol 5mg/ 5ml oral solution |
| 61340 | 19708411000001100 | Bisoprolol fumarate | Bisoprolol 5mg tablets (DE Pharmaceuticals) |
| 61564 | 21802311000001104 | Bisoprolol fumarate | Bisoprolol 3.75mg tablets (Waymade Healthcare Plc) |
| 61651 | 18506811000001102 | Bisoprolol fumarate | Bisoprolol 7.5mg tablets (Almus Pharmaceuticals Ltd) |
| 62361 | 17012211000001107 | Bisoprolol fumarate | Bisoprolol 1.25mg tablets (Chanelle Medical UK Ltd) |
| 62407 | 250065001000027102 | Bisoprolol Fumarate | Bisoprolol oral solution |
| 63493 | 17328511000001103 | Bisoprolol fumarate | Bisoprolol 2.5mg tablets (Actavis UK Ltd) |
| 63535 | 10285811000001100 | Bisoprolol fumarate | Bisoprolol 5mg tablets (Relonchem Ltd) |
| 63850 | 21108211000001103 | Bisoprolol fumarate | Bisoprolol 2.5mg tablets (Teva UK Ltd) |
| 64538 | 18743711000001108 | Bisoprolol fumarate | Bisoprolol 3.75mg tablets (Teva UK Ltd) |
| 64784 | 10438411000001104 | Bisoprolol fumarate | Bisoprolol 5mg tablets (Niche Generics Ltd) |
| 64850 | 19708011000001109 | Bisoprolol fumarate | Bisoprolol 2.5mg tablets (DE Pharmaceuticals) |
| 65805 | 21800411000001105 | Bisoprolol fumarate | Bisoprolol 1.25mg tablets (Waymade Healthcare Plc) |
| 65821 | 14159111000001101 | Bisoprolol fumarate | Bisoprolol 7.5mg/ 5ml oral suspension |
| 69156 | 22495711000001102 | Bisoprolol fumarate | Bisoprolol 3.75mg tablets (Medreich Plc) |
| 71472 | 21801411000001101 | Bisoprolol fumarate | Bisoprolol 5mg tablets (Waymade Healthcare Plc) |
| 72285 | 30001311000001100 | Bisoprolol fumarate | Bisoprolol 1.25mg tablets (Mawdsley-Brooks & Company Ltd) |
| 72540 | 12287411000001103 | Bisoprolol fumarate | Bisoprolol 10mg/ 5ml oral suspension |
| 73638 | 18497711000001100 | Bisoprolol fumarate | Bisoprolol 5mg tablets (Almus Pharmaceuticals Ltd) |
| 73641 | 18497911000001103 | Bisoprolol fumarate | Bisoprolol 10mg tablets (Almus Pharmaceuticals Ltd) |
| 73809 | 18506211000001103 | Bisoprolol fumarate | Bisoprolol 1.25mg tablets (Almus Pharmaceuticals Ltd) |
| 74742 | 15160311000001108 | Bisoprolol fumarate | Bisoprolol 625micrograms/ 5ml oral suspension |
| 75463 | | Bisoprolol fumarate | Bisoprolol 625micrograms/ 5ml oral solution |
| 76299 | | Bisoprolol fumarate | Bisoprolol 10mg tablets (Kent Pharmaceuticals Ltd) |
| 76322 | | Bisoprolol fumarate | Bisoprolol 10mg tablets (Almus Pharmaceuticals Ltd) |
| 76483 | | Bisoprolol fumarate | Bisoprolol 1.25mg tablets (Sigma Pharmaceuticals Plc) |
| 77219 | | Bisoprolol fumarate | Bisoprolol 7.5mg tablets (Waymade Healthcare Plc) |
| 5 | 318420003 | Atenolol | Atenolol 50mg tablets |
| 24 | 318421004 | Atenolol | Atenolol 100mg tablets |
| 26 | 318434003 | Atenolol | Atenolol 25mg tablets |
| 197 | 35903411000001106 | Atenolol | Atenolol 5mg/ 10ml solution for injection ampoules |
| 2432 | 423911000001107 | Atenolol | Tenormin LS 50mg tablets (AstraZeneca UK Ltd) |
| 2587 | 162411000001102 | Atenolol | Tenormin 100mg tablets (AstraZeneca UK Ltd) |
| 2590 | 317111000001101 | Atenolol | Tenormin 25mg tablets (AstraZeneca UK Ltd) |
| 6066 | 35903211000001107 | Atenolol | Atenolol 25mg/ 5ml oral solution sugar free |
| 7429 | 9111000001107 | Atenolol | Tenormin 5mg/ 10ml solution for injection ampoules (AstraZeneca UK Ltd) |
| 10191 | 271911000001106 | Atenolol | Atenix 50 tablets (Ashbourne Pharmaceuticals Ltd) |
| 13394 | 373311000001100 | Atenolol | Tenormin 25mg/ 5ml syrup (AstraZeneca UK Ltd) |
| 15176 | 182175001000027108 | Atenolol | Totamol 50mg Tablet (C P Pharmaceuticals Ltd) |
| 15730 | 182185001000027103 | Atenolol | Totamol 100mg Tablet (C P Pharmaceuticals Ltd) |
| 17322 | 482511000001101 | Atenolol | Atenix 25 tablets (Ashbourne Pharmaceuticals Ltd) |
| 18950 | 182195001000027104 | Atenolol | Totamol 25mg Tablet (C P Pharmaceuticals Ltd) |
| 19172 | 600811000001101 | Atenolol | Atenolol 25mg tablets (IVAX Pharmaceuticals UK Ltd) |

| | | | |
|---|---|---|---|
| 19182 | 852111000001108 | Atenolol | Atenolol 50mg tablets (IVAX Pharmaceuticals UK Ltd) |
| 19191 | 857511000001101 | Atenolol | Atenolol 100mg tablets (Teva UK Ltd) |
| 20502 | 181611000001107 | Atenolol | Atenix 100 tablets (Ashbourne Pharmaceuticals Ltd) |
| 20728 | 234895001000027106 | Atenolol | Atenamin 25mg Tablet (OPD Pharm) |
| 21133 | 234905001000027103 | Atenolol | Atenamin 50mg Tablet (OPD Pharm) |
| 24191 | 874711000001101 | Atenolol | Antipressan 50mg tablets (Teva UK Ltd) |
| 24195 | 734011000001102 | Atenolol | Antipressan 100mg tablets (Teva UK Ltd) |
| 26211 | 877011000001106 | Atenolol | Antipressan 25mg tablets (Teva UK Ltd) |
| 29368 | 623811000001100 | Atenolol | Atenolol 25mg tablets (Teva UK Ltd) |
| 29398 | 234915001000027101 | Atenolol | Atenamin 100mg Tablet (OPD Pharm) |
| 30636 | 182145001000027107 | Atenolol | Vasaten 50mg Tablet (Shire Pharmaceuticals Ltd) |
| 31536 | 907211000001102 | Atenolol | Atenolol 25mg tablets (Kent Pharmaceuticals Ltd) |
| 31934 | 646611000001108 | Atenolol | Atenolol 100mg tablets (IVAX Pharmaceuticals UK Ltd) |
| 33079 | 35311000001104 | Atenolol | Atenolol 100mg tablets (Mylan) |
| 33085 | 422011000001104 | Atenolol | Atenolol 100mg tablets (A A H Pharmaceuticals Ltd) |
| 33092 | 58311000001109 | Atenolol | Atenolol 50mg tablets (A A H Pharmaceuticals Ltd) |
| 33184 | 417811000001102 | Atenolol | Atenolol 100mg tablets (Wockhardt UK Ltd) |
| 33650 | 225111000001106 | Atenolol | Atenolol 50mg tablets (Mylan) |
| 33657 | 10811000001107 | Atenolol | Atenolol 25mg tablets (A A H Pharmaceuticals Ltd) |
| 33850 | 401411000001109 | Atenolol | Atenolol 50mg tablets (Actavis UK Ltd) |
| 34265 | 884011000001109 | Atenolol | Atenolol 50mg tablets (Sandoz Ltd) |
| 34365 | 674511000001104 | Atenolol | Atenolol 50mg tablets (Teva UK Ltd) |
| 34443 | 335611000001106 | Atenolol | Atenolol 50mg tablets (Wockhardt UK Ltd) |
| 34492 | 774211000001105 | Atenolol | Atenolol 25mg tablets (Mylan) |
| 34575 | 375411000001106 | Atenolol | Atenolol 25mg tablets (Wockhardt UK Ltd) |
| 34585 | 393011000001109 | Atenolol | Atenolol 25mg tablets (Sandoz Ltd) |
| 34695 | 790911000001104 | Atenolol | Atenolol 50mg tablets (Kent Pharmaceuticals Ltd) |
| 34754 | 282111000001106 | Atenolol | Atenolol 100mg tablets (Sandoz Ltd) |
| 34882 | 38985001000027109 | Atenolol | Atenolol 50mg Tablet (Berk Pharmaceuticals Ltd) |
| 34976 | 13740111000001101 | Atenolol | Atenolol 25mg tablets (Tillomed Laboratories Ltd) |
| 36261 | 13740311000001104 | Atenolol | Atenolol 50mg tablets (Tillomed Laboratories Ltd) |
| 44858 | 244111000001105 | Atenolol | Atenolol 25mg tablets (Actavis UK Ltd) |
| 46908 | 721711000001100 | Atenolol | Atenolol 100mg tablets (Kent Pharmaceuticals Ltd) |
| 46931 | 275811000001103 | Atenolol | Atenolol 100mg tablets (Actavis UK Ltd) |
| 47870 | 9791011000001101 | Atenolol | Atenolol 25mg tablets (Almus Pharmaceuticals Ltd) |
| 49953 | 15986211000001103 | Atenolol | Atenolol 25mg tablets (Bristol Laboratories Ltd) |
| 50702 | 47711000001105 | Atenolol | Atenolol 25mg tablets (Alliance Healthcare (Distribution) Ltd) |
| 51643 | 11560811000001105 | Atenolol | Atenolol 25mg/ 5ml oral solution sugar free (Alliance Healthcare (Distribution) Ltd) |
| 51998 | 18280111000001102 | Atenolol | Atenolol 25mg tablets (Strides Shasun (UK) Ltd) |
| 52310 | 20137611000001100 | Atenolol | Atenolol 25mg tablets (Crescent Pharma Ltd) |
| 52500 | 9791711000001104 | Atenolol | Atenolol 50mg tablets (Almus Pharmaceuticals Ltd) |
| 53204 | 263511000001107 | Atenolol | Atenolol 50mg tablets (Alliance Healthcare (Distribution) Ltd) |
| 53215 | 15986411000001104 | Atenolol | Atenolol 50mg tablets (Bristol Laboratories Ltd) |

| | | | |
|---|---|---|---|
| 53414 | 18458611000001100 | Atenolol | Atenolol 50mg tablets (Accord Healthcare Ltd) |
| 53802 | 15070211000001102 | Atenolol | Atenolol 25mg tablets (Sigma Pharmaceuticals Plc) |
| 53826 | 14801811000001107 | Atenolol | Atenolol 25mg tablets (Boston Healthcare Ltd) |
| 54542 | 15968011000001105 | Atenolol | Atenolol 25mg tablets (Zanza Laboratories Ltd) |
| 54752 | 18279911000001104 | Atenolol | Atenolol 50mg tablets (Strides Shasun (UK) Ltd) |
| 55778 | 17788311000001101 | Atenolol | Atenolol 50mg tablets (Phoenix Healthcare Distribution Ltd) |
| 56445 | 16183211000001105 | Atenolol | Atenolol 25mg/ 5ml oral solution sugar free (A A H Pharmaceuticals Ltd) |
| 57817 | 11177311000001101 | Atenolol | Atenolol 50mg tablets (Zentiva) |
| 59695 | 14802011000001109 | Atenolol | Atenolol 50mg tablets (Boston Healthcare Ltd) |
| 59982 | 18458411000001103 | Atenolol | Atenolol 25mg tablets (Accord Healthcare Ltd) |
| 61573 | 12015011000001105 | Atenolol | Atenolol 25mg/ 5ml oral solution |
| 62325 | 21778611000001100 | Atenolol | Atenolol 25mg tablets (Waymade Healthcare Plc) |
| 64973 | 29771711000001105 | Atenolol | Atenolol 50mg tablets (Sigma Pharmaceuticals Plc) |
| 66548 | 22612611000001101 | Atenolol | Atenolol 50mg tablets (DE Pharmaceuticals) |
| 69526 | 12079211000001106 | Atenolol | Atenolol 50mg/ 5ml oral solution |
| 70135 | 22612411000001104 | Atenolol | Atenolol 25mg tablets (DE Pharmaceuticals) |
| 71026 | 11399511000001103 | Atenolol | Atenolol 100mg tablets (Almus Pharmaceuticals Ltd) |
| 72043 | 21778811000001101 | Atenolol | Atenolol 50mg tablets (Waymade Healthcare Plc) |
| 72810 | 12014311000001109 | Atenolol | Atenolol 10mg/ 5ml oral solution |
| 73151 | 28945211000001109 | Atenolol | Atenolol 50mg tablets (Crescent Pharma Ltd) |
| 76593 | | Atenolol | Atenolol 25mg/ 5ml oral solution sugar free (DE Pharmaceuticals) |
| 77198 | | Atenolol | Atenolol 100mg tablets (Phoenix Healthcare Distribution Ltd) |
| 77613 | | Atenolol | Atenolol 25mg/ 5ml oral suspension |
| 7620 | 318414004 | Acebutolol | Acebutolol 400mg tablets |
| 8023 | 298111000001105 | Acebutolol | Sectral 400mg tablets (Sanofi) |
| 8113 | 318413005 | Acebutolol | Acebutolol 200mg capsules |
| 8172 | 318412000 | Acebutolol | Acebutolol 100mg capsules |
| 8555 | 925711000001108 | Acebutolol | Sectral 200mg capsules (Sanofi) |
| 12296 | 632811000001105 | Acebutolol | Sectral 100mg capsules (Sanofi) |
| 45309 | 9703611000001105 | Acebutolol | Acebutolol 400mg tablets (A A H Pharmaceuticals Ltd) |
| 65438 | 9703211000001108 | Acebutolol | Acebutolol 100mg capsules (A A H Pharmaceuticals Ltd) |
| 72545 | 9703411000001107 | Acebutolol | Acebutolol 200mg capsules (A A H Pharmaceuticals Ltd) |
| 77435 | | Acebutolol | Sectral 400mg tablets (Mawdsley-Brooks & Company Ltd) |
| 3041 | 108205001000027101 | | SOTALOL 40 MG INJ |
| 4021 | 177775001000027104 | | PROPRANOLOL 20 MG TAB |
| 7491 | 177255001000027105 | | LABETALOL TAB |
| 12119 | 132615001000027106 | | SOTALOL S/ R 80 MG TAB |
| 12497 | 166645001000027109 | | OXPRENOLOL 10 MG TAB |
| 16669 | 72615001000027105 | | LOPRESOR SR 200 MG TAB |
| 17876 | 112415001000027107 | | METOPROLOL FUMARATE 190 MG TAB |
| 18722 | | | CARVEDILOL |
| 19810 | 181045001000027103 | | BEDRANOL SR 80 MG CAP |
| 20813 | | | BETALOC S.A. |

| | | | |
|---|---|---|---|
| 22634 | 135155001000027102 | | PROPRANOLOL 10 MG SUS |
| 22796 | | | CARVEDILOL 3.125 MG |
| 23598 | | | SOTALOL S/ R |
| 23604 | | | PROPRANOLOL S/ R |
| 24378 | | | BETALOC S.A. (CALENDAR PACK) |
| 24677 | | | ATENOLOL |
| 25037 | | | ATENOLOL |
| 25052 | | | HALF-INDERAL LA |
| 25818 | 164975001000027100 | | PROPRANOLOL 30 MG SUS |
| 26105 | 165015001000027106 | | PROPRANOLOL paed 4 MG TAB |
| 26290 | 99325001000027104 | | SOTACOR 40 MG INJ |
| 26788 | 135175001000027104 | | PROPRANOLOL 2.5 MG ELI |
| 27036 | 164995001000027109 | | PROPRANOLOL POWDERS 5 MG POW |
| 28493 | 100955001000027108 | | METOPROLOL FUMARATE 95 MG TAB |
| 29803 | 135135001000027101 | | PROPRANOLOL 3 MG ELI |
| 32470 | 135035001000027107 | | PROPRANOLOL 1 MG LIQ |
| 44310 | | | INDERAL |
| 46493 | 135125001000027103 | | PROPRANOLOL 15 MG SYR |
| **Betablockers in combination with calcium channel blockers** | | | |
| 15117 | 156825001000027102 | Nifedipine/ Atenolol | Nifedipine with atenolol 20mg + 50mg Capsule |
| 1684 | 3142711000001107 | Atenolol/ Nifedipine | Beta-Adalat modified-release capsules (Bayer Plc) |
| 4542 | 35903311000001104 | Atenolol/ Nifedipine | Atenolol 50mg / Nifedipine 20mg modified-release capsules |
| 8642 | 3142511000001102 | Atenolol/ Nifedipine | Tenif 50mg/ 20mg modified-release capsules (AstraZeneca UK Ltd) |
| 52728 | 16141111000001106 | Atenolol/ Nifedipine | Beta-Adalat modified-release capsules (Lexon (UK) Ltd) |
| 61719 | 10490311000001102 | Atenolol/ Nifedipine | Beta-Adalat modified-release capsules (Waymade Healthcare Plc) |
| 68020 | 14208611000001108 | Atenolol/ Nifedipine | Beta-Adalat modified-release capsules (Sigma Pharmaceuticals Plc) |
| 74039 | 14625611000001108 | Atenolol/ Nifedipine | Tenif 50mg/ 20mg modified-release capsules (Sigma Pharmaceuticals Plc) |
| 76440 | | Atenolol/ Nifedipine | Beta-Adalat modified-release capsules (Dowelhurst Ltd) |
| **Betablockers and Diuretics** | | | |
| 8623 | 38405001000027107 | Timolol maleate/ Bendroflumethiazide | Prestim Tablet (ICN Pharmaceuticals France S.A.) |
| 12517 | 144255001000027109 | Timolol Maleate/ Bendroflumethiazide | Timolol maleate with bendroflumethiazide 20mg + 5mg Tablet |
| 12651 | 318556002 | Timolol maleate/ Bendroflumethiazide | Timolol 10mg / Bendroflumethiazide 2.5mg tablets |
| 19142 | 148855001000027103 | Timolol Maleate/ Bendroflumethiazide | Bendroflumethiazide 2.5mg with Timolol maleate 10mg tablets |
| 21025 | 144325001000027103 | Timolol Maleate/ Bendroflumethiazide | Prestim forte Tablet (LEO Pharma) |
| 25363 | 98411000001109 | Timolol maleate/ Bendroflumethiazide | Prestim tablets (Meda Pharmaceuticals Ltd) |
| 3691 | 141295001000027106 | Sotalol / Hydrochlorothiazide | Sotalol 160mg with hydrochlorothiazide 25mg tablet |

| | | | |
|---|---|---|---|
| 8061 | 141285001000027107 | Sotalol / Hydrochlorothiazide | Sotalol 80mg with hydrochlorothiazide 12.5mg tablet |
| 12456 | 29205001000027105 | Sotalol / Hydrochlorothiazide | Sotazide Tablet (Bristol-Myers Squibb Pharmaceuticals Ltd) |
| 15042 | 141405001000027109 | Sotalol / Hydrochlorothiazide | Tolerzide Tablet (Bristol-Myers Squibb Pharmaceuticals Ltd) |
| 17783 | 75195001000027107 | Propranolol / Spironolactone | Spiroprop Tablet (Pharmacia Ltd) |
| 4796 | 333111000001104 | Propranolol / Bendroflumethiazide | Inderetic 80mg/ 2.5mg capsules (AstraZeneca UK Ltd) |
| 8369 | 350811000001100 | Propranolol / Bendroflumethiazide | Inderex 160mg/ 5mg modified-release capsules (AstraZeneca UK Ltd) |
| 8987 | 35932411000001108 | Propranolol / Bendroflumethiazide | Propranolol 160mg modified-release / Bendroflumethiazide 5mg capsules |
| 12054 | 318584007 | Propranolol / Bendroflumethiazide | Propranolol 80mg / Bendroflumethiazide 2.5mg capsules |
| 22912 | 148735001000027104 | Propranolol / Bendroflumethiazide | Bendroflumethiazide 2.5mg with Propanolol 80mg capsules |
| 23131 | 148745001000027103 | Propranolol / Bendroflumethiazide | Bendroflumethiazide 5mg with Propanolol 160mg modified-release capsules |
| 25462 | 103335001000027100 | Pindolol/ Clopamide | Clopamide 5mg with Pindolol 10mg tablets |
| 77387 | | Pindolol/ Clopamide | Viskaldix tablets (Waymade Healthcare Plc) |
| 24832 | 39675001000027101 | Penbutolol/ Furosemide | Lasipressin Tablet (Hoechst UK Ltd) |
| 26529 | 108015001000027108 | Penbutolol/ Furosemide | Furosemide with penbutolol Tablet |
| 4429 | 3444411000001107 | Oxprenolol / Cyclopenthiazide | Trasidrex modified-release tablets (Mercury Pharma Group Ltd) |
| 8673 | 189585001000027103 | Oxprenolol / Cyclopenthiazide | Oxprenolol with cyclopenthiazide 160mg+0.25mg Modified-release tablet |
| 13871 | 36091611000001101 | Oxprenolol / Cyclopenthiazide | Co-prenozide 160mg/ 0.25mg modified-release tablets |
| 52145 | 104605001000027104 | Oxprenolol / Cyclopenthiazide | Cyclopenthiazide 0.25mg with oxprenolol 160mg modified-release tablets |
| 5330 | 3886211000001102 | Nadolol/ Bendroflumethiazide | Corgaretic 40mg tablets (Sanofi-Synthelabo Ltd) |
| 11338 | 148805001000027106 | Nadolol/ Bendroflumethiazide | Bendroflumethiazide 5mg with Nadolol 40mg tablets |
| 14438 | 4057911000001102 | Nadolol/ Bendroflumethiazide | Corgaretic 80mg tablets (Sanofi-Synthelabo Ltd) |
| 23134 | 318549008 | Nadolol/ Bendroflumethiazide | Nadolol 40mg / Bendroflumethiazide 5mg tablets |
| 27946 | 318550008 | Nadolol/ Bendroflumethiazide | Nadolol 80mg / Bendroflumethiazide 5mg tablets |
| 69334 | 148815001000027108 | Nadolol/ Bendroflumethiazide | Bendroflumethiazide 5mg with Nadolol 80mg tablets |
| 7066 | 318546001 | Metoprolol tartrate/ Hydrochlorothiazide | Metoprolol 100mg / Hydrochlorothiazide 12.5mg tablets |
| 10627 | 2977611000001106 | Metoprolol tartrate/ Hydrochlorothiazide | Co-Betaloc tablets (Pfizer Ltd) |
| 18287 | 3853411000001104 | Metoprolol tartrate/ Hydrochlorothiazide | Co-Betaloc SA tablets (Pfizer Ltd) |
| 20093 | 36035311000001101 | Metoprolol tartrate/ Hydrochlorothiazide | Metoprolol 200mg modified-release / Hydrochlorothiazide 25mg tablets |
| 29427 | 157645001000027106 | Metoprolol Tartrate/ Hydrochlorothiazide | Hydrochlorothiazide with metoprolol tartrate 12.5mg with 100mg tablet |
| 33659 | 157635001000027105 | Metoprolol Tartrate/ Hydrochlorothiazide | Hydrochlorothiazide with metoprolol tartrate 25mg with 200mg Modified-release tablet |
| 8147 | 17805001000027109 | Metoprolol Tartrate/ Chlortalidone | Lopresoretic Tablet (Novartis Pharmaceuticals UK Ltd) |
| 15488 | 157605001000027101 | Metoprolol Tartrate/ Chlortalidone | Metoprolol tartrate with chlortalidone Tablet |
| 9143 | 3638411000001101 | Clopamide/ Pindolol | Viskaldix tablets (AMCo) |
| 14057 | 318552000 | Clopamide/ Pindolol | Pindolol 10mg / Clopamide 5mg tablets |
| 581 | 148505001000027107 | Chlortalidone/ Atenolol | Atenolol 50mg with Chlortalidone 12.5mg tablets |
| 1788 | 148515001000027109 | Chlortalidone/ Atenolol | Atenolol 100mg with Chlortalidone 25mg tablets |
| 16786 | 101705001000027101 | Chlortalidone/ Atenolol | Chlortalidone 25mg with Atenolol 100mg tablets |

| | | | |
|---|---|---|---|
| 19055 | 101695001000027105 | Chlortalidone/ Atenolol | Chlortalidone 12.5mg with Atenolol 50mg tablets |
| 17149 | 4542911000001104 | Bisoprolol fumarate/ Hydrochlorothiazide | Monozide 10 tablets (Wyeth Pharmaceuticals) |
| 17462 | 318597009 | Bisoprolol fumarate/ Hydrochlorothiazide | Bisoprolol 10mg / Hydrochlorothiazide 6.25mg tablets |
| 1124 | 288711000001105 | Atenolol/ Chlortalidone | Tenoretic 100mg/ 25mg tablets (AstraZeneca UK Ltd) |
| 1288 | 825011000001103 | Atenolol/ Chlortalidone | Tenoret 50mg/ 12.5mg tablets (AstraZeneca UK Ltd) |
| 5721 | 377211005 | Atenolol/ Chlortalidone | Co-tenidone 100mg/ 25mg tablets |
| 9783 | 318575007 | Atenolol/ Chlortalidone | Co-tenidone 50mg/ 12.5mg tablets |
| 13526 | 101411000001108 | Atenolol/ Chlortalidone | Atenix Co 100 tablets (Ashbourne Pharmaceuticals Ltd) |
| 21873 | 629011000001109 | Atenolol/ Chlortalidone | Atenix Co 50 tablets (Ashbourne Pharmaceuticals Ltd) |
| 24280 | 212825001000027107 | Atenolol/ Chlortalidone | Totaretic 100mg+25mg Tablet (C P Pharmaceuticals Ltd) |
| 26248 | 298311000001107 | Atenolol/ Chlortalidone | Tenchlor 100mg/ 25mg tablets (Teva UK Ltd) |
| 26741 | 212815001000027103 | Atenolol/ Chlortalidone | Totaretic 50mg+12.5mg Tablet (C P Pharmaceuticals Ltd) |
| 31470 | 439411000001104 | Atenolol/ Chlortalidone | Tenchlor 50mg/ 12.5mg tablets (Teva UK Ltd) |
| 31708 | 653811000001101 | Atenolol/ Chlortalidone | Co-tenidone 50mg/ 12.5mg tablets (Actavis UK Ltd) |
| 32094 | 884711000001106 | Atenolol/ Chlortalidone | Co-tenidone 50mg/ 12.5mg tablets (A A H Pharmaceuticals Ltd) |
| 34012 | 134211000001101 | Atenolol/ Chlortalidone | Co-tenidone 100mg/ 25mg tablets (IVAX Pharmaceuticals UK Ltd) |
| 34034 | 576211000001104 | Atenolol/ Chlortalidone | Co-tenidone 50mg/ 12.5mg tablets (IVAX Pharmaceuticals UK Ltd) |
| 34449 | 587011000001102 | Atenolol/ Chlortalidone | Co-tenidone 50mg/ 12.5mg tablets (Mylan) |
| 34825 | 143311000001108 | Atenolol/ Chlortalidone | Co-tenidone 50mg/ 12.5mg tablets (Teva UK Ltd) |
| 34899 | 793911000001107 | Atenolol/ Chlortalidone | Co-tenidone 100mg/ 25mg tablets (A A H Pharmaceuticals Ltd) |
| 37725 | 594311000001107 | Atenolol/ Chlortalidone | Co-tenidone 100mg/ 25mg tablets (Mylan) |
| 41572 | 485811000001105 | Atenolol/ Chlortalidone | Co-tenidone 100mg/ 25mg tablets (Teva UK Ltd) |
| 46952 | 588911000001102 | Atenolol/ Chlortalidone | Co-tenidone 100mg/ 25mg tablets (Actavis UK Ltd) |
| 62537 | 23858011000001104 | Atenolol/ Chlortalidone | Co-tenidone 100mg/ 25mg tablets (DE Pharmaceuticals) |
| 77414 | | Atenolol/ Chlortalidone | Tenoretic 100mg/ 25mg tablets (Dowelhurst Ltd) |
| 77466 | | Atenolol/ Chlortalidone | Co-tenidone 50mg/ 12.5mg tablets (Kent Pharmaceuticals Ltd) |
| 9178 | 318560004 | Atenolol/ Bendroflumethiazide | Atenolol 25mg / Bendroflumethiazide 1.25mg capsules |
| 18743 | 721411000001106 | Atenolol/ Bendroflumethiazide | Tenben 25mg/ 1.25mg capsules (Galen Ltd) |
| 3526 | 159515001000027103 | Atenolol/ Amiloride / Hydrochlorothiazide | Amiloride with atenolol with hydrochlorothiazide capsules |
| 4983 | 156785001000027105 | Atenolol/ Amiloride / Hydrochlorothiazide | Atenolol with amiloride and hydrochlorothiazide capsules |
| 7543 | 237011000001100 | Atenolol/ Amiloride / Hydrochlorothiazide | Kalten capsules (M & A Pharmachem Ltd) |
| 28177 | 156755001000027103 | Atenolol/ Amiloride / Hydrochlorothiazide | Hydrochlorothiazide with atenolol and amiloride Capsule |
| 8189 | 878911000001106 | Acebutolol / Hydrochlorothiazide | Secadrex 200mg/ 12.5mg tablets (Sanofi) |
| 14126 | 318586009 | Acebutolol / Hydrochlorothiazide | Acebutolol 200mg / Hydrochlorothiazide 12.5mg tablets |
| 8765 | 156575001000027104 | | ATENOLOL/ CHLORTHALIDONE 50 MG TAB |
| 19003 | 132525001000027106 | | SPIRONOLACTONE/ PROPRANOLOL 50 MG TAB |
| 22151 | | | METOPROLOL 100MG/ CHLORTHALIDONE 12.5MG |

| | | | |
|---|---|---|---|
| 24520 | 145355001000027108 | | HYDROCHLOROTHIAZIDE / METOPROLOL TARTRATE 25 MG TAB |
| 25764 | | | PINDOLOL 10MG/ CLOPAMIDE 5MG |
| 27086 | 188055001000027109 | | NADOLOL 80MG/ BENDROFLUAZIDE 5MG MG TAB |
| 41892 | | | METOPROLOL 100MG/ HYDROCHLOROTHIAZ.12.5MG |
| **Calcium channel blockers** | | | |
| 700 | 7418511000001104 | Verapamil | Vera-Til SR 120mg tablets (Tillomed Laboratories Ltd) |
| 1118 | 318204006 | Verapamil | Verapamil 40mg tablets |
| 1120 | 318205007 | Verapamil | Verapamil 80mg tablets |
| 1298 | 318213008 | Verapamil | Verapamil 240mg modified-release tablets |
| 1574 | 36565011000001105 | Verapamil | Verapamil 120mg modified-release capsules |
| 1747 | 318206008 | Verapamil | Verapamil 120mg tablets |
| 1748 | 867011000001103 | Verapamil | Cordilox 120mg tablets (IVAX Pharmaceuticals UK Ltd) |
| 3057 | 140411000001109 | Verapamil | Securon 120mg tablets (Abbott Laboratories Ltd) |
| 3342 | 540111000001108 | Verapamil | Securon SR 240mg tablets (Mylan) |
| 3343 | 846611000001102 | Verapamil | Half Securon SR 120mg tablets (Mylan) |
| 3943 | 318250009 | Verapamil | Verapamil 240mg modified-release capsules |
| 6510 | 219711000001103 | Verapamil | Univer 120mg modified-release capsules (Teva UK Ltd) |
| 8524 | 81585001000027101 | Verapamil | Securon 40mg Tablet (Abbott Laboratories Ltd) |
| 8759 | 68645001000027109 | Verapamil | Verapamil 120mg modified release tablets |
| 8884 | 557111000001106 | Verapamil | Cordilox 40mg tablets (IVAX Pharmaceuticals UK Ltd) |
| 8945 | 65411000001102 | Verapamil | Univer 240mg modified-release capsules (Teva UK Ltd) |
| 8975 | 36149411000001103 | Verapamil | Verapamil 180mg modified-release capsules |
| 9569 | 35367911000001103 | Verapamil | Verapamil 120mg modified-release tablets |
| 10688 | 318248001 | Verapamil | Verapamil 160mg tablets |
| 10832 | 81595001000027102 | Verapamil | Securon 80mg Tablet (Abbott Laboratories Ltd) |
| 11777 | 4139411000001106 | Verapamil | Verapamil 40mg/ 5ml oral solution sugar free |
| 11972 | 675611000001104 | Verapamil | Vertab SR 240 tablets (Chiesi Ltd) |
| 12104 | 783511000001101 | Verapamil | Cordilox 160mg tablets (IVAX Pharmaceuticals UK Ltd) |
| 12392 | 391811000001102 | Verapamil | Univer 180mg modified-release capsules (Teva UK Ltd) |
| 13251 | 7418711000001109 | Verapamil | Vera-Til SR 240mg tablets (Tillomed Laboratories Ltd) |
| 13856 | 7855011000001100 | Verapamil | Verapress MR 240mg tablets (Actavis UK Ltd) |
| 13965 | 740811000001102 | Verapamil | Cordilox MR 240mg tablets (Teva UK Ltd) |
| 16328 | 525311000001101 | Verapamil | Verapress MR 240mg tablets (Dexcel-Pharma Ltd) |
| 16677 | 258911000001104 | Verapamil | Cordilox 80mg tablets (IVAX Pharmaceuticals UK Ltd) |
| 17599 | 7852611000001103 | Verapamil | Verapress MR 240mg tablets (Sandoz Ltd) |
| 19175 | 422111000001103 | Verapamil | Verapamil 40mg tablets (IVAX Pharmaceuticals UK Ltd) |
| 19325 | 6105001000027101 | Verapamil | Cordilox 2.5mg/ ml Injection (IVAX Pharmaceuticals UK Ltd) |
| 19457 | 3634111000001100 | Verapamil | Ranvera MR 240mg tablets (Ranbaxy (UK) Ltd) |
| 19459 | 4579511000001103 | Verapamil | Verapamil 240mg modified-release tablets (A A H Pharmaceuticals Ltd) |
| 22826 | 69645001000027106 | Verapamil | Securon 160mg Tablet (Abbott Laboratories Ltd) |
| 23872 | 44155001000027103 | Verapamil | Berkatens 40mg Tablet (Berk Pharmaceuticals Ltd) |
| 25059 | 44165001000027107 | Verapamil | Berkatens 80mg Tablet (Berk Pharmaceuticals Ltd) |

| | | | |
|---|---|---|---|
| 26252 | 44225001000027102 | Verapamil | Berkatens 160mg Tablet (Berk Pharmaceuticals Ltd) |
| 26674 | 36149611000001100 | Verapamil | Verapamil 5mg/ 2ml solution for injection ampoules |
| 27295 | 4399811000001108 | Verapamil | Securon IV 5mg/ 2ml solution for injection ampoules (Mylan) |
| 28843 | 22045001000027100 | Verapamil | Verapamil hc 80mg Tablet (Celltech Pharma Europe Ltd) |
| 28844 | 44175001000027100 | Verapamil | Berkatens 120mg Tablet (Berk Pharmaceuticals Ltd) |
| 29637 | 10910611000001109 | Verapamil | Verapress MR 240mg tablets (Teva UK Ltd) |
| 30462 | 782111000001104 | Verapamil | Ethimil MR 240mg tablets (Genus Pharmaceuticals Ltd) |
| 31490 | 4129511000001107 | Verapamil | Zolvera 40mg/ 5ml oral solution (Rosemont Pharmaceuticals Ltd) |
| 31711 | 100911000001107 | Verapamil | Verapamil 80mg tablets (A A H Pharmaceuticals Ltd) |
| 32590 | 90311000001104 | Verapamil | Verapamil 40mg tablets (Mylan) |
| 33471 | 797711000001100 | Verapamil | Verapamil 40mg tablets (Actavis UK Ltd) |
| 34959 | 413611000001109 | Verapamil | Verapamil 40mg tablets (A A H Pharmaceuticals Ltd) |
| 35729 | 279811000001100 | Verapamil | Verapamil 80mg tablets (Teva UK Ltd) |
| 39009 | 454211000001101 | Verapamil | Verapamil 40mg tablets (Teva UK Ltd) |
| 40405 | 590011000001102 | Verapamil | Verapamil 120mg tablets (Teva UK Ltd) |
| 41586 | 7911000001103 | Verapamil | Verapamil 80mg tablets (Actavis UK Ltd) |
| 41679 | 534011000001101 | Verapamil | Verapamil 80mg tablets (IVAX Pharmaceuticals UK Ltd) |
| 41693 | 485611000001106 | Verapamil | Verapamil 120mg tablets (Mylan) |
| 42625 | 16449211000001100 | Verapamil | Vera-Til SR 120mg tablets (Actavis UK Ltd) |
| 43879 | 16438111000001103 | Verapamil | Vera-Til SR 240mg tablets (Actavis UK Ltd) |
| 45051 | 66545001000027106 | Verapamil | Verapamil hc 240mg Modified-release tablet (Actavis UK Ltd) |
| 45308 | 9208311000001103 | Verapamil | Verapamil 240mg modified-release tablets (Mylan) |
| 46009 | 887611000001108 | Verapamil | Verapamil 120mg tablets (Kent Pharmaceuticals Ltd) |
| 46884 | 136715001000027100 | Verapamil | Verapamil hc 240mg Modified-release tablet (Sandoz Ltd) |
| 46955 | 803411000001104 | Verapamil | Verapamil 80mg tablets (Mylan) |
| 47222 | 13924111000001102 | Verapamil | Verapamil 120mg modified-release tablets (A A H Pharmaceuticals Ltd) |
| 47230 | 9527111000001101 | Verapamil | Verapamil 240mg modified-release tablets (Teva UK Ltd) |
| 51461 | 5407511000001101 | Verapamil | Securon SR 240mg tablets (Waymade Healthcare Plc) |
| 59264 | 18611011000001108 | Verapamil | Securon SR 240mg tablets (DE Pharmaceuticals) |
| 62552 | 223411000001100 | Verapamil | Verapamil 80mg tablets (Alliance Healthcare (Distribution) Ltd) |
| 67293 | 18633611000001108 | Verapamil | Half Securon SR 120mg tablets (Mawdsley-Brooks & Company Ltd) |
| 68531 | 13015411000001109 | Verapamil | Verapamil 40mg/ 5ml oral suspension |
| 71009 | 81311000001105 | Verapamil | Verapamil 40mg tablets (Kent Pharmaceuticals Ltd) |
| 73467 | 87665001000027108 | Verapamil | Verapamil hc 80mg Tablet (Ranbaxy (UK) Ltd) |
| 76547 | | Verapamil | Verapamil 80mg tablets (Waymade Healthcare Plc) |
| 76554 | | Verapamil | Verapamil 120mg modified-release tablets (DE Pharmaceuticals) |
| 77506 | | Verapamil | Verapamil 240mg modified-release tablets (Kent Pharmaceuticals Ltd) |
| 77557 | | Verapamil | Securon SR 240mg tablets (Mawdsley-Brooks & Company Ltd) |
| 29116 | 237645001000027104 | Perhexiline Maleate | Perhexiline maleate 100mg tablet |
| 61230 | 504305001000027109 | Perhexiline Maleate | Perhexiline maleate 100mg tablets |
| 75641 | | Perhexiline Maleate | Pexsig 100mg tablets (Imported (New Zealand)) |
| 18038 | 36031911000001102 | Nisoldipine | Nisoldipine 20mg modified-release tablets |
| 19129 | 3877011000001100 | Nisoldipine | Syscor MR 10 tablets (Forest Laboratories UK Ltd) |

| | | | |
|---|---|---|---|
| 23805 | 36031811000001107 | Nisoldipine | Nisoldipine 10mg modified-release tablets |
| 23823 | 36032011000001109 | Nisoldipine | Nisoldipine 30mg modified-release tablets |
| 31336 | 4069211000001108 | Nisoldipine | Syscor MR 30 tablets (Forest Laboratories UK Ltd) |
| 31337 | 4070011000001100 | Nisoldipine | Syscor MR 20 tablets (Forest Laboratories UK Ltd) |
| 10595 | 3879211000001104 | Nimodipine | Nimotop 30mg tablets (Bayer Plc) |
| 11547 | 323273000 | Nimodipine | Nimodipine 30mg tablets |
| 269 | 319222004 | Nifedipine | Nifedipine 5mg capsules |
| 410 | 319274001 | Nifedipine | Nifedipine 10mg modified-release tablets |
| 452 | 319223009 | Nifedipine | Nifedipine 10mg capsules |
| 541 | 881811000001100 | Nifedipine | Adalat LA 20mg tablets (Bayer Plc) |
| 662 | 27111000001107 | Nifedipine | Adalat 5mg capsules (Bayer Plc) |
| 737 | 319273007 | Nifedipine | Nifedipine 20mg modified-release capsules |
| 1262 | 129175001000027109 | Nifedipine | Nifedipine 12 20mg Modified-release tablet |
| 1300 | 55365001000027100 | Nifedipine | Nifensar xl 20mg Modified-release tablet (Rhone-Poulenc Rorer Ltd) |
| 1449 | 214905001000027101 | Nifedipine | Nifedipine 24 30mg Modified-release tablet |
| 1854 | 125735001000027108 | Nifedipine | Adalat la 30mg Tablet (Bayer Plc) |
| 2280 | 569011000001108 | Nifedipine | Adalat retard 10mg tablets (Bayer Plc) |
| 2343 | 5011000001109 | Nifedipine | Adalat retard 20mg tablets (Bayer Plc) |
| 2521 | 782511000001108 | Nifedipine | Adalat 10mg capsules (Bayer Plc) |
| 2605 | 319272002 | Nifedipine | Nifedipine 10mg modified-release capsules |
| 2746 | 126411000001108 | Nifedipine | Coracten SR 10mg capsules (UCB Pharma Ltd) |
| 3711 | 74111000001103 | Nifedipine | Adipine MR 20 tablets (Chiesi Ltd) |
| 3712 | 162811000001100 | Nifedipine | Coracten XL 30mg capsules (UCB Pharma Ltd) |
| 3930 | 319276004 | Nifedipine | Nifedipine 60mg modified-release tablets |
| 4227 | 125745001000027109 | Nifedipine | Adalat la 60mg Tablet (Bayer Plc) |
| 4239 | 833611000001109 | Nifedipine | Adipine MR 10 tablets (Chiesi Ltd) |
| 4856 | 389611000001101 | Nifedipine | Coracten SR 20mg capsules (UCB Pharma Ltd) |
| 4939 | 3381511000001105 | Nifedipine | Coracten XL 60mg capsules (UCB Pharma Ltd) |
| 5162 | 319247002 | Nifedipine | Nifedipine 30mg modified-release capsules |
| 5181 | 865011000001105 | Nifedipine | Angiopine MR 20mg tablets (Ashbourne Pharmaceuticals Ltd) |
| 5277 | 188711000001108 | Nifedipine | Fortipine LA 40 tablets (AMCo) |
| 5806 | 385611000001104 | Nifedipine | Tensipine MR 20 tablets (Thornton & Ross Ltd) |
| 7541 | 280811000001100 | Nifedipine | Nifopress Retard 20mg tablets (AMCo) |
| 8213 | 197535001000027103 | Nifedipine | Nifedipine 24 20mg Modified-release tablet |
| 9269 | 319233001 | Nifedipine | Nifedipine 40mg modified-release tablets |
| 9485 | 677411000001108 | Nifedipine | Hypolar Retard 20 tablets (Sandoz Ltd) |
| 9553 | 630411000001107 | Nifedipine | Slofedipine XL 60 tablets (Zentiva) |
| 9573 | 2882011000001104 | Nifedipine | Slofedipine XL 30mg tablets (Zentiva) |
| 9750 | 319248007 | Nifedipine | Nifedipine 60mg modified-release capsules |
| 10135 | 235325001000027108 | Nifedipine | Nifedipress mr 10mg Modified-release tablet (Sandoz Ltd) |
| 10136 | 619111000001101 | Nifedipine | Nifedipress MR 20 tablets (Dexcel-Pharma Ltd) |
| 10246 | 9049911000001105 | Nifedipine | Adipine XL 60mg tablets (Chiesi Ltd) |
| 11512 | 904011000001104 | Nifedipine | Nifedipress MR 10 tablets (Dexcel-Pharma Ltd) |

| | | | |
|---|---|---|---|
| 11769 | 17011000001100 | Nifedipine | Calchan MR 20 tablets (Ranbaxy (UK) Ltd) |
| 12606 | 19988501000027103 | Nifedipine | Nifelease 20mg Modified-release tablet (Eastern Pharmaceuticals Ltd) |
| 12613 | 21591501000027109 | Nifedipine | Unipine xl 30mg Modified-release tablet (Genus Pharmaceuticals Ltd) |
| 13139 | 9049711000001108 | Nifedipine | Adipine XL 30mg tablets (Chiesi Ltd) |
| 13672 | 568911000001104 | Nifedipine | Angiopine MR 10mg tablets (Ashbourne Pharmaceuticals Ltd) |
| 13699 | 22257501000027106 | Nifedipine | Angiopine la 40mg Tablet (Ashbourne Pharmaceuticals Ltd) |
| 14861 | 627111000001104 | Nifedipine | Calchan MR 10 tablets (Ranbaxy (UK) Ltd) |
| 15715 | 23087501000027104 | Nifedipine | Genalat retard 20mg Modified-release tablet (Wyeth Pharmaceuticals) |
| 16073 | 7856211000001104 | Nifedipine | Nifedipress MR 10 tablets (Teva UK Ltd) |
| 17325 | 25911000001100 | Nifedipine | Cardilate MR 10mg tablets (Teva UK Ltd) |
| 17338 | 22717501000027101 | Nifedipine | Nifedotard 20 mr 20mg Modified-release tablet (Galen Ltd) |
| 17342 | 23401501000027109 | Nifedipine | Nivaten retard 10mg Modified-release tablet (Actavis UK Ltd) |
| 17448 | 22603501000027105 | Nifedipine | Nifedipress mr 10mg Modified-release tablet (Sterwin Medicines) |
| 19170 | 413111000001101 | Nifedipine | Tensipine MR 10 tablets (Thornton & Ross Ltd) |
| 20257 | 905711000001103 | Nifedipine | Cardilate MR 20mg tablets (IVAX Pharmaceuticals UK Ltd) |
| 20311 | 7777501000027106 | Nifedipine | Nifedipress mr 20mg Modified-release tablet (Generics (UK) Ltd) |
| 20591 | 7856611000001102 | Nifedipine | Nifedipress MR 20 tablets (Teva UK Ltd) |
| 20878 | 811311000001107 | Nifedipine | Angiopine 10 capsules (Ashbourne Pharmaceuticals Ltd) |
| 21216 | 4878111000001101 | Nifedipine | Hypolar Retard 10mg tablets (Sandoz Ltd) |
| 21245 | 23203501000027107 | Nifedipine | Nifedipress mr 10mg Modified-release tablet (Actavis UK Ltd) |
| 21872 | 16963501000027100 | Nifedipine | Angiopine 5mg Capsule (Ashbourne Pharmaceuticals Ltd) |
| 21886 | 7855311000001102 | Nifedipine | Nifedipress MR 20 tablets (Actavis UK Ltd) |
| 22019 | 21026501000027101 | Nifedipine | Calanif 10mg Capsule (Berk Pharmaceuticals Ltd) |
| 22142 | 17025501000027101 | Nifedipine | Calcilat 10mg Capsule (Eastern Pharmaceuticals Ltd) |
| 22217 | 22231501000027101 | Nifedipine | Nimodrel 10mg modified-release tablet (Opus Pharmaceuticals Ltd) |
| 22696 | 843411000001105 | Nifedipine | Slofedipine 20mg tablets (Sterwin Medicines) |
| 23736 | 2881811000001101 | Nifedipine | Hypolar XL 30 tablets (Sandoz Ltd) |
| 24228 | 22232501000027105 | Nifedipine | Nimodrel 20mg modified-release tablet (Opus Pharmaceuticals Ltd) |
| 25132 | 9485111000001105 | Nifedipine | Nifopress MR 20mg tablets (Teva UK Ltd) |
| 25646 | 23402501000027100 | Nifedipine | Nivaten retard 20mg Modified-release tablet (Actavis UK Ltd) |
| 25919 | 10054311000001109 | Nifedipine | Nifedipine 20mg modified-release tablets (A A H Pharmaceuticals Ltd) |
| 26265 | 21027501000027108 | Nifedipine | Calanif 5mg Capsule (Berk Pharmaceuticals Ltd) |
| 26774 | 24297501000027102 | Nifedipine | Nifedipine 10mg/ 5ml Oral suspension |
| 28688 | 10054111000001107 | Nifedipine | Nifedipine 10mg modified-release tablets (A A H Pharmaceuticals Ltd) |
| 30199 | 319275000 | Nifedipine | Nifedipine 30mg modified-release tablets |
| 30473 | 309611000001102 | Nifedipine | Coroday MR 20mg tablets (Mylan) |
| 33025 | 10189111000001106 | Nifedipine | Nimodrel XL 30mg tablets (Zurich Pharmaceuticals) |
| 34101 | 10308501000027109 | Nifedipine | Nifedipine mr 20mg Modified-release tablet (IVAX Pharmaceuticals UK Ltd) |
| 34115 | 15502501000027103 | Nifedipine | Nifedipine 60mg Modified-release tablet |
| 34146 | 10307501000027104 | Nifedipine | Nifedipine mr 10mg Modified-release tablet (IVAX Pharmaceuticals UK Ltd) |
| 34187 | 9363501000027105 | Nifedipine | Nifedipine 10mg Modified-release tablet (Generics (UK) Ltd) |
| 34247 | 8795001000027103 | Nifedipine | Nifedipine 10mg Capsule (Berk Pharmaceuticals Ltd) |
| 34522 | 720511000001103 | Nifedipine | Nifedipine 5mg capsules (A A H Pharmaceuticals Ltd) |

| | | | |
|---|---|---|---|
| 34607 | 814611000001100 | Nifedipine | Nifedipine 5mg capsules (IVAX Pharmaceuticals UK Ltd) |
| 34975 | 168411000001100 | Nifedipine | Nifedipine 5mg capsules (Teva UK Ltd) |
| 35646 | 10751411000001101 | Nifedipine | Neozipine XL 60mg tablets (Kent Pharmaceuticals Ltd) |
| 37025 | 319277008 | Nifedipine | Nifedipine 20mg modified-release tablets |
| 37184 | 13401911000001109 | Nifedipine | Valni XL 30mg tablets (Zentiva) |
| 37530 | 10751211000001100 | Nifedipine | Neozipine XL 30mg tablets (Kent Pharmaceuticals Ltd) |
| 37726 | 8670011000001105 | Nifedipine | Nifedipine 100mg/ 5ml oral suspension |
| 38107 | 185705001000027105 | Nifedipine | Nifedipine sr 30mg Tablet (Hillcross Pharmaceuticals Ltd) |
| 39800 | 13402111000001101 | Nifedipine | Valni XL 60mg tablets (Zentiva) |
| 40074 | 129135001000027107 | Nifedipine | Nifedipine 20mg Capsule |
| 41979 | 232235001000027104 | Nifedipine | Adipine la 30mg Modified-release tablet (Chiesi Ltd) |
| 42912 | 295511000001109 | Nifedipine | Nifedipine 10mg capsules (Teva UK Ltd) |
| 43222 | 693311000001101 | Nifedipine | Valni 20 Retard tablets (Tillomed Laboratories Ltd) |
| 43410 | 197555001000027104 | Nifedipine | Nifedipine extra 60mg Modified-release tablet |
| 43511 | 498311000001106 | Nifedipine | Nifedipine 10mg capsules (A A H Pharmaceuticals Ltd) |
| 43515 | 639811000001100 | Nifedipine | Nifedipine 10mg capsules (Actavis UK Ltd) |
| 43753 | 2881311000001105 | Nifedipine | Adalat LA 30mg tablets (Bayer Plc) |
| 43818 | 235511000001104 | Nifedipine | Adalat LA 60mg tablets (Bayer Plc) |
| 45685 | 17666011000001106 | Nifedipine | Adanif XL 30mg tablets (Focus Pharmaceuticals Ltd) |
| 46445 | 652811000001109 | Nifedipine | Nifedipine 10mg capsules (IVAX Pharmaceuticals UK Ltd) |
| 46887 | 17666211000001101 | Nifedipine | Adanif XL 60mg tablets (Focus Pharmaceuticals Ltd) |
| 47027 | 116995001000027105 | Nifedipine | Nifedipine 10mg Modified-release tablet (Kent Pharmaceuticals Ltd) |
| 47217 | 232245001000027103 | Nifedipine | Adipine la 60mg Modified-release tablet (Chiesi Ltd) |
| 47285 | 185845001000027103 | Nifedipine | Nifedipine xl 60mg Tablet (Hillcross Pharmaceuticals Ltd) |
| 47529 | 9096811000001100 | Nifedipine | Nifedipine 20mg/ ml oral drops |
| 47614 | 10639711000001103 | Nifedipine | Nifedipine 30mg modified-release tablets (A A H Pharmaceuticals Ltd) |
| 47707 | 247555001000027108 | Nifedipine | Nifedipine Oral solution |
| 47887 | 10189311000001108 | Nifedipine | Nimodrel XL 60mg tablets (Zurich Pharmaceuticals) |
| 49338 | 10056111000001100 | Nifedipine | Nifedipine 20mg modified-release tablets (Alliance Healthcare (Distribution) Ltd) |
| 49762 | 10055911000001109 | Nifedipine | Nifedipine 10mg modified-release tablets (Alliance Healthcare (Distribution) Ltd) |
| 51917 | 14043411000001104 | Nifedipine | Adalat LA 60 tablets (Sigma Pharmaceuticals Plc) |
| 52017 | 17489111000001103 | Nifedipine | Adalat LA 30 tablets (Mawdsley-Brooks & Company Ltd) |
| 53278 | 17574611000001108 | Nifedipine | Adalat LA 30 tablets (Necessity Supplies Ltd) |
| 53357 | 8670111000001106 | Nifedipine | Nifedipine 10mg/ 5ml oral suspension |
| 53500 | 13824711000001105 | Nifedipine | Adalat LA 30 tablets (DE Pharmaceuticals) |
| 53629 | 16135011000001106 | Nifedipine | Adalat retard 20mg tablets (Lexon (UK) Ltd) |
| 53990 | 8670311000001108 | Nifedipine | Nifedipine 5mg/ 5ml oral suspension |
| 55455 | 18283911000001107 | Nifedipine | Nifedipine 10mg capsules (Strides Shasun (UK) Ltd) |
| 55824 | 88575001000027100 | Nifedipine | Nifedipine 20mg Modified-release tablet (Berk Pharmaceuticals Ltd) |
| 56469 | 17575011000001102 | Nifedipine | Adalat LA 60 tablets (Necessity Supplies Ltd) |
| 57531 | 5335911000001104 | Nifedipine | Adalat LA 60 tablets (Waymade Healthcare Plc) |
| 57653 | 14043011000001108 | Nifedipine | Adalat LA 20 tablets (Sigma Pharmaceuticals Plc) |
| 58557 | 17574211000001106 | Nifedipine | Adalat LA 20 tablets (Necessity Supplies Ltd) |

| | | | |
|---|---|---|---|
| 58990 | 21868911000001101 | Nifedipine | Nifedipine 10mg modified-release tablets (Cubic Pharmaceuticals Ltd) |
| 59163 | 21868511000001108 | Nifedipine | Nifedipine 20mg modified-release tablets (Cubic Pharmaceuticals Ltd) |
| 60856 | 24102511000001108 | Nifedipine | Nifedipine 10mg modified-release tablets (Sigma Pharmaceuticals Plc) |
| 63041 | 868811000001108 | Nifedipine | Nifedipine 10mg capsules (Mylan) |
| 63246 | 28407711000001101 | Nifedipine | Nifedipine 10mg modified-release tablets (AM Distributions (Yorkshire) Ltd) |
| 66191 | 13825311000001105 | Nifedipine | Adalat retard 20mg tablets (DE Pharmaceuticals) |
| 66236 | 117035001000027106 | Nifedipine | Nifedipine 20mg Modified-release tablet (Kent Pharmaceuticals Ltd) |
| 67074 | 13824311000001106 | Nifedipine | Adalat 10mg capsules (DE Pharmaceuticals) |
| 69202 | 12862411000001109 | Nifedipine | Nifedipine 2mg/ 5ml oral suspension |
| 69668 | 12862511000001108 | Nifedipine | Nifedipine 30mg/ 5ml oral suspension |
| 71339 | 5439111000001108 | Nifedipine | Adalat LA 30 tablets (Dowelhurst Ltd) |
| 71601 | 12303311000001109 | Nifedipine | Nifedipine 2.5mg/ 5ml oral suspension |
| 71653 | 34685811000001103 | Nifedipine | Nidef 60mg modified-release tablets (Morningside Healthcare Ltd) |
| 71760 | 24615511000001100 | Nifedipine | Nifedipine 10mg modified-release tablets (Ethigen Ltd) |
| 72253 | 24102911000001101 | Nifedipine | Nifedipine 30mg modified-release tablets (Sigma Pharmaceuticals Plc) |
| 72321 | 34685211000001104 | Nifedipine | Nidef 30mg modified-release tablets (Morningside Healthcare Ltd) |
| 73992 | 5440311000001102 | Nifedipine | Adalat LA 60 tablets (Dowelhurst Ltd) |
| 74001 | 10459411000001108 | Nifedipine | Adalat LA 20 tablets (Dowelhurst Ltd) |
| 74061 | 5335411000001107 | Nifedipine | Adalat LA 30 tablets (Waymade Healthcare Plc) |
| 74330 | 26854411000001105 | Nifedipine | Nifedipine 20mg modified-release tablets (Ennogen Healthcare Ltd) |
| 74648 | 8835001000027107 | Nifedipine | Nifedipine 10mg Capsule (C P Pharmaceuticals Ltd) |
| 75109 | 88585001000027105 | Nifedipine | Nifedipine 10mg Modified-release tablet (Berk Pharmaceuticals Ltd) |
| 76488 | | Nifedipine | Nifedipine 5mg capsules (Strides Pharma UK Ltd) |
| 76539 | | Nifedipine | Nifedipin-ratiopharm 20mg/ ml oral drops (Imported (Germany)) |
| 77134 | | Nifedipine | Adalat 10mg capsules (Lexon (UK) Ltd) |
| 77418 | | Nifedipine | Adalat retard 20mg tablets (Waymade Healthcare Plc) |
| 77447 | | Nifedipine | Adalat 10mg capsules (Necessity Supplies Ltd) |
| 77555 | | Nifedipine | Adalat LA 30 tablets (Sigma Pharmaceuticals Plc) |
| 77962 | | Nifedipine | Adalat LA 20 tablets (Lexon (UK) Ltd) |
| 2926 | 319217004 | Nicardipine | Nicardipine 20mg capsules |
| 3302 | 540311000001105 | Nicardipine | Cardene SR 30mg capsules (Astellas Pharma Ltd) |
| 5477 | 319216008 | Nicardipine | Nicardipine 30mg modified-release capsules |
| 7562 | 291111000001102 | Nicardipine | Cardene 30mg capsules (Astellas Pharma Ltd) |
| 8201 | 319218009 | Nicardipine | Nicardipine 30mg capsules |
| 9386 | 319215007 | Nicardipine | Nicardipine 45mg modified-release capsules |
| 11943 | 344811000001108 | Nicardipine | Cardene 20mg capsules (Astellas Pharma Ltd) |
| 12875 | 118811000001102 | Nicardipine | Cardene SR 45mg capsules (Astellas Pharma Ltd) |
| 45292 | 743711000001101 | Nicardipine | Nicardipine 30mg capsules (A A H Pharmaceuticals Ltd) |
| 73646 | 13760311000001107 | Nicardipine | Nicardipine 20mg capsules (Tillomed Laboratories Ltd) |
| 73968 | 3656311000001105 | Nicardipine | Nicardipine 20mg capsules (Teva UK Ltd) |
| 1529 | 226015001000027104 | Mibefradil | Posicor 50mg Tablet (Roche Products Ltd) |
| 3931 | 226025001000027108 | Mibefradil | Posicor 100mg Tablet (Roche Products Ltd) |
| 15652 | 226705001000027103 | Mibefradil | Mibefradil 50mg Tablet |

| | | | |
|---|---|---|---|
| 22241 | 226715001000027101 | Mibefradil | Mibefradil 100mg Tablet |
| 19013 | 155925001000027108 | Lidoflazine | Clinium 120mg Tablet (LEO Pharma) |
| 30758 | 155905001000027101 | Lidoflazine | Lidoflazine 120mg Tablet |
| 5570 | 20011000001105 | Lercanidipine | Zanidip 10mg tablets (Recordati Pharmaceuticals Ltd) |
| 5593 | 319316005 | Lercanidipine | Lercanidipine 10mg tablets |
| 13243 | 10225911000001102 | Lercanidipine | Lercanidipine 20mg tablets |
| 14300 | 10198711000001102 | Lercanidipine | Zanidip 20mg tablets (Recordati Pharmaceuticals Ltd) |
| 47331 | 16666611000001103 | Lercanidipine | Lercanidipine 10mg tablets (Mylan) |
| 56767 | 16666411000001101 | Lercanidipine | Lercanidipine 20mg tablets (Mylan) |
| 57444 | 22337311000001108 | Lercanidipine | Lercanidipine 10mg tablets (Aptil Pharma Ltd) |
| 59233 | 16607211000001104 | Lercanidipine | Lercanidipine 20mg tablets (Actavis UK Ltd) |
| 61611 | 24363711000001103 | Lercanidipine | Lercanidipine 10mg tablets (DE Pharmaceuticals) |
| 63917 | 16732211000001104 | Lercanidipine | Lercanidipine 20mg tablets (A A H Pharmaceuticals Ltd) |
| 64227 | 16606811000001100 | Lercanidipine | Lercanidipine 10mg tablets (Actavis UK Ltd) |
| 64424 | 16640011000001100 | Lercanidipine | Lercanidipine 20mg tablets (Zentiva) |
| 65659 | 16726111000001106 | Lercanidipine | Lercanidipine 20mg tablets (Teva UK Ltd) |
| 69239 | 16732011000001109 | Lercanidipine | Lercanidipine 10mg tablets (A A H Pharmaceuticals Ltd) |
| 70732 | 15608611000001100 | Lercanidipine | Lercanidipine 20mg tablets (Alliance Healthcare (Distribution) Ltd) |
| 70827 | 16725911000001102 | Lercanidipine | Lercanidipine 10mg tablets (Teva UK Ltd) |
| 71018 | 20911211000001100 | Lercanidipine | Lercanidipine 10mg tablets (Arrow Generics Ltd) |
| 71030 | 20911411000001101 | Lercanidipine | Lercanidipine 20mg tablets (Arrow Generics Ltd) |
| 76672 | | Lercanidipine | Lercanidipine 20mg tablets (Sigma Pharmaceuticals Plc) |
| 77363 | | Lercanidipine | Zanidip 10mg tablets (Dowelhurst Ltd) |
| 3221 | 319301007 | Lacidipine | Lacidipine 4mg tablets |
| 5158 | 319300008 | Lacidipine | Lacidipine 2mg tablets |
| 9670 | 333011000001100 | Lacidipine | Motens 4mg tablets (GlaxoSmithKline UK Ltd) |
| 11966 | 910911000001108 | Lacidipine | Motens 2mg tablets (GlaxoSmithKline UK Ltd) |
| 56994 | 18565211000001108 | Lacidipine | Lacidipine 4mg tablets (Teva UK Ltd) |
| 57680 | 18642611000001102 | Lacidipine | Lacidipine 4mg tablets (A A H Pharmaceuticals Ltd) |
| 60699 | 14736811000001107 | Lacidipine | Lacidipine 2mg tablets (Sigma Pharmaceuticals Plc) |
| 70990 | 18642411000001100 | Lacidipine | Lacidipine 2mg tablets (A A H Pharmaceuticals Ltd) |
| 77450 | | Lacidipine | Motens 2mg tablets (Lexon (UK) Ltd) |
| 8257 | 3689711000001108 | Isradipine | Prescal 2.5mg tablets (Novartis Pharmaceuticals UK Ltd) |
| 8310 | 319280009 | Isradipine | Isradipine 2.5mg tablets |
| 491 | 319291002 | Felodipine | Felodipine 2.5mg modified-release tablets |
| 501 | 319287007 | Felodipine | Felodipine 5mg modified-release tablets |
| 568 | 319288002 | Felodipine | Felodipine 10mg modified-release tablets |
| 7280 | 48511000001101 | Felodipine | Plendil 10mg modified-release tablets (AstraZeneca UK Ltd) |
| 9334 | 562711000001104 | Felodipine | Plendil 2.5mg modified-release tablets (AstraZeneca UK Ltd) |
| 9437 | 490211000001101 | Felodipine | Plendil 5mg modified-release tablets (AstraZeneca UK Ltd) |
| 10153 | 235995001000027107 | Felodipine | Felendil xl 5mg Modified-release tablet (Ratiopharm UK Ltd) |
| 14305 | 5638811000001106 | Felodipine | Vascalpha 10mg modified-release tablets (Actavis UK Ltd) |
| 17557 | 4785111000001103 | Felodipine | Felotens XL 5mg tablets (Thornton & Ross Ltd) |

| | | | |
|---|---|---|---|
| 17566 | 4785511000001107 | Felodipine | Felotens XL 10mg tablets (Thornton & Ross Ltd) |
| 20459 | 236005001000027102 | Felodipine | Felendil xl 10mg Modified-release tablet (Ratiopharm UK Ltd) |
| 24365 | 7887011000001104 | Felodipine | Cardioplen XL 5mg tablets (Chiesi Ltd) |
| 24366 | 7887511000001107 | Felodipine | Cardioplen XL 10mg tablets (Chiesi Ltd) |
| 25572 | 4972811000001103 | Felodipine | Felogen XL 5mg tablets (Mylan) |
| 26337 | 3800711000001105 | Felodipine | Cabren 10mg modified-release tablets (Teva UK Ltd) |
| 28721 | 8090111000001106 | Felodipine | Neofel XL 5mg tablets (Kent Pharmaceuticals Ltd) |
| 29044 | 8089811000001107 | Felodipine | Neofel XL 10mg tablets (Kent Pharmaceuticals Ltd) |
| 29145 | 243075001000027100 | Felodipine | Felendil xl 2.5mg Modified-release tablet (Ratiopharm UK Ltd) |
| 30557 | 4973011000001100 | Felodipine | Felogen XL 10mg tablets (Mylan) |
| 30915 | 3800311000001106 | Felodipine | Cabren 2.5mg modified-release tablets (Teva UK Ltd) |
| 30991 | 3800511000001100 | Felodipine | Cabren 5mg modified-release tablets (Teva UK Ltd) |
| 32922 | 152805001000027108 | Felodipine | Felodipine 10mg Modified-release tablet (Sandoz Ltd) |
| 33091 | 16183811000001106 | Felodipine | Felodipine 10mg modified-release tablets (A A H Pharmaceuticals Ltd) |
| 33932 | 7387911000001103 | Felodipine | Parmid XL 5mg tablets (Sandoz Ltd) |
| 35084 | 5638311000001102 | Felodipine | Vascalpha 5mg modified-release tablets (Actavis UK Ltd) |
| 35592 | 11506711000001103 | Felodipine | Cardioplen XL 2.5mg tablets (Chiesi Ltd) |
| 36620 | 7388311000001103 | Felodipine | Parmid XL 10mg tablets (Sandoz Ltd) |
| 37897 | 13127311000001107 | Felodipine | Felotens XL 2.5mg tablets (Thornton & Ross Ltd) |
| 38434 | 10284611000001100 | Felodipine | Keloc SR 10mg tablets (Teva UK Ltd) |
| 39357 | 11493211000001105 | Felodipine | Neofel XL 2.5mg tablets (Kent Pharmaceuticals Ltd) |
| 40633 | 18250911000001108 | Felodipine | Vascalpha 5mg modified-release tablets (Almus Pharmaceuticals Ltd) |
| 43394 | 9359611000001103 | Felodipine | Pinefeld XL 10mg tablets (Tillomed Laboratories Ltd) |
| 43512 | 16183611000001107 | Felodipine | Felodipine 5mg modified-release tablets (A A H Pharmaceuticals Ltd) |
| 43790 | 18252511000001100 | Felodipine | Vascalpha 10mg modified-release tablets (Almus Pharmaceuticals Ltd) |
| 44859 | 142895001000027103 | Felodipine | Felodipine sr 5mg Tablet (Approved Prescription Services Ltd) |
| 48009 | 152785001000027109 | Felodipine | Felodipine 5mg Modified-release tablet (Sandoz Ltd) |
| 55306 | 5008511000001107 | Felodipine | Folpik XL 5mg tablets (Teva UK Ltd) |
| 55740 | 18167311000001103 | Felodipine | Neofel XL 2.5mg tablets (Actavis UK Ltd) |
| 58339 | 18681211000001100 | Felodipine | Neofel XL 2.5mg tablets (Almus Pharmaceuticals Ltd) |
| 60569 | 23594111000001104 | Felodipine | Felodipine 2.5mg modified-release tablets (Waymade Healthcare Plc) |
| 60652 | 24221811000001105 | Felodipine | Parmid XL 2.5mg tablets (Sandoz Ltd) |
| 60884 | 23916011000001102 | Felodipine | Felodipine 2.5mg modified-release tablets (Phoenix Healthcare Distribution Ltd) |
| 63331 | 13565311000001101 | Felodipine | Folpik XL 2.5mg tablets (Teva UK Ltd) |
| 64474 | 16183411000001109 | Felodipine | Felodipine 2.5mg modified-release tablets (A A H Pharmaceuticals Ltd) |
| 64504 | 17621211000001102 | Felodipine | Plendil 5mg modified-release tablets (Necessity Supplies Ltd) |
| 64719 | 29860311000001105 | Felodipine | Felodipine 2.5mg modified-release tablets (Sigma Pharmaceuticals Plc) |
| 64760 | 23915811000001100 | Felodipine | Felodipine 10mg modified-release tablets (Phoenix Healthcare Distribution Ltd) |
| 64917 | 23594511000001108 | Felodipine | Felodipine 10mg modified-release tablets (Waymade Healthcare Plc) |
| 65349 | 5008911000001100 | Felodipine | Folpik XL 10mg tablets (Teva UK Ltd) |
| 66095 | 30127711000001105 | Felodipine | Felodipine 5mg modified-release tablets (Mawdsley-Brooks & Company Ltd) |
| 66910 | 30822411000001105 | Felodipine | Felodipine 2.5mg modified-release tablets (DE Pharmaceuticals) |
| 68181 | 29860511000001104 | Felodipine | Felodipine 5mg modified-release tablets (Sigma Pharmaceuticals Plc) |

| | | | |
|---|---|---|---|
| 68499 | 142935001000027102 | Felodipine | Felodipine sr 10mg Tablet (Approved Prescription Services Ltd) |
| 68828 | 23977111000001107 | Felodipine | Felodipine 5mg modified-release tablets (DE Pharmaceuticals) |
| 69206 | 18029311000001107 | Felodipine | Felodipine 2.5mg/ 5ml oral solution |
| 72181 | 18029411000001100 | Felodipine | Felodipine 5mg/ 5ml oral solution |
| 72221 | 23594311000001102 | Felodipine | Felodipine 5mg modified-release tablets (Waymade Healthcare Plc) |
| 73715 | 23915611000001104 | Felodipine | Felodipine 5mg modified-release tablets (Phoenix Healthcare Distribution Ltd) |
| 74004 | 5575011000001104 | Felodipine | Plendil 5mg modified-release tablets (Dowelhurst Ltd) |
| 74012 | 5401711000001102 | Felodipine | Plendil 2.5mg modified-release tablets (Waymade Healthcare Plc) |
| 75345 | 34447411000001102 | Felodipine | Felodipine 2.5mg/ 5ml oral suspension |
| 219 | 418945005 | Diltiazem | Diltiazem 120mg modified-release tablets |
| 517 | 71075001000027107 | Diltiazem | Adizem sr 120mg Modified-release capsule (Napp Pharmaceuticals Ltd) |
| 536 | 111175001000027101 | Diltiazem | Tildiem la 200mg Modified-release capsule (Sanofi) |
| 636 | 319185009 | Diltiazem | Diltiazem 60mg modified-release capsules |
| 793 | 196035001000027100 | Diltiazem | Adizem xl 240mg Capsule (Napp Pharmaceuticals Ltd) |
| 939 | 103611000001105 | Diltiazem | Tildiem Retard 90mg tablets (Sanofi) |
| 1130 | 407011000001105 | Diltiazem | Viazem XL 300mg capsules (Thornton & Ross Ltd) |
| 1289 | 383911000001109 | Diltiazem | Tildiem Retard 120mg tablets (Sanofi) |
| 1538 | 155445001000027109 | Diltiazem | Diltiazem 60mg tablets |
| 1686 | 319182007 | Diltiazem | Diltiazem 90mg modified-release capsules |
| 1836 | 319205001 | Diltiazem | Diltiazem 60mg modified-release tablets |
| 1995 | 71715001000027101 | Diltiazem | Diltiazem 12hr 120mg modified-release capsules |
| 2453 | 155455001000027107 | Diltiazem | Diltiazem 60mg modified-release capsules |
| 2528 | 130211000001109 | Diltiazem | Slozem 120mg capsules (Merck Serono Ltd) |
| 2592 | 32311000001105 | Diltiazem | Viazem XL 120mg capsules (Thornton & Ross Ltd) |
| 2663 | 181565001000027106 | Diltiazem | Diltiazem 240mg modified-release capsules |
| 2686 | 196445001000027108 | Diltiazem | Dilzem xl mr 240mg Modified-release capsule (Elan Pharma) |
| 2811 | 71085001000027102 | Diltiazem | Adizem sr 180mg Modified-release capsule (Napp Pharmaceuticals Ltd) |
| 2888 | 762011000001102 | Diltiazem | Tildiem 60mg modified-release tablets (Sanofi) |
| 3061 | 71725001000027105 | Diltiazem | Diltiazem 12hr 180mg modified-release capsules |
| 3118 | 181545001000027100 | Diltiazem | Adizem sr 90mg Modified-release capsule (Napp Pharmaceuticals Ltd) |
| 3370 | 196425001000027105 | Diltiazem | Dilzem xl mr 120mg Modified-release capsule (Elan Pharma) |
| 3676 | 196435001000027107 | Diltiazem | Dilzem xl mr 180mg Modified-release capsule (Elan Pharma) |
| 4308 | 193225001000027104 | Diltiazem | Dilzem sr 90mg Capsule (Elan Pharma) |
| 4408 | 599811000001104 | Diltiazem | Slozem 240mg capsules (Merck Serono Ltd) |
| 4635 | 319187001 | Diltiazem | Diltiazem 200mg modified-release capsules |
| 4732 | 319180004 | Diltiazem | Diltiazem 90mg modified-release tablets |
| 4808 | 319186005 | Diltiazem | Diltiazem 240mg modified-release capsules |
| 4852 | 181525001000027103 | Diltiazem | Adizem sr 120mg Modified-release tablet (Napp Pharmaceuticals Ltd) |
| 4923 | 197585001000027106 | Diltiazem | Diltiazem 24hr 180mg modified-release capsules |
| 5054 | 336611000001101 | Diltiazem | Angitil SR 180 capsules (Ethypharm UK Ltd) |
| 5194 | 193235001000027102 | Diltiazem | Dilzem sr 120mg Capsule (Elan Pharma) |
| 5234 | 119211000001108 | Diltiazem | Slozem 180mg capsules (Merck Serono Ltd) |
| 5296 | 111165001000027108 | Diltiazem | Tildiem la 300mg Modified-release capsule (Sanofi) |

| | | | |
|---|---|---|---|
| 5326 | 198095001000027102 | Diltiazem | Diltiazem 24hr 300mg modified-release capsules |
| 5348 | 319181000 | Diltiazem | Diltiazem 300mg modified-release capsules |
| 5513 | 193215001000027108 | Diltiazem | Dilzem sr 60mg Capsule (Elan Pharma) |
| 6309 | 71095001000027103 | Diltiazem | Adizem xl 300mg Capsule (Napp Pharmaceuticals Ltd) |
| 7398 | 648211000001107 | Diltiazem | Viazem XL 360mg capsules (Thornton & Ross Ltd) |
| 8558 | 196015001000027106 | Diltiazem | Adizem xl 120mg Capsule (Napp Pharmaceuticals Ltd) |
| 9240 | 196025001000027102 | Diltiazem | Adizem xl 180mg Capsule (Napp Pharmaceuticals Ltd) |
| 9374 | 181535001000027101 | Diltiazem | Adizem 60mg Modified-release tablet (Napp Pharmaceuticals Ltd) |
| 9410 | 298811000001103 | Diltiazem | Angitil SR 120 capsules (Ethypharm UK Ltd) |
| 9708 | 197575001000027101 | Diltiazem | Diltiazem 24hr 120mg modified-release capsules |
| 9723 | 219611000001107 | Diltiazem | Calcicard CR 90mg tablets (Teva UK Ltd) |
| 10267 | 2886511000001108 | Diltiazem | Adizem-XL 200mg capsules (Napp Pharmaceuticals Ltd) |
| 11223 | 857011000001109 | Diltiazem | Angitil SR 90 capsules (Ethypharm UK Ltd) |
| 11770 | 417111000001109 | Diltiazem | Dilzem SR 60 capsules (Teva UK Ltd) |
| 11922 | 8456911000001108 | Diltiazem | Diltiazem 60mg/ 5ml oral suspension |
| 11973 | 104111000001100 | Diltiazem | Calcicard CR 120mg tablets (Teva UK Ltd) |
| 12639 | 70315001000027107 | Diltiazem | Diltiazem 90mg Modified-release tablet (Actavis UK Ltd) |
| 12705 | 440711000001104 | Diltiazem | Angiozem CR 90mg tablets (Ashbourne Pharmaceuticals Ltd) |
| 13027 | 886511000001107 | Diltiazem | Viazem XL 240mg capsules (Thornton & Ross Ltd) |
| 13033 | 75111000001104 | Diltiazem | Angitil XL 240 capsules (Ethypharm UK Ltd) |
| 13075 | 254911000001106 | Diltiazem | Dilzem XL 180 capsules (Teva UK Ltd) |
| 13127 | 733511000001107 | Diltiazem | Dilzem XL 240 capsules (Teva UK Ltd) |
| 13240 | 243111000001108 | Diltiazem | Dilzem XL 120 capsules (Teva UK Ltd) |
| 13302 | 682311000001105 | Diltiazem | Dilzem SR 90 capsules (Teva UK Ltd) |
| 13410 | 672311000001103 | Diltiazem | Angiozem 60mg modified-release tablets (Ashbourne Pharmaceuticals Ltd) |
| 13926 | 319198000 | Diltiazem | Diltiazem 360mg modified-release capsules |
| 15221 | 199355001000027107 | Diltiazem | Dilcardia xl 180mg Modified-release capsule (Generics (UK) Ltd) |
| 15288 | 467111000001105 | Diltiazem | Angitil XL 300 capsules (Ethypharm UK Ltd) |
| 16038 | 5711000001106 | Diltiazem | Dilzem SR 120 capsules (Teva UK Ltd) |
| 16850 | 144811000001101 | Diltiazem | Angiozem CR 120mg tablets (Ashbourne Pharmaceuticals Ltd) |
| 17406 | 813611000001103 | Diltiazem | Zemtard 180 XL capsules (Galen Ltd) |
| 17425 | 105211000001107 | Diltiazem | Zemtard 120 XL capsules (Galen Ltd) |
| 17492 | 866811000001107 | Diltiazem | Zemtard 300 XL capsules (Galen Ltd) |
| 17586 | 550211000001104 | Diltiazem | Slozem 300mg capsules (Merck Serono Ltd) |
| 17666 | 33211000001108 | Diltiazem | Viazem XL 180mg capsules (Thornton & Ross Ltd) |
| 18379 | 353011000001101 | Diltiazem | Dilcardia SR 90mg capsules (Mylan) |
| 18403 | 95745001000027106 | Diltiazem | Diltiazem 180mg Modified-release capsule (Hillcross Pharmaceuticals Ltd) |
| 18404 | 4772111000001102 | Diltiazem | Diltiazem 60mg modified-release capsules (A A H Pharmaceuticals Ltd) |
| 18830 | 256611000001109 | Diltiazem | Disogram SR 90mg capsules (Ranbaxy (UK) Ltd) |
| 18834 | 469811000001106 | Diltiazem | Disogram SR 60mg capsules (Ranbaxy (UK) Ltd) |
| 18852 | 116711000001106 | Diltiazem | Disogram SR 120mg capsules (Ranbaxy (UK) Ltd) |
| 18874 | 713411000001103 | Diltiazem | Disogram SR 180mg capsules (Ranbaxy (UK) Ltd) |
| 18975 | 152715001000027107 | Diltiazem | Calcicard 60mg Tablet (3M Health Care Ltd) |

| | | | |
|---|---|---|---|
| 19426 | 108711000001107 | Diltiazem | Disogram SR 240mg capsules (Ranbaxy (UK) Ltd) |
| 19440 | 34711000001104 | Diltiazem | Disogram SR 300mg capsules (Ranbaxy (UK) Ltd) |
| 20642 | 228225001000027107 | Diltiazem | Bi-carzem sr 60mg Modified-release capsule (Tillomed Laboratories Ltd) |
| 20890 | 345411000001107 | Diltiazem | Zemtard 240 XL capsules (Galen Ltd) |
| 21145 | 937011000001101 | Diltiazem | Dilcardia SR 60mg capsules (Mylan) |
| 21763 | 215411000001106 | Diltiazem | Diltiazem 60mg modified-release tablets (A A H Pharmaceuticals Ltd) |
| 21773 | 96705001000027103 | Diltiazem | Diltiazem 60mg Tablet (Generics (UK) Ltd) |
| 21778 | 391911000001107 | Diltiazem | Diltiazem 60mg modified-release tablets (Teva UK Ltd) |
| 21795 | 865711000001107 | Diltiazem | Retalzem 60 modified-release tablets (Kent Pharmaceuticals Ltd) |
| 21918 | 527711000001108 | Diltiazem | Optil 60mg modified-release tablets (Opus Pharmaceuticals Ltd) |
| 22619 | 155475001000027105 | Diltiazem | Britiazim 60mg Modified-release tablet (Thames Laboratories Ltd) |
| 23233 | 228235001000027109 | Diltiazem | Bi-carzem sr 90mg Modified-release capsule (Tillomed Laboratories Ltd) |
| 23733 | 159965001000027107 | Diltiazem | Optil sr 90mg Modified-release capsule (Opus Pharmaceuticals Ltd) |
| 25777 | 723811000001107 | Diltiazem | Dilcardia SR 120mg capsules (Mylan) |
| 26267 | 159975001000027100 | Diltiazem | Optil sr 120mg Modified-release capsule (Opus Pharmaceuticals Ltd) |
| 26269 | 159985001000027105 | Diltiazem | Optil sr 180mg Modified-release capsule (Opus Pharmaceuticals Ltd) |
| 26270 | 203585001000027108 | Diltiazem | Optil xl 300mg Modified-release capsule (Opus Pharmaceuticals Ltd) |
| 26309 | 203575001000027103 | Diltiazem | Optil xl 240mg Modified-release capsule (Opus Pharmaceuticals Ltd) |
| 26460 | 199365001000027103 | Diltiazem | Dilcardia xl 240mg Modified-release capsule (Generics (UK) Ltd) |
| 26463 | 215655001000027103 | Diltiazem | Zemret xl 240mg Capsule (Neo Laboratories Ltd) |
| 26759 | 721811000001108 | Diltiazem | Zildil SR 60mg capsules (Chanelle Medical UK Ltd) |
| 27135 | 155305001000027109 | Diltiazem | Diltiazem sr 90mg Capsule (Hillcross Pharmaceuticals Ltd) |
| 27136 | 10065211000001107 | Diltiazem | Diltiazem 90mg modified-release tablets (A A H Pharmaceuticals Ltd) |
| 27401 | 8886211000001101 | Diltiazem | Kenzem SR 90mg capsules (Kent Pharmaceuticals Ltd) |
| 27685 | 129475001000027105 | Diltiazem | Diltiazem 300mg Capsule (PLIVA Pharma Ltd) |
| 28949 | 228245001000027105 | Diltiazem | Bi-carzem sr 120mg Modified-release capsule (Tillomed Laboratories Ltd) |
| 29676 | 214485001000027100 | Diltiazem | Calazem 60mg Modified-release tablet (Berk Pharmaceuticals Ltd) |
| 30197 | 319183002 | Diltiazem | Diltiazem 120mg modified-release capsules |
| 30242 | 319184008 | Diltiazem | Diltiazem 180mg modified-release capsules |
| 31489 | 233535001000027103 | Diltiazem | Bi-carzem xl 240mg Capsule (Tillomed Laboratories Ltd) |
| 31676 | 70325001000027103 | Diltiazem | Diltiazem 120mg Modified-release tablet (Actavis UK Ltd) |
| 31737 | 655411000001104 | Diltiazem | Zildil SR 120mg capsules (Chanelle Medical UK Ltd) |
| 32089 | 12155001000027107 | Diltiazem | Diltiazem 120mg Modified-release capsule (Hillcross Pharmaceuticals Ltd) |
| 32262 | 12115001000027102 | Diltiazem | Diltiazem 60mg Tablet (C P Pharmaceuticals Ltd) |
| 32658 | 199345001000027109 | Diltiazem | Dilcardia xl 120mg Modified-release capsule (Generics (UK) Ltd) |
| 32870 | 673611000001103 | Diltiazem | Diltiazem 60mg modified-release tablets (Sterwin Medicines) |
| 34377 | 95755001000027109 | Diltiazem | Diltiazem 90mg Modified-release capsule (Hillcross Pharmaceuticals Ltd) |
| 34475 | 45985001000027103 | Diltiazem | Diltiazem 90mg Modified-release tablet (IVAX Pharmaceuticals UK Ltd) |
| 34581 | 152545001000027104 | Diltiazem | Diltiazem 60mg Modified-release tablet (Kent Pharmaceuticals Ltd) |
| 34824 | 45995001000027104 | Diltiazem | Diltiazem 120mg Modified-release tablet (IVAX Pharmaceuticals UK Ltd) |
| 35696 | 8886511000001103 | Diltiazem | Kenzem SR 120mg capsules (Kent Pharmaceuticals Ltd) |
| 36583 | 215645001000027101 | Diltiazem | Zemret xl 180mg Capsule (Neo Laboratories Ltd) |
| 36664 | 215665001000027107 | Diltiazem | Zemret xl 300mg Capsule (Neo Laboratories Ltd) |

| | | | |
|---|---|---|---|
| 37774 | 8885711000001100 | Diltiazem | Kenzem SR 60mg capsules (Kent Pharmaceuticals Ltd) |
| 38066 | 89675001000027109 | Diltiazem | Diltiazem 60mg Modified-release tablet (Lagap) |
| 38545 | 261611000001107 | Diltiazem | Tildiem LA 200 capsules (Sanofi) |
| 38632 | 2887011000001102 | Diltiazem | Adizem-SR 90mg capsules (Napp Pharmaceuticals Ltd) |
| 38634 | 2886111000001104 | Diltiazem | Adizem-XL 300mg capsules (Napp Pharmaceuticals Ltd) |
| 38818 | 2887311000001104 | Diltiazem | Adizem-SR 120mg capsules (Napp Pharmaceuticals Ltd) |
| 38831 | 2886711000001103 | Diltiazem | Adizem-SR 180mg capsules (Napp Pharmaceuticals Ltd) |
| 38855 | 2938011000001101 | Diltiazem | Adizem-XL 180mg capsules (Napp Pharmaceuticals Ltd) |
| 38865 | 2937811000001108 | Diltiazem | Adizem-XL 120mg capsules (Napp Pharmaceuticals Ltd) |
| 38876 | 893111000001107 | Diltiazem | Tildiem LA 300 capsules (Sanofi) |
| 38882 | 2886311000001102 | Diltiazem | Adizem-XL 240mg capsules (Napp Pharmaceuticals Ltd) |
| 38964 | 2885611000001102 | Diltiazem | Adizem-SR 120mg tablets (Napp Pharmaceuticals Ltd) |
| 39171 | 540011000001107 | Diltiazem | Bi-Carzem SR 60mg capsules (Tillomed Laboratories Ltd) |
| 39298 | 764511000001100 | Diltiazem | Bi-Carzem SR 90mg capsules (Tillomed Laboratories Ltd) |
| 41489 | 580711000001101 | Diltiazem | Bi-Carzem SR 120mg capsules (Tillomed Laboratories Ltd) |
| 41635 | 544111000001104 | Diltiazem | Diltiazem 60mg modified-release tablets (IVAX Pharmaceuticals UK Ltd) |
| 42731 | 155325001000027102 | Diltiazem | Diltiazem sr 120mg Capsule (Hillcross Pharmaceuticals Ltd) |
| 42804 | 129425001000027106 | Diltiazem | Diltiazem 180mg Capsule (PLIVA Pharma Ltd) |
| 42819 | 163525001000027109 | Diltiazem | Diltiazem xl 240mg Capsule (Hillcross Pharmaceuticals Ltd) |
| 43430 | 10065011000001102 | Diltiazem | Diltiazem 120mg modified-release tablets (A A H Pharmaceuticals Ltd) |
| 44192 | 823511000001107 | Diltiazem | Zemret 240 XL capsules (Tillomed Laboratories Ltd) |
| 44887 | 233545001000027102 | Diltiazem | Bi-carzem xl 300mg Capsule (Tillomed Laboratories Ltd) |
| 45759 | 129455001000027107 | Diltiazem | Diltiazem 240mg Capsule (PLIVA Pharma Ltd) |
| 46937 | 896111000001102 | Diltiazem | Diltiazem 60mg modified-release tablets (Actavis UK Ltd) |
| 47415 | 203125001000027107 | Diltiazem | Diltiazem sr 60mg Capsule (Hillcross Pharmaceuticals Ltd) |
| 47530 | 591411000001102 | Diltiazem | Horizem SR 60mg capsules (Horizon lifecare) |
| 47608 | 640311000001101 | Diltiazem | Zemret 300 XL capsules (Tillomed Laboratories Ltd) |
| 47724 | 884811000001103 | Diltiazem | Bi-Carzem XL 240mg capsules (Tillomed Laboratories Ltd) |
| 47732 | 924711000001109 | Diltiazem | Zemret 180 XL capsules (Tillomed Laboratories Ltd) |
| 48272 | 10054911000001105 | Diltiazem | Diltiazem 60mg modified-release capsules (Alliance Healthcare (Distribution) Ltd) |
| 48282 | 18311411000001106 | Diltiazem | Diltiazem 90mg modified-release capsules (A A H Pharmaceuticals Ltd) |
| 48288 | 18311611000001109 | Diltiazem | Diltiazem 120mg modified-release capsules (A A H Pharmaceuticals Ltd) |
| 48457 | 10055111000001106 | Diltiazem | Diltiazem 90mg modified-release capsules (Alliance Healthcare (Distribution) Ltd) |
| 48870 | 19871811000001100 | Diltiazem | Adizem-SR 90mg capsules (DE Pharmaceuticals) |
| 49001 | 10055311000001108 | Diltiazem | Diltiazem 120mg modified-release tablets (Alliance Healthcare (Distribution) Ltd) |
| 49289 | 10054711000001108 | Diltiazem | Diltiazem 120mg modified-release capsules (Alliance Healthcare (Distribution) Ltd) |
| 49390 | 10055511000001102 | Diltiazem | Diltiazem 90mg modified-release tablets (Alliance Healthcare (Distribution) Ltd) |
| 51261 | 18481611000001101 | Diltiazem | Tildiem Retard 120mg tablets (Mawdsley-Brooks & Company Ltd) |
| 52276 | 19872011000001103 | Diltiazem | Adizem-XL 180mg capsules (DE Pharmaceuticals) |
| 52701 | 18481911000001107 | Diltiazem | Tildiem LA 200 capsules (Mawdsley-Brooks & Company Ltd) |
| 54799 | 17465311000001101 | Diltiazem | Tildiem LA 300 capsules (Mawdsley-Brooks & Company Ltd) |
| 55257 | 8457011000001107 | Diltiazem | Diltiazem 60mg/ 5ml oral solution |
| 56467 | 14005111000001104 | Diltiazem | Tildiem 60mg modified-release tablets (DE Pharmaceuticals) |

| | | | |
|---|---|---|---|
| 56758 | 21864611000001102 | Diltiazem | Diltiazem 90mg modified-release capsules (Cubic Pharmaceuticals Ltd) |
| 57208 | 21864811000001103 | Diltiazem | Diltiazem 120mg modified-release capsules (Cubic Pharmaceuticals Ltd) |
| 57594 | 5424211000001104 | Diltiazem | Tildiem 60mg modified-release tablets (Waymade Healthcare Plc) |
| 57859 | 21865011000001108 | Diltiazem | Diltiazem 90mg modified-release tablets (Cubic Pharmaceuticals Ltd) |
| 59098 | 16158611000001109 | Diltiazem | Dilzem XL 180 capsules (Lexon (UK) Ltd) |
| 59585 | 21965711000001100 | Diltiazem | Uard 120XL capsules (Ennogen Healthcare Ltd) |
| 59863 | 16158811000001108 | Diltiazem | Dilzem XL 240 capsules (Lexon (UK) Ltd) |
| 60415 | 17537211000001101 | Diltiazem | Dilzem XL 180 capsules (Sigma Pharmaceuticals Plc) |
| 60620 | 23929811000001107 | Diltiazem | Adizem-XL 240mg capsules (Waymade Healthcare Plc) |
| 61010 | 23586211000001104 | Diltiazem | Diltiazem 120mg modified-release tablets (Cubic Pharmaceuticals Ltd) |
| 61245 | 24597111000001107 | Diltiazem | Diltiazem 60mg modified-release capsules (Sigma Pharmaceuticals Plc) |
| 61532 | 24596711000001105 | Diltiazem | Diltiazem 120mg modified-release capsules (Sigma Pharmaceuticals Plc) |
| 62064 | 24523511000001104 | Diltiazem | Diltiazem 120mg modified-release tablets (Mawdsley-Brooks & Company Ltd) |
| 62065 | 24117911000001101 | Diltiazem | Diltiazem 90mg modified-release tablets (Colorama Pharmaceuticals Ltd) |
| 62207 | 23928711000001108 | Diltiazem | Adizem-SR 120mg capsules (Waymade Healthcare Plc) |
| 62912 | 28405011000001105 | Diltiazem | Diltiazem 120mg modified-release capsules (AM Distributions (Yorkshire) Ltd) |
| 65504 | 29955011000001104 | Diltiazem | Adizem-SR 180mg capsules (Lexon (UK) Ltd) |
| 65602 | 23929211000001106 | Diltiazem | Adizem-XL 120mg capsules (Waymade Healthcare Plc) |
| 65636 | 29955211000001109 | Diltiazem | Adizem-XL 120mg capsules (Lexon (UK) Ltd) |
| 66048 | 14005611000001107 | Diltiazem | Tildiem Retard 120mg tablets (DE Pharmaceuticals) |
| 66172 | 21965911000001103 | Diltiazem | Uard 180XL capsules (Ennogen Healthcare Ltd) |
| 66635 | 18574511000001102 | Diltiazem | Dilzem XL 180 capsules (DE Pharmaceuticals) |
| 66701 | 24101311000001106 | Diltiazem | Diltiazem 240mg modified-release capsules (DE Pharmaceuticals) |
| 66834 | 21966111000001107 | Diltiazem | Uard 240XL capsules (Ennogen Healthcare Ltd) |
| 66850 | 25711011000001108 | Diltiazem | Diltiazem 240mg modified-release capsules (Icarus Pharmaceuticals Ltd) |
| 67317 | 18189911000001107 | Diltiazem | Dilzem XL 120 capsules (Mawdsley-Brooks & Company Ltd) |
| 67344 | 24343511000001100 | Diltiazem | Diltiazem 300mg modified-release capsules (Ennogen Pharma Ltd) |
| 67890 | 21965511000001105 | Diltiazem | Uard 300XL capsules (Ennogen Healthcare Ltd) |
| 68054 | 526711000001105 | Diltiazem | Diltiazem 60mg modified-release tablets (Alliance Healthcare (Distribution) Ltd) |
| 68429 | 24615311000001106 | Diltiazem | Diltiazem 120mg modified-release capsules (Ethigen Ltd) |
| 69028 | 23969211000001102 | Diltiazem | Diltiazem 60mg modified-release capsules (DE Pharmaceuticals) |
| 69108 | 29956011000001108 | Diltiazem | Adizem-XL 300mg capsules (Lexon (UK) Ltd) |
| 69116 | 30139111000001101 | Diltiazem | Diltiazem 60mg modified-release tablets (DE Pharmaceuticals) |
| 69277 | 29955811000001105 | Diltiazem | Adizem-XL 240mg capsules (Lexon (UK) Ltd) |
| 70306 | 24524011000001109 | Diltiazem | Diltiazem 180mg modified-release capsules (Mawdsley-Brooks & Company Ltd) |
| 70961 | 14005411000001109 | Diltiazem | Tildiem Retard 90mg tablets (DE Pharmaceuticals) |
| 71342 | 10530911000001104 | Diltiazem | Dilzem XL 240 capsules (Waymade Healthcare Plc) |
| 71413 | 4773111000001108 | Diltiazem | Diltiazem 120mg modified-release capsules (A A H Pharmaceuticals Ltd) |
| 71702 | 8443011000001104 | Diltiazem | Diltiazem 60mg/ 5ml oral solution (Special Order) |
| 71969 | 24597911000001105 | Diltiazem | Diltiazem 300mg modified-release capsules (Sigma Pharmaceuticals Plc) |
| 72091 | 24102211000001105 | Diltiazem | Diltiazem 180mg modified-release capsules (DE Pharmaceuticals) |
| 72488 | 33424611000001105 | Diltiazem | Diltiazem 300mg modified-release capsules (A A H Pharmaceuticals Ltd) |
| 72839 | 24410111000001104 | Diltiazem | Diltiazem 300mg modified-release capsules (DE Pharmaceuticals) |

| | | | |
|---|---|---|---|
| 74022 | 5426711000001102 | Diltiazem | Tildiem LA 200 capsules (Waymade Healthcare Plc) |
| 74029 | 5619211000001106 | Diltiazem | Tildiem Retard 90mg tablets (Dowelhurst Ltd) |
| 74053 | 17537411000001102 | Diltiazem | Dilzem XL 240 capsules (Sigma Pharmaceuticals Plc) |
| 74689 | 24342811000001105 | Diltiazem | Diltiazem 240mg modified-release capsules (Ennogen Pharma Ltd) |
| 74915 | 22409111000001104 | Diltiazem | Diltiazem 120mg modified-release capsules (DE Pharmaceuticals) |
| 77077 | | Diltiazem | Diltiazem 10mg/ 5ml oral solution |
| 77442 | | Diltiazem | Tildiem 60mg modified-release tablets (Dowelhurst Ltd) |
| 77947 | | Diltiazem | Diltiazem 120mg/ 5ml oral solution |
| 29 | 182975001000027106 | Amlodipine | Amlodipine besilate 5mg tablets |
| 71 | 182985001000027101 | Amlodipine | Amlodipine besilate 10mg tablets |
| 729 | 237575001000027105 | Amlodipine | Amlodipine maleate 5mg tablets |
| 749 | 319283006 | Amlodipine | Amlodipine 5mg tablets |
| 3917 | 172711000001100 | Amlodipine | Istin 5mg tablets (Pfizer Ltd) |
| 5914 | 408111000001107 | Amlodipine | Istin 10mg tablets (Pfizer Ltd) |
| 6477 | 237585001000027100 | Amlodipine | Amlodipine maleate 10mg tablets |
| 6856 | 319284000 | Amlodipine | Amlodipine 10mg tablets |
| 16162 | 8278311000001107 | Amlodipine | Amlodipine 5mg/ 5ml oral suspension |
| 17640 | 8046211000001107 | Amlodipine | Amlostin 5mg tablets (Discovery Pharmaceuticals) |
| 31761 | 8046411000001106 | Amlodipine | Amlostin 10mg tablets (Discovery Pharmaceuticals) |
| 32595 | 7305311000001108 | Amlodipine | Amlodipine 5mg tablets (A A H Pharmaceuticals Ltd) |
| 32917 | 7305711000001107 | Amlodipine | Amlodipine 5mg tablets (IVAX Pharmaceuticals UK Ltd) |
| 34093 | 7305011000001105 | Amlodipine | Amlodipine 10mg tablets (A A H Pharmaceuticals Ltd) |
| 36202 | 8038411000001106 | Amlodipine | Amlodipine 10mg tablets (Actavis UK Ltd) |
| 39804 | 11008811000001105 | Amlodipine | Amlodipine 5mg tablets (Dr Reddy's Laboratories (UK) Ltd) |
| 39914 | 7333311000001100 | Amlodipine | Amlodipine 5mg tablets (Teva UK Ltd) |
| 42210 | 9557311000001100 | Amlodipine | Amlodipine 10mg tablets (Zentiva) |
| 43470 | 14768911000001108 | Amlodipine | Amlodipine 5mg tablets (Wockhardt UK Ltd) |
| 43880 | 11398511000001109 | Amlodipine | Amlodipine 5mg tablets (Almus Pharmaceuticals Ltd) |
| 45070 | 8278111000001105 | Amlodipine | Amlodipine 10mg/ 5ml oral suspension |
| 45279 | 7376311000001107 | Amlodipine | Amlodipine 5mg tablets (Sandoz Ltd) |
| 46233 | 243185001000027100 | Amlodipine | Amlodipine Oral solution |
| 46724 | 13892511000001100 | Amlodipine | Amlodipine 5mg/ 5ml oral solution |
| 47002 | 266105001000027102 | Amlodipine | Amlodipine 10mg/ 5ml sugar free Oral suspension |
| 49636 | 19704911000001101 | Amlodipine | Amlodipine 10mg tablets (DE Pharmaceuticals) |
| 52440 | 20478011000001105 | Amlodipine | Amlodipine 10mg/ 5ml oral solution |
| 53868 | 8038211000001107 | Amlodipine | Amlodipine 5mg tablets (Actavis UK Ltd) |
| 54515 | 7304811000001100 | Amlodipine | Amlodipine 10mg tablets (Alliance Healthcare (Distribution) Ltd) |
| 54633 | 15981411000001105 | Amlodipine | Amlodipine 5mg tablets (Bristol Laboratories Ltd) |
| 54654 | 11399011000001106 | Amlodipine | Amlodipine 10mg tablets (Almus Pharmaceuticals Ltd) |
| 54696 | 7376511000001101 | Amlodipine | Amlodipine 10mg tablets (Sandoz Ltd) |
| 54983 | 8278211000001104 | Amlodipine | Amlodipine 2.5mg/ 5ml oral suspension |
| 56147 | 18457811000001100 | Amlodipine | Amlodipine 10mg tablets (Accord Healthcare Ltd) |
| 56334 | 15981211000001106 | Amlodipine | Amlodipine 10mg tablets (Bristol Laboratories Ltd) |

| | | | |
|---|---|---|---|
| 58580 | 19833311000001108 | Amlodipine | Amlodipine 10mg tablets (APC Pharmaceuticals & Chemicals (Europe) Ltd) |
| 59001 | 7391311000001109 | Amlodipine | Amlodipine 10mg tablets (Mylan) |
| 59762 | 7333511000001106 | Amlodipine | Amlodipine 10mg tablets (Teva UK Ltd) |
| 60244 | 17779511000001109 | Amlodipine | Amlodipine 10mg tablets (Phoenix Healthcare Distribution Ltd) |
| 61374 | 11712011000001102 | Amlodipine | Amlodipine 4mg/ 5ml oral suspension |
| 61422 | 18458011000001107 | Amlodipine | Amlodipine 5mg tablets (Accord Healthcare Ltd) |
| 63515 | 11009211000001104 | Amlodipine | Amlodipine 10mg tablets (Dr Reddy's Laboratories (UK) Ltd) |
| 64166 | 29826311000001101 | Amlodipine | Amlodipine 5mg/ 5ml oral solution sugar free |
| 64327 | 7391211000001101 | Amlodipine | Amlodipine 5mg tablets (Mylan) |
| 64418 | 22080611000001107 | Amlodipine | Amlodipine 5mg tablets (Waymade Healthcare Plc) |
| 64441 | 7304611000001104 | Amlodipine | Amlodipine 5mg tablets (Alliance Healthcare (Distribution) Ltd) |
| 64447 | 19164711000001109 | Amlodipine | Amlodipine 5mg tablets (Somex Pharma) |
| 64606 | 10287111000001103 | Amlodipine | Amlodipine 5mg tablets (Focus Pharmaceuticals Ltd) |
| 64623 | 29826211000001109 | Amlodipine | Amlodipine 10mg/ 5ml oral solution sugar free |
| 65745 | 19705311000001103 | Amlodipine | Amlodipine 5mg tablets (DE Pharmaceuticals) |
| 66430 | 7378711000001105 | Amlodipine | Amlodipine 5mg tablets (Kent Pharmaceuticals Ltd) |
| 66574 | 17779111000001100 | Amlodipine | Amlodipine 5mg tablets (Phoenix Healthcare Distribution Ltd) |
| 66817 | 14769111000001103 | Amlodipine | Amlodipine 10mg tablets (Wockhardt UK Ltd) |
| 67662 | 13917011000001100 | Amlodipine | Istin 10mg tablets (DE Pharmaceuticals) |
| 68221 | 7378911000001107 | Amlodipine | Amlodipine 10mg tablets (Kent Pharmaceuticals Ltd) |
| 68311 | 29932511000001100 | Amlodipine | Amlodipine 5mg/ 5ml oral solution sugar free (A A H Pharmaceuticals Ltd) |
| 70999 | 19833111000001106 | Amlodipine | Amlodipine 5mg tablets (APC Pharmaceuticals & Chemicals (Europe) Ltd) |
| 71344 | 5332311000001106 | Amlodipine | Amlodipine 5mg tablets (Waymade Healthcare Plc) |
| 71353 | 5450111000001102 | Amlodipine | Amlodipine 5mg tablets (Dowelhurst Ltd) |
| 71939 | 10286711000001100 | Amlodipine | Amlodipine 10mg tablets (Focus Pharmaceuticals Ltd) |
| 72049 | 29992111000001103 | Amlodipine | Amlodipine 5mg tablets (Mawdsley-Brooks & Company Ltd) |
| 72199 | 35134611000001100 | Amlodipine | Amlodipine 5mg tablets (Aurobindo Pharma Ltd) |
| 72430 | 29781411000001107 | Amlodipine | Amlodipine 10mg/ 5ml oral solution sugar free (Alliance Healthcare (Distribution) Ltd) |
| 73612 | 30821811000001103 | Amlodipine | Amlodipine 5mg/ 5ml oral solution sugar free (DE Pharmaceuticals) |
| 74108 | 15064611000001101 | Amlodipine | Amlodipine 5mg tablets (Sigma Pharmaceuticals Plc) |
| 74121 | 30216911000001104 | Amlodipine | Amlodipine 5mg/ 5ml oral solution sugar free (Thame Laboratories Ltd) |
| 74795 | 19164911000001106 | Amlodipine | Amlodipine 10mg tablets (Somex Pharma) |
| 74820 | 15064211000001103 | Amlodipine | Amlodipine 10mg tablets (Sigma Pharmaceuticals Plc) |
| 74821 | 13764411000001106 | Amlodipine | Amlodipine 5mg tablets (Apotex UK Ltd) |
| 75387 | 10378211000001106 | Amlodipine | Amlodipine 5mg tablets (Arrow Generics Ltd) |
| 75609 | | Amlodipine | Amlodipine 5mg/ 5ml oral solution (Special Order) |
| 75862 | | Amlodipine | Amlodipine 5mg/ 5ml oral suspension sugar free |
| 76714 | | Amlodipine | Amlodipine 10mg/ 5ml oral solution sugar free (Thame Laboratories Ltd) |
| 77416 | | Amlodipine | Istin 10mg tablets (Dowelhurst Ltd) |
| 77420 | | Amlodipine | Istin 5mg tablets (DE Pharmaceuticals) |
| 77446 | | Amlodipine | Istin 10mg tablets (Waymade Healthcare Plc) |
| 7823 | 188995001000027108 | | NIFEDIPINE TAB 5 mg |
| 8024 | 96995001000027108 | | DILTIAZEM XL 300 MG CAP |

| | | | |
|---|---|---|---|
| 9094 | 118925001000027105 | | DILTIAZEM SR 300 MG CAP |
| 9211 | 95195001000027109 | | ADIZEM-XL 180 MG CAP |
| 10897 | 96235001000027102 | | VERAPAMIL S/ F 40 MG/ 5ML SOL |
| 15659 | 151485001000027101 | | DILTIAZEM S/ R 180 CAP |
| 18631 | | | VERAPAMIL MR |
| 18690 | 133445001000027107 | | SECURON (CALENDAR PACK) 120 MG TAB |
| 19015 | 171845001000027100 | | ADIZEM CONTINUS 120 MG TAB |
| 21496 | 166105001000027100 | | PERHEXILINE MALEATE 100 MG TAB |
| 21665 | | | CORDILOX |
| 23458 | | | VERAPAMIL SR |
| 23730 | 163275001000027103 | | VERAPAMIL 100 MG TAB |
| 25026 | | | NIFEDIPINE RETARD |
| 25027 | | | ADALAT RETARD 10 |
| 25044 | | | NIFEDIPINE RETARD |
| 25055 | | | NIFEDIPINE |
| 27910 | | | ADALAT 5 |
| 30491 | | | DILTIAZEM |
| 36684 | 193285001000027105 | | SLOFEDIPINE 20 MG TAB |
| **Diuretics** | | | |
| 4044 | 348911000001105 | Xipamide | Diurexan 20mg tablets (Meda Pharmaceuticals Ltd) |
| 7618 | 317970008 | Xipamide | Xipamide 20mg tablets |
| 9223 | 145145001000027101 | Triamterene/ Hydrochlorothiazide | Triamterene with hydrochlorothiazide 50mg + 25mg Tablet |
| 15127 | 116085001000027104 | Triamterene/ Hydrochlorothiazide | Hydrochlorothiazide with triamterene 25mgwith50mg Tablet |
| 2961 | 25411000001108 | Triamterene/ Furosemide | Frusene 50mg/ 40mg tablets (Orion Pharma (UK) Ltd) |
| 3050 | 107985001000027109 | Triamterene/ Furosemide | Furosemide with triamterene 40mgwith50mg Tablet |
| 11265 | 318101005 | Triamterene/ Furosemide | Triamterene 50mg / Furosemide 40mg tablets |
| 28157 | 40985001000027105 | Triamterene/ Chlortalidone | Kalspare ls Tablet (Dominion Pharma) |
| 37294 | 145265001000027105 | Triamterene/ Chlortalidone | Triamterene with chlortalidone 50mg + 25mg Tablet |
| 7136 | 714911000001108 | Triamterene/ Benzthiazide | Dytide capsules (Mercury Pharma Group Ltd) |
| 7740 | 318098006 | Triamterene/ Benzthiazide | Triamterene 50mg / Benzthiazide 25mg capsules |
| 2179 | 318082004 | Triamterene | Triamterene 50mg capsules |
| 4068 | 3907911000001100 | Triamterene | Dytac 50mg capsules (AMCo) |
| 8052 | 318041004 | Torasemide | Torasemide 5mg tablets |
| 10066 | 3700311000001109 | Torasemide | Torem 5mg tablets (Meda Pharmaceuticals Ltd) |
| 11268 | 3699711000001107 | Torasemide | Torem 2.5mg tablets (Meda Pharmaceuticals Ltd) |
| 11487 | 318040003 | Torasemide | Torasemide 2.5mg tablets |
| 18096 | 318042006 | Torasemide | Torasemide 10mg tablets |
| 22658 | 3700811000001100 | Torasemide | Torem 10mg tablets (Meda Pharmaceuticals Ltd) |
| 40190 | 198075001000027106 | Torasemide | Torasemide iv 20mg/ 4ml Intravenous injection |
| 40738 | 198025001000027107 | Torasemide | Torem iv 10mg/ 2ml Intravenous injection (Boehringer Mannheim UK Ltd) |
| 40898 | 5889911000001108 | Torasemide | Torasemide 5mg tablets (A A H Pharmaceuticals Ltd) |
| 46525 | 5592211000001107 | Torasemide | Torasemide 5mg tablets (Teva UK Ltd) |
| 1297 | 4669111000001107 | Spironolactone/ Hydroflumethiazide | Aldactide 50 tablets (Pfizer Ltd) |

| | | | |
|---|---|---|---|
| 2001 | 762511000001105 | Spironolactone/ Hydroflumethiazide | Aldactide 25 tablets (Pfizer Ltd) |
| 7961 | 141515001000027104 | Spironolactone/ Hydroflumethiazide | Spironolactone 50mg with hydroflumethiazide 50mg tablet |
| 8521 | 141505001000027101 | Spironolactone/ Hydroflumethiazide | Spironolactone 25mg with hydroflumethiazide 25mg tablet |
| 11384 | 318128000 | Spironolactone/ Hydroflumethiazide | Co-flumactone 50mg/ 50mg tablets |
| 15811 | 318127005 | Spironolactone/ Hydroflumethiazide | Co-flumactone 25mg/ 25mg tablets |
| 25505 | 196775001000027101 | Spironolactone/ Hydroflumethiazide | Spiro-co 50mg+50mg Tablet (IVAX Pharmaceuticals UK Ltd) |
| 29529 | 116225001000027101 | Spironolactone/ Hydroflumethiazide | Hydroflumethiazide with spironolactone 25mg+25mg Tablet |
| 31131 | 196765001000027108 | Spironolactone/ Hydroflumethiazide | Spiro-co 25mg+25mg Tablet (IVAX Pharmaceuticals UK Ltd) |
| 45916 | 116235001000027104 | Spironolactone/ Hydroflumethiazide | Hydroflumethiazide with spironolactone 50mg+50mg Tablet |
| 4661 | 318102003 | Spironolactone/ Furosemide | Spironolactone 50mg / Furosemide 20mg capsules |
| 7441 | 3645811000001107 | Spironolactone/ Furosemide | Lasilactone 20mg/ 50mg capsules (Sanofi) |
| 53508 | 12424811000001104 | Spironolactone/ Chlorothiazide | Spironolactone 5mg/ 5ml / Chlorothiazide 50mg/ 5ml oral suspension |
| 692 | 318056008 | Spironolactone | Spironolactone 25mg tablets |
| 708 | 318057004 | Spironolactone | Spironolactone 50mg tablets |
| 787 | 141475001000027103 | Spironolactone | Spironolactone 100mg capsule |
| 2142 | 318058009 | Spironolactone | Spironolactone 100mg tablets |
| 2389 | 930511000001105 | Spironolactone | Aldactone 25mg tablets (Pfizer Ltd) |
| 4161 | 84985001000027102 | Spironolactone | Spiroctan 25mg Tablet (Roche Products Ltd) |
| 4960 | 921811000001103 | Spironolactone | Aldactone 50mg tablets (Pfizer Ltd) |
| 6815 | 196545001000027101 | Spironolactone | Spironolactone 50mg/ 5ml oral suspension sugar free |
| 7952 | 421611000001100 | Spironolactone | Aldactone 100mg tablets (Pfizer Ltd) |
| 7991 | 85005001000027103 | Spironolactone | Spiroctan 100mg Capsule (Roche Products Ltd) |
| 10214 | 196365001000027109 | Spironolactone | Spironolactone 5mg/ 5ml oral suspension sugar free |
| 11156 | 40795001000027101 | Spironolactone | Spirolone 25mg Tablet (Berk Pharmaceuticals Ltd) |
| 11519 | 196385001000027107 | Spironolactone | Spironolactone 25mg/ 5ml oral suspension sugar free |
| 12946 | 196375001000027102 | Spironolactone | Spironolactone 10mg/ 5ml oral suspension sugar free |
| 13264 | 8727011000001103 | Spironolactone | Spironolactone 15mg/ 5ml oral suspension |
| 14109 | 196555001000027103 | Spironolactone | Spironolactone 100mg/ 5ml oral solution sugar free |
| 15052 | 84995001000027103 | Spironolactone | Spiroctan 50mg Tablet (Roche Products Ltd) |
| 17902 | 40815001000027104 | Spironolactone | Spirolone 100mg Tablet (Berk Pharmaceuticals Ltd) |
| 17950 | 40805001000027101 | Spironolactone | Spirolone 50mg Tablet (Berk Pharmaceuticals Ltd) |
| 19195 | 60755001000027100 | Spironolactone | Spironolactone 50mg Tablet (Wyeth Pharmaceuticals) |
| 21911 | 155055001000027102 | Spironolactone | Spirospare 25mg Tablet (Ashbourne Pharmaceuticals Ltd) |
| 23091 | 3411000001104 | Spironolactone | Spirospare 100 tablets (Ashbourne Pharmaceuticals Ltd) |
| 25494 | 75095001000027104 | Spironolactone | Diatensec 50mg Tablet (Pharmacia Ltd) |
| 29397 | 87695001000027105 | Spironolactone | Spiretic 100mg Tablet (DDSA Pharmaceuticals Ltd) |
| 31219 | 670811000001106 | Spironolactone | Spironolactone 100mg tablets (A A H Pharmaceuticals Ltd) |
| 31529 | 82311000001101 | Spironolactone | Spironolactone 25mg tablets (Teva UK Ltd) |
| 32837 | 330511000001102 | Spironolactone | Spironolactone 50mg tablets (Teva UK Ltd) |
| 34296 | 63111000001106 | Spironolactone | Spironolactone 25mg tablets (A A H Pharmaceuticals Ltd) |
| 34347 | 672411000001105 | Spironolactone | Spironolactone 25mg tablets (Actavis UK Ltd) |
| 34908 | 474611000001102 | Spironolactone | Spironolactone 25mg tablets (IVAX Pharmaceuticals UK Ltd) |
| 35789 | 20575001000027108 | Spironolactone | Spironolactone 25mg Tablet (Celltech Pharma Europe Ltd) |

| | | | |
|---|---|---|---|
| 41074 | 9803611000001107 | Spironolactone | Spironolactone 25mg tablets (Almus Pharmaceuticals Ltd) |
| 41592 | 38611000001103 | Spironolactone | Spironolactone 100mg tablets (Actavis UK Ltd) |
| 41660 | 610611000001103 | Spironolactone | Spironolactone 100mg tablets (Teva UK Ltd) |
| 41706 | 121211000001101 | Spironolactone | Spironolactone 50mg tablets (IVAX Pharmaceuticals UK Ltd) |
| 43514 | 481811000001107 | Spironolactone | Spironolactone 50mg tablets (A A H Pharmaceuticals Ltd) |
| 45078 | 60285001000027101 | Spironolactone | Spironolactone 25mg/ 5ml Oral solution sugar free (Rosemont Pharmaceuticals Ltd) |
| 46674 | 60315001000027106 | Spironolactone | Spironolactone 50mg/ 5ml Oral suspension sugar free (Rosemont Pharmaceuticals Ltd) |
| 46990 | 8726411000001105 | Spironolactone | Spironolactone 50mg/ 5ml oral suspension |
| 47018 | 8726511000001109 | Spironolactone | Spironolactone 25mg/ 5ml oral suspension |
| 47687 | 87685001000027106 | Spironolactone | Spiretic 25mg Tablet (DDSA Pharmaceuticals Ltd) |
| 49388 | 8727611000001105 | Spironolactone | Spironolactone 100mg/ 5ml oral suspension |
| 50079 | 8727311000001100 | Spironolactone | Spironolactone 10mg/ 5ml oral suspension |
| 50370 | 8726311000001103 | Spironolactone | Spironolactone 5mg/ 5ml oral suspension |
| 51652 | 13583411000001106 | Spironolactone | Spironolactone 25mg tablets (DE Pharmaceuticals) |
| 51720 | 13894811000001108 | Spironolactone | Spironolactone 25mg/ 5ml oral solution |
| 51933 | 13894911000001103 | Spironolactone | Spironolactone 50mg/ 5ml oral solution |
| 52366 | 13895011000001103 | Spironolactone | Spironolactone 5mg/ 5ml oral solution |
| 52970 | 13894711000001100 | Spironolactone | Spironolactone 10mg/ 5ml oral solution |
| 53253 | 8705811000001105 | Spironolactone | Spironolactone 50mg/ 5ml oral suspension (Drug Tariff Special Order) |
| 54120 | 13374211000001104 | Spironolactone | Spironolactone 4mg/ 5ml oral suspension |
| 56067 | 13374111000001105 | Spironolactone | Spironolactone 4mg/ 5ml oral solution |
| 56274 | 13355011000001105 | Spironolactone | Spironolactone 4.5mg/ 5ml oral suspension |
| 56536 | 13894611000001109 | Spironolactone | Spironolactone 100mg/ 5ml oral solution |
| 57104 | 13326311000001102 | Spironolactone | Spironolactone 200mg/ 5ml oral suspension |
| 57556 | 13325611000001102 | Spironolactone | Spironolactone 12mg/ 5ml oral solution |
| 57933 | 13374011000001109 | Spironolactone | Spironolactone 40mg/ 5ml oral suspension |
| 58077 | 13385811000001105 | Spironolactone | Spironolactone 8mg/ 5ml oral suspension |
| 58225 | 13326111000001104 | Spironolactone | Spironolactone 2.5mg/ 5ml oral suspension |
| 58757 | 13326511000001108 | Spironolactone | Spironolactone 20mg/ 5ml oral suspension |
| 60343 | 235211000001102 | Spironolactone | Spironolactone 25mg tablets (Kent Pharmaceuticals Ltd) |
| 60660 | 13354811000001100 | Spironolactone | Spironolactone 3mg/ 5ml oral suspension |
| 61025 | 13326411000001109 | Spironolactone | Spironolactone 20mg/ 5ml oral solution |
| 63309 | 13384811000001109 | Spironolactone | Spironolactone 6mg/ 5ml oral suspension |
| 65582 | 13354711000001108 | Spironolactone | Spironolactone 3mg/ 5ml oral solution |
| 65822 | 13325311000001107 | Spironolactone | Spironolactone 12.5mg/ 5ml oral suspension |
| 67913 | 606411000001101 | Spironolactone | Spironolactone 50mg tablets (Kent Pharmaceuticals Ltd) |
| 69473 | 13354211000001101 | Spironolactone | Spironolactone 3.5mg/ 5ml oral suspension |
| 71010 | 20326811000001106 | Spironolactone | Spironolactone 25mg tablets (Genesis Pharmaceuticals Ltd) |
| 71398 | 13325211000001104 | Spironolactone | Spironolactone 12.5mg/ 5ml oral solution |
| 73208 | 13385411000001108 | Spironolactone | Spironolactone 7mg/ 5ml oral suspension |
| 73644 | 11010911000001104 | Spironolactone | Spironolactone 25mg tablets (Dr Reddy's Laboratories (UK) Ltd) |
| 74154 | 13353811000001103 | Spironolactone | Spironolactone 2mg/ 5ml oral suspension |
| 74285 | 8707011000001106 | Spironolactone | Spironolactone 25mg/ 5ml oral suspension (Drug Tariff Special Order) |

| | | | |
|---|---|---|---|
| 74631 | 46011000001105 | Spironolactone | Spironolactone 50mg tablets (Actavis UK Ltd) |
| 75413 | | Spironolactone | Spironolactone 8mg/ 5ml oral solution (Special Order) |
| 75488 | | Spironolactone | Spironolactone 250mg/ 5ml oral suspension |
| 75593 | | Spironolactone | Spironolactone 10mg/ 5ml Oral solution sugar free (Rosemont Pharmaceuticals Ltd) |
| 76195 | | Spironolactone | Spironolactone 5mg/ 5ml Oral suspension sugar free (Rosemont Pharmaceuticals Ltd) |
| 76321 | | Spironolactone | Spironolactone 100mg tablets (Genesis Pharmaceuticals Ltd) |
| 77604 | | Spironolactone | Spironolactone 7.5mg/ 5ml oral suspension |
| 77610 | | Spironolactone | Spironolactone 6.25mg/ 5ml oral suspension |
| 7582 | 3704811000001105 | Potassium chloride/ Furosemide | Lasikal modified-release tablets (Borg Medicare) |
| 7734 | 4540011000001102 | Potassium chloride/ Furosemide | Diumide-K Continus tablets (Teofarma) |
| 8102 | 4557711000001102 | Potassium chloride/ Furosemide | Furosemide 40mg / Potassium chloride 600mg (potassium 8mmol) modified-release tablets |
| 10781 | 37075001000027100 | Potassium chloride/ Furosemide | Lasix with k Tablet (Hoechst Marion Roussel) |
| 17960 | 36061311000001109 | Potassium chloride/ Furosemide | Furosemide 20mg / Potassium chloride 750mg (potassium 10mmol) modified-release tablets |
| 18983 | 3875001000027105 | Potassium Chloride/ Clopamide | Brinaldix k Effervescent tablet (Berk Pharmaceuticals Ltd) |
| 22839 | 209335001000027100 | Potassium Chloride/ Clopamide | Clopamide with Potassium effervescent tablets |
| 8891 | 14465001000027109 | Potassium Chloride/ Chlortalidone | Hygroton -k Tablet (Novartis Pharmaceuticals UK Ltd) |
| 1776 | 3290211000001109 | Potassium chloride/ Bumetanide | Burinex K modified-release tablets (LEO Pharma) |
| 6160 | 35913011000001100 | Potassium chloride/ Bumetanide | Bumetanide 500microgram / Potassium chloride 573mg (potassium 7.7mmol) modified-release tablets |
| 12360 | 20915001000027105 | Polythiazide | "Nephril 1mg Tablet (Pfizer Ltd)" |
| 12926 | 317967009 | Polythiazide | "Polythiazide 1mg tablets" |
| 7709 | 39735001000027105 | Piretanide | Arelix 6mg Capsule (Hoechst Marion Roussel) |
| 12367 | 135585001000027102 | Piretanide | Piretanide 6mg capsule |
| 4332 | 317965001 | Metolazone | Metolazone 5mg tablets |
| 4334 | 115735001000027107 | Metolazone | Metolazone 500microgram low dose Tablet |
| 8602 | 375611000001109 | Metolazone | Metenix 5mg tablets (Sanofi) |
| 19352 | 176195001000027101 | Metolazone | Xuret 0.5mg Tablet (Galen Ltd) |
| 49752 | 13005911000001103 | Metolazone | Metolazone 2.5mg/ 5ml oral solution |
| 53674 | 374173002 | Metolazone | Metolazone 2.5mg tablets |
| 54329 | 13006411000001102 | Metolazone | Metolazone 5mg/ 5ml oral suspension |
| 54643 | 251255001000027106 | Metolazone | Metolazone Oral solution |
| 55777 | 13006311000001109 | Metolazone | Metolazone 5mg/ 5ml oral solution |
| 61846 | 503445001000027107 | Metolazone | Zaroxolyn 2.5mg tablets (IDIS) |
| 68432 | 13006011000001106 | Metolazone | Metolazone 2.5mg/ 5ml oral suspension |
| 69009 | 21574611000001109 | Metolazone | Zaroxolyn 5mg tablets (Imported (Canada)) |
| 75260 | 21574911000001103 | Metolazone | Zaroxolyn 2.5mg tablets (Imported (Canada)) |
| 18267 | 124815001000027107 | Methyclothiazide | "Enduron 5mg Tablet (Abbott Laboratories Ltd)" |
| 20057 | 124785001000027106 | Methyclothiazide | "Methyclothiazide 5mg Tablet" |
| 31013 | 124005001000027109 | Mersalyl | Mersalyl 50mg/ ml Injection |
| 15457 | 2515001000027106 | Mefruside | Baycaron 25mg Tablet (Bayer Plc) |
| 17143 | 123565001000027105 | Mefruside | Mefruside 25mg Tablet |
| 2612 | 317956008 | Indapamide hemihydrate | Indapamide 2.5mg tablets |
| 7641 | 321811000001109 | Indapamide hemihydrate | Natrilix 2.5mg tablets (Servier Laboratories Ltd) |

| | | | |
|---|---|---|---|
| 26256 | 209045001000027100 | Indapamide hemihydrate | Opumide 2.5mg Tablet (Opus Pharmaceuticals Ltd) |
| 26275 | 424311000001108 | Indapamide hemihydrate | Nindaxa 2.5 tablets (Ashbourne Pharmaceuticals Ltd) |
| 27957 | 207505001000027101 | Indapamide hemihydrate | Natramid 2.5mg Tablet (Trinity Pharmaceuticals Ltd) |
| 33083 | 711711000001108 | Indapamide hemihydrate | Indapamide 2.5mg tablets (Teva UK Ltd) |
| 34551 | 450511000001107 | Indapamide hemihydrate | Indapamide 2.5mg tablets (Mylan) |
| 40907 | 8139411000001107 | Indapamide hemihydrate | Indapamide 2.5mg tablets (Genus Pharmaceuticals Ltd) |
| 42906 | 10437611000001100 | Indapamide hemihydrate | Indapamide 2.5mg tablets (Niche Generics Ltd) |
| 43516 | 397311000001100 | Indapamide hemihydrate | Indapamide 2.5mg tablets (Actavis UK Ltd) |
| 48079 | 214811000001107 | Indapamide hemihydrate | Indapamide 2.5mg tablets (Zentiva) |
| 48099 | 56211000001102 | Indapamide hemihydrate | Indapamide 2.5mg tablets (A A H Pharmaceuticals Ltd) |
| 49529 | 17932311000001102 | Indapamide hemihydrate | Indapamide 2.5mg tablets (Phoenix Healthcare Distribution Ltd) |
| 54316 | 598411000001109 | Indapamide hemihydrate | Indapamide 2.5mg tablets (Alliance Healthcare (Distribution) Ltd) |
| 55259 | 146711000001100 | Indapamide hemihydrate | Indapamide 2.5mg tablets (Kent Pharmaceuticals Ltd) |
| 56296 | 14784911000001103 | Indapamide hemihydrate | Indapamide 2.5mg tablets (Boston Healthcare Ltd) |
| 56760 | 18283111000001109 | Indapamide hemihydrate | Indapamide 2.5mg tablets (Strides Shasun (UK) Ltd) |
| 70509 | 13579511000001104 | Indapamide hemihydrate | Indapamide 2.5mg tablets (DE Pharmaceuticals) |
| 77406 | | Indapamide hemihydrate | Natrilix 2.5mg tablets (Mawdsley-Brooks & Company Ltd) |
| 3056 | 456611000001108 | Indapamide | Natrilix SR 1.5mg tablets (Servier Laboratories Ltd) |
| 5112 | 317954006 | Indapamide | Indapamide 1.5mg modified-release tablets |
| 39447 | 14693611000001102 | Indapamide | Varbim XL 1.5mg tablets (Teva UK Ltd) |
| 41861 | 14242211000001100 | Indapamide | Tensaid XL 1.5mg tablets (Mylan) |
| 41885 | 13824811000001102 | Indapamide | Ethibide XL 1.5mg tablets (Genus Pharmaceuticals Ltd) |
| 43184 | 15600711000001107 | Indapamide | Mapemid XL 1.5mg tablets (Teva UK Ltd) |
| 44168 | 15436111000001104 | Indapamide | Indipam XL 1.5mg tablets (Actavis UK Ltd) |
| 46675 | 14033311000001105 | Indapamide | Indapamide 1.5mg modified-release tablets (A A H Pharmaceuticals Ltd) |
| 59616 | 16737911000001105 | Indapamide | Rawel XL 1.5mg tablets (Consilient Health Ltd) |
| 60020 | 22058211000001103 | Indapamide | Indapamide 1.5mg modified-release tablets (Waymade Healthcare Plc) |
| 62066 | 24331611000001104 | Indapamide | Cardide SR 1.5mg tablets (Teva UK Ltd) |
| 62771 | 24175711000001103 | Indapamide | Indapamide 1.5mg modified-release tablets (DE Pharmaceuticals) |
| 64066 | 21578911000001105 | Indapamide | Indapamide 2.5mg/ 5ml oral suspension |
| 74172 | 15101611000001109 | Indapamide | Indapamide 1.5mg modified-release tablets (Sigma Pharmaceuticals Plc) |
| 77449 | | Indapamide | Natrilix SR 1.5mg tablets (Waymade Healthcare Plc) |
| 12110 | 116195001000027108 | Hydroflumethiazide | "Hydroflumethiazide 50mg Tablet" |
| 13525 | 80235001000027106 | Hydroflumethiazide | "Hydrenox 50mg Tablet (Knoll Ltd)" |
| 1721 | 132811000001106 | Hydrochlorothiazide/ Triamterene | Dyazide 50mg/ 25mg tablets (AMCo) |
| 5416 | 410896007 | Hydrochlorothiazide/ Triamterene | Co-triamterzide 50mg/ 25mg tablets |
| 8897 | 191211000001106 | Hydrochlorothiazide/ Triamterene | Triam-Co 50mg/ 25mg tablets (IVAX Pharmaceuticals UK Ltd) |
| 18726 | 721311000001104 | Hydrochlorothiazide/ Triamterene | Triamaxco 50mg/ 25mg tablets (Ashbourne Pharmaceuticals Ltd) |
| 47804 | 12779411000001108 | Hydrochlorothiazide/ Triamterene | Co-triamterzide 50mg/ 25mg tablets (A A H Pharmaceuticals Ltd) |
| 67801 | 16160111000001109 | Hydrochlorothiazide/ Triamterene | Dyazide 50mg/ 25mg tablets (Lexon (UK) Ltd) |
| 77386 | | Hydrochlorothiazide/ Triamterene | Dyazide 50mg/ 25mg tablets (Dowelhurst Ltd) |
| 924 | 318121006 | Hydrochlorothiazide/ Amiloride | Co-amilozide 2.5mg/ 25mg tablets |
| 1251 | 314211000001100 | Hydrochlorothiazide/ Amiloride | Moduret 25 tablets (Merck Sharp & Dohme Ltd) |

| | | | |
|---|---|---|---|
| 34367 | 925611000001104 | Hydrochlorothiazide/ Amiloride | Co-amilozide 2.5mg/ 25mg tablets (Wockhardt UK Ltd) |
| 60354 | 495611000001107 | Hydrochlorothiazide/ Amiloride | Co-amilozide 2.5mg/ 25mg tablets (Kent Pharmaceuticals Ltd) |
| 542 | 376209006 | Hydrochlorothiazide | "Hydrochlorothiazide 25mg tablets" |
| 3517 | 376508004 | Hydrochlorothiazide | "Hydrochlorothiazide 50mg tablets" |
| 12440 | 4544011000001109 | Hydrochlorothiazide | "Hydrosaluric 25mg tablets (Merck Sharp & Dohme Ltd)" |
| 13363 | 10555001000027106 | Hydrochlorothiazide | "Esidrex 50mg Tablet (Novartis Pharmaceuticals UK Ltd)" |
| 16632 | 4546211000001109 | Hydrochlorothiazide | "Hydrosaluric 50mg tablets (Merck Sharp & Dohme Ltd)" |
| 17252 | 10545001000027108 | Hydrochlorothiazide | "Esidrex 25mg Tablet (Novartis Pharmaceuticals UK Ltd)" |
| 48132 | 250545001000027106 | Hydrochlorothiazide | "Hydrochlorothiazide Capsule" |
| 57488 | 246525001000027108 | Hydrochlorothiazide | "Hydrochlorothiazide Oral solution" |
| 62516 | 24594211000001108 | Hydrochlorothiazide | "Hydrochlorothiazide 12.5mg tablets" |
| 1369 | 107935001000027101 | Furosemide/ Amiloride | Furosemide with amiloride 40mg+5mg Tablet |
| 4211 | 107945001000027100 | Furosemide/ Amiloride | Furosemide with amiloride 20mg+2.5mg Tablet |
| 5220 | 107955001000027102 | Furosemide/ Amiloride | Furosemide with amiloride 80mg+10mg Tablet |
| 9456 | 159445001000027103 | Furosemide/ Amiloride | Amiloride 5mg / furosemide 40mg tablets |
| 15874 | 159435001000027104 | Furosemide/ Amiloride | Amiloride 2.5mg / furosemide 20mg tablets |
| 18497 | 159455001000027100 | Furosemide/ Amiloride | Amiloride 10mg / furosemide 80mg tablets |
| 47647 | 245685001000027108 | Furosemide/ Amiloride | Co-amilofruse oral liquid |
| 6 | 317972000 | Furosemide | Furosemide 40mg tablets |
| 55 | 317971007 | Furosemide | Furosemide 20mg tablets |
| 562 | 107885001000027101 | Furosemide | Furosemide 10mg/ ml Injection |
| 3248 | 317973005 | Furosemide | Furosemide 500mg tablets |
| 3287 | 199505001000027101 | Furosemide | Furosemide 1mg/ ml Oral solution |
| 4182 | 701111000001105 | Furosemide | Lasix 5mg/ 5ml oral solution (Borg Medicare) |
| 4258 | 9611000001104 | Furosemide | Lasix 20mg/ 2ml solution for injection ampoules (Sanofi) |
| 4705 | 145335001000027109 | Furosemide | Furosemide 20mg/ 2ml Injection |
| 5249 | 36564411000001102 | Furosemide | Furosemide 50mg/ 5ml oral solution sugar free |
| 5728 | 318007007 | Furosemide | Furosemide 40mg/ 5ml oral solution sugar free |
| 5868 | 602811000001100 | Furosemide | Frusol 20mg/ 5ml oral solution (Rosemont Pharmaceuticals Ltd) |
| 6118 | 318006003 | Furosemide | Furosemide 20mg/ 5ml oral solution sugar free |
| 7606 | 79411000001107 | Furosemide | Lasix 40mg tablets (Sanofi) |
| 7799 | 701611000001102 | Furosemide | Lasix 20mg tablets (Borg Medicare) |
| 9680 | 494811000001105 | Furosemide | Frusol 40mg/ 5ml oral solution (Rosemont Pharmaceuticals Ltd) |
| 10392 | 829911000001102 | Furosemide | Lasix 500mg tablets (Sanofi) |
| 10422 | 37095001000027109 | Furosemide | Lasix 50mg/ 5ml Injection (Hoechst UK Ltd) |
| 12318 | 153285001000027109 | Furosemide | Lasix 250mg/ 25ml Injection (Hoechst Marion Roussel) |
| 14761 | 99311000001108 | Furosemide | Frusid 40mg tablets (Dr Reddy's Laboratories (UK) Ltd) |
| 14837 | 855611000001109 | Furosemide | Frusol 50mg/ 5ml oral solution (Rosemont Pharmaceuticals Ltd) |
| 16206 | 714311000001107 | Furosemide | Froop 40mg tablets (Ashbourne Pharmaceuticals Ltd) |
| 18716 | 40555001000027108 | Furosemide | Dryptal 10mg/ ml Injection (Berk Pharmaceuticals Ltd) |
| 19056 | 60235001000027109 | Furosemide | Furosemide 50mg/ 5ml sugar free Oral solution (Rosemont Pharmaceuticals Ltd) |
| 19192 | 118425001000027108 | Furosemide | Furosemide 40mg Tablet (M & A Pharmachem Ltd) |
| 19194 | 17211000001105 | Furosemide | Furosemide 20mg tablets (Teva UK Ltd) |

| | | | |
|---|---|---|---|
| 19258 | 36061511000001103 | Furosemide | Furosemide 50mg/ 5ml solution for injection ampoules |
| 20538 | 1795001000027107 | Furosemide | Frumax 40mg Tablet (Ashbourne Pharmaceuticals Ltd) |
| 21849 | 8805001000027103 | Furosemide | Dryptal 40mg Tablet (Berk Pharmaceuticals Ltd) |
| 24835 | 23365001000027103 | Furosemide | Min-i-jet furosemide 10mg/ ml Injection (Celltech Pharma Europe Ltd) |
| 25334 | 119611000001105 | Furosemide | Furosemide 500mg tablets (A A H Pharmaceuticals Ltd) |
| 25717 | 147711000001102 | Furosemide | Furosemide 40mg tablets (Mylan) |
| 26292 | 82155001000027100 | Furosemide | Diuresal 40mg Tablet (Lagap) |
| 27447 | 445111000001108 | Furosemide | Furosemide 40mg tablets (Wockhardt UK Ltd) |
| 27690 | 100611000001101 | Furosemide | Furosemide 40mg tablets (A A H Pharmaceuticals Ltd) |
| 27696 | 765411000001103 | Furosemide | Furosemide 40mg tablets (Kent Pharmaceuticals Ltd) |
| 27926 | 229711000001109 | Furosemide | Furosemide 20mg tablets (Mylan) |
| 29780 | 13295001000027107 | Furosemide | Furosemide 20mg Tablet (C P Pharmaceuticals Ltd) |
| 30625 | 202711000001106 | Furosemide | Furosemide 20mg tablets (A A H Pharmaceuticals Ltd) |
| 30875 | 3419371000001108 | Furosemide | Furosemide 250mg/ 25ml solution for injection ampoules |
| 31548 | 631611000001100 | Furosemide | Furosemide 20mg tablets (Actavis UK Ltd) |
| 32277 | 36061611000001104 | Furosemide | Furosemide 80mg/ 8ml solution for injection pre-filled syringes |
| 32896 | 8751711000001106 | Furosemide | Furosemide 40mg tablets (Ranbaxy (UK) Ltd) |
| 32918 | 662811000001104 | Furosemide | Furosemide 20mg tablets (Sandoz Ltd) |
| 34006 | 351511000001105 | Furosemide | Furosemide 40mg tablets (Actavis UK Ltd) |
| 34374 | 472211000001108 | Furosemide | Furosemide 40mg tablets (Teva UK Ltd) |
| 34557 | 856211000001101 | Furosemide | Furosemide 40mg tablets (IVAX Pharmaceuticals UK Ltd) |
| 35162 | 36061411000001102 | Furosemide | Furosemide 20mg/ 2ml solution for injection ampoules |
| 36190 | 318010000 | Furosemide | Furosemide 5mg/ 5ml oral solution sugar free |
| 40247 | 46575001000027100 | Furosemide | Furosemide 10mg/ ml Injection (Martindale Pharmaceuticals Ltd) |
| 41292 | 539911000001101 | Furosemide | Furosemide 20mg tablets (Wockhardt UK Ltd) |
| 41405 | 752711000001106 | Furosemide | Furosemide 500mg tablets (Teva UK Ltd) |
| 41828 | 822211000001103 | Furosemide | Furosemide 500mg tablets (Actavis UK Ltd) |
| 42388 | 10305811000001105 | Furosemide | Furosemide 40mg/ 5ml oral solution sugar free (Focus Pharmaceuticals Ltd) |
| 42488 | 10214011000001105 | Furosemide | Furosemide 40mg/ 5ml oral solution sugar free (A A H Pharmaceuticals Ltd) |
| 46116 | 58505001000027108 | Furosemide | Furosemide 10mg/ ml Injection (Antigen Pharmaceuticals) |
| 46699 | 9801311000001103 | Furosemide | Furosemide 40mg tablets (Almus Pharmaceuticals Ltd) |
| 46948 | 10429011000001100 | Furosemide | Furosemide 40mg tablets (Arrow Generics Ltd) |
| 47815 | 13335001000027109 | Furosemide | Furosemide 20mg Tablet (Celltech Pharma Europe Ltd) |
| 49268 | 12088211000001109 | Furosemide | Furosemide 50mg/ 5ml oral suspension |
| 51983 | 13893511000001107 | Furosemide | Furosemide 5mg/ 5ml oral suspension |
| 52045 | 20176911000001106 | Furosemide | Furosemide 250mg/ 5ml solution for injection vials |
| 52887 | 725611000001107 | Furosemide | Furosemide 20mg/ 2ml solution for injection ampoules (A A H Pharmaceuticals Ltd) |
| 52900 | 2898811000001106 | Furosemide | Furosemide 80mg/ 8ml solution for injection Minijet pre-filled syringes (UCB Pharma Ltd) |
| 53967 | 16052611000001100 | Furosemide | Furosemide 20mg tablets (Bristol Laboratories Ltd) |
| 54825 | 15089311000001105 | Furosemide | Furosemide 20mg tablets (Sigma Pharmaceuticals Plc) |
| 55738 | 5088711000001101 | Furosemide | Furosemide 50mg/ 5ml solution for injection ampoules (Hameln Pharmaceuticals Ltd) |
| 56051 | 503711000001106 | Furosemide | Furosemide 20mg tablets (Kent Pharmaceuticals Ltd) |
| 56375 | 18461911000001109 | Furosemide | Furosemide 40mg tablets (Accord Healthcare Ltd) |

| | | | |
|---|---|---|---|
| 57600 | 120011000001102 | Furosemide | Furosemide 20mg/ 2ml solution for injection ampoules (Alliance Healthcare (Distribution) Ltd) |
| 57610 | 10305611000001106 | Furosemide | Furosemide 20mg/ 5ml oral solution sugar free (Focus Pharmaceuticals Ltd) |
| 58078 | 12088911000001100 | Furosemide | Furosemide 8mg/ 5ml oral solution |
| 58224 | 12035011000001100 | Furosemide | Furosemide 10mg/ 5ml oral solution |
| 59030 | 12036211000001104 | Furosemide | Furosemide 20mg/ 5ml oral solution |
| 59290 | 121411000001102 | Furosemide | Furosemide 20mg tablets (Alliance Healthcare (Distribution) Ltd) |
| 59884 | 17911411000001106 | Furosemide | Furosemide 20mg tablets (Phoenix Healthcare Distribution Ltd) |
| 59911 | 139011000001107 | Furosemide | Furosemide 40mg tablets (Alliance Healthcare (Distribution) Ltd) |
| 59939 | 12036311000001107 | Furosemide | Furosemide 20mg/ 5ml oral suspension |
| 60291 | 19309711000001101 | Furosemide | Furosemide 40mg tablets (AMCo) |
| 60465 | 13893411000001108 | Furosemide | Furosemide 5mg/ 5ml oral solution |
| 61365 | 12087811000001106 | Furosemide | Furosemide 40mg/ 5ml oral suspension |
| 61475 | 23909911000001105 | Furosemide | Furosemide 20mg tablets (DE Pharmaceuticals) |
| 63237 | 14789111000001100 | Furosemide | Furosemide 20mg tablets (Boston Healthcare Ltd) |
| 64255 | 12036811000001103 | Furosemide | Furosemide 2mg/ 5ml oral solution |
| 64677 | 23910211000001105 | Furosemide | Furosemide 40mg tablets (DE Pharmaceuticals) |
| 64745 | 12089011000001109 | Furosemide | Furosemide 8mg/ 5ml oral suspension |
| 65583 | 12087311000001102 | Furosemide | Furosemide 3mg/ 5ml oral solution |
| 66017 | 9800811000001104 | Furosemide | Furosemide 20mg tablets (Almus Pharmaceuticals Ltd) |
| 66149 | 12087511000001108 | Furosemide | Furosemide 4.5mg/ 5ml oral solution |
| 67910 | 12087711000001103 | Furosemide | Furosemide 40mg/ 5ml oral solution |
| 68068 | 8519911000001102 | Furosemide | Furosemide 4mg/ 5ml oral solution |
| 69338 | 60225001000027107 | Furosemide | Furosemide 40mg/ 5ml sugar free Oral solution (Rosemont Pharmaceuticals Ltd) |
| 69445 | 18706911000001106 | Furosemide | Furosemide 40mg/ 5ml oral solution sugar free (Sigma Pharmaceuticals Plc) |
| 70650 | 34537111000001101 | Furosemide | Furosemide 50mg/ 5ml solution for injection ampoules (Peckforton Pharmaceuticals Ltd) |
| 71406 | 12035811000001106 | Furosemide | Furosemide 1mg/ 5ml oral solution |
| 71950 | 22018711000001104 | Furosemide | Furosemide 20mg tablets (Waymade Healthcare Plc) |
| 73171 | 246455001000027101 | Furosemide | Furosemide Oral solution |
| 73480 | 60215001000027103 | Furosemide | Furosemide 20mg/ 5ml sugar free Oral solution (Rosemont Pharmaceuticals Ltd) |
| 74381 | 12087411000001109 | Furosemide | Furosemide 3mg/ 5ml oral suspension |
| 74677 | 30133911000001101 | Furosemide | Furosemide 40mg tablets (Mawdsley-Brooks & Company Ltd) |
| 74926 | 28986611000001105 | Furosemide | Furosemide 20mg tablets (Crescent Pharma Ltd) |
| 75855 | | Furosemide | Furosemide 10mg/ 5ml oral suspension |
| 76157 | | Furosemide | Furosemide 50mg/ 5ml oral solution |
| 76323 | | Furosemide | Furosemide 40mg tablets (Crescent Pharma Ltd) |
| 76934 | | Furosemide | Furosemide 6.5mg/ 5ml oral solution |
| 77333 | | Furosemide | Aluzine 20mg Tablet (M A Steinhard Ltd) |
| 77566 | | Furosemide | Furosemide 40mg tablets (Bristol Laboratories Ltd) |
| 12354 | 99155001000027108 | Etacrynic Acid | Etacrynic 50mg tablets |
| 18650 | 9585001000027101 | Etacrynic Acid | Edecrin 50mg Tablet (Merck Sharp & Dohme Ltd) |
| 32002 | 179865001000027103 | Etacrynic Acid | Etacrynic 50mg/ vial injection |
| 1170 | 317940003 | Cyclopenthiazide | "Cyclopenthiazide 500microgram tablets" |
| 2046 | 487811000001100 | Cyclopenthiazide | "Navidrex 500microgram tablets (AMCo)" |

| | | | |
|---|---|---|---|
| 12546 | 40975001000027100 | Chlortalidone/ Triamterene | Kalspare Tablet (Dominion Pharma) |
| 12547 | 318100006 | Chlortalidone/ Triamterene | Triamterene 50mg / Chlortalidone 50mg tablets |
| 16498 | 3252011000001105 | Chlortalidone/ Triamterene | Kalspare tablets (DHP Healthcare Ltd) |
| 605 | 317935006 | Chlortalidone | Chlortalidone 50mg tablets |
| 3054 | 14435001000027104 | Chlortalidone | Hygroton 100mg Tablet (Alliance Pharmaceuticals Ltd) |
| 3548 | 101645001000027102 | Chlortalidone | Chlortalidone 100mg tablets |
| 3997 | 285911000001101 | Chlortalidone | Hygroton 50mg tablets (Alliance Pharmaceuticals Ltd) |
| 74942 | 8359911000001106 | Chlortalidone | Chlortalidone 50mg/ 5ml oral suspension |
| 75383 | 376346007 | Chlortalidone | Chlortalidone 25mg tablets |
| 6816 | 408039005 | Chlorothiazide | "Chlorothiazide 250mg/5ml oral suspension" |
| 8836 | 101315001000027103 | Chlorothiazide | "Chlorothiazide 500mg tablets" |
| 13246 | 8358511000001105 | Chlorothiazide | "Chlorothiazide 150mg/5ml oral suspension" |
| 17720 | 27705001000027105 | Chlorothiazide | "Saluric 500mg Tablet (Merck Sharp & Dohme Ltd)" |
| 33724 | 7652811000001107 | Chlorothiazide | "Diuril 250mg/5ml oral suspension (Imported (United States))" |
| 54341 | 12015711000001107 | Chlorothiazide | "Chlorothiazide 5mg/5ml oral suspension" |
| 54679 | 395516007 | Chlorothiazide | "Chlorothiazide 250mg tablets" |
| 55889 | 243885001000027106 | Chlorothiazide | "Chlorothiazide oral solution" |
| 56804 | 8358711000001100 | Chlorothiazide | "Chlorothiazide 25mg/5ml oral suspension" |
| 59834 | 12503011000001109 | Chlorothiazide | "Chlorothiazide 250mg/5ml oral solution" |
| 60603 | 8359111000001108 | Chlorothiazide | "Chlorothiazide 50mg/5ml oral suspension" |
| 63227 | 12016611000001108 | Chlorothiazide | "Chlorothiazide 70mg/5ml oral solution" |
| 64798 | 12431311000001107 | Chlorothiazide | "Chlorothiazide 120mg/5ml oral solution" |
| 71871 | 8359211000001102 | Chlorothiazide | "Chlorothiazide 60mg/5ml oral suspension" |
| 73441 | 12502211000001105 | Chlorothiazide | "Chlorothiazide 200mg/5ml oral solution" |
| 74153 | 12431011000001109 | Chlorothiazide | "Chlorothiazide 10mg/5ml oral solution" |
| 75957 | | Chlorothiazide | "Chlorothiazide 5mg/5ml oral solution" |
| 76031 | | Chlorothiazide | "Chlorothiazide 250mg/5ml oral solution (Special Order)" |
| 77162 | | Chlorothiazide | "Chlorothiazide 12.5mg/5ml oral solution" |
| 77850 | | Chlorothiazide | "Chlorothiazide 500mg/5ml oral solution" |
| 2493 | 33911000001104 | Bumetanide/ Amiloride | Burinex A 5mg/ 1mg tablets (LEO Pharma) |
| 2495 | 189885001000027104 | Bumetanide/ Amiloride | Bumetanide with Amiloride tablets |
| 14587 | 318097001 | Bumetanide/ Amiloride | Amiloride 5mg / Bumetanide 1mg tablets |
| 814 | 318021009 | Bumetanide | Bumetanide 1mg tablets |
| 2788 | 846411000001100 | Bumetanide | Burinex 1mg tablets (LEO Pharma) |
| 5218 | 318023007 | Bumetanide | Bumetanide 1mg/ 5ml oral solution sugar free |
| 7806 | 318022002 | Bumetanide | Bumetanide 5mg tablets |
| 12226 | 37875001000027101 | Bumetanide | Burinex 1mg/ 5ml Oral solution (LEO Pharma) |
| 12294 | 521811000001107 | Bumetanide | Burinex 5mg tablets (LEO Pharma) |
| 15341 | 35125001000027107 | Bumetanide | Burinex 0.5mg/ ml Injection (LEO Pharma) |
| 19300 | 35912911000001108 | Bumetanide | Bumetanide 2mg/ 4ml solution for injection ampoules |
| 30913 | 232615001000027108 | Bumetanide | Betinex 1mg Tablet (Berk Pharmaceuticals Ltd) |
| 31932 | 689011000001100 | Bumetanide | Bumetanide 1mg tablets (C P Pharmaceuticals Ltd) |
| 32091 | 43411000001106 | Bumetanide | Bumetanide 1mg tablets (A A H Pharmaceuticals Ltd) |

| | | | |
|---|---|---|---|
| 34613 | 102011000001107 | Bumetanide | Bumetanide 5mg tablets (Teva UK Ltd) |
| 34934 | 402111000001109 | Bumetanide | Bumetanide 1mg tablets (Mylan) |
| 36767 | 226011000001101 | Bumetanide | Bumetanide 1mg tablets (IVAX Pharmaceuticals UK Ltd) |
| 39602 | 539611000001107 | Bumetanide | Bumetanide 1mg tablets (Actavis UK Ltd) |
| 45305 | 274211000001100 | Bumetanide | Bumetanide 1mg tablets (Teva UK Ltd) |
| 55548 | 731811000001106 | Bumetanide | Bumetanide 1mg tablets (Alliance Healthcare (Distribution) Ltd) |
| 62024 | 376211000001101 | Bumetanide | Bumetanide 5mg tablets (A A H Pharmaceuticals Ltd) |
| 63555 | 17810611000001109 | Bumetanide | Bumetanide 1mg tablets (Phoenix Healthcare Distribution Ltd) |
| 66195 | 9793511000001108 | Bumetanide | Bumetanide 1mg tablets (Almus Pharmaceuticals Ltd) |
| 73152 | 21804811000001102 | Bumetanide | Bumetanide 1mg tablets (Waymade Healthcare Plc) |
| 73195 | 22622811000001107 | Bumetanide | Bumetanide 1mg tablets (DE Pharmaceuticals) |
| 74814 | 14106511000001105 | Bumetanide | Bumetanide 1mg tablets (Niche Generics Ltd) |
| 30272 | 122075001000027106 | Benzthiazide/ Triamterene | Benthiazide with Triamterene capsules |
| 2 | 317919004 | Bendroflumethiazide | "Bendroflumethiazide 2.5mg tablets" |
| 58 | 317920005 | Bendroflumethiazide | "Bendroflumethiazide 5mg tablets" |
| 1209 | 817911000001100 | Bendroflumethiazide | "Neo-Naclex 5mg tablets (Mercury Pharma Group Ltd)" |
| 7351 | 8306811000001101 | Bendroflumethiazide | "Bendroflumethiazide 2.5mg/5ml oral suspension" |
| 7698 | 120911000001103 | Bendroflumethiazide | "Aprinox 5mg tablets (Amdipharm Plc)" |
| 8526 | 672111000001100 | Bendroflumethiazide | "Aprinox 2.5mg tablets (AMCo)" |
| 18973 | 35075001000027101 | Bendroflumethiazide | "Centyl 2.5mg Tablet (Edwin Burgess Ltd)" |
| 21803 | 3215001000027107 | Bendroflumethiazide | "Berkozide 2.5mg Tablet (Berk Pharmaceuticals Ltd)" |
| 21867 | 3225001000027103 | Bendroflumethiazide | "Berkozide 5mg Tablet (Berk Pharmaceuticals Ltd)" |
| 23427 | 501911000001107 | Bendroflumethiazide | "Bendroflumethiazide 5mg tablets (A A H Pharmaceuticals Ltd)" |
| 24189 | 197885001000027105 | Bendroflumethiazide | "Neo-bendromax 2.5mg Tablet (Ashbourne Pharmaceuticals Ltd)" |
| 24190 | 197895001000027109 | Bendroflumethiazide | "Neo-bendromax 5mg Tablet (Ashbourne Pharmaceuticals Ltd)" |
| 27256 | 454811000001100 | Bendroflumethiazide | "Bendroflumethiazide 2.5mg tablets (Wockhardt UK Ltd)" |
| 27689 | 464311000001103 | Bendroflumethiazide | "Bendroflumethiazide 2.5mg tablets (IVAX Pharmaceuticals UK Ltd)" |
| 29991 | 35085001000027106 | Bendroflumethiazide | "Centyl 5mg Tablet (Edwin Burgess Ltd)" |
| 31670 | 779811000001105 | Bendroflumethiazide | "Bendroflumethiazide 2.5mg tablets (Teva UK Ltd)" |
| 31820 | 159711000001108 | Bendroflumethiazide | "Bendroflumethiazide 5mg tablets (Wockhardt UK Ltd)" |
| 33415 | 914811000001108 | Bendroflumethiazide | "Bendroflumethiazide 2.5mg tablets (Mylan)" |
| 33651 | 772511000001103 | Bendroflumethiazide | "Bendroflumethiazide 2.5mg tablets (A A H Pharmaceuticals Ltd)" |
| 34059 | 1711000001109 | Bendroflumethiazide | "Bendroflumethiazide 2.5mg tablets (Actavis UK Ltd)" |
| 34124 | 720911000001105 | Bendroflumethiazide | "Bendroflumethiazide 5mg tablets (Actavis UK Ltd)" |
| 34602 | 17195711000001104 | Bendroflumethiazide | "Bendroflumethiazide 2.5mg tablets (Sovereign Medical Ltd)" |
| 34803 | 113045001000027101 | Bendroflumethiazide | "Bendroflumethiazide 2.5mg Tablet (Regent Laboratories Ltd)" |
| 40149 | 320511000001104 | Bendroflumethiazide | "Bendroflumethiazide 5mg tablets (IVAX Pharmaceuticals UK Ltd)" |
| 40886 | 9792811000001103 | Bendroflumethiazide | "Bendroflumethiazide 2.5mg tablets (Almus Pharmaceuticals Ltd)" |
| 41517 | 436711000001100 | Bendroflumethiazide | "Bendroflumethiazide 5mg tablets (Teva UK Ltd)" |
| 46302 | 18149011000001101 | Bendroflumethiazide | "Neo-Naclex 2.5mg tablets (AMCo)" |
| 47844 | 750511000001102 | Bendroflumethiazide | "Bendroflumethiazide 2.5mg tablets (Kent Pharmaceuticals Ltd)" |
| 53812 | 244535001000027101 | Bendroflumethiazide | "Bendroflumethiazide oral solution" |
| 64907 | 9793111000001104 | Bendroflumethiazide | "Bendroflumethiazide 5mg tablets (Almus Pharmaceuticals Ltd)" |

| | | | |
|---|---|---|---|
| 66517 | 8307011000001105 | Bendroflumethiazide | "Bendroflumethiazide 1.25mg/5ml oral suspension" |
| 67737 | 11009711000001106 | Bendroflumethiazide | "Bendroflumethiazide 2.5mg tablets (Dr Reddy's Laboratories (UK) Ltd)" |
| 67738 | 11010111000001102 | Bendroflumethiazide | "Bendroflumethiazide 5mg tablets (Dr Reddy's Laboratories (UK) Ltd)" |
| 67780 | 22617011000001105 | Bendroflumethiazide | "Bendroflumethiazide 5mg tablets (DE Pharmaceuticals)" |
| 70989 | 20323211000001103 | Bendroflumethiazide | "Bendroflumethiazide 2.5mg tablets (Genesis Pharmaceuticals Ltd)" |
| 72042 | 288611000001101 | Bendroflumethiazide | "Bendroflumethiazide 2.5mg tablets (Alliance Healthcare (Distribution) Ltd)" |
| 72083 | 21782811000001109 | Bendroflumethiazide | "Bendroflumethiazide 5mg tablets (Waymade Healthcare Plc)" |
| 72914 | 6685001000027101 | Bendroflumethiazide | "Bendroflumethiazide 2.5mg Tablet (Celltech Pharma Europe Ltd)" |
| 77681 | | Bendroflumethiazide | "Bendroflumethiazide 5mg tablets (Mylan)" |
| 348 | 19555001000027107 | Amiloride / Hydrochlorothiazide | Moduretic Tablet (Bristol-Myers Squibb Pharmaceuticals Ltd) |
| 923 | 377566005 | Amiloride / Hydrochlorothiazide | Co-amilozide 5mg/ 50mg tablets |
| 2002 | 159475001000027102 | Amiloride / Hydrochlorothiazide | Amiloride 5mg / hydrochlorothiazide 50mg tablets |
| 3293 | 19565001000027103 | Amiloride / Hydrochlorothiazide | Moduretic Oral solution (Bristol-Myers Squibb Pharmaceuticals Ltd) |
| 3701 | 159485001000027107 | Amiloride / Hydrochlorothiazide | Amiloride 2.5mg / hydrochlorothiazide 25mg tablets |
| 4034 | 159495001000027106 | Amiloride / Hydrochlorothiazide | Amiloride 5mg / hydrochlorothiazide 50mg/ 5ml solution |
| 8058 | 116145001000027100 | Amiloride / Hydrochlorothiazide | Normetic Tablet (Abbott Laboratories Ltd) |
| 18361 | 712211000001108 | Amiloride / Hydrochlorothiazide | Amilmaxco 5mg/ 50mg tablets (Ashbourne Pharmaceuticals Ltd) |
| 18733 | 169595001000027101 | Amiloride / Hydrochlorothiazide | Co-amilozide 5mg with 50mg/ ml oral solution |
| 19890 | 116045001000027108 | Amiloride / Hydrochlorothiazide | Hydrochlorothiazide with amiloride 25mgwith2.5mg Tablet |
| 20066 | 636611000001107 | Amiloride / Hydrochlorothiazide | Amil-Co 5mg/ 50mg tablets (IVAX Pharmaceuticals UK Ltd) |
| 22923 | 116065001000027102 | Amiloride / Hydrochlorothiazide | Hydrochlorothiazide with amiloride 50mg with 5mg Tablet |
| 24008 | 181425001000027109 | Amiloride / Hydrochlorothiazide | Vasetic Tablet (Shire Pharmaceuticals Ltd) |
| 25500 | 152595001000027107 | Amiloride / Hydrochlorothiazide | Hypertane 50 Tablet (Schwarz Pharma Ltd) |
| 26219 | 211855001000027109 | Amiloride / Hydrochlorothiazide | Zida-co 5mg+50mg Tablet (Opus Pharmaceuticals Ltd) |
| 26220 | 151865001000027108 | Amiloride / Hydrochlorothiazide | Delvas Tablet (Berk Pharmaceuticals Ltd) |
| 31150 | 831011000001103 | Amiloride / Hydrochlorothiazide | Co-amilozide 5mg/ 50mg tablets (IVAX Pharmaceuticals UK Ltd) |
| 41556 | 26611000001101 | Amiloride / Hydrochlorothiazide | Co-amilozide 5mg/ 50mg tablets (Teva UK Ltd) |
| 42142 | 453811000001103 | Amiloride / Hydrochlorothiazide | Moduretic 5mg/ 50mg tablets (Merck Sharp & Dohme Ltd) |
| 46916 | 17611000001107 | Amiloride / Hydrochlorothiazide | Co-amilozide 5mg/ 50mg tablets (A A H Pharmaceuticals Ltd) |
| 62249 | 552711000001105 | Amiloride / Hydrochlorothiazide | Co-amilozide 5mg/ 50mg tablets (Alliance Healthcare (Distribution) Ltd) |
| 62700 | 17885211000001100 | Amiloride / Hydrochlorothiazide | Co-amilozide 5mg/ 50mg tablets (Phoenix Healthcare Distribution Ltd) |
| 73337 | 366811000001105 | Amiloride / Hydrochlorothiazide | Co-amilozide 5mg/ 50mg tablets (Wockhardt UK Ltd) |
| 74017 | 5392111000001103 | Amiloride / Hydrochlorothiazide | Moduretic 5mg/ 50mg tablets (Waymade Healthcare Plc) |
| 75069 | 30015911000001100 | Amiloride / Hydrochlorothiazide | Co-amilozide 2.5mg/ 25mg tablets (Mawdsley-Brooks & Company Ltd) |
| 56 | 318136009 | Amiloride / Furosemide | Co-amilofruse 5mg/ 40mg tablets |
| 193 | 318135008 | Amiloride / Furosemide | Co-amilofruse 2.5mg/ 20mg tablets |
| 211 | 40605001000027105 | Amiloride / Furosemide | Frumil 40mg+5mg Tablet (Helios Healthcare Ltd) |
| 1301 | 40615001000027107 | Amiloride / Furosemide | Frumil ls 20mg+2.5mg Tablet (Helios Healthcare Ltd) |
| 2772 | 678511000001106 | Amiloride / Furosemide | Lasoride 5mg/ 40mg tablets (Sanofi) |
| 3793 | 318137000 | Amiloride / Furosemide | Co-amilofruse 10mg/ 80mg tablets |
| 4873 | 818511000001106 | Amiloride / Furosemide | Fru-Co 5mg/ 40mg tablets (Teva UK Ltd) |
| 9431 | 222555001000027108 | Amiloride / Furosemide | Frusemek 40mg+5mg Tablet (Approved Prescription Services Ltd) |
| 13435 | 82611000001106 | Amiloride / Furosemide | Frumil Forte 10mg/ 80mg tablets (Sanofi) |

| | | | |
|---|---|---|---|
| 18332 | 196835001000027101 | Amiloride / Furosemide | Aridil 20mg+2.5mg Tablet (C P Pharmaceuticals Ltd) |
| 21938 | 331311000001103 | Amiloride / Furosemide | Froop Co 5mg/ 40mg tablets (Ashbourne Pharmaceuticals Ltd) |
| 25965 | 213911000001104 | Amiloride / Furosemide | Co-amilofruse 2.5mg/ 20mg tablets (Wockhardt UK Ltd) |
| 28129 | 891211000001100 | Amiloride / Furosemide | Co-amilofruse 5mg/ 40mg tablets (Teva UK Ltd) |
| 30773 | 47575001000027101 | Amiloride / Furosemide | Co-amilofruse 5mg+40mg Tablet (Berk Pharmaceuticals Ltd) |
| 31773 | 380811000001106 | Amiloride / Furosemide | Co-amilofruse 5mg/ 40mg tablets (Wockhardt UK Ltd) |
| 33527 | 428711000001104 | Amiloride / Furosemide | Co-amilofruse 5mg/ 40mg tablets (Mylan) |
| 33658 | 11311000001108 | Amiloride / Furosemide | Co-amilofruse 5mg/ 40mg tablets (A A H Pharmaceuticals Ltd) |
| 34280 | 901811000001108 | Amiloride / Furosemide | Co-amilofruse 2.5mg/ 20mg tablets (Sandoz Ltd) |
| 34622 | 457611000001105 | Amiloride / Furosemide | Co-amilofruse 10mg/ 80mg tablets (Wockhardt UK Ltd) |
| 38901 | 550711000001106 | Amiloride / Furosemide | Frumil LS 20mg/ 2.5mg tablets (Sanofi) |
| 39807 | 427411000001106 | Amiloride / Furosemide | Frumil 40mg/ 5mg tablets (Sanofi) |
| 41533 | 242811000001109 | Amiloride / Furosemide | Co-amilofruse 2.5mg/ 20mg tablets (Teva UK Ltd) |
| 41719 | 667111000001108 | Amiloride / Furosemide | Co-amilofruse 5mg/ 40mg tablets (Actavis UK Ltd) |
| 43508 | 295411000001105 | Amiloride / Furosemide | Co-amilofruse 5mg/ 40mg tablets (Sandoz Ltd) |
| 57908 | 571911000001103 | Amiloride / Furosemide | Co-amilofruse 5mg/ 40mg tablets (Kent Pharmaceuticals Ltd) |
| 59412 | 21939011000001108 | Amiloride / Furosemide | Co-amilofruse 5mg/ 40mg tablets (Waymade Healthcare Plc) |
| 60258 | 19189511000001103 | Amiloride / Furosemide | Co-amilofruse 2.5mg/ 20mg tablets (Milpharm Ltd) |
| 71348 | 5372111000001107 | Amiloride / Furosemide | Frumil 40mg/ 5mg tablets (Waymade Healthcare Plc) |
| 71377 | 10855211000001104 | Amiloride / Furosemide | Frumil LS 20mg/ 2.5mg tablets (Waymade Healthcare Plc) |
| 73993 | 14244511000001107 | Amiloride / Furosemide | Frumil 40mg/ 5mg tablets (Sigma Pharmaceuticals Plc) |
| 74800 | 16182411000001100 | Amiloride / Furosemide | Frumil 40mg/ 5mg tablets (Lexon (UK) Ltd) |
| 2255 | 535711000001100 | Amiloride / Cyclopenthiazide | Navispare 2.5mg/ 250microgram tablets (AMCo) |
| 5727 | 318096005 | Amiloride / Cyclopenthiazide | Amiloride 2.5mg / Cyclopenthiazide 250microgram tablets |
| 1060 | 318052005 | Amiloride | Amiloride 5mg tablets |
| 9935 | 35900111000001108 | Amiloride | Amiloride 5mg/ 5ml oral solution sugar free |
| 13352 | 18905001000027107 | Amiloride | Midamor 5mg Tablet (MSD Thomas Morson Pharmaceuticals) |
| 24893 | 181285001000027104 | Amiloride | Amilospare Tablet (Ashbourne Pharmaceuticals Ltd) |
| 26217 | 156675001000027106 | Amiloride | Berkamil 5mg Tablet (Berk Pharmaceuticals Ltd) |
| 31375 | 799711000001108 | Amiloride | Amilamont 5mg/ 5ml oral solution sugar free (Rosemont Pharmaceuticals Ltd) |
| 33837 | 382611000001108 | Amiloride | Amiloride 5mg tablets (A A H Pharmaceuticals Ltd) |
| 34324 | 446011000001103 | Amiloride | Amiloride 5mg tablets (Teva UK Ltd) |
| 34750 | 754911000001104 | Amiloride | Amiloride 5mg tablets (Actavis UK Ltd) |
| 41630 | 3955001000027101 | Amiloride | Amiloride 5mg Tablet (IVAX Pharmaceuticals UK Ltd) |
| 43523 | 505611000001103 | Amiloride | Amiloride 5mg tablets (Mylan) |
| 44254 | 86595001000027100 | Amiloride | Amiloride 5.67mg tablets |
| 46930 | 533611000001105 | Amiloride | Amiloride 5mg tablets (Wockhardt UK Ltd) |
| 60149 | 8274111000001104 | Amiloride | Amiloride 5mg/ 5ml oral suspension |
| 76165 | | Amiloride | Amiloride 5mg tablets (Accord Healthcare Ltd) |
| 36519 | 10595001000027100 | Potassium Chloride/ Hydrochlorothiazide | "Esidrex -k Tablet (Novartis Pharmaceuticals UK Ltd)" |
| 1125 | 20765001000027100 | Potassium Chloride/ Cyclopenthiazide | "Navidrex -k Tablet (Novartis Pharmaceuticals UK Ltd)" |

| | | | |
|---|---|---|---|
| 2833 | 104585001000027108 | Potassium Chloride/ Cyclopenthiazide | "CYCLOPENTHIAZIDE -K tablets" |
| 1211 | 35910111000001106 | Potassium chloride/ Bendroflumethiazide | "Bendroflumethiazide 2.5mg / Potassium chloride 630mg (potassium 8.4mmol) modified-release tablets" |
| 1213 | 3638211000001100 | Potassium chloride/ Bendroflumethiazide | "Neo-Naclex-K modified-release tablets (Mercury Pharma Group Ltd)" |
| 2979 | 4955001000027104 | Potassium chloride/ Bendroflumethiazide | "Centyl k Tablet (Edwin Burgess Ltd)" |
| 17561 | 35910011000001105 | Bendroflumethiazide/ Potassium chloride | "Bendroflumethiazide 2.5mg / Potassium chloride 573mg (potassium 7.7mmol) modified-release tablets" |
| 20426 | 4965001000027108 | Bendroflumethiazide/ Potassium chloride | "Centyl k 2.5mg+7.7mmol Tablet (Edwin Burgess Ltd)" |
| 20431 | 3932711000001105 | Bendroflumethiazide/ Potassium chloride | "Centyl K modified-release tablets (LEO Pharma)" |
| 2681 | 185595001000027107 | | AMILORIDE 10 MG TAB |
| 3285 | 96205001000027106 | | AMILORIDE S/ F 5 MG/ 5ML SOL |
| 3962 | 187005001000027106 | | TRIAMTERENE 50MG HYDROCHLOROTHIAZIDE25MG TAB |
| 10796 | 188965001000027106 | | CHLORTHALIDONE 25MG/ POTASSIUM6.7MMOL S/ R MG TAB |
| 15053 | 96925001000027103 | | SPIRONOLACTONE 10 MG/ 5ML LIQ |
| 15602 | 167105001000027101 | | NATRILIX 5 MG TAB |
| 17721 | 158125001000027104 | | ALDACTIDE 100 MG TAB |
| 19611 | | | AMILORIDE 5MG/ HYDROCHLORTHIAZIDE 50MG |
| 19683 | | | BURINEX K |
| 19695 | | | AMILORIDE |
| 19721 | | | AMILORIDE 5MG/ HYDROCHLORTHIAZIDE 50MG |
| 20160 | 172315001000027106 | | CHLORTHALIDONE 500 MG TAB |
| 20513 | 167925001000027106 | | LASIX 10 MG INJ |
| 20779 | | | MODURETIC |
| 21848 | 4605001000027106 | | AMILOSPARE 5 MG TAB |
| 22539 | | | LASIX (2ML) |
| 23256 | | | LASIX PAED |
| 25630 | | | BRINALDIX K EFFERVESCENT |
| 26328 | | | LASIX (25ML) |
| 26675 | | | XIPAMIDE |
| 27555 | | | BURINEX |
| 29242 | 179705001000027102 | | CLOREXOLONE 10 MG TAB |
| 30368 | 172785001000027107 | | CHLOROTHIAZIDE/ SPIRONOLACTONE SACHETS 100 MG |
| 41889 | | | TRIAMTERENE 50MG HYDROCHLOROTHIAZIDE25MG |
| 13472 | 149755001000027102 | | "ESIDREX-K TAB" |
| 22242 | | | NAVIDREX |
| 22525 | 154075001000027109 | | "CHLOROTHIAZIDE 250 MG SYR" |
| 23492 | | | "CYCLOPENTHIAZIDE 250MCG/K 8.1MMOL" |
| 26120 | 154245001000027103 | | "CHLOROTHIAZIDE 50 MG SUS" |

| | | | |
|---|---|---|---|
| 27489 | 170715001000027104 | | "CHLOROTHIAZIDE 25 MG LIQ" |
| 31235 | 154235001000027104 | | "CHLOROTHIAZIDE SACHETS 60 MG" |
| **Renin inhibitors** | | | |
| 36629 | 425960005 | Aliskiren hemifumarate | Aliskiren 150mg tablets |
| 36878 | 11960911000001108 | Aliskiren hemifumarate | Rasilez 150mg tablets (Noden Pharma DAC) |
| 36879 | 11961711000001103 | Aliskiren hemifumarate | Rasilez 300mg tablets (Noden Pharma DAC) |
| 36909 | 425669009 | Aliskiren hemifumarate | Aliskiren 300mg tablets |
| **ORAL BISPHOSPHONATES** | | | |
| 9208 | 325979009 | Tiludronate disodium | Tiludronic acid 200mg tablets |
| 9525 | 4122511000001103 | Tiludronate disodium | Skelid 200mg tablets (Sanofi) |
| 3680 | 3848211000001108 | Sodium clodronate | Loron 400mg capsules (Roche Products Ltd) |
| 4868 | 325965001 | Sodium clodronate | Sodium clodronate 400mg capsules |
| 4927 | 3847911000001100 | Sodium clodronate | Bonefos 400mg capsules (Bayer Plc) |
| 5629 | 325972000 | Sodium clodronate | Sodium clodronate 800mg tablets |
| 6568 | 325970008 | Sodium clodronate | Sodium clodronate 520mg tablets |
| 9189 | 920611000001107 | Sodium clodronate | Bonefos 800mg tablets (Bayer Plc) |
| 11244 | 3813811000001107 | Sodium clodronate | Loron 520mg tablets (Intrapharm Laboratories Ltd) |
| 39043 | 11550411000001104 | Sodium clodronate | Clasteon 400mg capsules (Kent Pharmaceuticals Ltd) |
| 54989 | 20540511000001106 | Sodium clodronate | Clasteon 800mg tablets (Beacon Pharmaceuticals Ltd) |
| 45280 | 247875001000027107 | Risedronate Sodium/ Calcium Carbonate | Risedronate sodium 35mg & calcium carbonate 1250mg tablet |
| 6058 | 408027002 | Risedronate sodium | Risedronate sodium 35mg tablets |
| 6084 | 215955001000027104 | Risedronate sodium | Actonel once a week 35mg Tablet (Procter & Gamble (Health & Beauty Care) Ltd) |
| 6634 | 325983009 | Risedronate sodium | Risedronate sodium 5mg tablets |
| 7089 | 325984003 | Risedronate sodium | Risedronate sodium 30mg tablets |
| 7527 | 892511000001105 | Risedronate sodium | Actonel 5mg tablets (Warner Chilcott UK Ltd) |
| 7546 | 3778711000001100 | Risedronate sodium | Actonel 30mg tablets (Warner Chilcott UK Ltd) |
| 44511 | 4028511000001107 | Risedronate sodium | Actonel Once a Week 35mg tablets (Warner Chilcott UK Ltd) |
| 48013 | 18448311000001108 | Risedronate sodium | Risedronate sodium 35mg tablets (A A H Pharmaceuticals Ltd) |
| 52373 | 18626011000001106 | Risedronate sodium | Risedronate sodium 35mg tablets (Phoenix Healthcare Distribution Ltd) |
| 56431 | 18597911000001107 | Risedronate sodium | Risedronate sodium 35mg tablets (Actavis UK Ltd) |
| 56663 | 21887311000001101 | Risedronate sodium | Risedronate sodium 35mg tablets (Waymade Healthcare Plc) |
| 58618 | 16131211000001104 | Risedronate sodium | Risedronate sodium 35mg/ 5ml oral solution |
| 59449 | 19215711000001107 | Risedronate sodium | Risedronate sodium 35mg tablets (Bluefish Pharmaceuticals AB) |
| 59916 | 20889411000001109 | Risedronate sodium | Risedronate sodium 35mg tablets (Sandoz Ltd) |
| 60288 | 16255311000001105 | Risedronate sodium | Actonel Once a Week 35mg tablets (Mawdsley-Brooks & Company Ltd) |
| 61313 | 18448111000001106 | Risedronate sodium | Risedronate sodium 30mg tablets (A A H Pharmaceuticals Ltd) |

| | | | |
|---|---|---|---|
| 63802 | 19863711000001101 | Risedronate sodium | Actonel Once a Week 35mg tablets (Lexon (UK) Ltd) |
| 64431 | 18359911000001105 | Risedronate sodium | Risedronate sodium 30mg tablets (Aspire Pharma Ltd) |
| 65971 | 16131311000001107 | Risedronate sodium | Risedronate sodium 35mg/ 5ml oral suspension |
| 66028 | 13211011000001106 | Risedronate sodium | Actonel 35mg tablets (Teva UK Ltd) |
| 67078 | 18308311000001108 | Risedronate sodium | Risedronate sodium 35mg tablets (Teva UK Ltd) |
| 69630 | 20536311000001101 | Risedronate sodium | Risedronate sodium 35mg tablets (Almus Pharmaceuticals Ltd) |
| 69929 | 18344211000001102 | Risedronate sodium | Risedronate sodium 35mg tablets (Alliance Healthcare (Distribution) Ltd) |
| 69958 | 3361481000001104 | Risedronate sodium | Risedronate sodium 35mg tablets (Mylan) |
| 71209 | 30879111000001108 | Risedronate sodium | Risedronate sodium 35mg tablets (Mawdsley-Brooks & Company Ltd) |
| 73454 | 18359411000001102 | Risedronate sodium | Risedronate sodium 35mg tablets (Aspire Pharma Ltd) |
| 73989 | 13098211000001102 | Risedronate sodium | Actonel Once a Week 35mg tablets (Dowelhurst Ltd) |
| 74805 | 13823311000001109 | Risedronate sodium | Actonel Once a Week 35mg tablets (DE Pharmaceuticals) |
| 76178 | | Risedronate sodium | Actonel 5mg tablets (Sigma Pharmaceuticals Plc) |
| 76190 | | Risedronate sodium | Risedronate sodium 5mg tablets (A A H Pharmaceuticals Ltd) |
| 7112 | 9544911000001107 | Ibandronic sodium monohydrate | Bonviva 150mg tablets (Roche Products Ltd) |
| 7146 | 9553111000001105 | Ibandronic sodium monohydrate | Ibandronic acid 150mg tablets |
| 10193 | 410948006 | Ibandronic sodium monohydrate | Ibandronic acid 50mg tablets |
| 26913 | 7540111000001106 | Ibandronic sodium monohydrate | Bondronat 50mg tablets (Roche Products Ltd) |
| 47911 | 19371411000001104 | Ibandronic sodium monohydrate | Iasibon 50mg tablets (Aspire Pharma Ltd) |
| 51342 | 13837111000001101 | Ibandronic sodium monohydrate | Bonviva 150mg tablets (DE Pharmaceuticals) |
| 54453 | 19866411000001102 | Ibandronic sodium monohydrate | Bonviva 150mg tablets (Lexon (UK) Ltd) |
| 56030 | 20641211000001107 | Ibandronic sodium monohydrate | Ibandronic acid 150mg tablets (A A H Pharmaceuticals Ltd) |
| 56369 | 22086511000001105 | Ibandronic sodium monohydrate | Ibandronic acid 150mg tablets (Zentiva) |
| 57980 | 19295711000001108 | Ibandronic sodium monohydrate | Ibandronic acid 50mg tablets (Actavis UK Ltd) |
| 59587 | 23166211000001109 | Ibandronic sodium monohydrate | Ibandronic acid 150mg tablets (Ranbaxy (UK) Ltd) |
| 67159 | 20596011000001106 | Ibandronic sodium monohydrate | Ibandronic acid 50mg tablets (Teva UK Ltd) |
| 71000 | 20971311000001104 | Ibandronic sodium monohydrate | Ibandronic acid 150mg tablets (Alliance Healthcare (Distribution) Ltd) |
| 75425 | | Ibandronic sodium monohydrate | Ibandronic acid 50mg tablets (DE Pharmaceuticals) |
| 75644 | | Ibandronic sodium monohydrate | Quodixor 150mg tablets (Aspire Pharma Ltd) |
| 76545 | | Ibandronic sodium monohydrate | Bonviva 150mg tablets (Mawdsley-Brooks & Company Ltd) |
| 77225 | | Ibandronic sodium monohydrate | Ibandronic acid 150mg tablets (Teva UK Ltd) |
| 78002 | | Ibandronic sodium monohydrate | Ibandronic acid 150mg tablets (Mylan) |
| 766 | 3356811000001108 | Etidronate disodium | Didronel 200mg tablets (Warner Chilcott UK Ltd) |
| 4680 | 325951003 | Etidronate disodium | Etidronate disodium 200mg tablets |
| 63371 | 5196211000001102 | Etidronate disodium | Etidronate disodium 200mg tablets (Mylan) |
| 77070 | | Etidronate disodium | Etidronate disodium 100mg/ 5ml oral suspension |
| 77996 | | Etidronate disodium | Didronel 400mg tablets (Mawdsley-Brooks & Company Ltd) |
| 53169 | 224555001000027100 | Disodium Etidronate | Etidronate disodium 400mg Tablet |
| 54436 | 247235001000027103 | Disodium Etidronate | Etidronate disodium Oral solution |
| 37575 | 249915001000027103 | Colecalciferol/ Risedronate Sodium/ Calcium Carbonate | Risedronate sodium 35mg with calcium carbonate 2500mg & colecalciferol 22micrograms tablets and granules |
| 7224 | 9526611000001107 | Colecalciferol/ Alendronate sodium | Alendronic acid 70mg / Colecalciferol 70microgram tablets |
| 10227 | 9523811000001102 | Colecalciferol/ Alendronate sodium | Fosavance tablets (Merck Sharp & Dohme Ltd) |

| | | | |
|---|---|---|---|
| 66485 | 16182011000001109 | Colecalciferol/ Alendronate sodium | Fosavance tablets (Lexon (UK) Ltd) |
| 70927 | 34741911000001106 | Colecalciferol/ Alendronate sodium | Alendronic acid 70mg / Colecalciferol 70microgram tablets (Creo Pharma Ltd) |
| 76864 | | Colecalciferol/ Alendronate sodium | Alendronic acid 70mg / Colecalciferol 140microgram tablets |
| 45787 | 18683211000001101 | Alendronic acid | Alendronic acid 70mg/ 100ml oral solution unit dose sugar free |
| 52564 | 18680211000001106 | Alendronic acid | Alendronic acid 70mg/ 100ml oral solution unit dose sugar free (Rosemont Pharmaceuticals Ltd) |
| 55295 | 19185711000001106 | Alendronic acid | Alendronic acid 70mg/ 100ml oral solution unit dose sugar free (Alliance Healthcare (Distribution) Ltd) |
| 55998 | 20920711000001107 | Alendronic acid | Alendronic acid 70mg/ 75ml oral solution unit dose |
| 60144 | 21755811000001108 | Alendronic acid | Alendronic acid 70mg/ 100ml oral solution unit dose sugar free (Waymade Healthcare Plc) |
| 72208 | 20005911000001103 | Alendronic acid | Alendronic acid 70mg/ 100ml oral solution unit dose sugar free (A A H Pharmaceuticals Ltd) |
| 544 | 417211000001103 | Alendronate sodium | Fosamax Once Weekly 70mg tablets (Merck Sharp & Dohme Ltd) |
| 663 | 726311000001107 | Alendronate sodium | Fosamax 10mg tablets (Merck Sharp & Dohme Ltd) |
| 688 | 134599008 | Alendronate sodium | Alendronic acid 70mg tablets |
| 782 | 3145911000001101 | Alendronate sodium | Fosamax 5mg tablets (Merck Sharp & Dohme Ltd) |
| 2298 | 325974004 | Alendronate sodium | Alendronic acid 10mg tablets |
| 7530 | 325977006 | Alendronate sodium | Alendronic acid 5mg tablets |
| 35937 | 9221911000001101 | Alendronate sodium | Alendronic acid 70mg tablets (A A H Pharmaceuticals Ltd) |
| 37217 | 9251711000001102 | Alendronate sodium | Alendronic acid 10mg tablets (Teva UK Ltd) |
| 37218 | 9188811000001108 | Alendronate sodium | Alendronic acid 70mg tablets (Teva UK Ltd) |
| 40449 | 9836811000001109 | Alendronate sodium | Alendronic acid 70mg tablets (PLIVA Pharma Ltd) |
| 43958 | 9830711000001104 | Alendronate sodium | Alendronic acid 70mg tablets (Actavis UK Ltd) |
| 46245 | 9554311000001109 | Alendronate sodium | Alendronic acid 70mg tablets (Mylan) |
| 47380 | 10447611000001104 | Alendronate sodium | Alendronic acid 70mg tablets (Arrow Generics Ltd) |
| 50278 | 13441411000001109 | Alendronate sodium | Alendronic acid 70mg tablets (Wockhardt UK Ltd) |
| 50880 | 18264111000001108 | Alendronate sodium | Fosamax 10mg tablets (Necessity Supplies Ltd) |
| 51877 | 9299611000001102 | Alendronate sodium | Alendronic acid 70mg tablets (Alliance Healthcare (Distribution) Ltd) |
| 52284 | 14240211000001101 | Alendronate sodium | Fosamax 10mg tablets (Sigma Pharmaceuticals Plc) |
| 52624 | 17756411000001101 | Alendronate sodium | Alendronic acid 70mg tablets (Phoenix Healthcare Distribution Ltd) |
| 52834 | 18455911000001100 | Alendronate sodium | Alendronic acid 70mg tablets (Accord Healthcare Ltd) |
| 54566 | 9252411000001103 | Alendronate sodium | Alendronic acid 10mg tablets (A A H Pharmaceuticals Ltd) |
| 55965 | 9990311000001102 | Alendronate sodium | Alendronic acid 70mg tablets (Zentiva) |
| 56061 | 10435111000001104 | Alendronate sodium | Alendronic acid 10mg tablets (Actavis UK Ltd) |
| 56260 | 9208911000001102 | Alendronate sodium | Alendronic acid 70mg tablets (Kent Pharmaceuticals Ltd) |
| 56730 | 17963711000001100 | Alendronate sodium | Alendronic acid 70mg tablets (Almus Pharmaceuticals Ltd) |
| 57875 | 16181811000001107 | Alendronate sodium | Fosamax Once Weekly 70mg tablets (Lexon (UK) Ltd) |
| 58744 | 13880811000001102 | Alendronate sodium | Fosamax Once Weekly 70mg tablets (DE Pharmaceuticals) |
| 59079 | 17961911000001104 | Alendronate sodium | Alendronic acid 10mg tablets (Almus Pharmaceuticals Ltd) |
| 59247 | 18264311000001105 | Alendronate sodium | Fosamax Once Weekly 70mg tablets (Necessity Supplies Ltd) |
| 59485 | 18455711000001102 | Alendronate sodium | Alendronic acid 10mg tablets (Accord Healthcare Ltd) |
| 59555 | 9452511000001100 | Alendronate sodium | Alendronic acid 10mg tablets (Alliance Healthcare (Distribution) Ltd) |
| 61686 | 19701111000001102 | Alendronate sodium | Alendronic acid 70mg tablets (DE Pharmaceuticals) |
| 63008 | 24111211000001100 | Alendronate sodium | Alendronic acid 70mg tablets (Somex Pharma) |
| 63175 | 17756211000001100 | Alendronate sodium | Alendronic acid 10mg tablets (Phoenix Healthcare Distribution Ltd) |

| | | | |
|---|---|---|---|
| 64331 | 19700911000001106 | Alendronate sodium | Alendronic acid 10mg tablets (DE Pharmaceuticals) |
| 65008 | 30317811000001101 | Alendronate sodium | Alendronic acid 70mg effervescent tablets sugar free |
| 65905 | 9554111000001107 | Alendronate sodium | Alendronic acid 10mg tablets (Mylan) |
| 66203 | 30316711000001106 | Alendronate sodium | Binosto 70mg effervescent tablets (Internis Pharmaceuticals Ltd) |
| 69995 | 15631411000001101 | Alendronate sodium | Alendronic acid 35mg/ 5ml oral solution |
| 71851 | 15060811000001107 | Alendronate sodium | Alendronic acid 70mg tablets (Sigma Pharmaceuticals Plc) |
| 71963 | 15060411000001105 | Alendronate sodium | Alendronic acid 10mg tablets (Sigma Pharmaceuticals Plc) |
| 72541 | 15166911000001107 | Alendronate sodium | Alendronic acid 70mg/ 5ml oral solution |
| 73560 | 10688611000001108 | Alendronate sodium | Alendronic acid 10mg tablets (PLIVA Pharma Ltd) |
| 74859 | 5366911000001104 | Alendronate sodium | Fosamax Once Weekly 70mg tablets (Waymade Healthcare Plc) |
| 75094 | 21756211000001101 | Alendronate sodium | Alendronic acid 10mg tablets (Waymade Healthcare Plc) |
| 77297 | | Alendronate sodium | Alendronic acid 70mg tablets (Focus Pharmaceuticals Ltd) |
| 110 | 151675001000027109 | | DIDRONEL 100 MG TAB |
| 468 | 3352411000001105 | | Didronel PMO tablets (Warner Chilcott UK Ltd) |
| 3046 | 81085001000027103 | | DISODIUM ETIDRONATE 200 MG TAB |
| 11368 | 36133111000001109 | | Calcium carbonate 1.25g effervescent tablets and Disodium etidronate 400mg tablets |
| 37833 | 13208811000001109 | | Actonel Combi 35mg tablets and 1000mg/ 880unit effervescent granules sachets (Teva UK Ltd) |
| **STATINS** | | | |
| 66505 | 32234311000001109 | Simvastatin/ Fenofibrate | Fenofibrate 145mg / Simvastatin 40mg tablets |
| 66780 | 32234211000001101 | Simvastatin/ Fenofibrate | Fenofibrate 145mg / Simvastatin 20mg tablets |
| 69528 | 32170911000001100 | Simvastatin/ Fenofibrate | Cholib 145mg/ 20mg tablets (Mylan) |
| 70486 | 32169911000001103 | Simvastatin/ Fenofibrate | Cholib 145mg/ 40mg tablets (Mylan) |
| 7552 | 414177002 | Simvastatin/ Ezetimibe | Simvastatin 20mg / Ezetimibe 10mg tablets |
| 10172 | 414178007 | Simvastatin/ Ezetimibe | Simvastatin 40mg / Ezetimibe 10mg tablets |
| 10183 | 240715001000027105 | Simvastatin/ Ezetimibe | Simvastatin 40mg with ezetimibe 10mg tablet |
| 10206 | 240725001000027101 | Simvastatin/ Ezetimibe | Simvastatin 80mg with ezetimibe 10mg tablet |
| 11815 | 240705001000027108 | Simvastatin/ Ezetimibe | Simvastatin 20mg with ezetimibe 10mg tablet |
| 14219 | 414179004 | Simvastatin/ Ezetimibe | Simvastatin 80mg / Ezetimibe 10mg tablets |
| 16186 | 9310611000001103 | Simvastatin/ Ezetimibe | Inegy 10mg/ 80mg tablets (Merck Sharp & Dohme Ltd) |
| 17059 | 9310311000001108 | Simvastatin/ Ezetimibe | Inegy 10mg/ 40mg tablets (Merck Sharp & Dohme Ltd) |
| 21020 | 9309911000001100 | Simvastatin/ Ezetimibe | Inegy 10mg/ 20mg tablets (Merck Sharp & Dohme Ltd) |
| 25 | 319997009 | Simvastatin | Simvastatin 20mg tablets |
| 42 | 319996000 | Simvastatin | Simvastatin 10mg tablets |
| 51 | 320000009 | Simvastatin | Simvastatin 40mg tablets |
| 802 | 4896711000001108 | Simvastatin | Simvador 40mg tablets (Discovery Pharmaceuticals) |
| 818 | 242705001000027101 | Simvastatin | Simvastatin 20mg/ 5ml oral solution sugar free |
| 2718 | 108111000001106 | Simvastatin | Zocor 10mg tablets (Merck Sharp & Dohme Ltd) |
| 5148 | 320006003 | Simvastatin | Simvastatin 80mg tablets |
| 6168 | 859611000001107 | Simvastatin | Zocor 40mg tablets (Merck Sharp & Dohme Ltd) |
| 7196 | 776811000001104 | Simvastatin | Zocor 20mg tablets (Merck Sharp & Dohme Ltd) |
| 9920 | 4896511000001103 | Simvastatin | Simvador 20mg tablets (Discovery Pharmaceuticals) |
| 13041 | 4896211000001101 | Simvastatin | Simvador 10mg tablets (Discovery Pharmaceuticals) |

| | | | |
|---|---|---|---|
| 22579 | 113211000001106 | Simvastatin | Zocor 80mg tablets (Merck Sharp & Dohme Ltd) |
| 31930 | 238335001000027101 | Simvastatin | Zocor heart-pro 10mg Tablet (McNeil Products Ltd) |
| 32909 | 4579211000001101 | Simvastatin | Simvastatin 80mg tablets (A A H Pharmaceuticals Ltd) |
| 33082 | 4578811000001107 | Simvastatin | Simvastatin 20mg tablets (A A H Pharmaceuticals Ltd) |
| 34312 | 4464011000001108 | Simvastatin | Simvastatin 20mg tablets (Mylan) |
| 34316 | 4380911000001108 | Simvastatin | Simvastatin 20mg tablets (Teva UK Ltd) |
| 34353 | 4464211000001103 | Simvastatin | Simvastatin 40mg tablets (Mylan) |
| 34366 | 4466511000001106 | Simvastatin | Simvastatin 20mg tablets (IVAX Pharmaceuticals UK Ltd) |
| 34376 | 4381111000001104 | Simvastatin | Simvastatin 40mg tablets (Teva UK Ltd) |
| 34381 | 4466711000001101 | Simvastatin | Simvastatin 40mg tablets (IVAX Pharmaceuticals UK Ltd) |
| 34476 | 134095001000027108 | Simvastatin | Simvastatin 20mg Tablet (Ratiopharm UK Ltd) |
| 34481 | 4466211000001108 | Simvastatin | Simvastatin 10mg tablets (IVAX Pharmaceuticals UK Ltd) |
| 34502 | 4579011000001106 | Simvastatin | Simvastatin 40mg tablets (A A H Pharmaceuticals Ltd) |
| 34535 | 4463811000001100 | Simvastatin | Simvastatin 10mg tablets (Mylan) |
| 34545 | 134145001000027108 | Simvastatin | Simvastatin 40mg Tablet (Ratiopharm UK Ltd) |
| 34560 | 134055001000027102 | Simvastatin | Simvastatin 10mg Tablet (Ratiopharm UK Ltd) |
| 34746 | 136355001000027109 | Simvastatin | Simvastatin 20mg Tablet (Niche Generics Ltd) |
| 34814 | 4480511000001107 | Simvastatin | Simvastatin 20mg tablets (Wockhardt UK Ltd) |
| 34879 | 136385001000027102 | Simvastatin | Simvastatin 40mg Tablet (Niche Generics Ltd) |
| 34891 | 4467111000001104 | Simvastatin | Simvastatin 20mg tablets (Kent Pharmaceuticals Ltd) |
| 34907 | 4480711000001102 | Simvastatin | Simvastatin 40mg tablets (Wockhardt UK Ltd) |
| 34955 | 4578011000001101 | Simvastatin | Simvastatin 10mg tablets (A A H Pharmaceuticals Ltd) |
| 34969 | 4437211000001100 | Simvastatin | Simvastatin 40mg tablets (Actavis UK Ltd) |
| 37434 | 4580311000001109 | Simvastatin | Simvastatin 40mg tablets (Sandoz Ltd) |
| 39060 | 11551511000001107 | Simvastatin | Simvastatin 20mg tablets (Dexcel-Pharma Ltd) |
| 39652 | 256505001000027107 | Simvastatin | Simvastatin 40mg/ 5ml oral solution sugar free |
| 39675 | 183305001000027103 | Simvastatin | Simvastatin 20mg/ 5ml Oral suspension (Martindale Pharmaceuticals Ltd) |
| 39870 | 15158611000001106 | Simvastatin | Simvador 80mg tablets (Discovery Pharmaceuticals) |
| 40340 | 4380611000001102 | Simvastatin | Simvastatin 10mg tablets (Teva UK Ltd) |
| 40601 | 4465311000001105 | Simvastatin | Simvastatin 20mg tablets (Ranbaxy (UK) Ltd) |
| 41657 | 5476111000001102 | Simvastatin | Simvastatin 80mg tablets (Teva UK Ltd) |
| 44528 | 17305411000001100 | Simvastatin | Simvastatin 20mg/ 5ml oral suspension sugar free (Rosemont Pharmaceuticals Ltd) |
| 44650 | 11551711000001102 | Simvastatin | Simvastatin 40mg tablets (Dexcel-Pharma Ltd) |
| 44878 | 7630211000001106 | Simvastatin | Ranzolont 10mg tablets (Ranbaxy (UK) Ltd) |
| 45219 | 4467311000001102 | Simvastatin | Simvastatin 40mg tablets (Kent Pharmaceuticals Ltd) |
| 45235 | 4580111000001107 | Simvastatin | Simvastatin 20mg tablets (Sandoz Ltd) |
| 45245 | 4437011000001105 | Simvastatin | Simvastatin 20mg tablets (Actavis UK Ltd) |
| 45346 | 10414211000001101 | Simvastatin | Simvastatin 40mg tablets (Arrow Generics Ltd) |
| 46878 | 9804311000001100 | Simvastatin | Simvastatin 40mg tablets (Almus Pharmaceuticals Ltd) |
| 46956 | 10414411000001102 | Simvastatin | Simvastatin 80mg tablets (Arrow Generics Ltd) |
| 47774 | 10413811000001103 | Simvastatin | Simvastatin 10mg tablets (Arrow Generics Ltd) |
| 47948 | 13762911000001107 | Simvastatin | Simvastatin 10mg tablets (Tillomed Laboratories Ltd) |
| 48018 | 10414011000001106 | Simvastatin | Simvastatin 20mg tablets (Arrow Generics Ltd) |

| | | | |
|---|---|---|---|
| 48051 | 4466911000001104 | Simvastatin | Simvastatin 10mg tablets (Kent Pharmaceuticals Ltd) |
| 48058 | 4465011000001107 | Simvastatin | Simvastatin 10mg tablets (Ranbaxy (UK) Ltd) |
| 48078 | 4436811000001101 | Simvastatin | Simvastatin 10mg tablets (Actavis UK Ltd) |
| 48221 | 17369311000001105 | Simvastatin | Simvastatin 20mg/ 5ml oral suspension sugar free |
| 48431 | 17429811000001102 | Simvastatin | Simvastatin 40mg/ 5ml oral suspension sugar free |
| 48867 | 4574811000001104 | Simvastatin | Simvastatin 40mg tablets (Alliance Healthcare (Distribution) Ltd) |
| 49061 | 16067211000001108 | Simvastatin | Simvastatin 40mg tablets (Bristol Laboratories Ltd) |
| 49062 | 4574611000001103 | Simvastatin | Simvastatin 20mg tablets (Alliance Healthcare (Distribution) Ltd) |
| 49587 | 9804011000001103 | Simvastatin | Simvastatin 80mg tablets (Almus Pharmaceuticals Ltd) |
| 50483 | 10297511000001106 | Simvastatin | Simvastatin 40mg tablets (Relonchem Ltd) |
| 50564 | 10297311000001100 | Simvastatin | Simvastatin 20mg tablets (Relonchem Ltd) |
| 50670 | 19197811000001107 | Simvastatin | Simvastatin 40mg tablets (Milpharm Ltd) |
| 50703 | 18468511000001107 | Simvastatin | Simvastatin 40mg tablets (Accord Healthcare Ltd) |
| 50754 | 19733411000001104 | Simvastatin | Simvastatin 20mg tablets (Medreich Plc) |
| 50882 | 10618911000001100 | Simvastatin | Simvastatin 40mg tablets (Somex Pharma) |
| 51085 | 19733211000001103 | Simvastatin | Simvastatin 10mg tablets (Medreich Plc) |
| 51166 | 19733611000001101 | Simvastatin | Simvastatin 40mg tablets (Medreich Plc) |
| 51233 | 4574411000001101 | Simvastatin | Simvastatin 10mg tablets (Alliance Healthcare (Distribution) Ltd) |
| 51483 | 19197611000001108 | Simvastatin | Simvastatin 20mg tablets (Milpharm Ltd) |
| 51715 | 15188311000001107 | Simvastatin | Simvastatin 10mg tablets (Sigma Pharmaceuticals Plc) |
| 52098 | 4465611000001100 | Simvastatin | Simvastatin 40mg tablets (Ranbaxy (UK) Ltd) |
| 52257 | 18468011000001104 | Simvastatin | Simvastatin 20mg tablets (Accord Healthcare Ltd) |
| 52625 | 4480311000001101 | Simvastatin | Simvastatin 10mg tablets (Wockhardt UK Ltd) |
| 52676 | 8722011000001100 | Simvastatin | Simvastatin 10mg/ 5ml oral suspension |
| 52812 | 15171911000001107 | Simvastatin | Simvastatin 20mg tablets (Sigma Pharmaceuticals Plc) |
| 52953 | 16066911000001102 | Simvastatin | Simvastatin 20mg tablets (Bristol Laboratories Ltd) |
| 52962 | 19733811000001102 | Simvastatin | Simvastatin 80mg tablets (Medreich Plc) |
| 53087 | 10618711000001102 | Simvastatin | Simvastatin 20mg tablets (Somex Pharma) |
| 53340 | 16451611000001102 | Simvastatin | Zocor 40mg tablets (Lexon (UK) Ltd) |
| 53415 | 19197411000001105 | Simvastatin | Simvastatin 10mg tablets (Milpharm Ltd) |
| 53676 | 13763111000001103 | Simvastatin | Simvastatin 20mg tablets (Tillomed Laboratories Ltd) |
| 53822 | 16066711000001104 | Simvastatin | Simvastatin 10mg tablets (Bristol Laboratories Ltd) |
| 53908 | 11551311000001101 | Simvastatin | Simvastatin 10mg tablets (Dexcel-Pharma Ltd) |
| 53966 | 17793911000001109 | Simvastatin | Simvastatin 40mg tablets (Phoenix Healthcare Distribution Ltd) |
| 54240 | 15172411000001109 | Simvastatin | Simvastatin 40mg tablets (Sigma Pharmaceuticals Plc) |
| 54266 | 8722111000001104 | Simvastatin | Simvastatin 20mg/ 5ml oral suspension |
| 54493 | 10296911000001102 | Simvastatin | Simvastatin 10mg tablets (Relonchem Ltd) |
| 54606 | 18757511000001106 | Simvastatin | Simvastatin 20mg/ 5ml oral suspension sugar free (A A H Pharmaceuticals Ltd) |
| 54655 | 18467811000001106 | Simvastatin | Simvastatin 10mg tablets (Accord Healthcare Ltd) |
| 54819 | 17305711000001106 | Simvastatin | Simvastatin 40mg/ 5ml oral suspension sugar free (Rosemont Pharmaceuticals Ltd) |
| 54947 | 9804811000001109 | Simvastatin | Simvastatin 20mg tablets (Almus Pharmaceuticals Ltd) |
| 54976 | 10618511000001107 | Simvastatin | Simvastatin 10mg tablets (Somex Pharma) |
| 54985 | 13894411000001106 | Simvastatin | Simvastatin 40mg/ 5ml oral suspension |

| | | | |
|---|---|---|---|
| 55452 | 17785511000001104 | Simvastatin | Simvastatin 20mg tablets (Phoenix Healthcare Distribution Ltd) |
| 56065 | 21898011000001102 | Simvastatin | Simvastatin 20mg/ 5ml oral suspension sugar free (Waymade Healthcare Plc) |
| 56481 | 14732411000001109 | Simvastatin | Zocor 10mg tablets (Sigma Pharmaceuticals Plc) |
| 56494 | 14732811000001106 | Simvastatin | Zocor 20mg tablets (Sigma Pharmaceuticals Plc) |
| 57329 | 16091011000001105 | Simvastatin | Simvastatin 25mg/ 5ml oral suspension |
| 57568 | 16451411000001100 | Simvastatin | Zocor 10mg tablets (Lexon (UK) Ltd) |
| 58315 | 21899311000001105 | Simvastatin | Simvastatin 20mg tablets (Waymade Healthcare Plc) |
| 58755 | 17928611000001107 | Simvastatin | Simvastatin 10mg tablets (Phoenix Healthcare Distribution Ltd) |
| 61155 | 18758211000001107 | Simvastatin | Simvastatin 40mg/ 5ml oral suspension sugar free (A A H Pharmaceuticals Ltd) |
| 61321 | 4579911000001105 | Simvastatin | Simvastatin 10mg tablets (Sandoz Ltd) |
| 61360 | 9805011000001104 | Simvastatin | Simvastatin 10mg tablets (Almus Pharmaceuticals Ltd) |
| 61665 | 21899011000001107 | Simvastatin | Simvastatin 10mg tablets (Waymade Healthcare Plc) |
| 62137 | 21899611000001100 | Simvastatin | Simvastatin 40mg tablets (Waymade Healthcare Plc) |
| 64104 | 28988511000001107 | Simvastatin | Simvastatin 20mg tablets (Crescent Pharma Ltd) |
| 64180 | 28918311000001104 | Simvastatin | Simvastatin 10mg tablets (Crescent Pharma Ltd) |
| 64307 | 28918111000001101 | Simvastatin | Simvastatin 40mg tablets (Crescent Pharma Ltd) |
| 64968 | 30097311000001108 | Simvastatin | Simvastatin 10mg tablets (DE Pharmaceuticals) |
| 65181 | 30097711000001107 | Simvastatin | Simvastatin 40mg tablets (DE Pharmaceuticals) |
| 65679 | 30097511000001102 | Simvastatin | Simvastatin 20mg tablets (DE Pharmaceuticals) |
| 65901 | 10733711000001109 | Simvastatin | Simvastatin 40mg tablets (Zentiva) |
| 65925 | 17841911000001100 | Simvastatin | Simvastatin 20mg/ 5ml oral suspension sugar free (Alliance Healthcare (Distribution) Ltd) |
| 67098 | 32494611000001109 | Simvastatin | Simvastatin 10mg tablets (Brown & Burk UK Ltd) |
| 67745 | 10733011000001107 | Simvastatin | Simvastatin 10mg tablets (Zentiva) |
| 67773 | 10733511000001104 | Simvastatin | Simvastatin 20mg tablets (Zentiva) |
| 68563 | 32495011000001103 | Simvastatin | Simvastatin 40mg tablets (Brown & Burk UK Ltd) |
| 68686 | 33555611000001103 | Simvastatin | Simvastatin 20mg tablets (Genesis Pharmaceuticals Ltd) |
| 69413 | 32494811000001108 | Simvastatin | Simvastatin 20mg tablets (Brown & Burk UK Ltd) |
| 71773 | 32495311000001100 | Simvastatin | Simvastatin 80mg tablets (Brown & Burk UK Ltd) |
| 72050 | 33654911000001100 | Simvastatin | Simvastatin 10mg tablets (Genesis Pharmaceuticals Ltd) |
| 75134 | 4464511000001100 | Simvastatin | Simvastatin 80mg tablets (Mylan) |
| 76481 | | Simvastatin | Simvastatin 80mg Tablet (Dexcel-Pharma Ltd) |
| 76594 | | Simvastatin | Simvastatin 40mg tablets (Tillomed Laboratories Ltd) |
| 77357 | | Simvastatin | Zocor 20mg tablets (Waymade Healthcare Plc) |
| 77358 | | Simvastatin | Zocor 10mg tablets (Waymade Healthcare Plc) |
| 77470 | | Simvastatin | Zocor 10mg tablets (Necessity Supplies Ltd) |
| 77471 | | Simvastatin | Zocor 20mg tablets (Necessity Supplies Ltd) |
| 713 | 408036003 | Rosuvastatin calcium | Rosuvastatin 10mg tablets |
| 6213 | 408037007 | Rosuvastatin calcium | Rosuvastatin 20mg tablets |
| 7347 | 4171011000001104 | Rosuvastatin calcium | Crestor 10mg tablets (AstraZeneca UK Ltd) |
| 7554 | 409108001 | Rosuvastatin calcium | Rosuvastatin 5mg tablets |
| 9897 | 408024009 | Rosuvastatin calcium | Rosuvastatin 40mg tablets |
| 9930 | 4172111000001108 | Rosuvastatin calcium | Crestor 40mg tablets (AstraZeneca UK Ltd) |
| 15252 | 4171311000001101 | Rosuvastatin calcium | Crestor 20mg tablets (AstraZeneca UK Ltd) |

| | | | |
|---|---|---|---|
| 17688 | 9747511000001107 | Rosuvastatin calcium | Crestor 5mg tablets (AstraZeneca UK Ltd) |
| 53460 | 13857911000001102 | Rosuvastatin calcium | Crestor 10mg tablets (DE Pharmaceuticals) |
| 57763 | 10769311000001104 | Rosuvastatin calcium | Rosuvastatin 10mg tablets (Waymade Healthcare Plc) |
| 57999 | 16155011000001105 | Rosuvastatin calcium | Crestor 40mg tablets (Lexon (UK) Ltd) |
| 58617 | 16075311000001101 | Rosuvastatin calcium | Rosuvastatin 20mg/ 5ml oral suspension |
| 59447 | 10513311000001102 | Rosuvastatin calcium | Crestor 20mg tablets (Waymade Healthcare Plc) |
| 59452 | 12561111000001105 | Rosuvastatin calcium | Rosuvastatin 5mg tablets (Waymade Healthcare Plc) |
| 60160 | 18202311000001102 | Rosuvastatin calcium | Rosuvastatin 5mg tablets (Mawdsley-Brooks & Company Ltd) |
| 70308 | 14212311000001103 | Rosuvastatin calcium | Crestor 20mg tablets (Sigma Pharmaceuticals Plc) |
| 71014 | 11580111000001101 | Rosuvastatin calcium | Rosuvastatin 20mg tablets (Waymade Healthcare Plc) |
| 73025 | 35027811000001104 | Rosuvastatin calcium | Rosuvastatin 20mg tablets (Mylan) |
| 74552 | 35183311000001102 | Rosuvastatin calcium | Rosuvastatin 10mg tablets (Milpharm Ltd) |
| 75971 | | Rosuvastatin calcium | Rosuvastatin 10mg tablets (Sandoz Ltd) |
| 76120 | | Rosuvastatin calcium | Rosuvastatin 10mg tablets (Teva UK Ltd) |
| 490 | 320012008 | Pravastatin sodium | Pravastatin 10mg tablets |
| 730 | 320013003 | Pravastatin sodium | Pravastatin 20mg tablets |
| 1219 | 320014009 | Pravastatin sodium | Pravastatin 40mg tablets |
| 1221 | 802411000001108 | Pravastatin sodium | Lipostat 10mg tablets (Bristol-Myers Squibb Pharmaceuticals Ltd) |
| 1223 | 535011000001102 | Pravastatin sodium | Lipostat 40mg tablets (Bristol-Myers Squibb Pharmaceuticals Ltd) |
| 3690 | 454111000001107 | Pravastatin sodium | Lipostat 20mg tablets (Bristol-Myers Squibb Pharmaceuticals Ltd) |
| 32921 | 163075001000027105 | Pravastatin sodium | Pravastatin 10mg Tablet (Dr Reddy's Laboratories (UK) Ltd) |
| 34820 | 7977111000001100 | Pravastatin sodium | Pravastatin 40mg tablets (A A H Pharmaceuticals Ltd) |
| 36377 | 7943411000001102 | Pravastatin sodium | Pravastatin 20mg tablets (Teva UK Ltd) |
| 40382 | 7976911000001100 | Pravastatin sodium | Pravastatin 20mg tablets (A A H Pharmaceuticals Ltd) |
| 43218 | 7943211000001101 | Pravastatin sodium | Pravastatin 10mg tablets (Teva UK Ltd) |
| 47988 | 8113311000001107 | Pravastatin sodium | Pravastatin 40mg tablets (Mylan) |
| 48097 | 7943611000001104 | Pravastatin sodium | Pravastatin 40mg tablets (Teva UK Ltd) |
| 50925 | 15174911000001105 | Pravastatin sodium | Pravastatin 10mg tablets (Sigma Pharmaceuticals Plc) |
| 51676 | 19732111000001103 | Pravastatin sodium | Pravastatin 40mg tablets (Medreich Plc) |
| 51890 | 19731911000001106 | Pravastatin sodium | Pravastatin 20mg tablets (Medreich Plc) |
| 52755 | 8027311000001103 | Pravastatin sodium | Pravastatin 20mg tablets (Alliance Healthcare (Distribution) Ltd) |
| 54435 | 9807511000001103 | Pravastatin sodium | Pravastatin 40mg tablets (Almus Pharmaceuticals Ltd) |
| 54607 | 9806911000001107 | Pravastatin sodium | Pravastatin 20mg tablets (Almus Pharmaceuticals Ltd) |
| 55912 | 8027611000001108 | Pravastatin sodium | Pravastatin 40mg tablets (Alliance Healthcare (Distribution) Ltd) |
| 56146 | 21850611000001104 | Pravastatin sodium | Pravastatin 10mg tablets (Waymade Healthcare Plc) |
| 56607 | 21850811000001100 | Pravastatin sodium | Pravastatin 20mg tablets (Waymade Healthcare Plc) |
| 56735 | 8113111000001105 | Pravastatin sodium | Pravastatin 20mg tablets (Mylan) |
| 56893 | 18465611000001100 | Pravastatin sodium | Pravastatin 40mg tablets (Accord Healthcare Ltd) |
| 56916 | 11410811000001100 | Pravastatin sodium | Pravastatin 40mg tablets (PLIVA Pharma Ltd) |
| 57108 | 21851011000001102 | Pravastatin sodium | Pravastatin 40mg tablets (Waymade Healthcare Plc) |
| 57137 | 9806311000001106 | Pravastatin sodium | Pravastatin 10mg tablets (Almus Pharmaceuticals Ltd) |
| 57296 | 17915711000001103 | Pravastatin sodium | Pravastatin 20mg tablets (Phoenix Healthcare Distribution Ltd) |
| 57397 | 18465211000001102 | Pravastatin sodium | Pravastatin 10mg tablets (Accord Healthcare Ltd) |

| | | | |
|---|---|---|---|
| 59508 | 18465411000001103 | Pravastatin sodium | Pravastatin 20mg tablets (Accord Healthcare Ltd) |
| 60251 | 7997711000001109 | Pravastatin sodium | Pravastatin 10mg tablets (Sandoz Ltd) |
| 61134 | 15175111000001106 | Pravastatin sodium | Pravastatin 20mg tablets (Sigma Pharmaceuticals Plc) |
| 62979 | 8099711000001100 | Pravastatin sodium | Pravastatin 40mg tablets (Kent Pharmaceuticals Ltd) |
| 63074 | 11410611000001104 | Pravastatin sodium | Pravastatin 20mg tablets (PLIVA Pharma Ltd) |
| 63787 | 13761111000001104 | Pravastatin sodium | Pravastatin 10mg tablets (Tillomed Laboratories Ltd) |
| 67829 | 7998011000001108 | Pravastatin sodium | Pravastatin 20mg tablets (Sandoz Ltd) |
| 68156 | 7976411000001108 | Pravastatin sodium | Pravastatin 10mg tablets (A A H Pharmaceuticals Ltd) |
| 71015 | 19731611000001100 | Pravastatin sodium | Pravastatin 10mg tablets (Medreich Plc) |
| 72048 | 7959111000001108 | Pravastatin sodium | Pravastatin 40mg tablets (Actavis UK Ltd) |
| 72149 | 14957811000001102 | Pravastatin sodium | Pravastatin 5mg/ 5ml oral suspension |
| 75826 | | Pravastatin sodium | Pravastatin 20mg tablets (Actavis UK Ltd) |
| 77394 | | Pravastatin sodium | Lipostat 20mg tablets (Dowelhurst Ltd) |
| 379 | 320022002 | Fluvastatin sodium | Fluvastatin 20mg capsules |
| 2137 | 320023007 | Fluvastatin sodium | Fluvastatin 40mg capsules |
| 5985 | 378111000001106 | Fluvastatin sodium | Lescol XL 80mg tablets (Novartis Pharmaceuticals UK Ltd) |
| 8380 | 84811000001104 | Fluvastatin sodium | Lescol 20mg capsules (Novartis Pharmaceuticals UK Ltd) |
| 9153 | 409611000001108 | Fluvastatin sodium | Lescol 40mg capsules (Novartis Pharmaceuticals UK Ltd) |
| 11627 | 36566411000001105 | Fluvastatin sodium | Fluvastatin 80mg modified-release tablets |
| 53770 | 14584511000001106 | Fluvastatin sodium | Fluvastatin 40mg capsules (A A H Pharmaceuticals Ltd) |
| 59278 | 14036511000001107 | Fluvastatin sodium | Fluvastatin 20mg capsules (Zentiva) |
| 62148 | 16237911000001102 | Fluvastatin sodium | Fluvastatin 20mg capsules (Actavis UK Ltd) |
| 67328 | 16499511000001107 | Fluvastatin sodium | Lescol XL 80mg tablets (Mawdsley-Brooks & Company Ltd) |
| 71029 | 20289511000001104 | Fluvastatin sodium | Fluvastatin 40mg capsules (Sandoz Ltd) |
| 72308 | 14037111000001100 | Fluvastatin sodium | Fluvastatin 20mg capsules (Alliance Healthcare (Distribution) Ltd) |
| 73383 | 16238111000001104 | Fluvastatin sodium | Fluvastatin 40mg capsules (Actavis UK Ltd) |
| 74085 | 14765111000001100 | Fluvastatin sodium | Lescol 40mg capsules (Sigma Pharmaceuticals Plc) |
| 77306 | | Fluvastatin sodium | Nandovar XL 80mg tablets (Sandoz Ltd) |
| 77425 | | Fluvastatin sodium | Lescol 20mg capsules (Sigma Pharmaceuticals Plc) |
| 77472 | | Fluvastatin sodium | Lescol 40mg capsules (Lexon (UK) Ltd) |
| 420 | 320035006 | Cerivastatin sodium | Cerivastatin 100microgram tablets |
| 4961 | 226245001000027101 | Cerivastatin sodium | Lipobay 300microgram Tablet (Bayer Plc) |
| 5009 | 320036007 | Cerivastatin sodium | Cerivastatin 200microgram tablets |
| 5251 | 320037003 | Cerivastatin sodium | Cerivastatin 300microgram tablets |
| 5278 | 320041004 | Cerivastatin sodium | Cerivastatin 400microgram tablets |
| 9315 | 226225001000027104 | Cerivastatin sodium | Lipobay 100microgram Tablet (Bayer Plc) |
| 9316 | 226235001000027102 | Cerivastatin sodium | Lipobay 200microgram Tablet (Bayer Plc) |
| 18442 | 201215001000027108 | Cerivastatin sodium | Lipobay 400microgram Tablet (Bayer Plc) |
| 31658 | 134491009 | Cerivastatin sodium | Cerivastatin 800microgram tablets |
| 53813 | 4535911000001106 | Cerivastatin sodium | Lipobay 100microgram tablets (Bayer Plc) |
| 55207 | 4537511000001108 | Cerivastatin sodium | Lipobay 200microgram tablets (Bayer Plc) |
| 58480 | 4566311000001105 | Cerivastatin sodium | Lipobay 300microgram tablets (Bayer Plc) |
| 62132 | 4538111000001103 | Cerivastatin sodium | Lipobay 400microgram tablets (Bayer Plc) |

| | | | |
|---|---|---|---|
| 28 | 320029006 | Atorvastatin calcium trihydrate | Atorvastatin 10mg tablets |
| 75 | 320030001 | Atorvastatin calcium trihydrate | Atorvastatin 20mg tablets |
| 745 | 320031002 | Atorvastatin calcium trihydrate | Atorvastatin 40mg tablets |
| 2955 | 484211000001108 | Atorvastatin calcium trihydrate | Lipitor 40mg tablets (Pfizer Ltd) |
| 3411 | 643911000001108 | Atorvastatin calcium trihydrate | Lipitor 10mg tablets (Pfizer Ltd) |
| 5775 | 134489001 | Atorvastatin calcium trihydrate | Atorvastatin 80mg tablets |
| 7374 | 232011000001102 | Atorvastatin calcium trihydrate | Lipitor 20mg tablets (Pfizer Ltd) |
| 17683 | 756111000001109 | Atorvastatin calcium trihydrate | Lipitor 80mg tablets (Pfizer Ltd) |
| 47065 | 19722511000001105 | Atorvastatin calcium trihydrate | Atorvastatin 20mg chewable tablets sugar free |
| 47090 | 19722411000001106 | Atorvastatin calcium trihydrate | Atorvastatin 10mg chewable tablets sugar free |
| 47630 | 19719611000001109 | Atorvastatin calcium trihydrate | Lipitor 20mg chewable tablets (Pfizer Ltd) |
| 47721 | 19719311000001104 | Atorvastatin calcium trihydrate | Lipitor 10mg chewable tablets (Pfizer Ltd) |
| 48346 | 20528611000001105 | Atorvastatin calcium trihydrate | Atorvastatin 60mg tablets |
| 48518 | 14158611000001100 | Atorvastatin calcium trihydrate | Atorvastatin 10mg/ 5ml oral solution |
| 48973 | 20528511000001106 | Atorvastatin calcium trihydrate | Atorvastatin 30mg tablets |
| 49558 | 20491911000001107 | Atorvastatin calcium trihydrate | Atorvastatin 20mg tablets (A A H Pharmaceuticals Ltd) |
| 49751 | 20508311000001106 | Atorvastatin calcium trihydrate | Atorvastatin 40mg tablets (Alliance Healthcare (Distribution) Ltd) |
| 50236 | 20576911000001102 | Atorvastatin calcium trihydrate | Atorvastatin 10mg tablets (Zentiva) |
| 50272 | 20978811000001102 | Atorvastatin calcium trihydrate | Atorvastatin 40mg tablets (Pfizer Ltd) |
| 50788 | 20978111000001109 | Atorvastatin calcium trihydrate | Atorvastatin 20mg tablets (Pfizer Ltd) |
| 50790 | 20448411000001102 | Atorvastatin calcium trihydrate | Atorvastatin 20mg tablets (Dexcel-Pharma Ltd) |
| 50963 | 20494711000001108 | Atorvastatin calcium trihydrate | Atorvastatin 40mg tablets (Teva UK Ltd) |
| 51134 | 20491711000001105 | Atorvastatin calcium trihydrate | Atorvastatin 10mg tablets (A A H Pharmaceuticals Ltd) |
| 51200 | 20570211000001109 | Atorvastatin calcium trihydrate | Atorvastatin 40mg tablets (Arrow Generics Ltd) |
| 51359 | 20569511000001105 | Atorvastatin calcium trihydrate | Atorvastatin 20mg tablets (Arrow Generics Ltd) |
| 51622 | 20529611000001101 | Atorvastatin calcium trihydrate | Atorvastatin 20mg tablets (Consilient Health Ltd) |
| 51876 | 20529811000001102 | Atorvastatin calcium trihydrate | Atorvastatin 40mg tablets (Consilient Health Ltd) |
| 52097 | 20573211000001104 | Atorvastatin calcium trihydrate | Atorvastatin 40mg tablets (Wockhardt UK Ltd) |
| 52168 | 20496611000001108 | Atorvastatin calcium trihydrate | Atorvastatin 20mg tablets (Aspire Pharma Ltd) |
| 52211 | 20482911000001102 | Atorvastatin calcium trihydrate | Atorvastatin 20mg tablets (Actavis UK Ltd) |
| 52397 | 20982611000001106 | Atorvastatin calcium trihydrate | Atorvastatin 40mg tablets (Dr Reddy's Laboratories (UK) Ltd) |
| 52398 | 20492311000001102 | Atorvastatin calcium trihydrate | Atorvastatin 40mg tablets (A A H Pharmaceuticals Ltd) |
| 52459 | 20483311000001108 | Atorvastatin calcium trihydrate | Atorvastatin 80mg tablets (Actavis UK Ltd) |
| 52460 | 20496911000001102 | Atorvastatin calcium trihydrate | Atorvastatin 40mg tablets (Aspire Pharma Ltd) |
| 52821 | 20982911000001100 | Atorvastatin calcium trihydrate | Atorvastatin 80mg tablets (Dr Reddy's Laboratories (UK) Ltd) |
| 53594 | 16507511000001107 | Atorvastatin calcium trihydrate | Lipitor 80mg tablets (Mawdsley-Brooks & Company Ltd) |
| 53772 | 20508511000001100 | Atorvastatin calcium trihydrate | Atorvastatin 80mg tablets (Alliance Healthcare (Distribution) Ltd) |
| 53887 | 20483111000001106 | Atorvastatin calcium trihydrate | Atorvastatin 40mg tablets (Actavis UK Ltd) |
| 53890 | 20979511000001106 | Atorvastatin calcium trihydrate | Atorvastatin 80mg tablets (Pfizer Ltd) |
| 54535 | 20977811000001101 | Atorvastatin calcium trihydrate | Atorvastatin 10mg tablets (Pfizer Ltd) |
| 54992 | 14158711000001109 | Atorvastatin calcium trihydrate | Atorvastatin 10mg/ 5ml oral suspension |
| 55032 | 20448211000001101 | Atorvastatin calcium trihydrate | Atorvastatin 10mg tablets (Dexcel-Pharma Ltd) |
| 55034 | 14158911000001106 | Atorvastatin calcium trihydrate | Atorvastatin 40mg/ 5ml oral suspension |

| | | | |
|---|---|---|---|
| 55444 | 20577411000001107 | Atorvastatin calcium trihydrate | Atorvastatin 40mg tablets (Zentiva) |
| 55727 | 20482711000001104 | Atorvastatin calcium trihydrate | Atorvastatin 10mg tablets (Actavis UK Ltd) |
| 56016 | 19719611000001109 | Atorvastatin calcium trihydrate | Lipitor 20mg chewable tablets (Pfizer Ltd) |
| 56097 | 19722411000001106 | Atorvastatin calcium trihydrate | Atorvastatin 10mg chewable tablets sugar free |
| 56165 | 19722511000001105 | Atorvastatin calcium trihydrate | Atorvastatin 20mg chewable tablets sugar free |
| 56182 | 20577611000001105 | Atorvastatin calcium trihydrate | Atorvastatin 80mg tablets (Zentiva) |
| 56248 | 14198211000001106 | Atorvastatin calcium trihydrate | Atorvastatin 20mg tablets (Sigma Pharmaceuticals Plc) |
| 56564 | 22047511000001102 | Atorvastatin calcium trihydrate | Atorvastatin 20mg tablets (Almus Pharmaceuticals Ltd) |
| 56841 | 20448611000001104 | Atorvastatin calcium trihydrate | Atorvastatin 40mg tablets (Dexcel-Pharma Ltd) |
| 57117 | 21779811000001108 | Atorvastatin calcium trihydrate | Atorvastatin 80mg tablets (Waymade Healthcare Plc) |
| 57348 | 20529411000001104 | Atorvastatin calcium trihydrate | Atorvastatin 10mg tablets (Consilient Health Ltd) |
| 57834 | 22613211000001109 | Atorvastatin calcium trihydrate | Atorvastatin 40mg tablets (DE Pharmaceuticals) |
| 57836 | 20495411000001101 | Atorvastatin calcium trihydrate | Atorvastatin 80mg tablets (Teva UK Ltd) |
| 58041 | 20494411000001102 | Atorvastatin calcium trihydrate | Atorvastatin 20mg tablets (Teva UK Ltd) |
| 58110 | 20577211000001108 | Atorvastatin calcium trihydrate | Atorvastatin 20mg tablets (Zentiva) |
| 58394 | 20508111000001109 | Atorvastatin calcium trihydrate | Atorvastatin 20mg tablets (Alliance Healthcare (Distribution) Ltd) |
| 58418 | 20492611000001107 | Atorvastatin calcium trihydrate | Atorvastatin 80mg tablets (A A H Pharmaceuticals Ltd) |
| 58742 | 20570911000001100 | Atorvastatin calcium trihydrate | Atorvastatin 80mg tablets (Arrow Generics Ltd) |
| 58834 | 13831111000001108 | Atorvastatin calcium trihydrate | Atorvastatin 10mg tablets (DE Pharmaceuticals) |
| 58868 | 14197911000001103 | Atorvastatin calcium trihydrate | Atorvastatin 10mg tablets (Sigma Pharmaceuticals Plc) |
| 59272 | 21779411000001106 | Atorvastatin calcium trihydrate | Atorvastatin 20mg tablets (Waymade Healthcare Plc) |
| 59331 | 13922611000001102 | Atorvastatin calcium trihydrate | Lipitor 10mg tablets (DE Pharmaceuticals) |
| 59357 | 22940511000001105 | Atorvastatin calcium trihydrate | Atorvastatin 10mg tablets (Ranbaxy (UK) Ltd) |
| 59446 | 22047711000001107 | Atorvastatin calcium trihydrate | Atorvastatin 40mg tablets (Almus Pharmaceuticals Ltd) |
| 59776 | 20497611000001105 | Atorvastatin calcium trihydrate | Atorvastatin 80mg tablets (Aspire Pharma Ltd) |
| 59859 | 20492811000001106 | Atorvastatin calcium trihydrate | Atorvastatin 10mg tablets (Teva UK Ltd) |
| 60464 | 14018311000001109 | Atorvastatin calcium trihydrate | Atorvastatin 20mg/ 5ml oral suspension |
| 60511 | 22941211000001101 | Atorvastatin calcium trihydrate | Atorvastatin 40mg tablets (Ranbaxy (UK) Ltd) |
| 60607 | 22613511000001107 | Atorvastatin calcium trihydrate | Atorvastatin 80mg tablets (DE Pharmaceuticals) |
| 60989 | 22202511000001100 | Atorvastatin calcium trihydrate | Atorvastatin 80mg tablets (Phoenix Healthcare Distribution Ltd) |
| 61149 | 21779211000001107 | Atorvastatin calcium trihydrate | Atorvastatin 10mg tablets (Waymade Healthcare Plc) |
| 62219 | 22613011000001104 | Atorvastatin calcium trihydrate | Atorvastatin 20mg tablets (DE Pharmaceuticals) |
| 62429 | 13831511000001104 | Atorvastatin calcium trihydrate | Atorvastatin 20mg tablets (DE Pharmaceuticals) |
| 62476 | 22047911000001109 | Atorvastatin calcium trihydrate | Atorvastatin 80mg tablets (Almus Pharmaceuticals Ltd) |
| 63140 | 20507911000001106 | Atorvastatin calcium trihydrate | Atorvastatin 10mg tablets (Alliance Healthcare (Distribution) Ltd) |
| 63249 | 20513411000001108 | Atorvastatin calcium trihydrate | Atorvastatin 80mg tablets (Consilient Health Ltd) |
| 63469 | 20512911000001107 | Atorvastatin calcium trihydrate | Atorvastatin 30mg tablets (Consilient Health Ltd) |
| 64067 | 14018211000001101 | Atorvastatin calcium trihydrate | Atorvastatin 20mg/ 5ml oral solution |
| 64702 | 21099211000001101 | Atorvastatin calcium trihydrate | Atorvastatin 30mg tablets (A A H Pharmaceuticals Ltd) |
| 64810 | 22202111000001109 | Atorvastatin calcium trihydrate | Atorvastatin 40mg tablets (Phoenix Healthcare Distribution Ltd) |
| 64825 | 22201511000001108 | Atorvastatin calcium trihydrate | Atorvastatin 10mg tablets (Phoenix Healthcare Distribution Ltd) |
| 64868 | 29772511000001108 | Atorvastatin calcium trihydrate | Atorvastatin 40mg tablets (Sigma Pharmaceuticals Plc) |
| 65193 | 22940911000001103 | Atorvastatin calcium trihydrate | Atorvastatin 20mg tablets (Ranbaxy (UK) Ltd) |

| | | | |
|---|---|---|---|
| 66963 | 29772911000001101 | Atorvastatin calcium trihydrate | Atorvastatin 80mg tablets (Sigma Pharmaceuticals Plc) |
| 67402 | 32395011000001108 | Atorvastatin calcium trihydrate | Atorvastatin 40mg tablets (Kent Pharmaceuticals Ltd) |
| 67573 | 22612811000001102 | Atorvastatin calcium trihydrate | Atorvastatin 10mg tablets (DE Pharmaceuticals) |
| 67660 | 22941511000001103 | Atorvastatin calcium trihydrate | Atorvastatin 80mg tablets (Ranbaxy (UK) Ltd) |
| 67846 | 22047311000001108 | Atorvastatin calcium trihydrate | Atorvastatin 10mg tablets (Almus Pharmaceuticals Ltd) |
| 68023 | 20496111000001100 | Atorvastatin calcium trihydrate | Atorvastatin 10mg tablets (Aspire Pharma Ltd) |
| 68048 | 22201711000001103 | Atorvastatin calcium trihydrate | Atorvastatin 20mg tablets (Phoenix Healthcare Distribution Ltd) |
| 68467 | 32394811000001103 | Atorvastatin calcium trihydrate | Atorvastatin 20mg tablets (Kent Pharmaceuticals Ltd) |
| 68785 | 33556411000001105 | Atorvastatin calcium trihydrate | Atorvastatin 10mg tablets (Mylan) |
| 68827 | 33556611000001108 | Atorvastatin calcium trihydrate | Atorvastatin 20mg tablets (Mylan) |
| 69093 | 20573911000001108 | Atorvastatin calcium trihydrate | Atorvastatin 80mg tablets (Wockhardt UK Ltd) |
| 69427 | 33556811000001107 | Atorvastatin calcium trihydrate | Atorvastatin 40mg tablets (Mylan) |
| 70693 | 29771911000001107 | Atorvastatin calcium trihydrate | Atorvastatin 10mg tablets (Sigma Pharmaceuticals Plc) |
| 70987 | 20982011000001104 | Atorvastatin calcium trihydrate | Atorvastatin 10mg tablets (Dr Reddy's Laboratories (UK) Ltd) |
| 71017 | 20982411000001108 | Atorvastatin calcium trihydrate | Atorvastatin 20mg tablets (Dr Reddy's Laboratories (UK) Ltd) |
| 72164 | 34961811000001104 | Atorvastatin calcium trihydrate | Atorvastatin 20mg tablets (Bristol Laboratories Ltd) |
| 72213 | 20572711000001103 | Atorvastatin calcium trihydrate | Atorvastatin 20mg tablets (Wockhardt UK Ltd) |
| 72641 | 14158811000001101 | Atorvastatin calcium trihydrate | Atorvastatin 40mg/ 5ml oral solution |
| 73520 | 13923011000001100 | Atorvastatin calcium trihydrate | Lipitor 20mg tablets (DE Pharmaceuticals) |
| 74518 | 20572511000001108 | Atorvastatin calcium trihydrate | Atorvastatin 10mg tablets (Wockhardt UK Ltd) |
| 75391 | 34682311000001101 | Atorvastatin calcium trihydrate | Atorvastatin 80mg/ 5ml oral suspension |
| 75622 | | Atorvastatin calcium trihydrate | Atorvastatin 60mg tablets (A A H Pharmaceuticals Ltd) |
| 77344 | | Atorvastatin calcium trihydrate | Atorvastatin 80mg tablets (Mylan) |
| 77427 | | Atorvastatin calcium trihydrate | Lipitor 10mg tablets (Waymade Healthcare Plc) |
| 77434 | | Atorvastatin calcium trihydrate | Lipitor 20mg tablets (Waymade Healthcare Plc) |
| 77782 | | Atorvastatin calcium trihydrate | Atorvastatin 40mg tablets (Bristol Laboratories Ltd) |
| 24509 | | | SIMVASTATIN |
| 29438 | | | SIMVASTATIN |
